# Cardiovascular drugs and COVID-19 clinical outcomes: a living systematic review and meta-analysis

**DOI:** 10.1101/2020.10.07.20208918

**Authors:** Innocent G Asiimwe, Sudeep Pushpakom, Richard M Turner, Ruwanthi Kolamunnage-Dona, Andrea Jorgensen, Munir Pirmohamed

**Affiliations:** The Wolfson Centre for Personalized Medicine, MRC Centre for Drug Safety Science, Department of Pharmacology and Therapeutics, Institute of Systems, Molecular and Integrative Biology, University of Liverpool, L69 3GL, Liverpool, United Kingdom; Department of Biostatistics, Institute of Population Health Sciences, University of Liverpool, L69 3GL, Liverpool, United Kingdom

**Author notes:** Correspondence to: Innocent G. Asiimwe; Munir Pirmohamed.

## Abstract

**OBJECTIVE:** To continually evaluate the rapidly evolving evidence base on the role of cardiovascular drugs in COVID-19 clinical outcomes (susceptibility to infection, hospitalization, hospitalization length, disease severity, and all-cause mortality).

**DESIGN:** Living systematic review and meta-analysis.

**DATA SOURCES:** Eligible publications identified from >500 databases indexed through 31^st^ July 2020 and additional studies from reference lists, with planned continual surveillance for at least two years.

**STUDY SELECTION:** Observational and interventional studies that report on the association between cardiovascular drugs and COVID-19 clinical outcomes.

**DATA EXTRACTION:** Single-reviewer extraction and quality evaluation (using ROBINS-I), with half the records independently extracted and evaluated by a second reviewer.

**RESULTS:** Of 23,427 titles screened, 175 studies were included in the quantitative synthesis. The most reported drug classes were angiotensin-converting enzyme inhibitors (ACEIs) and angiotensin receptor blockers (ARBs) with ACEI/ARB exposure being associated with higher odds of testing positive for COVID-19 (pooled unadjusted OR 1.15, 95% CI 1.02 to 1.30). Among patients with COVID-19, unadjusted estimates showed that ACEI/ARB exposure was associated with being hospitalized (OR 2.25, 1.70 to 2.98) and having severe disease (OR 1.50, 1.27 to 1.77) but not with the length of hospitalization (mean difference −0.45, −1.33 to 0.43 days) or all-cause mortality (OR 1.25, CI 0.98 to 1.58). However, after adjustment, ACEI/ARB exposure was not associated with testing positive for COVID-19 (pooled adjusted OR 1.01, 0.93 to 1.10), being hospitalized (OR 1.16, 0.80 to 1.68), having severe disease (1.04, 0.76 to 1.42), or all-cause mortality (0.86, 0.64 to 1.15). Similarly, subgroup analyses involving only hypertensive patients revealed that ACEI/ARB exposure was not associated with being hospitalized (OR 0.84, 0.58 to 1.22), disease severity (OR 0.88, 0.68 to 1.14) or all-cause mortality (OR 0.77, 0.54 to 1.12) while it decreased the length of hospitalization (mean difference −0.71, −1.11 to −0.30 days). After adjusting for relevant covariates, other cardiovascular drug classes were mostly not found to be associated with poor COVID-19 clinical outcomes. However, the validity of these findings is limited by a high level of heterogeneity in terms of effect sizes and a serious risk of bias, mainly due to confounding in the included studies.

**CONCLUSION:** Our comprehensive review shows that ACEI/ARB exposure is associated with COVID-19 outcomes such as susceptibility to infection, severity, and hospitalization in unadjusted analyses. However, after adjusting for potential confounding factors, this association is not evident. Patients on cardiovascular drugs should continue taking their medications as currently recommended. Higher quality evidence in the form of randomized controlled trials will be needed to determine any adverse or beneficial effects of cardiovascular drugs.

**PRIMARY FUNDING SOURCE:** None

**SYSTEMATIC REVIEW REGISTRATION:** PROSPERO (CRD42020191283)

## BACKGROUND

Coronavirus disease 2019 (COVID-19) was first reported on 8 December 2019 in Wuhan, Hubei province, China.^1^ It is caused by severe acute respiratory syndrome coronavirus 2 (SARS-CoV-2), which infects cells through the human angiotensin-converting enzyme 2 (ACE2) receptor.^2^ It was designated a pandemic by the World Health Organization on 11 March 2020^3^ and has since affected 188 countries, more than 31 million patients and led to over 957,000 deaths (as of 20 September 2020^4^). To put it into context, cardiovascular diseases such as ischemic heart disease, stroke and heart failure remain the leading causes of global deaths, being responsible for an estimated 17 ·8 million deaths in 2017.^5^ The interaction between COVID-19 and cardiovascular disease appears complex and bi-directional with cardiovascular disease increasing susceptibility to SARS-CoV-2 infection or COVID-19 severity and at the same time COVID-19 causing injury to the cardiovascular system in some patients.^6-9^ Consequently, the relationship between COVID-19 and cardiovascular drugs is of interest because: a) patients with increased susceptibility to SARS-CoV-2 infection may be taking these drugs, b) they may alleviate cardiovascular injury caused by COVID-19, and c) cardiovascular drugs such as angiotensin-converting enzyme inhibitors (ACEIs) or angiotensin receptor blockers (ARBs) may play a direct role in COVID-19 pathology.^2^

Recent systematic reviews and/or meta-analyses^10-21^ have characterized the relationship between COVID-19 outcomes (such as severity and mortality) and cardiovascular drugs. These reviews have, however, been limited in scope in terms of the COVID-19 outcomes and cardiovascular drugs studied. For example, most have focused on ACEIs and ARBs given their interaction with the ACE2 receptor that facilitates SARS-CoV-2 entry into cells.^2^ However, being a novel disease, a lot is still unknown about COVID-19 which makes a broader systematic review (in terms of the drugs studied) necessary. Moreover, there are emerging reports that other drug classes such as anticoagulants, calcium channel blockers and statins could be beneficial.^22-25^ Additionally, many cardiovascular disease patients are on combination therapies and a broader review may facilitate understanding of the interplay between the different classes of cardiovascular drugs. Lastly, evidence in this field is rapidly evolving which means that recently published reviews soon become outdated. To provide more comprehensive and up-to-date evidence, we have conducted a systematic review and meta-analysis to evaluate all the current evidence on the influence of cardiovascular drugs on COVID-19 clinical outcomes. Due to the rapidly evolving nature of this field, we will periodically update this baseline review for up to two years to reflect emerging evidence.

## METHODS

A predefined protocol (PROSPERO: CRD42020191283), based on the principles of the Cochrane Handbook for Systematic Reviews of Interventions^26^ with living systematic review considerations^27 28^ was followed. This report adheres to the Preferred Reporting Items for Systematic Reviews and Meta-Analyses (PRISMA) statement^29^ (Table S1).

### Identification of studies

A final search of the University of Liverpool’s DISCOVER platform (which links, through EBSCOhost, to sources from >500 databases including MEDLINE, Google Scholar, Scopus, the Web of Science and Cochrane Central Register of Controlled Trial libraries) was undertaken on 31st July 2020 using medical subject headings and text words related to “cardiovascular drugs” and “COVID-19” (Text S1). A separate MEDLINE search (Text S1) was conducted to ensure that the DISCOVER search was retrieving all eligible records. Lists of references from the identified studies and previous systematic reviews were hand-searched to identify additional eligible articles. Preprint servers (bioRxiv and medRxiv, Text S1), COVID-19 specific databases (such as the COVID-19 Clinical Trials registry and the World Health Organization database of COVID-19 publications), other registries/results databases (such as ClinicalTrials.gov and the International Clinical Trials Registry Platform) and grey literature were also searched to identify further eligible studies. After this baseline review is published, monthly searches will be conducted in the major bibliographic databases using the DISCOVER platform, with quarterly searches applying to the other sources (including additional preprint servers such as Authorea.com, Preprints from Lancet, and Preprints.org).

### Selection criteria

Both observational (e.g. cohorts/case-series and case-control studies) and interventional (e.g. randomised controlled trials) studies investigating the association between cardiovascular drugs and COVID-19 were included. Cardiovascular drugs were defined as those found in Chapter 2 (“Cardiovascular system”) of the British National Formulary^30^ while COVID-19 clinical outcomes included those outlined below. Unless translated text could be obtained, non-English studies were excluded.

### Outcomes

COVID-19 clinical outcomes included susceptibility to infection, disease severity (Text S2), hospitalization, hospitalization length, and all-cause mortality.

### Data extraction

Two reviewers (IGA for DISCOVER and SP for MEDLINE) independently screened titles and abstracts of the retrieved bibliographic records according to eligibility. Where no abstract was available, the full text was obtained unless the article could be confidently excluded by its title alone. Full texts of potentially eligible studies were retrieved, a data extraction form developed and piloted in a subset of ten randomly selected papers and used to extract relevant information (related to study design, patient characteristics, cardiovascular drugs, COVID-19 outcomes, and study quality). Data from all eligible studies were extracted (Text S3 for more details) and summarized by one reviewer (IGA). As a quality control measure, a second reviewer (SP or RT) independently extracted and evaluated half the records to ascertain consistency. Any disagreements were resolved by consensus.

### Assessment of study quality

To assess the quality of each included study estimate, the revised Cochrane risk-of-bias tool for randomized trials^31^ and ROBINS-I (Risk Of Bias In Non-randomised Studies - of Interventions) tool for non-randomized studies^32^ were used. When applying ROBINS-I, we bore in mind that our aim was to quantify the effect of starting and adhering to the intervention under study on our outcomes, and so co-interventions such as other cardiovascular drugs, steroids, antivirals or procedures like mechanical ventilation that could differ between intervention groups and so could impact the outcomes were considered. Based on several systematic reviews of risk factors for COVID-19 outcomes,^8 33-44^ the confounding domains we thought relevant to all or most studies included age, race, gender, cardiovascular comorbidities (including hypertension, diabetes, and obesity), smoking and clinical setting/location.

### Data synthesis

Where ≥ 2 studies reporting on the same exposure-outcome combination were reported, effect estimates were pooled by way of random-effects meta-analyses using R version 3.6.1^45^ (R meta package^46^). Odds ratios and mean differences (with 95% confidence intervals) were generated for dichotomous and continuous outcomes, respectively. Both unadjusted (or in the case of binary outcomes, count data, which is preferred to unadjusted odds ratios as it provides more reliable estimates^47^) and adjusted estimates were extracted and pooled separately. Where there was more than one adjusted estimate, the estimate adjusting for the most covariates was preferred. Since different studies adjust for different covariates, we did not limit our inclusion criteria to a given set of covariates. Where median values and ranges/interquartile ranges were provided (for example for length of hospitalization), they were used to estimate the mean values and standard deviations.^48^ Where necessary, means and standard deviations were combined using formulae available in the Cochrane Handbook.^49^ Forest plots were prepared for each exposure-outcome combination. Studies that could not be pooled due to being the only ones reporting on an exposure-outcome combination were also included as part of qualitative synthesis.

### Heterogeneity measures

The magnitude of inconsistency in the study results was assessed by visually examining forest plots and considering the *I*^2^ statistic.^26 50^ Arbitrarily-defined categories of heterogeneity were: *I*^2^ <30 %, low; *I*^2^ = 30–70 %, moderate; and *I*^2^ >70 %, high. Potential sources of heterogeneity were explored during the subgroup analyses (see below).

### Publication bias

Where enough (≥10) studies were available for a given exposure-outcome combination, publication bias was assessed using the linear regression test of funnel plot asymmetry (Egger’s test, implemented using the metabias function in the R meta package^46^). A p-value <0.1 was considered to suggest the presence of publication bias. When asymmetry was suggested by a visual assessment, we performed exploratory analyses to investigate and adjust for it (trim and fill analysis) using the trimfill function (R metafor package^51^).

### Sensitivity and subgroup analyses

Due to the need to provide results in real-time, many COVID-19 studies are being directly uploaded to preprint servers and/or undergoing rapid peer review. Therefore, in addition to a robust quality assessment, we conducted sensitivity analyses in which prominent/outlier studies were excluded to identify those studies whose retraction would significantly alter the pooled estimates. Prominent studies were identified through visual examination of the forest plots (criteria including accorded weights and whether individual estimates were consistent with other study estimates). Random effects meta-regression (Text S4) was conducted to both explore heterogeneity and inform sub-group analyses. Where possible, sub-group analyses based on drug sub-classes were also conducted.

### Confidence in cumulative evidence

The strength of the body of evidence and the quality and strength of recommendations was assessed according to the GRADE (Grading of Recommendations, Assessment, Development and Evaluations) criteria.^52 53^

### Review updating

After each monthly search, new evidence will be briefly summarized unless it changes the nature or strength of the conclusions, in which case a major update will be performed. The COVID-19 situation is extremely dynamic, and it is not possible to tell when we will be transitioning out of the living systematic review mode (updating for up to two years planned). The review scope and methods will, however, be reviewed at least once a year.

## RESULTS

### Study selection and characteristics

Of the 23,427 titles screened, 178 and 175 studies were included in the qualitative and quantitative syntheses respectively (Figure 1). The characteristics of the included studies are shown in Table S2. Out of the 178 studies, more than a third (*n* = 67, 38%) were preprints. Most studies (*n* = 163, 92%) had a cohort/case series design with 14 (8%) being case-control studies. Only one (<1%) study was an open-label randomized control trial; however, it conducted a retrospective/non-pre-specified interim analysis of its currently recruited trial participants. The most commonly reported drug exposure was with angiotensin-converting enzyme inhibitor (ACEI)/angiotensin receptor blocker (ARB) (ACEI/ARB), which therefore became the main focus.

**Figure 1.**
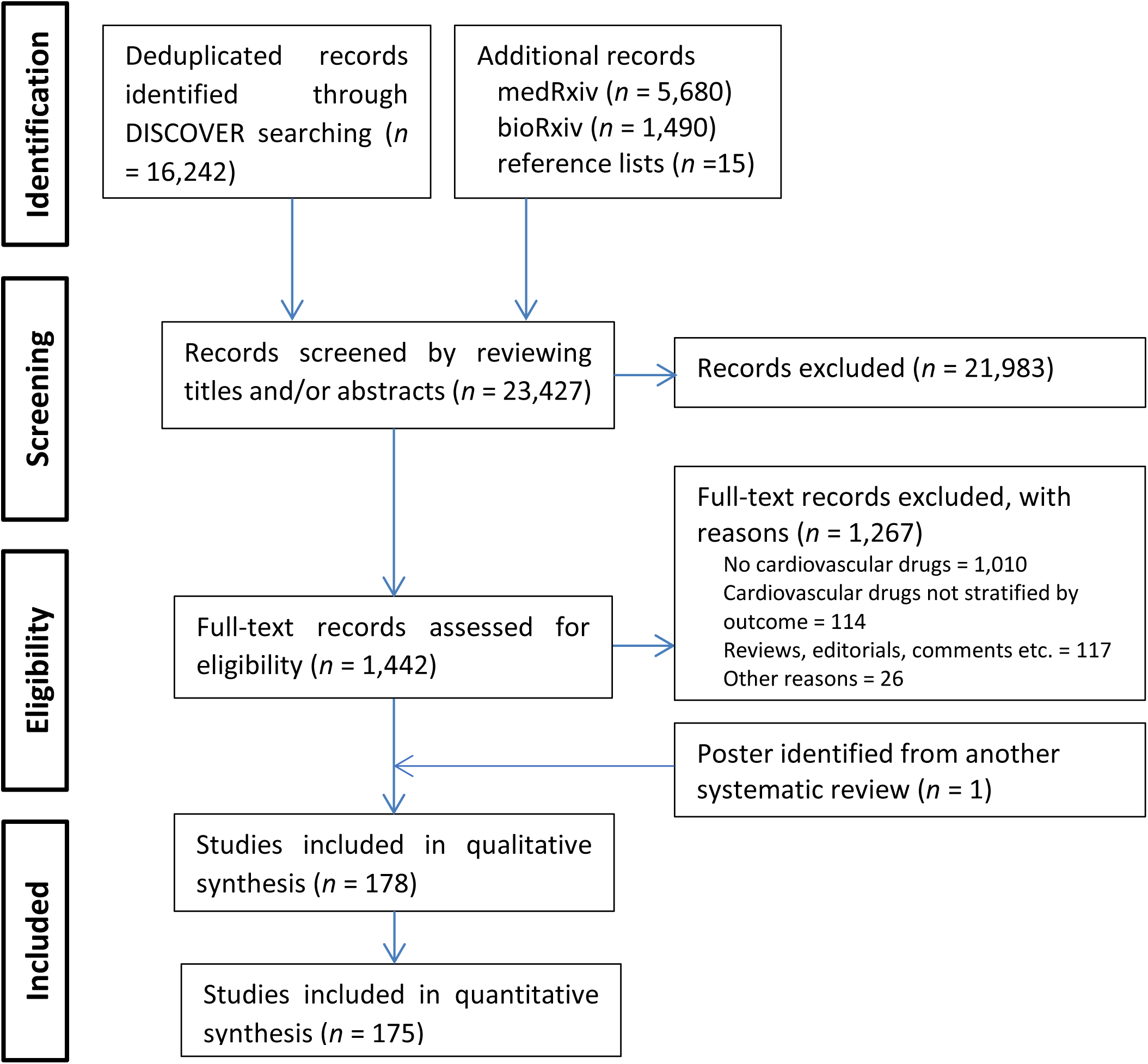
PRISMA Flow Chart of Included Studies.

The summary risk of bias assessment for ACEI/ARB exposure is shown in Figure S1 (unadjusted estimates, all five outcomes). All studies had serious risks of bias, mainly driven by confounding (only counts/unadjusted estimates considered, with no adjustment for potential confounders) and inappropriate selection of participants into the study (selection into the study was related to ACEI/ARB exposure and severity/hospitalization/mortality outcomes). The risk of bias in classification of interventions was generally considered low because of the chronic nature of most drug exposures while there was little information to assess the risk of bias due to deviations from intended interventions (we aimed to quantify the effect of starting and adhering to the intervention as explained under the methods section, although information on adherence was rarely reported). Although we did not examine each study’s individual analysis plan, the risk of bias due to missing data was generally considered to be low. We also considered the risk of bias in outcome measurement to be low, except for the susceptibility to infection outcome in which about 46% of the studies were at risk of differential misclassification (i.e. due to low testing rates, some participants considered negative could have been unknown COVID-19 cases). Although it could not be ruled out, bias during the selection of the reported result was considered moderate since in the primary meta-analyses the raw count data was used to calculate effect estimates. At least one of the first two domains was rated serious for all estimates which implied that the overall risk of bias rating for all studies had to be rated ‘serious’. Because all estimates were rated as having a serious risk of bias, no sensitivity analyses based on methodological rigor were performed (the risk of bias assessments for all individual estimates are shown in the corresponding forest plots).

### Meta-analysis

Table 1, Tables S3-S27 and Figures S2-S60 summarise the pooled estimates for the associations between all cardiovascular drug exposures and the various COVID-19 clinical outcomes. The text below is focused on the most reported drug (ACEI/ARB) exposure.

**Table 1.** Summary results for associations between cardiovascular drug exposure and COVID-19 outcomes*.

| Outcome | Exposure | All studies | Primary meta-analysis |  |  |  |  | Adjusted estimates (95% CI) | Reference Figures/ Tables |
| --- | --- | --- | --- | --- | --- | --- | --- | --- | --- |
| | | | Included studies | Sample size | Unadjusted estimates (95% CI), heterogeneity ($I^2$) | Covariates important in meta-regression Sub-group analyses | Egger's p | | |
| Susceptibility | ACEI/ARB | 31 | 24 | 1,362,182 | OR 1.15 (1.02; 1.30), $I^2=93\%$ | Country, study design<br>China-only ($n=2$): OR 0.70 (0.35; 1.40), $I^2=70\%$<br>Not in China ($n=22$): OR 1.19 (1.06; 1.35), $I^2=93\%$<br>Single centre ($n=10$): OR 1.51 (1.36; 1.69), $I^2=0\%$<br>Multicentre ($n=14$): OR 1.04 (0.91; 1.20), $I^2=95\%$<br>Cohorts ($n=16$): OR 1.33 (1.15; 1.55), $I^2=89\%$<br>Case-controls ($n=8$): OR 0.91 (0.75; 1.11), $I^2=94\%$ | 0.89 | OR ($n=6$): 1.01 (0.93; 1.10), $I^2=0\%$<br>HR ($n=2$): 1.05 (0.92; 1.20), $I^2=0\%$ | Figures 2, S2-S3; Table S3 |
| | ACEI | 20 | 17 | 818,018 | OR 1.09 (0.97; 1.23), $I^2=84\%$ | Study design<br>Single centre ($n=5$): OR 1.48 (1.25; 1.75), $I^2=0\%$<br>Multicentre ($n=12$): OR 1.02 (0.89; 1.17), $I^2=87\%$ | 0.76 | OR ($n=9$): 0.97 (0.87; 1.09), $I^2=0\%$ | Figures S4-S5; Table S4 |
| | ARB | 20 | 17 | 823,654 | OR 1.11 (0.96; 1.28), $I^2=90\%$ | Publication status, study design<br>Peer-reviewed ($n=9$): OR 1.24 (1.07; 1.43), $I^2=79\%$<br>Preprints ($n=8$): OR 0.96 (0.80; 1.16), $I^2=84\%$<br>Single centre ($n=5$): OR 1.42 (1.23; 1.63), $I^2=0\%$<br>Multicentre ($n=12$): OR 1.03 (0.87; 1.21), $I^2=93\%$<br>Cohorts ($n=10$): OR 1.30 (1.13; 1.51), $I^2=72\%$<br>Case-controls ($n=7$): OR 0.91 (0.74; 1.12), $I^2=92\%$ | 0.83 | OR ($n=9$): 0.90 (0.65; 1.24), $I^2=89\%$; when one outlier study removed, OR 1.04 (0.96; 1.12), $I^2=10\%$ | Figures S6-S7; Table S5 |
| | Anticoagulant | 9 | 9 | 333,417 | OR 1.17 (0.98; 1.41), $I^2=86\%$ | NA | NA | OR ($n=2$): 0.86 (0.47; 1.56), $I^2=95\%$ | Figures S8-S9 |
| | Antiplatelet | 6 | 6 | 417,227 | OR 1.07 (0.85; 1.35), $I^2=93\%$ | NA | NA | OR ($n=3$): 0.78 (0.32; 1.93), $I^2=87\%$ | Figures S8-S9 |
| | Beta blocker | 10 | 9 | 1,744,871 | OR 1.15 (1.02; 1.29), $I^2=84\%$ | NA | NA | OR ($n=5$): 0.94 (0.84; 1.06), $I^2=20\%$ | Figures S8-S9 |
| | CCB | 10 | 9 | 1,744,871 | OR 1.12 (0.93; 1.35), $I^2=95\%$ | NA | NA | OR ($n=5$): 1.06 (0.84; 1.35), $I^2=72\%$ | Figures S8-S9 |
| | Class III anti-arrhythmic (Amiodarone) | 3 | 3 | 102,242 | OR 0.65 (0.17; 2.47), $I^2=92\%$ | NA | NA | NA | Figure S8 |
| | Diuretics | 9 | 9 | 5,609,843 | OR 1.37 (1.16; 1.62), $I^2=96\%$ | Loop diuretics ($n=4$): OR 1.92 (1.35; 2.71), $I^2=97\%$<br>Thiazides ($n=5$): OR 1.14 (0.88; 1.47), $I^2=95\%$<br>Other<sup>†</sup> ($n=3$): OR 1.80 (1.47; 2.21), $I^2=45\%$ | NA | OR ($n=5$): 0.82 (0.53; 1.27), $I^2=83\%$ | Figures S8-S9 |
| | Lipid modifying drug | 11 | 11 | 708,933 | OR 1.16 (0.98; 1.38), $I^2=94\%$ | COVID-1 testing, diabetes mellitus<br>All tested ($n=4$): OR 0.96 (0.70; 1.31), $I^2=86\%$<br>Not all tested ($n=7$): OR 1.28 (1.10; 1.49), $I^2=88\%$<br>Statins ($n=7$): OR 1.25 (0.80; 1.95), $I^2=93\%$ | 0.81 | OR ($n=5$): 1.01 (0.65; 1.56), $I^2=82\%$ | Figures S10-S11; Table S6 |
| Hospitalization | ACEI/ARB | 23 | 20 | 27,413 | OR 2.25 (1.70; 2.98), $I^2=91\%$ | Age, hypertension<br>All hypertensive ($n=4$): OR 0.84 (0.58; 1.22), $I^2=66\%$ | 0.13 | OR ($n=6$): 1.16 (0.80; 1.68), $I^2=53\%$<br>HR ($n=2$): 1.00 (0.87; 1.14), $I^2=0\%$ | Figures 2-3, S12-S13; Table S7 |
| | ACEI | 15 | 12 | 11,159 | OR 2.51 (1.44; 4.36), $I^2=93\%$ | None | 0.85 | OR ($n=6$): 0.78 (0.47; 1.28), $I^2=73\%$<br>HR ($n=2$): 1.04 (0.64; 1.68), $I^2=84\%$ | Figures S14-S15; Table S8 |
| | ARB | 15 | 12 | 11,159 | OR 2.16 (1.16; 4.00), $I^2=94\%$ | Age, hypertension<br>All hypertensive ($n=3$): OR 0.77 (0.46; 1.29), $I^2=39\%$ | 0.63 | OR ($n=6$): 1.09 (0.67; 1.77), $I^2=51\%$<br>HR ($n=2$): 1.07 (0.79; 1.43), $I^2=55\%$ | Figures S16-S17; Table S9 |
| | Anticoagulants | 7 | 6 | 12,630 | OR 3.16 (1.66; 6.01), $I^2=89\%$ | NA | NA | NA | Figure S18 |
| | Antiplatelets | 5 | 4 | 12,367 | OR 3.27 (1.85; 5.76), $I^2=87\%$ | NA | NA | NA | Figure S18 |
| | Beta blocker | 7 | 7 | 14,158 | OR 2.47 (1.35; 4.55), $I^2=93\%$ | NA | NA | OR ($n=3$): 0.86 (0.38; 1.96), $I^2=77\%$ | Figures S18-S19 |
| | CCB | 6 | 5 | 13,135 | OR 2.38 (1.42; 3.98), $I^2=87\%$ | NA | NA | OR ($n=3$): 1.16 (0.83; 1.63), $I^2=0\%$ | Figures S18-S19 |
| | Diuretic | 5 | 5 | 34,774 | OR 3.09 (1.64; 5.82), $I^2=97\%$ | NA | NA | OR ($n=4$): 1.26 (0.90; 1.76), $I^2=0\%$ | Figures S18-S19 |
| | Lipid modifying drug | 5 | 5 | 13,093 | OR 3.88 (2.92; 5.17), $I^2=72\%$ | NA | NA | OR ($n=2$): 1.00 (0.31; 3.21), $I^2=86\%$ | Figures S18-S19 |

Table 1. Continued.
| Outcome | Exposure | All studies | Primary meta-analysis |  |  |  |  | Adjusted estimates (95% CI) | Reference Figures/ Tables |
| --- | --- | --- | --- | --- | --- | --- | --- | --- | --- |
| | | | Included studies | Sample size | Unadjusted estimates (95% CI), heterogeneity ($I^2$) | Covariates important in meta-regression Sub-group analyses | Egger's p | | |
| Hospitalization length | ACEI/ARB | 12 | 11 | 2,510 | MD -0.45 (-1.33; 0.43) days, $I^2=30\%$ | Age, hypertension<br>All hypertensive ($n=9$): MD -0.71 (-1.11; -0.30) days, $I^2=0\%$ | 0.94 | NA | Figures 2-3, S20-S21; Table S10 |
| Severity | ACEI/ARB | 76 | 60 | 45,394 | OR 1.50 (1.27; 1.77), $I^2=81\%$ | Country, setting, sample size, age, hypertension, diabetes mellitus<br>China-only ($n=14$): OR 0.97 (0.71; 1.33), $I^2=58\%$<br>Not in China ($n=46$): OR 1.70 (1.41; 2.04), $I^2=82\%$<br>South Korea-only ($n=2$): OR 4.04 (2.28; 7.16), $I^2=0\%$<br>Not in S.Korea ($n=58$): OR 1.45 (1.23; 1.72), $I^2=81\%$<br>All inpatient ($n=48$): OR 1.29 (1.12; 1.49), $I^2=61\%$<br>All hypertensive ($n=22$): OR 0.88 (0.68; 1.14), $I^2=77\%$<br>All diabetic ($n=2$): OR 1.27 (1.01; 1.60), $I^2=0\%$ | 0.15 | OR ($n=18$): 1.04 (0.76; 1.42), $I^2=65\%$<br>HR ($n=4$): 0.98 (0.72; 1.33), $I^2=68\%$ | Figure 2, 3, S22-S23; Table S11 |
| | ACEI | 35 | 32 | 25,962 | OR 1.67 (1.33; 2.10), $I^2=77\%$ | Setting, age, hypertension, diabetes mellitus<br>All inpatient ($n=24$): OR 1.27 (1.08; 1.50), $I^2=22\%$<br>All hypertensive ($n=11$): OR 0.89 (0.70; 1.13), $I^2=40\%$ | 0.37 | OR ($n=10$): 0.72 (0.46; 1.13), $I^2=53\%$<br>HR ($n=2$): 1.18 (1.07; 1.30), $I^2=0\%$ | Figures S24-S25; Table S12 |
| | ARB | 35 | 32 | 26,106 | OR 1.48 (1.19; 1.85), $I^2=79\%$ | Country, setting, sample size, age, hypertension, diabetes mellitus<br>China-only ($n=5$): OR 0.79 (0.48; 1.32), $I^2=66\%$<br>Not in China ($n=27$): OR 1.69 (1.34; 2.13), $I^2=78\%$<br>USA-only ($n=9$): OR 2.08 (1.26; 3.43), $I^2=89\%$<br>Not in USA ($n=23$): OR 1.28 (1.01; 1.63), $I^2=69\%$<br>All inpatient ($n=25$): OR 1.23 (1.04; 1.46), $I^2=34\%$<br>All hypertensive ($n=12$): OR 0.99 (0.81; 1.21), $I^2=39\%$ | 0.28 | OR ($n=12$): 1.12 (0.69; 1.82), $I^2=52\%$<br>HR ($n=2$): 1.05 (0.74; 1.48), $I^2=61\%$ | Figures S26-S27; Table S13 |
| | Anticoagulant | 14 | 14 | 14,779 | OR 1.99 (1.52; 2.62), $I^2=34\%$ | None | 0.20 | OR ($n=2$): 1.35 (0.80; 2.29), $I^2=0\%$ | Figures S28-S29; Table S14 |
| | Antiplatelet | 10 | 10 | 14,745 | OR 1.39 (0.86; 2.24), $I^2=79\%$ | Sample size, diabetes mellitus, publication status<br>Peer-reviewed ($n=5$): OR 0.73 (0.46; 1.15), $I^2=5\%$<br>Preprints ($n=5$): OR 2.43 (1.70; 3.45), $I^2=56\%$ | 0.004 | OR ($n=2$): 0.70 (0.39; 1.25), $I^2=0\%$ | Figures S30-S32; Table S15 |
| | Beta blocker | 12 | 12 | 20,524 | OR 1.63 (1.17; 2.27), $I^2=79\%$ | Sample size, age, gender, hypertension, diabetes mellitus<br>All hypertensive ($n=5$): OR 0.98 (0.80; 1.20), $I^2=0\%$ | 0.61 | OR ($n=5$): 1.19 (0.54; 2.65), $I^2=46\%$ | Figures S33-S34; Table S16 |
| | CCB | 15 | 14 | 16,910 | OR 1.55 (1.14; 2.09), $I^2=73\%$ | Country, sample size, age, hypertension, diabetes mellitus<br>China-only ($n=4$): OR 1.03 (0.71; 1.48), $I^2=24\%$<br>Not in China ($n=10$): OR 1.93 (1.40; 2.67), $I^2=65\%$<br>All hypertensive ($n=7$): OR 1.26 (1.06; 1.49), $I^2=0\%$ | 0.11 | OR ($n=6$): 1.31 (0.62; 2.80), $I^2=66\%$ | Figures S35-S36; Table S17 |
| | Diuretic | 9 | 9 | 19,928 | OR 1.33 (1.15; 1.53), $I^2=0\%$ | Loop diuretics ($n=3$): OR 1.66 (0.82; 3.37), $I^2=89\%$<br>Thiazides ($n=5$): OR 1.33 (1.11; 1.59), $I^2=12\%$<br>Other<sup>†</sup> ($n=4$): OR 1.20 (0.78; 1.85), $I^2=0\%$ | NA | OR ($n=5$): 0.91 (0.65; 1.27), $I^2=0\%$ | Figures S37 |
| | Lipid modifying drug | 16 | 15 | 17,155 | OR 1.68 (1.18; 2.38), $I^2=83\%$ | Publication status, sample size, age, hypertension, diabetes mellitus<br>Peer-reviewed ($n=7$): OR 1.22 (0.87; 1.72), $I^2=48\%$<br>Preprints ($n=8$): OR 2.14 (1.48; 3.11), $I^2=71\%$ | 0.72 | OR ($n=5$): 1.63 (0.57; 4.65), $I^2=72\%$ | Figures S38-S39; Table S18 |

**Table 1. Continued.**
| Outcome | Exposure | All studies | Primary meta-analysis |  |  |  |  | Adjusted estimates (95% CI) | Reference Figures/ Tables |
| --- | --- | --- | --- | --- | --- | --- | --- | --- | --- |
| | | | Included studies | Sample size | Unadjusted estimates (95% CI), heterogeneity ($I^2$) | Covariates important in meta-regression Sub-group analyses | Egger's p | | |
| All-cause mortality | ACEI/ARB | 73 | 40 | 43,099 | OR 1.25 (0.98; 1.58), $I^2=85\%$ | Country, sample size, age, hypertension<br>China-only ($n=12$): OR 0.63 (0.46; 0.85), $I^2=0\%$<br>Not in China ($n=28$): OR 1.54 (1.18; 1.99), $I^2=88\%$<br>All hypertensive ($n=18$): OR 0.77 (0.54; 1.12), $I^2=68\%$ | 0.20 | OR ($n=13$): 0.86 (0.64; 1.15), $I^2=4\%$<br>HR ($n=10$): 0.94 (0.79; 1.13), $I^2=29\%$ | Figures 2-3 S40-S41; Table S19 |
| | ACEI | 30 | 20 | 32,601 | OR 1.38 (1.03; 1.85), $I^2=79\%$ | Sample size, age, hypertension<br>All hypertensive ($n=6$): OR 0.90 (0.79; 1.04), $I^2=0\%$ | 0.73 | OR ($n=5$): 0.80 (0.46; 1.38), $I^2=58\%$<br>HR ($n=5$): 1.03 (0.90; 1.18), $I^2=0\%$ | Figures S42-S43; Table S20 |
| | ARB | 27 | 18 | 32,407 | OR 1.43 (1.05; 1.93), $I^2=84\%$ | Sample size, age, hypertension<br>All hypertensive ($n=6$): OR 0.97 (0.73; 1.29), $I^2=19\%$ | 0.90 | OR ($n=4$): OR 1.11 (0.94; 1.32), $I^2=51\%$<br>HR ($n=5$): 1.05 (0.90; 1.22), $I^2=10\%$ | Figures S44-S45; Table S21 |
| | Anticoagulant | 24 | 17 | 17,687 | OR 0.97 (0.48; 1.94), $I^2=96\%$ | Setting, sample size, gender<br>All inpatient ($n=13$): OR 0.82 (0.56; 1.18), $I^2=71\%$ | 0.28 | OR ($n=3$): OR 1.05 (0.51; 2.18), $I^2=82\%$ | Figures S46-S47; Table S22 |
| | Antiplatelet | 16 | 13 | 14,634 | OR 1.85 (1.08; 3.16), $I^2=91\%$ | Publication status, sample size, age, hypertension<br>Peer-reviewed ($n=10$): OR 1.50 (1.02; 2.19), $I^2=48\%$<br>Preprints ($n=3$): OR 4.99 (2.14; 11.64), $I^2=90\%$ | 0.12 | NA | Figures S48-S49; Table S23 |
| | Beta blocker | 13 | 13 | 18,583 | OR 2.16 (1.47; 3.20), $I^2=87\%$ | Sample size, age, hypertension<br>All hypertensive ($n=2$): OR 1.64 (0.59; 4.54), $I^2=54\%$ | 0.05 | OR ($n=2$): 1.44 (1.19; 1.75), $I^2=0\%$ | Figures S50-S52; Table S24 |
| | CCB | 15 | 12 | 17,970 | OR 1.43 (1.03; 1.99), $I^2=75\%$ | Sample size, age, hypertension, diabetes mellitus<br>All hypertensive ($n=4$): OR 1.06 (0.74; 1.53), $I^2=11\%$ | 0.04 | OR ($n=2$): 0.86 (0.60; 1.24), $I^2=10\%$<br>HR ($n=2$): 0.34 (0.04; 3.16), $I^2=84\%$ | Figures S53-S55; Table S25 |
| | Diuretic | 12 | 11 | 42,290 | OR 3.45 (2.30; 5.18), $I^2=92\%$ | Publication status, country, setting, age, hypertension<br>Peer-reviewed ($n=9$): OR 2.74 (1.82; 4.11), $I^2=81\%$<br>Preprints ($n=2$): OR 7.43 (3.02; 18.28), $I^2=98\%$<br>France-only ($n=2$): OR 1.40 (0.41; 4.80), $I^2=87\%$<br>Not in France ($n=9$): OR 4.28 (2.84; 6.44), $I^2=91\%$<br>Inpatient ($n=8$): OR 2.21 (1.55; 3.17), $I^2=63\%$<br>Loop diuretics ($n=5$): OR 6.07 (3.09; 11.90), $I^2=94\%$<br>Thiazides ($n=5$): OR 1.82 (0.90; 3.69), $I^2=93\%$<br>Other<sup>†</sup> ($n=5$): OR 2.53 (1.33; 4.82), $I^2=74\%$ | 0.18 | OR ($n=2$): 1.55 (1.25; 1.91), $I^2=0\%$<br>HR ($n=2$): 1.06 (0.93; 1.22), $I^2=0\%$ | Figures S56-S57; Table S26 |
| | Lipid modifying drug | 19 | 12 | 32,625 | 1.54 (1.09; 2.19), $I^2=90\%$ | None<br>Statins ($n=8$): OR 1.41 (0.97; 2.04), $I^2=79\%$ | 0.54 | OR ($n=4$): 1.08 (0.68; 1.71), $I^2=33\%$<br>HR ($n=4$): 0.67 (0.47; 0.96), $I^2=83\%$ | Figures S58-S59; Table S27 |
| | Vasopressors | 4 | 4 | 3,572 | OR 10.24 (1.56; 67.40), $I^2=94\%$ | NA | NA | NA | Figure S60 |

\*In terms of GRADE rating, all estimates were downgraded to moderate certainty due to a serious risk of bias for all. Estimates with heterogeneity ( $I^2 > 70$ ) were further downgraded to low certainty. <sup>†</sup>Refers to potassium-sparing diuretics and aldosterone antagonists. Abbreviations: ACEI = angiotensin-converting enzyme inhibitor, ARB = angiotensin receptor blocker, CCB = calcium channel blocker, HR = hazard ratio, MD = mean difference, NA = not applicable, OR = odds ratio.

#### Susceptibility to infection

Thirty-one studies reported count data and/or crude odds ratios (OR) for the association between ACEI/ARB exposure and susceptibility to infection (Figure S2). Seven studies were removed to minimize the inclusion of studies with overlapping data. The primary meta-analysis (24 studies) revealed that ACEIs/ARBs were associated with higher odds of testing positive for COVID-19 (pooled unadjusted OR 1.15, 95% CI 1.02 to 1.30, Figure 2). Heterogeneity was high (*I*^2^=93%) and could be explained by country (China versus other) and study design (multicentre vs single-centre, cohort/case series vs case-control) (Table S3). The pooled estimate was no longer statistically significant when analysis was restricted to only Chinese studies (*n* = 2), multicentre studies (*n* = 14), and case-control studies (*n* = 8) (Figure S2). The linear regression test of funnel plot asymmetry (Egger’s test, p = 0.89) was not significant (funnel plot in Figure S3). Six studies reported adjusted or propensity score– weighted odds ratios (pooled adjusted OR 1.01, 95% CI 0.93 to 1.10, *I*^2^=0%) while two studies reported adjusted hazards ratios (pooled adjusted HR 1.05, 95% CI 0.92 to 1.20, *I*^2^=0%) (Figure S2). Other cardiovascular drug classes included anticoagulants, antiplatelets, beta-blockers, calcium channel blockers, class III anti-arrhythmics (specifically amiodarone), diuretics and lipid modifying drugs. Except for beta-blockers and diuretics (unadjusted estimates), none of the other cardiovascular drug exposures (including ACEIs and ARBs assessed separately) were associated with susceptibility to infection as detailed in Table 1.

**Figure 2.**
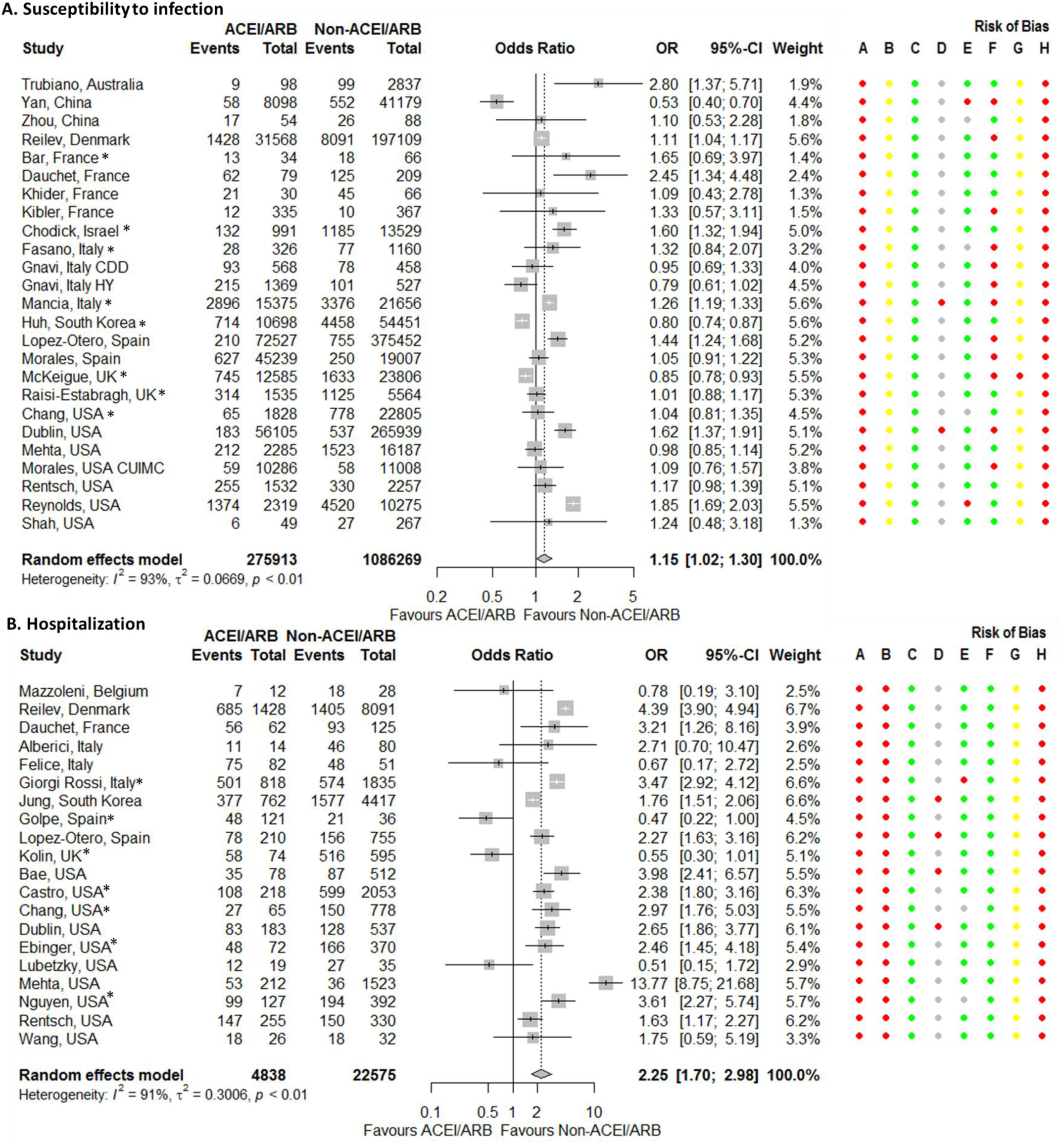
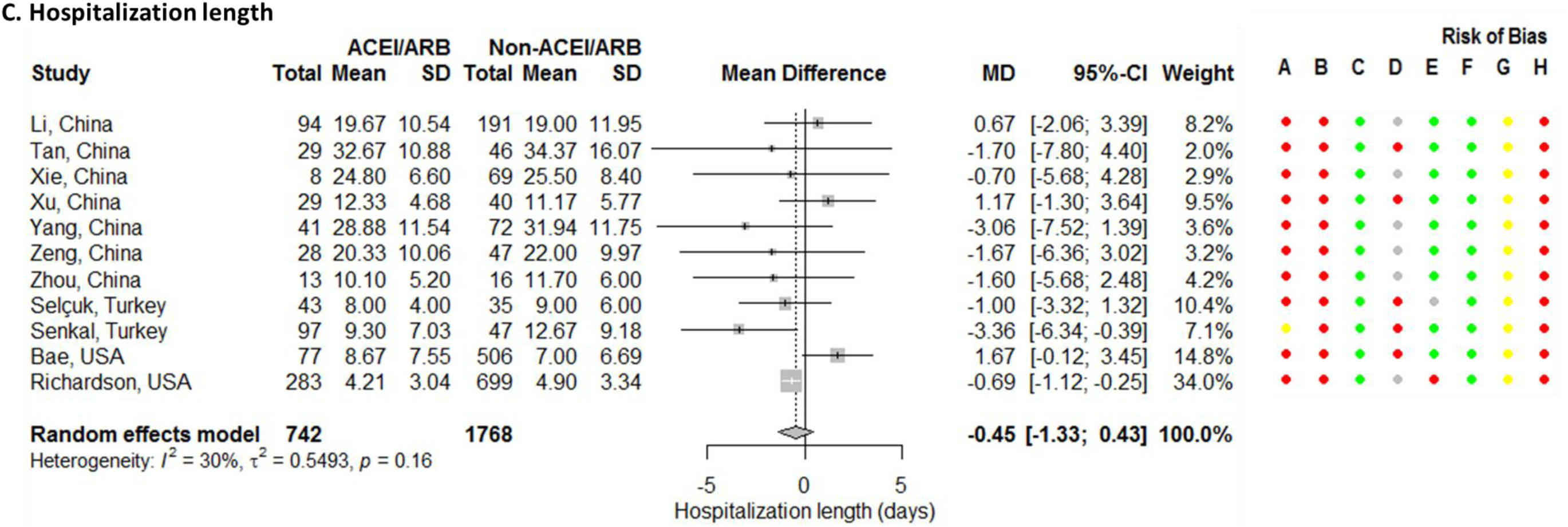
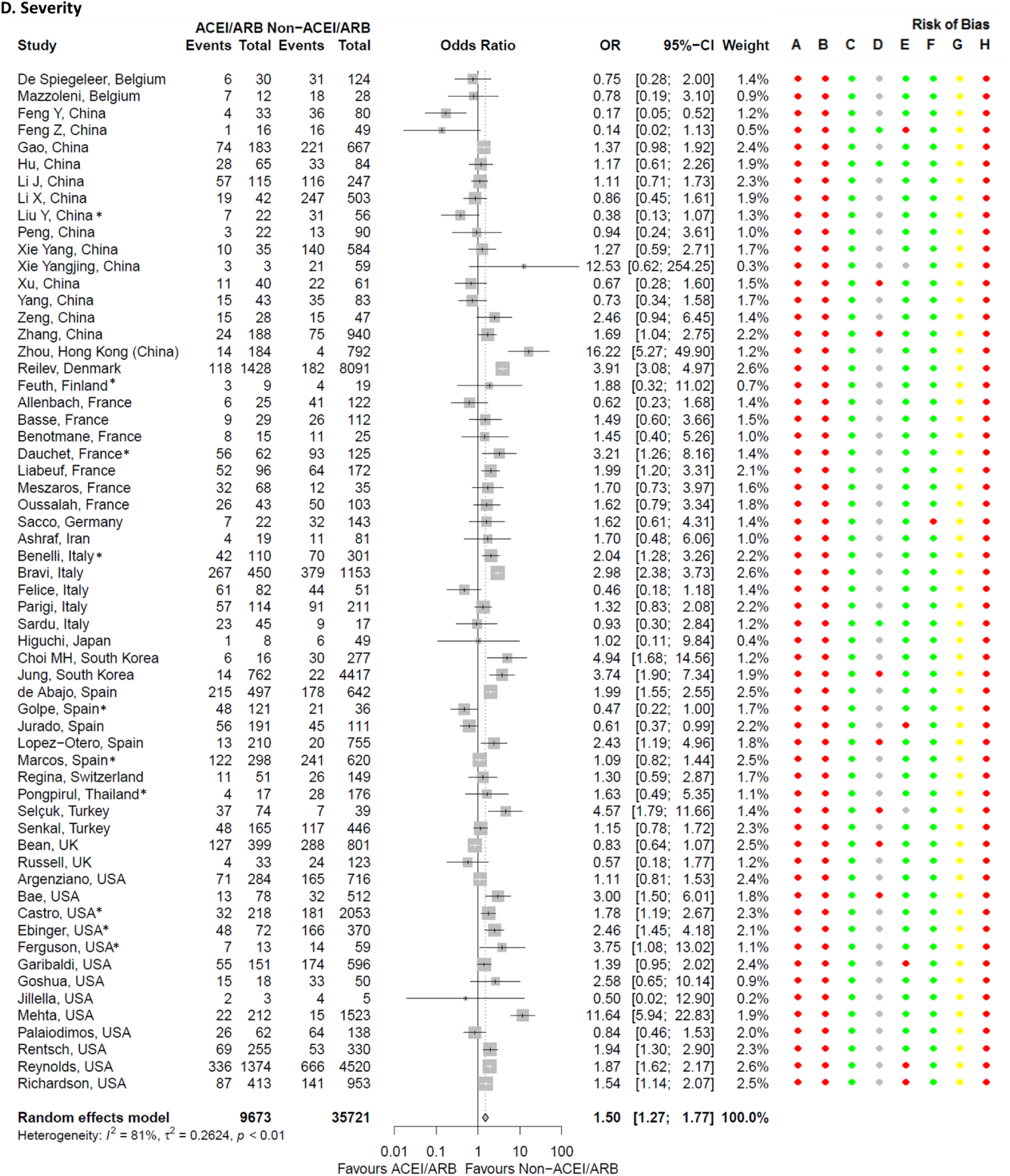
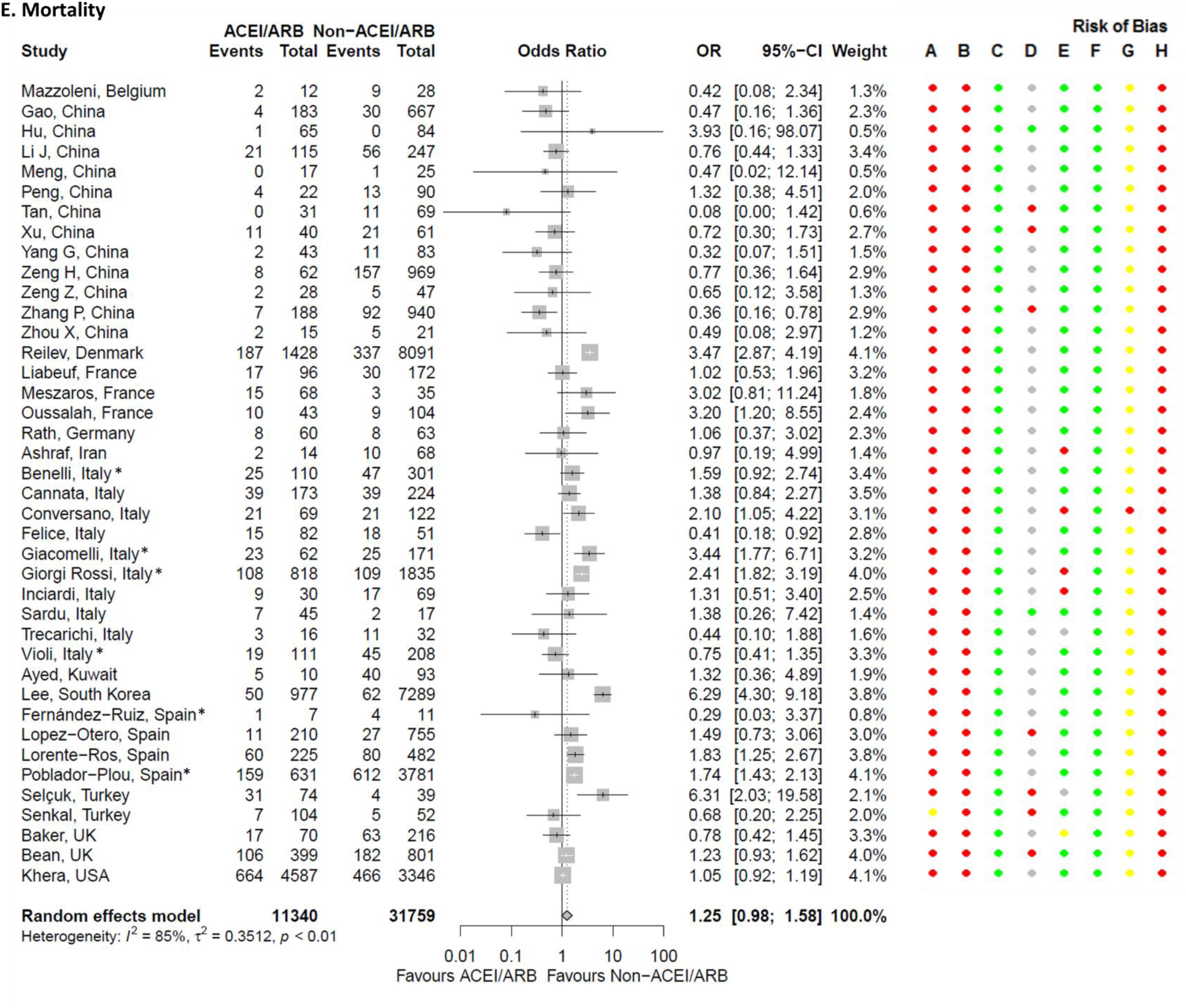
Forest plots for associations between COVID-19 outcomes and being on an angiotensin-converting enzyme inhibitor (ACEI) or angiotensin receptor blocker (ARB). Gnavi and Morales studies each provided two separate cohorts. *Estimates assume that none of the patients are taking both ACEIs and ARBs. Risk of bias legend. A = risk of bias due to confounding, B = risk of bias in selection of participants into the study, C = risk of bias in classification of interventions, D = risk of bias due to deviations from intended interventions, E = risk of bias due to missing data, F = risk of bias in measurement of outcomes, G = risk of bias in selection of the reported result, H = overall risk of bias. Color codes. Colour codes. Red = serious, yellow = moderate, green = low, grey = unclear. CDD = circulatory diseases/diabetes population, CUIMC = Columbia University Irving Medical Center, HY = hypertension population.

#### Hospitalization

Twenty-three studies explored the association between being hospitalized and being on ACEIs/ARBs (Figure S12). When three studies were excluded to reduce potentially overlapping data, ACEIs/ARBs were associated with higher odds of being hospitalized (pooled unadjusted OR 2.25, 95% CI 1.70 to 2.98, *I*^2^=91%, Figure 2). Heterogeneity could be explained by study-level mean/median ages and the proportion of patients with hypertension (Table S7). Four studies included only hypertensive patients and for these, the pooled estimate lost statistical significance (Figure 3). We did not conduct sub-group analyses based on age because no two studies included patients of a unique age category. Publication bias assessment for the 20 studies did not reveal funnel plot asymmetry (Figure S13, Egger’s test p-value = 0.13). The pooled adjusted odds ratio (6 studies) was not statistically significant at 1.16 (95% CI 0.80 to 1.68, *I*^2^=53%), a result which was similar to the pooled adjusted hazards ratio (1.00, 95% CI 0.87 to 1.14, *I*^2^=0%, 2 studies) (Figure S12). Other cardiovascular drugs were also associated with higher odds of being hospitalized in unadjusted, but not adjusted, estimates (Table 1).

**Figure 3.**
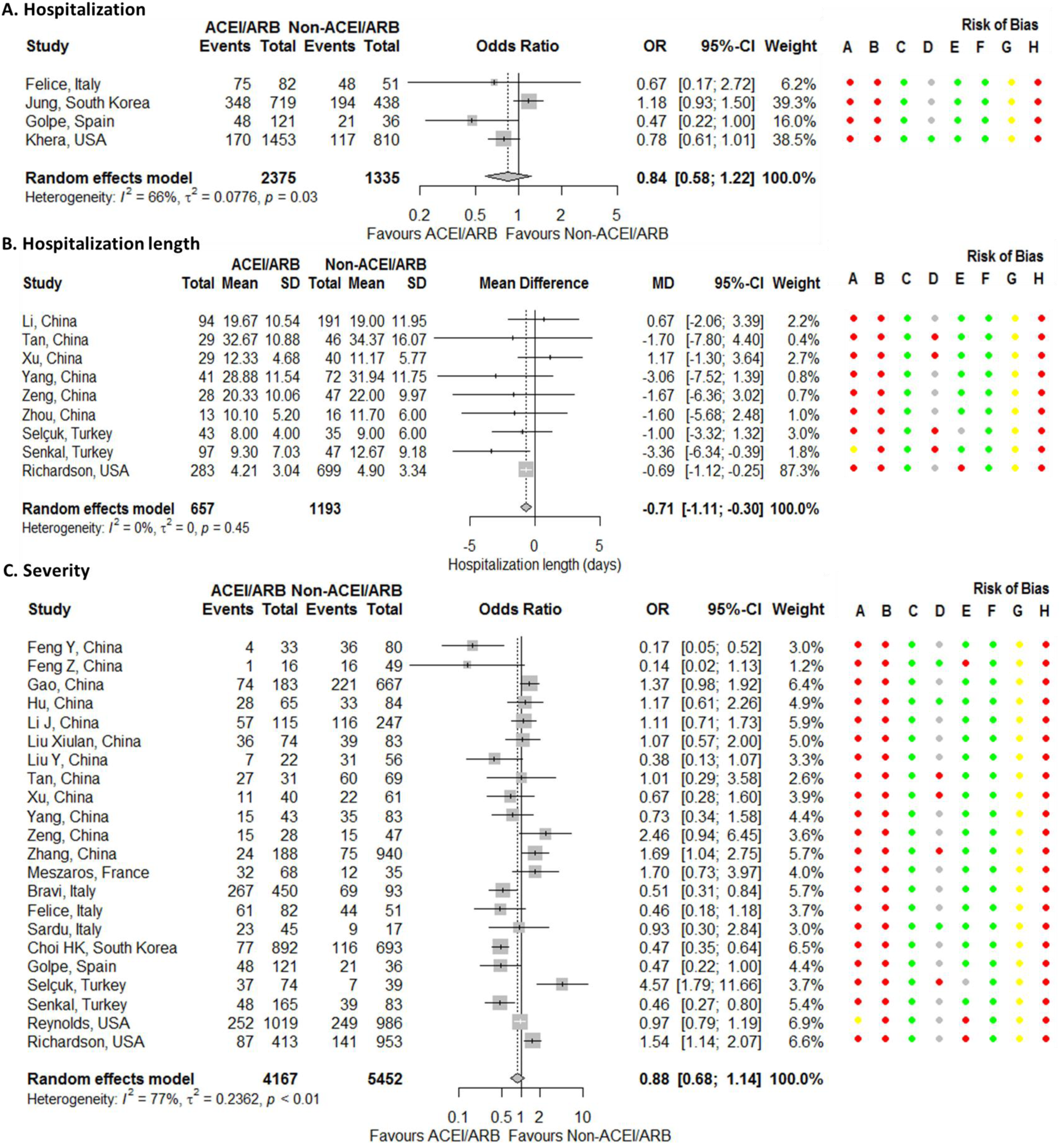
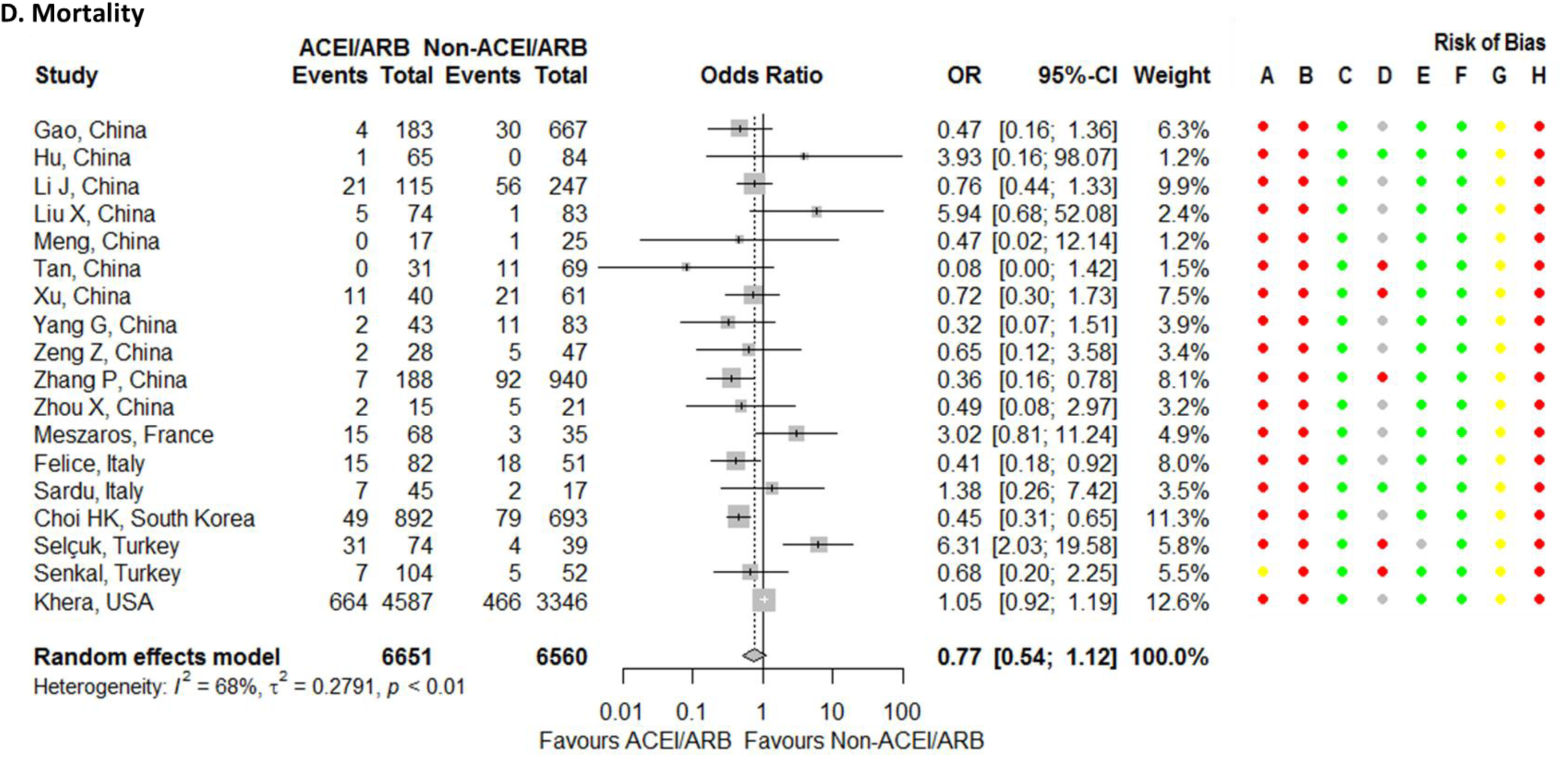
Forest plots for associations between COVID-19 outcomes and being on an angiotensin-converting enzyme inhibitor (ACEI) or angiotensin receptor blocker (ARB) – only hypertensive patients included. Risk of bias legend. A = risk of bias due to confounding, B = risk of bias in selection of participants into the study, C = risk of bias in classification of interventions, D = risk of bias due to deviations from intended interventions, E = risk of bias due to missing data, F = risk of bias in measurement of outcomes, G = risk of bias in selection of the reported result, H = overall risk of bias. Color codes. Colour codes. Red = serious, yellow = moderate, green = low, grey = unclear.

#### Hospitalization length

Twelve studies reported length of hospitalization (Figure S20) and for 11 of these without potentially overlapping datasets, ACEIs/ARBs were not significantly associated with longer hospitalization length (mean difference −0.45, 95% CI −1.33 to 0.43 days, *I*^2^=30%, Figure 2). When one study attributed 34% of the weight was excluded in a sensitivity analysis, the results were similar (mean difference −0.44, 95% CI −1.66 to 0.77 days, *I*^2^=32%). Heterogeneity was moderate and explainable by both age and proportion of patients with hypertension (Table S10). When 9 studies that included only hypertensive patients were pooled, ACEIs/ARBs were associated with shorter hospitalization length although this benefit was small. Egger’s test was not statistically significant (p = 0.94, funnel plot in Figure S21). This outcome was not assessed for other cardiovascular drug exposures.

#### Severity

Seventy-six studies reported the association between ACEIs/ARBs and severity outcomes (Figure S22). Sixteen studies were excluded due to having potentially overlapping data which resulted in a primary meta-analysis of 60 studies in which ACEIs/ARBs were associated with higher odds of severe disease (pooled OR 1.50, 95% CI 1.27 to 1.77, *I*^2^=81%, Figure 2). Heterogeneity could be explained by country, study setting, sample size, study-level mean/median ages, and the proportion of patients with hypertension and diabetes mellitus (Table S11). Sub-group analyses based on country (South Korea versus other), in-patient study setting, and the proportion of patients with diabetes mellitus arrived at similar conclusions while the pooled estimates were no longer significant when only Chinese studies or hypertensive cohorts were pooled (Figure 3, Figure S22). Publication bias assessment for the 60 studies revealed funnel plot symmetry (Figure S23, Egger’s test p = 0.15). Adjusted odds ratios were obtained from 18 studies (pooled adjusted OR 1.04, 95% CI 0.76 to 1.42, *I*^2^=65%) while hazard ratios were obtained from 4 studies (pooled adjusted HR 0.98, 95% CI 0.72 to 1.33, *I*^2^=68%) (Figure S22). Except for antiplatelet exposure, other cardiovascular drugs were associated with higher odds of severe disease in the unadjusted estimates, with statistical significance being lost when adjusted estimates were considered (Table 1).

#### All-cause mortality

Seventy-three studies reported the association between ACEI/ARB exposure and all-cause mortality (Figure S40). Because some studies had potentially overlapping datasets, only 40 were included in the primary meta-analysis with ACEIs/ARBs not being associated with higher odds of all-cause mortality (pooled OR 1.25, 95% CI 0.98 to 1.58, Figure 2). Heterogeneity was high (*I*^2^=85%) and could be accounted for by country, sample size, study-level mean/median ages, and the proportion of patients with hypertension (Table S19). The direction of the pooled estimate was reversed and was statistically significant in 12 China-only studies (Figure S40) whilst when restricting the analysis to hypertensive cohorts only (*n* = 18), the result remained non-significant (Figure 3). Egger’s test was not statistically significant (p = 0.20, funnel plot in Figure S41). The pooled adjusted odds ratio (13 studies) was 0.86 (95% CI 0.64 to 1.15, *I*^2^=4%) while the pooled adjusted hazards ratio (10 studies) was 0.94 (95% CI 0.79 to 1.13, *I*^2^=29%) (Figure S40). Except for anticoagulants, other cardiovascular drugs were associated with higher odds of all-cause mortality in the unadjusted estimates. Statistical significance was lost for ACEIs, ARBs, anticoagulants, calcium channel blockers, and lipid-modifying drugs when adjusted ORs were pooled. When adjusted hazards ratios were considered, ACEIs, ARBs, calcium channel blockers and diuretics were not associated with all-cause mortality while lipid-modifying drugs (statins) decreased the odds of dying (HR 0.67, 95% CI 0.47 to 0.96, *I*^2^=83%, Table 1).

## DISCUSSION

We have conducted a systematic review and meta-analysis to evaluate the current evidence on the influence of cardiovascular drugs on five COVID-19 clinical outcomes. The most reported drug classes were angiotensin-converting enzyme inhibitors (ACEIs) or angiotensin receptor blockers (ARBs) with ACEI/ARB exposure being associated with higher odds of testing positive for COVID-19, which contradicts a previous estimate by Zhang et al. (0.99, 95% CI, 0.95 to 1.04).^10^ Our study, which included 24 studies for this outcome, is however more comprehensive than Zhang et al.’s which included only 3 studies for the same outcome. Among COVID-19 patients, ACEI/ARB exposure was associated with being hospitalized, having severe disease but not hospitalization length or all-cause mortality. Diaz-Arocutipa et al.^14^ who explored these four outcomes reported similar results for hospitalization length (mean difference −0.96 days, 95%CI −2.50 to 0.57, *n* = 5 studies) and mortality (OR 1.11, 95%CI 0.77-1.60, *n* = 22 studies) but not for being hospitalized (OR 1.83, 95%CI 0.95-3.52, *n* = 4 studies) or having severe disease (OR 0.79, 95%CI 0.59-1.07, *n* = 18 studies) – differences we again attribute to our review being bigger in terms of the number of studies included. With a higher rate of hospitalization and more severe disease, one would respectively expect longer hospital stay and increased mortality, which makes our results seem counter intuitive. However, the length of hospitalization outcome excluded patients who died or those who were still hospitalized by the time of analysis, which may have contributed to the observed discrepancy. It is also important to note that these results are from pooling unadjusted estimates which did not account for confounding factors such as cardiovascular comorbidities. For instance, because hypertension might necessitate ACEI/ARB use, and hypertension contributes to poor COVID-19 clinical outcomes,^8 36-39^ estimates that do not adjust for hypertension might be spuriously elevated as seen above (an example of “confounding by indication”). Indeed, when subgroup analyses that included only hypertensive patients were conducted, ACEI/ARB exposure was no longer associated with being hospitalized or having severe disease. Lastly, co-interventions such as steroids and remdesivir that could influence these results could not be accounted for since studies rarely reported these co-interventions and stratified them by cardiovascular drug exposure.

We also reported pooled adjusted estimates in which ACEI/ARB exposure was not associated with higher odds of testing positive for COVID-19, being hospitalized, having severe disease, or all-cause mortality. Diaz-Arocutipa et al.^14^ reported similar estimates for all-cause mortality (adjusted hazards ratio 0.83, 95%CI 0.49 to 1.38, *n* = 3 studies) but not severity (adjusted OR 0.56, 95%CI 0.37 to 0.87, *n* = 4 studies). Although pooling adjusted estimates can protect against the effect of confounders present in unadjusted estimates, these pooled adjusted estimates should still be cautiously interpreted since many did not include adjustment for important confounders, and odds/hazard ratios that adjust for different sets of covariates may not be comparable.^47^ Further, adjusted odds/hazards ratios are expected to be further from zero (the “non-collapsibility” of effect estimates).^54^

Regarding other cardiovascular drug classes, this is the first review to be broad in this context (most previous reviews have focused solely on ACEIs/ARBs) with most other drugs not being associated with poor COVID-19 clinical outcomes in the pooled adjusted estimates. One key result is that lipid modifying drugs (statins in particular) appear to protect against all-cause mortality based on the adjusted hazards ratios, as has recently been reported.^55^ However, the number of included studies (*n* = 4) was small and the adjusted odds ratios were not statistically significant. The potential mechanisms in which cardiovascular drugs can influence COVID-19 outcomes have been previously discussed.^6-9 22- 25^

### Limitations of this review

For most of the meta-analyses, heterogeneity in effect estimates was high, which is similar to previous observations.^10 12-14 17 18 21^ Consequently, following GRADE rating,^52 53^ all estimates with high heterogeneity (*I*^2^ >70) were downgraded by one level (high to moderate certainty rating). Additionally, almost all estimates were ranked to be at a serious risk of bias. This differs from previous reviews in which many studies were considered to have low to moderate risk of bias,^10-18 20 21^ a contradiction we attribute to the risk of bias assessment tools used (whereas we used the ROBINS-I [Risk Of Bias In Non-randomised Studies – of Interventions] tool for non-randomized studies,^32^ other reviews used the Newcastle Ottawa Scale). Again following GRADE^52 53^ recommendations, the evidence certainty rating was downgraded by one level for estimates with a serious risk of bias (from high to moderate or from moderate to low). Despite our comprehensive search strategy and to facilitate timely publication, we did not contact study authors for data that was required to enable meta-analysis but not reported in the published paper, and therefore did not include studies that could potentially be eligible. We were also unable to include many studies from some preprint servers such as Authorea.com, Preprints from Lancet, and Preprints.org. We were however cognizant of the fact that this is a living systematic review and any studies that we excluded due to data needed for meta-analysis not being reported, or that we missed in our initial searches may be included in subsequent updates. We also included several preprint publications that have not been certified by peer review. This we felt necessary since many COVID-19 studies are being first published as preprints and we will be reassessing the preprints that will be peer-reviewed in future updates. We tried to exclude potentially overlapping data – however, we may have missed some overlapping data or inadvertently excluded non-overlapping data. The overall low contributions/assigned weights of the individual studies make the reported estimates robust to these errors. We also relied on single-reviewer extraction for half the studies, which could introduce bias from simple errors. Any new information/inconsistencies that come to our attention will be incorporated in subsequent reviews. Lastly, we could not explore the interplay of the various cardiovascular drugs because the number of studies for many drug classes was small. Once more, preferably high-quality, studies become available, we will compare how the different drug classes perform in combination and against each other (for example propensity score stratified HRs suggest that ACEIs/ARBs combined with other drugs are similar to calcium channel blockers/thiazides (HR 1.01, 95% CI 0.90 to 1.15) and ACEI combinations are more protective than ARB combinations (HR 0.88, 95% CI 0.79 to 0.99) in terms of susceptibility to infection^56^).

## Conclusions

Low-to moderate-certainty evidence suggests that cardiovascular drugs are not associated with poor COVID-19 clinical outcomes in high-risk patients such as those with hypertension. High quality evidence in the form of randomized controlled trials is urgently required and will be incorporated in updates of this review as soon as it is available (several trials are currently ongoing such as the 3 interventional phase III studies listed on ClinicalTrials.gov for Losartan [identifiers: NCT04343001, NCT04328012, NCT04349410], as of 30 August 2020). A better understanding of the interactions between cardiovascular drugs and COVID-19 pathology is required at cellular and molecular level to resolve the biological plausibility arguments that support both harm and benefit from ARB/ACEI exposure.^6-9^ As we await further evidence, patients on cardiovascular drugs should continue taking their medications as is recommended worldwide for ARBs/ACEIs.

## Data Availability

All relevant material is provided in the supplementary material.

## CONTRIBUTORS

MP conceived the study. All authors designed the study. IGA and SP screened titles and abstracts for inclusion. IGA, SP and RT collected the data. IGA analysed the data. IGA, SP and RT assessed the certainty of the evidence. IGA wrote the first draft, which all authors revised for critical content. All authors approved the final manuscript.

## PATIENT AND PUBLIC INVOLVEMENT

Patients or the public were not involved in the design, or conduct, or reporting, or dissemination plans of our research.

## ETHICAL APPROVAL

Not applicable. All the work was developed using published data.

## DATA SHARING

All relevant material is provided in the supplementary material.

## SUPPLEMENTARY MATERIALS

## References

1. Hui DS, E IA, Madani TA, et al. The continuing 2019-nCoV epidemic threat of novel coronaviruses to global health - The latest 2019 novel coronavirus outbreak in Wuhan, China. Int J Infect Dis 2020;91:264–66. doi: 10.1016/j.ijid.2020.01.009 [published Online First: 2020/01/19]

2. Hoffmann M, Kleine-Weber H, Schroeder S, et al. SARS-CoV-2 Cell Entry Depends on ACE2 and TMPRSS2 and Is Blocked by a Clinically Proven Protease Inhibitor. Cell 2020;181(2):271–80 e8. doi: 10.1016/j.cell.2020.02.052 [published Online First: 2020/03/07]

3. World Health Organisation. WHO Director-General’s opening remarks at the media briefing on COVID-19 - 11 March 2020 Online: World Health Organisation; 2020 [Available from: https://www.who.int/dg/speeches/detail/who-director-general-s-opening-remarks-at-the-media-briefing-on-covid-19---11-march-2020 accessed 6 June 2020.

4. Dong E, Du H, Gardner L. An interactive web-based dashboard to track COVID-19 in real time. Lancet Infect Dis 2020;20(5):533–34. doi: 10.1016/S1473-3099(20)30120-1 [published Online First: 2020/02/23]

5. Collaborators GBDCoD. Global, regional, and national age-sex-specific mortality for 282 causes of death in 195 countries and territories, 1980-2017: a systematic analysis for the Global Burden of Disease Study 2017. Lancet 2018;392(10159):1736–88. doi: 10.1016/S0140-6736(18)32203-7 [published Online First: 2018/11/30]

6. Clerkin KJ, Fried JA, Raikhelkar J, et al. COVID-19 and Cardiovascular Disease. Circulation 2020;141(20):1648–55. doi: 10.1161/CIRCULATIONAHA.120.046941 [published Online First: 2020/03/24]

7. Guzik TJ, Mohiddin SA, Dimarco A, et al. COVID-19 and the cardiovascular system: implications for risk assessment, diagnosis, and treatment options. Cardiovasc Res 2020 doi: 10.1093/cvr/cvaa106 [published Online First: 2020/05/01]

8. Zheng Z, Peng F, Xu B, et al. Risk factors of critical & mortal COVID-19 cases: A systematic literature review and meta-analysis. J Infect 2020 doi: 10.1016/j.jinf.2020.04.021 [published Online First: 2020/04/27]

9. Pranata R, Huang I, Lim MA, et al. Impact of Cerebrovascular and Cardiovascular Diseases on Mortality and Severity of COVID-19 - Systematic Review, Meta-analysis, and Meta-regression. J Stroke Cerebrovasc Dis 2020:104949. doi: 10.1016/j.jstrokecerebrovasdis.2020.104949 [published Online First: 2020/05/16]

10. Zhang X, Yu J, Pan LY, et al. ACEI/ARB use and risk of infection or severity or mortality of COVID-19: A systematic review and meta-analysis. Pharmacol Res 2020;158:104927. doi: 10.1016/j.phrs.2020.104927 [published Online First: 2020/05/19]

11. Flacco ME, Acuti Martellucci C, Bravi F, et al. Treatment with ACE inhibitors or ARBs and risk of severe/lethal COVID-19: a meta-analysis. Heart 2020 doi: 10.1136/heartjnl-2020-317336 [published Online First: 2020/07/03]

12. Grover A, Oberoi M. A systematic review and meta-analysis to evaluate the clinical outcomes in COVID-19 patients on angiotensin-converting enzyme inhibitors or angiotensin receptor blockers. Eur Heart J Cardiovasc Pharmacother 2020 doi: 10.1093/ehjcvp/pvaa064 [published Online First: 2020/06/17]

13. Pirola CJ, Sookoian S. Estimation of Renin-Angiotensin-Aldosterone-System (RAAS)-Inhibitor effect on COVID-19 outcome: A Meta-analysis. J Infect 2020 doi: 10.1016/j.jinf.2020.05.052 [published Online First: 2020/06/01]

14. Diaz-Arocutipa C, Saucedo-Chinchay J, Hernandez AV. Association Between ACEIs or ARBs Use and Clinical Outcomes in COVID-19 Patients: A Systematic Review and Meta-analysis. medRxiv 2020 doi: https://www.medrxiv.org/content/10.1101/2020.06.03.20120261v1 [published Online First: June 8]

15. Mackey K, King VJ, Gurley S, et al. Risks and Impact of Angiotensin-Converting Enzyme Inhibitors or Angiotensin-Receptor Blockers on SARS-CoV-2 Infection in Adults: A Living Systematic Review. Ann Intern Med 2020;173(3):195–203. doi: 10.7326/M20-1515 [published Online First: 2020/05/19]

16. Guo X, Zhu Y, Hong Y. Decreased Mortality of COVID-19 With Renin-Angiotensin-Aldosterone System Inhibitors Therapy in Patients With Hypertension: A Meta-Analysis. Hypertension 2020;76(2):e13–e14. doi: 10.1161/HYPERTENSIONAHA.120.15572 [published Online First: 2020/05/28]

17. Pranata R, Permana H, Huang I, et al. The use of renin angiotensin system inhibitor on mortality in patients with coronavirus disease 2019 (COVID-19): A systematic review and meta-analysis. Diabetes & metabolic syndrome 2020;14(5):983–90. doi: 10.1016/j.dsx.2020.06.047

18. Barochiner J, Martinez R. Use of inhibitors of the renin-angiotensin system in hypertensive patients and COVID-19 severity: A systematic review and meta-analysis. J Clin Pharm Ther 2020 doi: 10.1111/jcpt.13246 [published Online First: 2020/08/09]

19. Volpe M, Battistoni A. Systematic review of the role of renin-angiotensin system inhibitors in late studies on Covid-19: A new challenge overcome? Int J Cardiol 2020 doi: 10.1016/j.ijcard.2020.07.041 [published Online First: 2020/08/02]

20. Nunes JPL. Mortality and use of angiotensin converting enzyme inhibitors in Covid 19 disease – a systematic review. medRxiv 2020 doi: 10.1101/2020.05.29.20116483

21. Ssentongo A, Ssentongo P, Heilbrunn E, et al. Renin-angiotensin-aldosterone system inhibitors and mortality in patients with hypertension hospitalized for COVID19: systematic review &meta-analysis. medRxiv 2020 doi: 10.1101/2020.05.21.20107003

22. Muthuswamy B. COVID-19: Is it time to revisit the research on calcium channel drug targets? European Medical Journal Diabetes 2020 doi: 10.33590/emjdiabet/200608

23. Rodrigues-Diez RR, Tejera-Muñoz A, Marquez-Exposito L, et al. Statins: Could an old friend help in the fight against COVID-19? British journal of pharmacology 2020 doi: 10.1111/bph.15166

24. Sanchis-Gomar F, Perez-Quilis C, Favaloro EJ, et al. Statins and other drugs: Facing COVID-19 as a vascular disease. Pharmacological research 2020;159:105033. doi: 10.1016/j.phrs.2020.105033

25. Tang N, Bai H, Chen X, et al. Anticoagulant treatment is associated with decreased mortality in severe coronavirus disease 2019 patients with coagulopathy. J Thromb Haemost 2020;18(5):1094–99. doi: 10.1111/jth.14817 [published Online First: 2020/03/29]

26. Jpt H, Green S e. Cochrane Handbook for Systematic Reviews of Interventions Version 5.1.0 [updated March 2011]. In: The Cochrane Collaboration, ed. Online, 2011.

27. Elliott JH, Synnot A, Turner T, et al. Living systematic review: 1. Introduction-the why, what, when, and how. J Clin Epidemiol 2017;91:23–30. doi: 10.1016/j.jclinepi.2017.08.010 [published Online First: 2017/09/16]

28. Simmonds M, Salanti G, McKenzie J, et al. Living systematic reviews: 3. Statistical methods for updating meta-analyses. J Clin Epidemiol 2017;91:38–46. doi: 10.1016/j.jclinepi.2017.08.008 [published Online First: 2017/09/16]

29. Shamseer L, Moher D, Clarke M, et al. Preferred reporting items for systematic review and meta-analysis protocols (PRISMA-P) 2015: elaboration and explanation. BMJ 2015;350:g7647. doi: 10.1136/bmj.g7647

30. Joint Formulary Committee. British National Formulary 78 September 2019 – March 2020. 78 ed. London: BMJ Group and Pharmaceutical Press 2019.

31. Sterne JAC, Savovic J, Page MJ, et al. RoB 2: a revised tool for assessing risk of bias in randomised trials. BMJ 2019;366:l4898. doi: 10.1136/bmj.l4898 [published Online First: 2019/08/30]

32. Sterne JA, Hernan MA, Reeves BC, et al. ROBINS-I: a tool for assessing risk of bias in non-randomised studies of interventions. BMJ 2016;355:i4919. doi: 10.1136/bmj.i4919 [published Online First: 2016/10/14]

33. Emami A, Javanmardi F, Pirbonyeh N, et al. Prevalence of Underlying Diseases in Hospitalized Patients with COVID-19: a Systematic Review and Meta-Analysis. Arch Acad Emerg Med 2020;8(1):e35. [published Online First: 2020/04/02]

34. Parohan M, Yaghoubi S, Seraji A, et al. Risk factors for mortality in patients with Coronavirus disease 2019 (COVID-19) infection: a systematic review and meta-analysis of observational studies. Aging Male 2020:1–9. doi: 10.1080/13685538.2020.1774748 [published Online First: 2020/06/09]

35. Roncon L, Zuin M, Rigatelli G, et al. Diabetic patients with COVID-19 infection are at higher risk of ICU admission and poor short-term outcome. J Clin Virol 2020;127:104354. doi: 10.1016/j.jcv.2020.104354 [published Online First: 2020/04/20]

36. Singh AK, Gupta R, Misra A. Comorbidities in COVID-19: Outcomes in hypertensive cohort and controversies with renin angiotensin system blockers. Diabetes Metab Syndr 2020;14(4):283–87. doi: 10.1016/j.dsx.2020.03.016 [published Online First: 2020/04/14]

37. Tian W, Jiang W, Yao J, et al. Predictors of mortality in hospitalized COVID-19 patients: A systematic review and meta-analysis. J Med Virol 2020 doi: 10.1002/jmv.26050 [published Online First: 2020/05/23]

38. Wang B, Li R, Lu Z, et al. Does comorbidity increase the risk of patients with COVID-19: evidence from meta-analysis. Aging (Albany NY) 2020;12(7):6049–57. doi: 10.18632/aging.103000 [published Online First: 2020/04/09]

39. Yang J, Zheng Y, Gou X, et al. Prevalence of comorbidities and its effects in patients infected with SARS-CoV-2: a systematic review and meta-analysis. Int J Infect Dis 2020;94:91–95. doi: 10.1016/j.ijid.2020.03.017 [published Online First: 2020/03/17]

40. Tamara A, Tahapary DL. Obesity as a predictor for a poor prognosis of COVID-19: A systematic review. Diabetes Metab Syndr 2020;14(4):655–59. doi: 10.1016/j.dsx.2020.05.020 [published Online First: 2020/05/22]

41. Pan D, Sze S, Minhas JS, et al. The impact of ethnicity on clinical outcomes in COVID-19: A systematic review. EClinicalMedicine 2020;23:100404. doi: 10.1016/j.eclinm.2020.100404 [published Online First: 2020/07/08]

42. Guo FR. Active smoking is associated with severity of coronavirus disease 2019 (COVID-19): An update of a meta-analysis. Tob Induc Dis 2020;18:37. doi: 10.18332/tid/121915 [published Online First: 2020/05/10]

43. Vardavas CI, Nikitara K. COVID-19 and smoking: A systematic review of the evidence. Tob Induc Dis 2020;18:20. doi: 10.18332/tid/119324 [published Online First: 2020/03/25]

44. Mantovani A, Byrne CD, Zheng MH, et al. Diabetes as a risk factor for greater COVID-19 severity and in-hospital death: A meta-analysis of observational studies. Nutr Metab Cardiovasc Dis 2020;30(8):1236–48. doi: 10.1016/j.numecd.2020.05.014 [published Online First: 2020/06/24]

45. R: A language and environment for statistical computing. [program]. Vienna: R Foundation for Statistical Computing, 2019.

46. Schwarzer G. meta: An R package for meta-analysis. R News 2007;7(3):40–45.

47. Chang BH, Hoaglin DC. Meta-Analysis of Odds Ratios: Current Good Practices. Med Care 2017;55(4):328–35. doi: 10.1097/MLR.0000000000000696 [published Online First: 2017/02/09]

48. Wan X, Wang W, Liu J, et al. Estimating the sample mean and standard deviation from the sample size, median, range and/or interquartile range. BMC Med Res Methodol 2014;14:135. doi: 10.1186/1471-2288-14-135 [published Online First: 2014/12/20]

49. Higgins J, Green Se. Cochrane Handbook for Systematic Reviews of Interventions Version 5.1.0 [updated March 2011]. The Cochrane Collaboration, 2011. 2011.

50. Higgins JP, Thompson SG. Quantifying heterogeneity in a meta-analysis. Stat Med 2002;21(11):1539–58. doi: 10.1002/sim.1186

51. Viechtbauer W. Conducting meta-analyses in R with the metafor package. Journal of Statistical Software 2010;36(3):1–48.

52. Guyatt G, Oxman AD, Akl EA, et al. GRADE guidelines: 1. Introduction-GRADE evidence profiles and summary of findings tables. J Clin Epidemiol 2011;64(4):383–94. doi: 10.1016/j.jclinepi.2010.04.026

53. Guyatt GH, Oxman AD, Vist GE, et al. GRADE: an emerging consensus on rating quality of evidence and strength of recommendations. BMJ 2008;336(7650):924–6. doi: 10.1136/bmj.39489.470347.AD [published Online First: 2008/04/26]

54. Greenland S, Robins JM, Pearl J. Confounding and Collapsibility in Causal Inference. Statistical science 1999;14:29–46. doi: 10.1214/ss/1009211805

55. Kow CS, Hasan SS. Meta-analysis of Effectiveness of Statins in Patients with Severe COVID-19. The American Journal of Cardiology 2020 doi: 10.1016/j.amjcard.2020.08.004 [published Online First: 12 August]

56. Morales DR, Conover MM, You SC, et al. Renin-angiotensin system blockers and susceptibility to COVID-19: a multinational open science cohort study. MedRxiv 2020 doi: 10.1101/2020.06.11.20125849 [published Online First: 12 June]

