## Supplementary Methods, Figures and Tables for "Cardiovascular drugs and COVID-19 clinical outcomes: a living systematic review and meta-analysis"

### Table of contents

**Supplementary Text……………………………………………………………………………………2**

**Supplementary Tables…………………………………………………………………………………..7**

**Supplementary Figures………………………………………………………………………………….47**

**Supplementary References……………………………………………………………………………127**

### Supplementary Text

#### Text S1. Search strategies

DISCOVER

(coronavirus* OR corona-virus* OR 2019-nCoV* OR (2019 AND nCoV*) OR Covid* OR (severe AND acute AND respiratory AND syndrome*) OR nCoV* OR SARS-CoV* OR SARSCoV*) AND (cardiovascular* OR (angiotensin-converting AND enzyme AND inhibitor*) OR ACEI* OR (angiotensin AND receptor AND blocker*) OR (Angiotensin AND II AND receptor AND blocker*) OR ARB* OR (renin AND angiotensin AND aldosterone AND system AND inhibitor*) OR RAAI* OR antihypertensive* OR anti-hypertensive* OR diuretic* OR inotrop* OR anti-arrhythmic* OR antiarrhythmic* OR beta-adrenoceptor* OR beta-blocker* OR (beta AND blocker*) OR beta-adrenergic* OR *blocker* OR (RAS AND blocker*) OR anti-hypertensive* OR alpha-adrenoceptor* OR alpha-blocker* OR anti-anginal* OR antianginal* OR sympathomimetic* OR anticoagulant* OR anti-coagulant* OR antiplatelet* OR anti-platelet* OR fibrinolytic* OR (calcium AND channel AND blocker*) OR thrombolytic* OR lipid-lowering* OR *statin* OR *sartan OR *pril)

MEDLINE

((coronavirus$ or corona-virus$ or 2019-nCoV$ or 2019 nCoV$ or Covid$ or severe acute respiratory syndrome$ or nCoV$ OR SARS-CoV$ or SARSCoV$) and (cardiovascular$ or angiotensin-converting enzyme inhibitor$ or ACEI$ or angiotensin receptor blocker$ or Angiotensin II receptor blocker$ or ARB$ or renin angiotensin aldosterone system inhibitor$ or RAAI$ or antihypertensive$ or anti-hypertensive$ or diuretic$ or inotrop$ or anti-arrhythmic$ or antiarrhythmic$ or beta-adrenoceptor$ or beta-blocker$ or beta blocker$ or beta-adrenergic$ or $blocker$ or RAS blocker$ or anti-hypertensive$ or alpha-adrenoceptor$ or alpha-blocker$ or anti-anginal$ or antianginal$ or sympathomimetic$ or anticoagulant$ or anti-coagulant$ or antiplatelet$ or anti-platelet$ or fibrinolytic$ or calcium channel blocker$ or thrombolytic$ or lipid-lowering$ or statin$ or $sartan or $pril)).mp

medRxiv and bioRxiv PREPRINTS

All 'COVID-19 SARS-CoV-2 preprints from medRxiv and bioRxiv' (https://connect.medrxiv.org/relate/content/181) were scraped using the rvest package[1] in in R version 3.6.1.[2]

**Text S2. Severity definitions**

Many studies reported the four common classifications (mild, moderate, severe, and critical) based on societal, national or World Health Organization guidelines. For example, Xiong[3] reports the following categories:

a) mild ("*patients who had mild clinical symptoms without manifestation of viral pneumonia on chest CT scans*.")
b) Moderate ("*patients who had symptoms such as fever and respiratory tract symptoms, etc., with manifestation of viral pneumonia on chest CT scans*.")
c) Severe ("*adults who met any of the following criteria: (1) respiratory rate ≥30 breaths/min; (2) oxygen saturation ≤93% at rest state; and (3) arterial PO2/oxygen concentration ≤300 mm Hg. Patients with pulmonary lesion progression >50% within 24–48 hours on radiologic imaging were treated as severe cases*.")
d) Critical ("*patients that met any of the following criteria: (1) occurrence of respiratory failure requiring mechanical ventilation; (2) presence of shock; and (3) other organ failure that requires monitoring and treatment in the intensive care unit.*")

The four commonly reported severity classifications were dichotomized into not severe (for 'mild' and 'moderate' classifications) and severe (for 'severe' and 'critical classifications). Where these were not reported, severe cases were considered as those with acute respiratory disease syndrome, or those being taken to intensive care units and/or requiring oxygen/intubation/any form of ventilation/continuous renal-replacement therapy. Because many studies also admitted mild/moderate patients, hospitalization was not considered to be severe illness unless specifically pointed out e.g. by Ebinger[4] and Dauchet.[5] Also, because not all severe patients die, mortality alone was not considered severity unless it was part of a composite outcome that represented severity. If two different outcomes e.g. ICU and intubation were separately reported and it was neither reported that one was a subset of the other or the two were mutually exclusive, then the one experienced by most patients was chosen.

**Text S3. Data extraction strategies**

Overlapping/potentially overlapping datasets

Where two or more studies used the same dataset for a given exposure-outcome combination, then peer-reviewed publications and those reporting a larger number of patients were preferred. In instances where it was not obvious if the included patients were the same but there was a possibility of overlap (e.g. studies using the same biobank/national database or recruiting from the same hospital with overlapping recruitment periods), then such studies were 'clustered' together. For the primary meta-analyses, only one of these studies (the one with the largest sample size) was included. In some cases, however, a peer-reviewed study/study with the largest sample size could be excluded if:

a) Its exclusion could allow the inclusion of several smaller non-overlapping studies (whose combined sample size would be larger than the one study).
b) The peer-reviewed/larger study had very restrictive criteria e.g. reporting on only patients with a specific and relatively rare condition or on a narrow age-group.
c) For susceptibility to infection, all included patients were not tested for COVID-19 and yet a sub-study included only tested patients.

For transparency, pooled estimates in which all studies, regardless of any overlapping, were included are also reported.

ACEI/ARB exposure

Although some studies that assessed ACEIs and ARBs reported a combined (ACEIs/ARBs) estimate, others did not. During the evaluation of the ACEI/ARB exposure, three options for the latter studies included:

a) Exclusion
b) Including the drug class (ACEI vs ARB) that had the highest number of patients
c) Including both drug classes

Options a) and b) were ignored since both were wasteful in terms of not maximizing available data. For option c) however, an assumption had to be made that no patients were taking both drug classes. Indeed, combinations of ARBs and ACEIs are generally not recommended.[6] Moreover, studies reporting combinations such as Sacubitril/valsartan counted it once (as an ARB, in this case) (e.g. Fosbol,[7] Grasselli,[8] Schneeweiss[9]). In the studies that reported these drug classes both separately and when combined, the majority (e.g. Ashraf,[10] Benotmane,[11] Feng,[12] Gao,[13] Garibaldi,[14] Jung,[15] Oussalah,[16] Richardson,[17] Sardu[18], Tan[19]) had zero overlap (no patients taking both drugs) while it was low for others [e.g. Bravi[20] (29/450, 6.4%), Cariou[21] (6/737, 0.8%), Fosbol[7] (12/895, 1.3%), Jung[22]

(12/762, 2.0%) Li[23] (3/115, 2.6%0, Lopez-Otero[24] (2175/72527, 3.0%), Mehta[25] (2/212, 0.9%), Rentsch[26] (8/255, 3.1%) Reynolds[27] (31/1374, 2.3%), Senkal[28] (2/165, 1.2%) and Xu[29] (1/40, 2.5%)]. Despite likely having a low impact on the pooled estimates, the impact of this assumption on the pooled estimates was also checked by examining the respective forest plots (to check the weights accorded to these studies and whether their individual estimates were consistent with other study estimates). Study authors will also be contacted to verify the overlap.

**Text S4. Simple meta-regression**

To investigate heterogeneity and inform sub-group analysis, we considered study design (multicentre vs single-centre, cohort/case series vs case-control), sample size, clinical setting (included only inpatients), participant age (study-level median/mean), gender (study-level proportions), most reported comorbidities (hypertension and diabetes, study-level proportions), study location (given country vs other), and publication status (peer-reviewed journal vs preprint server) as covariates to a simple meta regression. For susceptibility to infection, we also considered the extent of testing (all controls tested vs some controls not tested). We planned to explore race (study-level proportions) but this was infrequently reported so we used study location (already considered) as a proxy. We also could not include severity definitions as a covariate in meta-regression because they were too heterogeneous. We set the number of studies per covariate parameter to ten[30] which implied that we could only conduct meta-regressions for drug exposure-outcome combinations that included at least 10 studies. When data was missing, mean imputation (i.e. estimates for studies with missing data were estimated as the mean measures across all other studies) was used. A p-value threshold of 0.05 was used. The simple meta-regression was implemented using the metareg function in the R meta package[31]).

### Supplementary Tables

#### Table S1. Preferred Reporting Items for Systematic Reviews and Meta-Analyses: The PRISMA Statement[32]

| Section and topic | Item No | Checklist item | Section and topic |
|---|---|---|---|
| TITLE | | | |
| Title | 1 | Identify the report as a systematic review, meta-analysis, or both. | Title page |
| ABSTRACT | | | |
| Structured summary | 2 | Provide a structured summary including, as applicable: background; objectives; data sources; study eligibility criteria, participants, and interventions; study appraisal and synthesis methods; results; limitations; conclusions and implications of key findings; systematic review registration number. | Abstract |
| INTRODUCTION | | | |
| Rationale | 3 | Describe the rationale for the review in the context of what is already known. | Introduction, paragraphs 1, 2 and 3 |
| Objectives | 4 | Provide an explicit statement of questions being addressed with reference to participants, interventions, comparisons, outcomes, and study design (PICOS). | Introduction, paragraph 3 |
| METHODS | | | |
| Protocol and registration | 5 | Indicate if a review protocol exists, if and where it can be accessed (e.g., Web address), and, if available, provide registration information including registration number. | Methods, paragraph 1 |
| Eligibility criteria | 6 | Specify study characteristics (e.g., PICOS, length of follow-up) and report characteristics (e.g., years considered, language, publication status) used as criteria for eligibility, giving rationale. | Methods (selection criteria) |
| Information sources | 7 | Describe all information sources (e.g., databases with dates of coverage, contact with study authors to identify additional studies) in the search and date last searched. | Methods (identification of studies) |
| Search | 8 | Present full electronic search strategy for at least one database, including any limits used, such that it could be repeated. | Text S1 |
| Study selection | 9 | State the process for selecting studies (i.e., screening, eligibility, included in systematic review, and, if applicable, included in the meta-analysis). | Methods (data extraction) |
| Data collection process | 10 | Describe method of data extraction from reports (e.g., piloted forms, independently, in duplicate) and any processes for obtaining and confirming data from investigators. | Methods (data extraction) |
| Data items | 11 | List and define all variables for which data were sought (e.g., PICOS, funding sources) and any assumptions and simplifications made. | Methods (data extraction) |
| Risk of bias in individual studies | 12 | Describe methods used for assessing risk of bias of individual studies (including specification of whether this was done at the study or outcome level), and how this information is to be used in any data synthesis. | Methods (assessment of study quality) |
| Summary measures | 13 | State the principal summary measures (e.g., risk ratio, difference in means). | Methods (data synthesis) |
| Synthesis of results | 14 | Describe the methods of handling data and combining results of studies, if done, including measures of consistency (e.g., $I^2$) for each meta-analysis. | Methods (data synthesis, heterogeneity measures) |

**Table S1. Continued**

| Section and topic | Item No | Checklist item | Section and topic |
|---|---|---|---|
| Risk of bias across studies | 15 | Specify any assessment of risk of bias that may affect the cumulative evidence (e.g., publication bias, selective reporting within studies). | Methods (publication bias) |
| Additional analyses | 16 | Describe methods of additional analyses (e.g., sensitivity or subgroup analyses, meta-regression), if done, indicating which were pre-specified. | Methods (sensitivity and subgroup analyses) |
| RESULTS | | | |
| Study selection | 17 | Give numbers of studies screened, assessed for eligibility, and included in the review, with reasons for exclusions at each stage, ideally with a flow diagram. ` | Figure 1 |
| Study characteristics | 18 | For each study, present characteristics for which data were extracted (e.g., study size, PICOS, follow-up period) and provide the citations. | Table S2 |
| Risk of bias within studies | 19 | Present data on risk of bias of each study and, if available, any outcome level assessment (see item 12). | Figure S1, all forest plots |
| Results of individual studies | 20 | For all outcomes considered (benefits or harms), present, for each study: (a) simple summary data for each intervention group (b) effect estimates and confidence intervals, ideally with a forest plot. | Table 1, all forest plots |
| Synthesis of results | 21 | Present results of each meta-analysis done, including confidence intervals and measures of consistency. | Table 1, all forest plots |
| Risk of bias across studies | 22 | Present results of any assessment of risk of bias across studies (see Item 15). | Table 1, all funnel plots |
| Additional analysis | 23 | Give results of additional analyses, if done (e.g., sensitivity or subgroup analyses, meta-regression [see Item 16]). | Tables 1, S3-S27, some forest plots |
| DISCUSSION | | | |
| Summary of evidence | 24 | Summarize the main findings including the strength of evidence for each main outcome; consider their relevance to key groups (e.g., healthcare providers, users, and policy makers). | Discussion, paragraph one |
| Limitations | 25 | Discuss limitations at study and outcome level (e.g., risk of bias), and at review-level (e.g., incomplete retrieval of identified research, reporting bias). | Discussion (limitations of this review) |
| Conclusions | 26 | Provide a general interpretation of the results in the context of other evidence, and implications for future research. | Discussion (conclusions) |
| FUNDING | | | |
| Funding | 27 | Describe sources of funding for the systematic review and other support (e.g., supply of data); role of funders for the systematic review. | Funder section. |

### Table S2. Studies included in the systematic review

| # | Cluster/ clustered with | First Author | Peer-reviewed | First published or posted, 2020 | Country | Design | Recruitment, 2020 | Data source | Sample size | Eligibility | Race | Age, years | Male % | HTN % | DM % | Obesity % | Drug classes | Outcomes (definition for severity, if any) |
|---|---|---|---|---|---|---|---|---|---|---|---|---|---|---|---|---|---|---|
| 1 | Other trials from 38 countries | Jung[15] | Yes | 9-Jul | 38 countries | Multicenter RC (calls it prospective) | Upto 7 May | NI | 324 | ICU COVID-19 patients ≥ 70 years | NI | 75 (70–93) | 69 | 65 | 29 | NI | ACEi/ARB | 30-day mortality |
| 2 | NA | Trubiano[33] | No | 2-Jul | Australia | Single-center RC (call it prospective) | 11 Mar to 22 Apr | EMR | 2935 | COVID-19 tested patients | NI | 39 (29–53) | 36 | 9 | 3 | NI | ACEI/ARB | Susceptibility to infection |
| 3 | NA | De Spiegeleer[34] | Yes | 15 Jun (on medrxiv 15 May) | Belgium | Multicentre RC | 1 Mar to 16 Apr | EHR | 154 | COVID-19 patients | NI | 86 ± 7 | 33 | 25 | 18 | NI | ACEIS, ARBs, LMDs | Severity (long-stay hospital admission or death), 14-day mortality |
| 4 | NA | Mazzoleni[35] | Yes | 24-Jul | Belgium | Single-center case series | 6 Mar to 14 Apr | Clinical medical records | 40 | COVID-19 patients with kidney replacement therapy | NI | 75 (68–83) | 58 | 93 | 65 | NI | ACEI/ARB | Hospitalization or severity (based on the COVID-19 severity index), mortality |
| 5 | Seventh Hospital | Chen M[36] | No | 27-Feb | China | Single-centre RC | 1 Jan 1 to 15 Feb | EHR | 123 | COVID-19 patients | NI | 58 ± 15 | 50 | 33 | 11 | NI | ACEIs, ARBs | Mortality |
| 6 | Central Hospital | Chen Y[37] | Yes | 14-May | China | Single-centre RC | 1 Jan to 17 Mar | EHR | 71 (subset) | COVID-19 patients, HTN, and DM | NI | 67 (61–76) | NR | 100 | 100 | NI | ACEIS, ARBs | Hospitalization, mortality |
| 7 | NA | Feng Y[12] | Yes | 1-Jun | China | Multicentre RC | Jan 1 to 15 Feb | Medical record review | 113 (subset) | COVID-19 patients | NI | 53 (40–64) for 476 | 57 for 476 | 100 | NI | NI | ACEIS, ARBs, other antihypertensives | Severity (critical/severe vs moderate - based on national guidelines) |
| 8 | NA | Feng Z[38] | No | 10-Apr | China | Multicentre RC | 17 Jan to 28 Feb | EHR | 65 (subset) | Consecutive hospitalized adult COVID-19 patients | NI | 57 (51–65) for 16 and 63 (53–69) for 49 | 51 | 100 | 31 | NI | ACEIs, ARBs | Severity (critical/severe vs mild/moderate - based on symptoms and the need for ICU/ventilation) |
| 9 | NA | Gao[13] | Yes | 4-Jun | China | Single-centre RC | 5 Feb to 15 Mar | EHR | 850 | Consecutive hospitalized patients with confirmed COVID-19 | NI | 64 ± 11 | 52 | 100 | 28 | NI | ACEIS, ARBs, antihypertensives | Severity (mild vs severe/critical - based on clinical symptoms and ventilation), mortality |
| 10 | Seventh Hospital | Guo[39] | Yes | 27-Mar | China | Single-center retrospective case series | 23 Jan to 23 Feb | EHR | 187 | COVID-19 patients | NI | 59 ± 15 | 49 | 33 | 15 | NI | ACEIS, ARBs | Mortality |
| 11 | Zhejiang | Hu[40] | Yes | 8-Jun | China | Multicentre RC | 17 Jan to 8 Feb | NI | 149 (subset) | COVID-19 patients with HTN | NI | 57 (50–66) | 59 | 100 | 20 | NI | ACEIS, ARBs | Severity (mild vs severe/critical - based on WHO guidance, ICU admission, mechanical ventilation), mortality |

| | | | | | | | | | | | | | | | | | |
|---|---|---|---|---|---|---|---|---|---|---|---|---|---|---|---|---|---|
| 12 | Renmin Hospital | Huang[41] | Yes | 30-Mar | China | Single-centre RC | 7 Feb to 3 Mar | EHR | 50 | Hospitalized COVID-19 patients with HTN | NI | 62 ± 15 | 54 | 100 | 8 | NI | ACEIS, ARBs | Severity (mild/moderate vs severe/critically ill - based on national guidelines, high flow oxygen or non-invasive ventilation, invasive ventilation), mortality |
| 13 | Zhejiang | Jiang[42] | Yes | 23-Jun | China | Multicenter RC | 19 Jan to 20 Feb | EMR | 131 | Hospitalized adult (≥18 years) COVID-19 patients | NI | 51 ± 16 | 53 | NI | NI | NI | Statins | Severity (mild/moderate/severe vs critically ill - based on national guidelines) |
| 14 | Central Hospital | Li J[23] | Yes | 23-Apr | China | Single-centre retrospective case series | 15 Jan to 15 Mar | Medical records | 362 (subset) | COVID-19 patients with HTN | NI | 66 (59–73) | 52 | 100 | 35 | NI | ACEIS, ARBs, CCBs, β-blockers | Hospitalization, severity (non-severe vs severe - based on national guidelines), mortality |
| 15 | Sino-French New City Branch of Tongji Hosp | Li T[43] | Yes | 15-Jul | China | Single-centre RC | 1 Feb to 31 Mar | EMR | 312 | Hospitalized COVID-19 patients ≥ 65 years | NI | 69 ± 7 | 60 | 57 | 39 | NI | ACEi | Severity (non-severe vs severe - based on societal guidelines) |
| 16 | Sino-French New City Branch of Tongji Hosp | Li X[44] | Yes | 12-Apr | China | Single-centre ambispective cohort | 26 Jan to 5 Feb | EHR | 545/7 | Consecutive hospitalized COVID-19 patients | NI | 60 (48–69) [for 548] | 51 [for 548] | 30 | 15 | NI | ACE/ARBs, anticoagulants | Severity (according to societal guidelines) |
| 17 | NA | Li Y[45] | Yes | 2-Jul | China | Single-centre retrospective case series | 16 Jan to 19 Feb | EMR | 11 | Hospitalized COVID-19 patients with new onset of CVD | NI | 75 (range 57–91) | 55 | 82 | 55 | NI | Antiplatelet | Severity (severe vs non-severe - based on societal guidelines), mortality |
| 18 | Shenzhen Third People's Hospital | Liu Y[46] | No | 27-Mar | China | Multicentre RC | 27 Dec 2019 to 29 Feb | Medical records | 78 | Adult COVID-19 patients with HTN | NI | 65.2 ± 10.7 | 55 | 100 | NI | NI | ACEIS, ARBs, β-blockers, CCB, diuretics | Severity (severe vs mild - based on national guidelines) |
| 19 | Renmin Hospital | Liu Xiaofan[47] | Yes | 16-Apr | China | Single-center RC | 5 Feb to 14 Mar | EMR | 34 | Consecutive hospitalized COVID-19 patients | NI | NI | NI | 100 | NI | NI | ACEi/ARB | Severity (moderate vs severe pneumonia) |
| 20 | Tongji Hospital | Liu Xiulan[48] | Yes | 20-Jul | China | Single-center retrospective case series | 25 Jan to 15 Mar | EMR | 157 | Hospitalized COVID-19 patients with HTN (using ACEis/ ARBs or CCBs) | NI | 66 ± 10 | 46 | 100 | 27 | NI | ACEi/ARB, CCB | Severity (mild/moderate vs severe/critically ill - based on national guidelines), mortality |
| 21 | Shenzhen Third People's Hospital | Meng[49] | Yes | 31-Mar | China | Single-centre retrospective review | 11 Jan to 23 Feb | EHR | 42 | COVID-19 patients with HTN | | 65 (56–69) | 57 | 100 | 14 | NI | ACEIS, ARBs | Severity (severe vs moderate - based on national guidelines), mortality |

| | | | | | | | | | | | | | | | | | | |
|---|---|---|---|---|---|---|---|---|---|---|---|---|---|---|---|---|---|---|
| 22 | Union Hospital | Peng[50] | Yes | 2-Mar | China | Single-centre RC | 20 Jan to 15 Feb | NI | 112 | Adult COVID-19 patients with CVDs | NI | 62 (55–67) | 47 | 82 | 20 | NI | ACEIS, ARBs | Severity (critical vs mild/severe - based on diagnosis and treatment standards), mortality |
| 23 | Tongji | Qin[51] | Yes | 29-May | China | Single-centre RC | 27 Jan to 5 Mar | Patients' medical records | 50 | Consecutive COVID-19 patients with stroke history | NI | 70 (64–80) | 60 | 76 | 26 | NI | Anticoagulants, antiplatelets, statins | Severity (mild vs severe, based on all-cause death, admission to ICU, and mechanical ventilation), mortality |
| 24 | NA | Shi[52] | No | 1-Apr | China | Single-centre RC | 1 Feb to 15 Mar | EHR | 42 | Hospitalized COVID-19 patients | NI | 69 (range 40–91) | 64 | 31 | 19 | NI | Anticoagulants | Hospitalization |
| 25 | Union Hospital | Tan[19] | Yes | 15-May | China | Single-centre RC | 28 Jan to 8 Apr | NI | 100 | Consecutive hospitalized COVID-19 patients with HTN | NI | 67 (57–71) | 51 | 10 0 | 28 | NI | ACEIs, ARBs | Severity (Critical/ severe vs mild/ moderate, ventilation, ARDS), hospitalization, mortality |
| 26 | Tongji | Tang[53] | Yes | 27-Mar | China | Single-centre RC | 1 Jan to 13 Feb | EHR | 449 | Patients with severe COVID-19 | NI | 65 ± 12 | 60 | 39 | 21 | NI | Anticoagulants | 28-day mortality |
| 27 | Tongji Hospital | Xie Yang[54] | No | 7-Jul | China | Single-center RC | 27 Jan to 8 Mar | EMR | 619 | Hospitalized COVID-19 patients | NI | 58 ± 14 | 48 | 30 | 14 | NI | ACEi/ARB | Hospitalization length, severity (mild/common vs severe/critically ill - based on national guidelines), mortality |
| 28 | Union Hospital | Xie Yangjing[55] | Yes | 13-Jun | China | Multicenter RC | 15 Feb to 14 Mar | EMR | 62 | Consecutive hospitalized COVID-19 patients | NI | 66 (53–73) | 44 | 39 | 21 | | ACEI/ARB | Severity (mild/ moderate vs severe/ critical - based on national guidelines) |
| 29 | Renmin Hospital | Xiong[3] | Yes | 8-May | China | Multicentre RC | 1 Jan to 10 Mar | Online registration system | 131 | Maintenance hemodialysis COVID-19 patients | | 63 ± 13 | 57 | NI | 23 | NI | ACEIS, ARBs | Severity (mild/moderate versus severe/critical - national guidelines) |
| 30 | NA | Xu[29] | Yes | 3-Jul | China | Single-center RC | 29 Dec 2019 to 15 Feb 2020 | EMR | 101 | Adult hospitalized COVID-19 patients with HTN (on anti-hypertensive treatment) | NI | 65 (58–73) | 52 | 10 0 | 19 | NI | ACEI/ARB | Hospitalization length, severity (ARDS, ICU admission, mechanical ventilation), mortality |
| 31 | Zhejiang | Yan[56] | No | 29-Apr | China | Multicentre/population-based case-control | 10 Jan to 28 Feb | EHR | 49277: 610 cases, 48667 controls | Consecutive adult COVID-19 patients, General population control group | NI | 50 ± 17 | 48 | 20 | 6 | NI | ACE/ARBs, antiplatelets, β-blockers, CCBs, diuretics, LMDs | Susceptibility to infection, severity (critical/severe vs mild/moderate - based on national guidelines) |

| | | | | | | | | | | | | | | | | | | |
|---|---|---|---|---|---|---|---|---|---|---|---|---|---|---|---|---|---|---|
| 32 | NA | Yang G[57] | Yes | 29 Apr (posted on medrxiv on 4 Apr) | China | Single-center RC (case series) | 5 Jan to 22 Feb | EHR | 126 (subset) | COVID-19 patients with HTN | NI | 66 (61–73) | 49 | 100 | 30 | NI | ARBs/ACE | Hospitalization, severity (critical/ severe vs mild/ moderate - based on national guidelines), mortality |
| **33** | NA | Yang W[58] | Yes | 8-Apr | China | Single-centre RC | 25 Jan to 25 Feb | EHR | 344 | COVID-19 patients admitted to ICU | NI | 64 (52–72) | 52 | 41 | 19 | NI | ACEis | Mortality |
| 34 | NA | Yang X[59] | Yes | 21-Feb | China | Single-center RC | 24 Dec 2019 to 26 Jan 2020 | EMR | 52 | ICU COVID-19 patients ≥ 70 years | NI | 60 ± 13 | 67 | NI | 17 | NI | Vasoconstrictors | 28-day mortality |
| 35 | NA | Yao[60] | Yes | 10-Jul | China | Single-center case-control | 28 Jan to 8 Mar | EMR | 248 | Consecutive hospitalized COVID-19 patients | NI | 63 ± 13 | 54 | 32 | 18 | NI | Anticoagulants | Mortality |
| 36 | Zhejiang | Ye[61] | Yes | 16-Jun | China | Multicenter RC | 17 Jan to 7 Feb | Medical records and patients' self-report | 142 | Hospitalized COVID-19 patients with HTN | NI | 58 ± 12 | 59 | 100 | NI | NI | ACEI/ARB | Severity (mild/ ordinary vs severe/ critical - based on national guidelines; ICU admission, shock, invasive ventilation, and death), mortality |
| 37 | NA | Yin[62] | No | 5-May | China | Single-centre retrospective case series | 4 Feb to 14 Apr | EHR | 106 | Hospitalized COVID-19 patients with neurological diseases | NI | 73 ± 12 | 60 | 68 | 35 | NI | Anticoagulants, antiplatelets, LMDs | Severity (mild/ moderate vs severe/ critically ill - based on national guidelines) |
| 38 | NA | Yu | No | Poster (from Zhang[63] system-atic review) | China | Multicentre RC | 17 Jan to 19 Feb | Medical record review | 276 | NI | NI | 60 (52–68) | 53 | NI | NI | NI | ACE/ARBs | Mortality |
| 39 | Tongji Hospital | Zeng H[64] | No | 16-Jun | China | Single-centre RC | 27 Jan to 8 Mar | EHR | 1031 | Hospitalized COVID-19 patients | NI | 67 ± 11 | 52 | 37 | 18 | NI | ACEIS, ARBs, CCB, LMDs | Mortality |
| 40 | NA | Zeng Z[65] | No | 11-Apr | China | Single-centre RC | 5 Jan to 8 Mar | Patient medical records | 75 | Adult COVID-19 patients with HTN | NI | 60 ± 15 | 47 (Table 4), 55 (Table 1) | 100 | 31 | NI | ACEIs, ARBs | Hospitalization, severity (severe vs non-severe - based on published literature), 28-day mortality |
| 41 | Tongji Hospital | Zhang L[66] | No | 14-Apr | China | Multicentre RC | 10 Jan to 30 Mar | Patient medical records | 96 | Hospitalized COVID-19 patients with HTN | NI | 67 (59–72) | 53 | 100 | NI | NI | Antihypertensives, CCBs | Mortality |
| 42 | Renmin Hospital, Seventh Hospital | Zhang P[67] | Yes | 17-Apr | China | Multicentre RC | 31 Dec 2019 to 20 Feb | EHR | 1128 | COVID-19 patients with HTN | | 64 (56–69) | 53 | 100 | 21 | NI | ACEIS, ARBs | Severity (ARDS, ventilation and extracorporeal membrane oxygenation), 28-day mortality |

| | | | | | | | | | | | | | | | | | | |
|---|---|---|---|---|---|---|---|---|---|---|---|---|---|---|---|---|---|---|
| 43 | Other studies recruiting from Hubei hospitals | Zhang XJ[68] | Yes | 24-Jun | China | Multicenter RC | 30 Dec 2019 to 17 Apr 2020 | EMR | 13981 | Hospitalized COVID-19 patients | NI | 57 ± 16 | 49 | 35 | 16 | NI | ACEI/ARBs, statins | Severity (invasive mechanical ventilation, ICU admission, ARDS), mortality |
| 44 | NA | Zhou H[69] | No | 10-Jul | China | Single-centre case-control | 10 Jan to 15 Mar | EMR and patient/family member interview | 142 | Hospitalized hemodialysis patients tested for COVID-19 | NI | 61 ± 11 | 69 | >30 | >27 | NI | ACEi/ARB | Susceptibility to infection |
| 45 | NA | Zhou X[70] | Yes | 13-May | China | Single-centre retrospective case series | 25 Jan to 20 Feb | Patient medical records | 36 | Hospitalized COVID-19 patients with HTN | NI | 65 ± 10 | 53 | 100 | 25 | NI | ACEIS, ARBs | Hospitalization, mortality |
| 46 | Danish national administrative registries | Fosbol[7] | Yes | 19-Jun | Denmark | Nation-wide nested case control | 1 Feb to 4 May | Danish national administrative registries | 6281 | Cases (COVID-19 patients with HTN), age- and sex-matched controls with prior HTN but not COVID-19 | NI | 74 (63–81) | 54 | 100 | 15 | NI | ACEIS, ARBs, CCBs | Susceptibility to infection, severity (ICU admission or death), 30-day mortality |
| 47 | Danish national administrative registries | Reilev[71] | No | 26-May | Denmark | Nationwide population-based cohort | 27 Feb to 30 Apr | Danish microbiology, administrative and health databases | 228677 | Consecutive COVID-19 patients | NI | 47 (31–60) for 219158, 49 (34–63) for 9519 | 37 | 24 | 7 | NI | Antihypertensives, ACEIS, ARBs, CCBs, β-blockers, diuretics, LMDs, antiplatelets, anticoagulants | Susceptibility to infection, hospitalization, severity (ICU admission), mortality |
| 48 | NA | Feuth[72] | No | 18-May | Finland | Single-centre RC | Admitted by 3 May | Hospital records | 28 | Hospitalized Covid-19 patients | NI | 56 (47–72) | 54 | 43 | 25 | 37 | ACEIS, ARBs | Severity (ICU admission) |
| 49 | NA | Allenbach[73] | Yes | 8-May | France | Single-center PC | From 16 Mar | Standardized form | 147 | Consecutive hospitalized adult COVID-19 patients | 63% White (of 135) | 77 (60–83) [for 152] | 61 | 52 | 25 | NI | ACEis | Severity (14-day ICU transfer/ventilation/death) |
| 50 | NA | Bar[74] | Yes | 10-Jun | France | Single-center RC | Mar to Apr | Patient records | 100 | COVID-19 tested adults | NI | 68 ± 16 | 41 | 57 | 10 | NI | ACEI/ARB | Susceptibility to infection |
| 51 | NA | Basse[75] | No | 19-May | France | Multicentre PC | 13 Mar 13 to 25 Apr | Institutional Redcap database | 141 | Cancer COVID-19 patients | NI | 62 (52–72) | 28 | 34 | 17 | 18 | ACEIs, ARBs, anticoagulants | Severity (28-day transfer to ICU or death) |
| 52 | NA | Benotmane[11] | No | 19-Jun | France | Single-center RC | 4 Mar to 7 Apr | EMR | 40 | Hospitalized COVID-19 kidney transplant patients | NI | 64 (55–68) | 78 | 83 | 48 | 50 | ACEi/ARB | Severity (non-severe vs severe) |
| 53 | Other French trials | Cariou[21] | Yes | 29-May | France | Multicentre RC | 10 Mar to 31 Mar | Inpatient medical files | 1317 | COVID-19 patients with DM | 62% Europid, 19% MENA, 17% | 70 ± 13 | 65 | 77 | 100 | 38 | ACEIS, ARBs, β-blockers, diuretics, LMDs | Severity (tracheal intubation for mechanical ventilation and/or death within 7 days |

| | | | | | | | | | | | | | | | | | | |
|---|---|---|---|---|---|---|---|---|---|---|---|---|---|---|---|---|---|---|
| | | | | | | | | | | | African, 2% Asian | | | | | | | of admission), 7-day mortality |
| 54 | NA | Dauchet[5] | No | 1-May | France | Single centre RC | 29 Feb to 5 Apr | EHR | 288 | Consecutive patients >35 years with suspected COVID-19 | NI | 55 ± 15 | 62 | NI | 14 | NI | Antihypertensives, ACEIs, ARBs | Susceptibility to infection, hospitalization, severity (ICU admission) |
| 55 | NA | Khider[76] | Yes | 18-Jun | France | Single-center RC (call it prospective) | 14 Mar to 20 Mar | Medical records | 96 | Consecutive adult COVID-19-suspected/tested hospitalized patients | NI | 66 ± 18 | 59 | 49 | 17 | NI | ACEi/ARB, anticoagulants, beta-blockers, CCBs, statins | Susceptibility to infection |
| 56 | NA | Kibler[77] | No | 16-Jun | France | Single-centre RC | 1 Jan to 8 May | Patient medical records | 702 | Patients with aortic stenosis who had undergone transcatheter aortic valve replacement | NI | 82 ± 7 | 44 | 83 | 30 | 26 | ACEIS, ARBs, anticoagulants, LMDs, antiplatelets, antiarrythmics | Susceptibility to infection, severity (hospitalization/death) |
| 57 | NA | Liabeuf[78] | Yes | 12-Jun | France | Single-centre RC | 28 Feb to 30 Mar | EMR | 268 | Consecutive hospitalized COVID-19 patients | NI | 73 (61–84) | 58 | 57 | 21 | 39 | ACEi/ARB, beta-blockers, CCB, diuretics | Severity (ICU admission or death), mortality |
| 58 | NA | Meszaros[79] | Yes | 3-Jun | France | Multicentre RC | 10 Mar to 18 Apr | Medical (admission and hospitalization) records | 103 | Consecutive hospitalized COVID-19 patients with HTN | NI | 72 ± 11 | 58 | 100 | NI (27% for 234) | NI | ACEi/ARB | Severity (severe vs non-severe - based on WHO guidelines), mortality |
| 59 | NA | Oussalah[16] | Yes | 5-Jun | France | Single-center RC | 1 Mar to 25 Mar | Nancy Biochemical Database | 146 | Consecutive hospitalized COVID-19 patients | NI | 65 (54–77) [for 149] | 61 [for 149] | 50% [for 133] | 29% [for 133] | NI | ACEi/ARBs | Severity (acute respiratory failure or intubation and mechanical ventilation), mortality |
| 60 | NA | Rath[80] | Yes | 14-Jun | Germany | Single-center RC (call it prospective) | Feb to Mar | NI | 123 | Consecutive hospitalized COVID-19 patients | NI | 68 ± 15 | 63 | 70 | 24 | 20 | ACEi/ARB, anticoagulants, antiplatelets, beta-blockers, CCBs, statins | 30-day mortality |
| 61 | NA | Rieder[81] | Yes | 2-Jul | Germany | Single-center RC (call it prospective) | 26 Mar to 20 Apr | EMR | 190 | Hospitalized COVID-19 suspected patients (all tested for COVID-19) | NI | median 60 | 53 | NI | NI | NI | Anticoagulation | Susceptibility to infection |
| 62 | NA | Sacco[82] | No | 24-Jul | Germany | Population cohort study | Feb to Jun | Online survey | 165 | COVID-19 adult patients | NI | Categorized (mode 40-59) | 33 | NI | NI | NI | ACEi/ARB, statins | Severity (grade 1/grade 2 vs grade 3/grade 4/grade 5 - based on a survey question) |
| 63 | NA | Cheung[83] | Yes | 8-Jun | Hong Kong | Single-centre RC | 1 Jan to 27 Apr | EHR | 734 | Adult COVID-19 patients | NI | NI | NI | NI | NI | NI | ACEIs/ARBs | Severity (severe pneumonia, critical |

| | | | | | | | | | | | | | | | | | | |
|---|---|---|---|---|---|---|---|---|---|---|---|---|---|---|---|---|---|---|
| | | | | | | | | | | | | | | | | | | complications, ventilatory support, ICU admission or death) |
| 64 | NA | Zhou[84] | No | 2-Jul | Hong Kong (China) | Multicenter RC | 1 Jan to 24 May | Clinical Data Analysis and Reporting System (CDARS) database | 976 | Consecutive hospitalized COVID-19 patients | 88% Chinese [for 1043] | 34 (32–36) [for 1043] | 54 [for 1043] | 20 [for 535] | 10 [for 535] | NI | ACEI/ARB | Severity (ICU admission) |
| 65 | NA | Ashraf[10] | No | 24-Apr | Iran | Single-centre RC | 22 Feb to 5 Mar | Patient medical records | 100 | Hospitalized COVID-19 patients | NI | 58 (48–68) | 65 (for 99) | 26 | 26 | NI | ACEIs, ARBs | Severity (critical vs noncritical - based on national guidelines), mortality |
| 66 | NA | Amit[85] | Yes | 18-Jul | Israel | Multicentre retrospective registry-based case series | 5 Mar to 27 Apr | Covid-19 ICU registry | 156 | Consecutive ICU COVID-19 patients | NI | 72 (60–82) | 69 | 54 | 40 | 70 | Anti-Fibrinolytics | Mortality |
| 67 | NA | Chodick[86] | Yes | 14-May | Israel | Single-centre cross-sectional study | 1 Jan to before 25 Mar | Healthcare database | 14520 | Participants tested for COVID-19 | NI | 37 ± 19 | 47 | 11 | 5 | 15 | ACEI/ARB | Susceptibility to infection |
| 68 | NA | Alberici[87] | Yes | 8-May | Italy | Multicentre cohort | 1 Mar to 3 Apr | NI | 94 | Hemodialysis patients with COVID-19 | NI | 72 (62–79) | 66 | 93 | 43 | NI | ACEI/ARB, Heparin (prophylactic) | Hospitalization |
| 69 | Brescia | Alberici[88] | Yes | 9-Apr | Italy | Single-centre consecutive case series | 27 Feb to 24 Mar | NI | 20 | Consecutive kidney transplant patients with COVID-19 | NI | 59 (51–64) | 80 | 85 | 15 | NI | ACEI/ARB | Mortality |
| 70 | NA | Benelli[89] | No | 30-Apr | Italy | Single-centre RC | 21 Feb to 13 Mar | Electronic database | 411 | Consecutive COVID-19 patients | NI | 67 ± 16 | 67 | 47 | 16 | NI | ACEIs, ARBs | Severity (ICU admission, CPAP or non-invasive ventilation), mortality |
| 71 | NA | Bravi[20] | Yes | 24 Jun (posted on medrxiv on 23 May) | Italy | Multicentre retrospective case-control | Until 24 Apr | EHR | 1603 | COVID-19 patients | NI | 58 ± 21 | 47 | 34 | 12 | NI | ACEIs, ARBs | Severity (mild vs severe/very severe/ lethal - based on hospitalization, ICU, and death) |
| 72 | NA | Cannata[90] | Yes | 5-Jun | Italy | Single-centre RC | Until 1 Apr | Prospective registry | 397 | Consecutive COVID-19 patients | NI | NI | NI | NI | NI | NI | ACEIS, ARBs | Mortality |
| 73 | NA | Conversano[91] | Yes | 8-May | Italy | Single centre retrospective case series | 27 Feb to 17 Mar | EHR | 191 | Hospitalized adult COVID-19 patients | NI | 63 ± 15 | 69 | 50 | 15 | NI | ACEIS, ARBs, β-blockers, CCB, diuretics, LMDs | Mortality |

| | | | | | | | | | | | | | | | | | | |
|---|---|---|---|---|---|---|---|---|---|---|---|---|---|---|---|---|---|---|
| 74 | NA | Di Micco[92] | Yes | 7-May | Italy | Multicentre RC | Feb to Mar | Medical records | 67 | Hospitalized COVID-19 patients | NI | Categorized | 70/67 | NI | NI | NI | Antiplatelets, anticoagulants | Severity (based on presence of SARS) |
| 75 | NA | Fasano[93] | Yes | 2-Jun | Italy | Single-center case-control | Between 20 Feb and 3 May | Patient electronic charts and phone interview | 1486 | COVID-19 patients with Parkinson's Disease | NI | 73 ± 10 | 57 | 39 | 8 | 11 | ACEI/ARB | Susceptibility to infection |
| 76 | NA | Felice[94] | Yes | 8-Jun | Italy | Single-centre retrospective consecutive cohort/series | 9 Mar to 31 Mar | Medical records | 133 | Consecutive COVID-19 patients with HTN | NI | 73 ± 13 | 65 | 100 | 26 | NI | ACEIS, ARBs | Hospitalization, severity (need for oxygen, ICU admission - based on PaO2/FiO2 ratio <250 and need for invasive or non-invasive ventilation and non-invasive ventilation), mortality |
| 77 | NA | Ferrante[95] | Yes | 8-Jul | Italy | Single-center RC | 25 Feb to 2 Apr | EHR | 332 | Consecutive COVID-19 patients undergoing chest CT on admission | NI | 70 (55–76) | 71 | 54 | 21 | NI | ACEi/ARB, statins | Mortality |
| 78 | NA | Giacomelli[96] | Yes | 22 May (posted on medrxiv on 6 May) | Italy | Single-centre PC | 21 Feb to 19 Mar | Patient clinical charts | 233 | Adult hospitalized COVID-19 patients | NI | 61 (50–72) | 69 | 36 | NI | 16 | ACEIs, ARBs, antihypertensives, antiplatelets, anticoagulants, antiarrhythmics, β-blockers, CCBs, diuretics, LMDs | Mortality |
| 79 | NA | Giorgi Rossi[97] | No | 16-Apr | Italy | Population-based RC (they call it prospective) | 27 Feb to 2 Apr | EHR | 2653 | Symptomatic COVID-19 patients | NI | 63 mean | 50 | 16 | 11 | 2 | ACEIs, ARBs | Hospitalization, mortality |
| 80 | NA | Gnavi[98] | Yes | 22-May | Italy | Population-nested case-control | 22 Feb to 23 Mar (cases) | Automated system of databases | 1896: 316 cases, 1580 controls | Patients with HTN (at risk of COVID-19) | NI | 71 ± 12 | 59 | 100 | NI | NI | ACEIS, ARBs | Susceptibility to infection |
| | | | | | | | | | 1026: 171 cases, 855 controls | Patients with CVDs and DM (at risk of COVID-19) | NI | 71 ± 11 | 78 | 0 | 36 | NI | | |
| **81** | NA | Grasselli[8] | Yes | 15-Jul | Italy | Multicentre RC | 20 Feb to 22 Apr | Regional Health | 3988 | Consecutive ICU COVID-19 patients | NI | 63 (56–69) | 80 | 42 | 13 | NI | ACEi/ARB, antiplatelets, anticoagulants, | Mortality |

| | | | | | | | | | | | | | | | | | | |
|---|---|---|---|---|---|---|---|---|---|---|---|---|---|---|---|---|---|---|
| | | | | | | | | System Database | | | | | | | | | beta-blockers, diuretics, statins | |
| 82 | NA | Iaccarino[99] | Yes | 22-Jun | Italy | Multicenter cross-sectional study | 9 Mar to 9 Apr | Online questionnaire completed using EMR | 1591 | Adult COVID-19 patients | 94% Italian nationality | 67 ± 0.4 | 65 | 55 | 17 | 6 | ACEI/ARBs, beta-blockers, CCBs, diuretics | Mortality |
| 83 | NA | Iacovoni[100] | Yes | 26-Jun | Italy | Multicentre RC | 1 Feb to 31 Mar | Medical records | 26 | Heart transplanted patients with COVID-19 | NI | 63 (22–77) | 77 | 58 | 15 | NI | Anticoagulant | Mortality |
| 84 | Brescia | Inciardi[101] | Yes | 8-May | Italy | Multicentre RC | 4 Mar to 25 Mar | Medical records | 99 | Consecutive hospitalized COVID-19 patients | NI | 67 ± 12 | 81 | 64 | 31 | 23 | ACEIs, ARBs, ARNIs, anticoagulants, LMDs | 14-day mortality |
| 85 | NA | Mancia[102] | Yes | 1-May | Italy | Population-based case-control | 21 Feb to 11 Mar | Healthcare use databases | 37031: 6272 cases, 30759 controls | Patients at risk of COVID-19 | NI | 68 ± 13 | 63 | 51 | NI | NI | Antihypertensives, ACEIS, ARBs, CCBs, β-blockers, diuretics, LMDs, antiplatelets, anticoagulants, digitalis, nitrate | Susceptibility to infection |
| 86 | NA | Oliva[103] | Yes | 28-Jul | Italy | Single-centre retrospective case series | 1 Mar to 30 Apr | Clinical reports | 7 | COVID-19 patients with Chlamydia pneumoniae (n = 5) or Mycoplasma pneumoniae (n = 2) co-infection | NI | 73 (45–79) | 57 | 29 | 29 | NI | Anticoagulants | Hospitalization length |
| 87 | NA | Parigi[104] | Yes | 16-Jul | Italy | Single-center RC | 22 Feb to 30 Mar | NI | 325 | Consecutive hospitalized COVID-19 patients | NI | 66 (55–75) | 69 | 51 | NI | NI | ACEI/ARB | Severity (supplemental oxygen, mechanical ventilation and death or ICU admission) |
| 88 | NA | Perotti[105] | No | 29-May | Italy | Multicenter PC | 25 Mar to 21 Apr | REDCap database | 46 | Severe Covid-19 patients treated with hyperimmune plasma | NI | 63 ± 12 | 61 | 48 | 22 | NI | Anticoagulants | 7-day mortality |
| 89 | NA | Russo[106] | Yes | 29-May | Italy | Multicentre RC | Feb to Apr | NI | 192 | Hospitalized COVID-19 patients | NI | 68 ± 15 | 60 | 58 | 22 | 14 | Antiplatelets, anticoagulants | Severity (ARDS), mortality |
| 90 | NA | Sardu[18] | Yes | 7-Jul | Italy | Multicenter RC (calls it prospective) | Before Feb 29 | EMR | 62 | Consecutive hospitalized COVID-19 patients (aged >18 and <80 years) with HTN | NI | 58 ± 18 | 66 | 100 | 26 | NI | ACEi/ARB, CCB | Severity (ARDS, ICU admission or mechanical ventilation), mortality |
| 91 | NA | Tedeschi[107] | Yes | 27-Apr | Italy | Multicentre PC | 22 Feb to 4 Apr | Medical records | 311 (subset) | Hospitalized COVID-19 patients with HTN | NI | 76 (67–83) | 72 | 100 | 24 | NI | ACEIs, ARBs | Mortality |

| | | | | | | | | | | | | | | | | | |
|---|---|---|---|---|---|---|---|---|---|---|---|---|---|---|---|---|---|
| 92 | NA | Trecarichi[108] | No | 2-Jul | Italy | Single-centre RC | 27 Mar to 6 May | Hospital charts and the laboratory database | 48 | Hospitalized COVID-19 patients | NI | 80 ± 12 | 54 | 46 | 23 | 10 | ACEi/ARB | Mortality |
| 93 | NA | Viecca[109] | Yes | 23-May | Italy | Single-centre case-control | 9 Apr to 16 Apr | NI | 10 | Adult hospitalized patients with severe Covid-19 with hypercoagulability | NI | 64 ± 13 | 80 | NI | NI | NI | Antiplatelets | Mortality |
| 94 | NA | Violi[110] | Yes | 22-Jun | Italy | Multicenter RC | Mar to Apr | Medical records | 319 | Consecutive hospitalized adult (≥18 years) COVID-19 patients | NI | 68 ± 17 | 61 | 55 | 19 | NI | ACEi/ARBs, antiplatelets, statins | Mortality |
| 95 | Trials from Italy, Spain, France, Switzerland, Netherlands, USA, UK, China | Garassino[111] | Yes | 12-Jun | Italy, Spain, France, Switzerland, Netherlands, USA, UK, China | Multicenter RC | 26 Mar to 12 Apr | REDCap (Research Electronic Data Capture) database | 195 | Consecutive COVID-19 patients with thoracic malignancies | 94% White, 6% other [for 200] | 68 (62–75) [for 200] | 70 [for 200] | 47 [for 200] | 15 [for 200] | NI | ACEI/ARB, anticoagulants, antiplatelets | Hospitalization, mortality |
| 96 | NA | Higuchi[112] | No | 30-Jul | Japan | Single-center RC | 10 Feb to 10 Jun | Medical records | 57 | Consecutive hospitalized COVID-19 patients ≥16 years | 100% Asian | 52 (35–70) | 56 | 28 | 23 | NI | ARB, CCB, statin | Severity (mechanical ventilation) |
| 97 | NA | Almazeedi[113] | Yes | 4-Jul | Kuwait | Single-centre RC | 24 Feb to 20 Apr | EMR | 1096 | Consecutive hospitalized COVID-19 patients | 48% Indians, 27% Kuwaitis and 7% Egyptians | 41 (25–57) | 81 | 16 | 14 | 22 | Vasopressors | Mortality |
| 98 | NA | Ayed[114] | No | 20-Jun | Kuwait | Single-centre RC | 1 Mar to 30 Apr | EMR | 103 | Adult COVID-19 patients admitted to ICU | NI | 53 (44–63) | 86 | 35 | 39 | NI | ACEis, beta blockers, antiplatelets, LMDs | Mortality |
| 99 | NA | Brouns[115] | Yes | 7-Jun | Netherlands | Multicentre retrospective case series | 20 Mar to 1 May | Patient records | 101 | Nursing Home Residents with COVID-19 | NI | 85 ± 8 | 33 | 52 | 14 | NI | Anticoagulants, antiplatelets | Mortality |
| 100 | NA | Middeldorp[116] | Yes | 5-May | Netherlands | Single-center RC | 2 Mar to 12 Apr | Patient records | 198 | Consecutive hospitalized COVID-19 patients | NI | 61 ± 14 | 66 | NI | NI | NI | Anticoagulants, antiplatelets | Severity (ICU admission/mechanical ventilation) |
| 101 | NA | Davies[117] | No | 3-Jul | South Africa | Population cohort study | 1 Mar to 9 Jun | Western Cape Provincial | 3460932 | All public sector patients aged ≥20 years | NI | Categorized | 42 | 16 | 8 | NI | Diuretics | Susceptibility to infection, |

| | | | | | | | | | | | | | | | | | | |
|---|---|---|---|---|---|---|---|---|---|---|---|---|---|---|---|---|---|---|
| | | | | | | | | Health Data Centre (WCPHDC) | | | | | | | | | | hospitalization, mortality |
| 102 | HIRA database | Choi HK[118] | No | 13-Jun | South Korea | Nationwide retrospective case-control study | Until 15 May | HIRA database | 1585: 892 cases, 693 controls | COVID-19 patients with HTN. Controls included HTN patients. | NI | 66 ± 14 | 26 | 10 0 | 45 | NI | ACEIs, ARBs | Severity (severe infection or death - severe infection based on need for mechanical ventilation, ICU, CRRT or ECMO treatment), mortality |
| 103 | NA | Choi MH[119] | Yes | 23-Jun | South Korea | Single-centre RC | 5 Mar to 18 Mar | EMR | 293 | Consecutive hospitalized COVID-19 patients | NI | 29 (24–47) | 73 | 10 | 7 | NI | ARB, beta-blockers, CCBs, diuretics, statins, isosorbide | Severity (mild vs moderate/severe - based on national guidelines) |
| 104 | Considered part of Korean nationwide studies | Chung[120] | Yes | 21-May | South Korea | Single-center RC | Before 24 Apr | EMR | 29 | Consecutive hospitalized adult (age >18 years) COVID-19 patients with DM | NI | 66 ± 9 | 44 | 34 | 10 0 | NI | RAAS | Severity (ARDS, septic shock, ICU admission, and mortality within 28 days) |
| 105 | HIRA database | Huh[121] | No | 8-May | South Korea | Nationwide case-control study | Until 8 Apr | HIRA database | 65149 | Adults tested for COVID-19 | NI | 48 ± 20 | 49 | 33 | 28 | NI | ACEIs, ARBs, antiplatelets, antiarrythmics, LMDs | Susceptibility to infection |
| 106 | Considered part of Korean nationwide studies | Hwang[122] | Yes | 8-Jul | South Korea | Multicentre RC | 1 Feb to 25 Mar | EMR | 103 | Hospitalized COVID-19 Patients | NI | 68 ± 15 | 50 | 55 | 34 | NI | ACEi/ARBs | Mortality |
| 107 | HIRA database | Jung[22] | Yes | 22-May | South Korea | Nationwide population-based RC | Until 8 Apr | HIRA database | 5179 | Adult COVID-19 patients | NI | 45 ± 18 | 44 | 22 | 17 | NI | ACEIS, ARBs | Severity (mechanical ventilation), mortality |
| 108 | HIRA database | Kim[123] | Yes | 19-Jun | South Korea | Multicenter RC | 1 Jan to 2 Apr | HIRA database | 137805 2 | Adults ≥ 40 years living in Daegu Metropolitan City | NI | Categor ized | 47 | NI | NI | NI | ACEIs/ARBs, beta-blockers, CCBs, diuretics | Susceptibility to infection |
| 109 | HIRA database | Lee[124] | No | 1-Apr | South Korea | Multicentre RC | 19 Jan to 16 Mar | HIRA database | 8266 | Hospitalized COVID-19 patients | NI | 44 ± 19 | 38 | 19 | 17 | NI | ACEIs, ARBs | 60-day mortality |
| 110 | HIRA database | Rhee[125] | No | 23-May | South Korea | Population-based RC | Until 17 May | HIRA database | 832 | COVID-19 patients with DM | NI | 62 ± 16 | 53 | 68 | 10 0 | NI | ACEIs, ARBs | Severity (mild vs severe/lethal - based on ICU/death) |
| 111 | Other Spanish trials | Amat-Santos[126] | Yes | 26-May | Spain | Multicentre open-label RCT (retrospective analysis of RCT data, non-pre-specified | 1 Jan to 1 Apr | Phone calls and EHRs | 102 | Adult aortic stenosis patients successfully treated TAVR | NI | 82 ± 6 | 57 | 54 | 21 | NI | Ramipril, anticoagulants, LMDs | Susceptibility to infection, mortality |

| | | | | | | | | | | | | | | | | | | |
|---|---|---|---|---|---|---|---|---|---|---|---|---|---|---|---|---|---|---|
| | | | | | | interim analysis) | | | | | | | | | | | | |
| 112 | NA | Ayerbe[127] | Yes | 31 May (posted on medrxiv on 29 May) | Spain | Multicentre RC | 1 Mar to 20 Apr | Patient clinical records | 2019 | COVID-19 patients | NI | 68 ± 16 | 61 | NI | NI | NI | Heparin | Mortality |
| 113 | Other Madrid trials | Bernaola [128] | No | 21-Jul | Spain | Multicentre RC | 24 Feb to 24 Apr | HM hospital network database | 1645 | Hospitalized COVID-19 Patients | NI | Categorized (mode 60-79) | 62 | 30 | 13 | NI | Anticoagulants, antiplatelets | Severity (intubation or death), mortality |
| 114 | Other Spanish trials | de Abajo[129] | Yes | 14-May | Spain | Multicentre retrospective consecutive series | 1 Mar to 24 Mar | EHR | 1139 | Adult hospitalized COVID-19 patients | NI | 69 ± 15 | 61 | NI | NI | NI | ACEIS, ARBs | Hospitalization, severity (death and ICU) |
| 115 | NA | Fernández-Ruiz[130] | Yes | 16-Apr | Spain | Single-center case series | 5 Mar to 23 Mar | EMR | 18 | Adult COVID-19 patients with solid organ transplants | NI | 71 (63–75) | 78 | 72 | 50 | 11 | ACEI/ARB | Mortality |
| 116 | NA | Golpe[131] | Yes | 25-Jun | Spain | Single-center RC | 9 Mar to 1 Apr | EMR | 157 | Consecutive COVID-19 patients with HTN | NI | 70 ± 12 | 46 | 100 | 34 | NI | ACEI/ARB, beta blocker, CCB, diuretics | Hospitalization/severity |
| 117 | NA | Jurado[132] | No | 16-May | Spain | Multicenter RC | During the second half of March | EHR | 574 | Consecutive hospitalized COVID-19 patients | NI | 63 ± 16 | 59 | 51 | 22 | NI | ACEIs, ARBs | Severity (severe vs mild/moderate - based on acute respiratory distress, shock, ICU admission, physician consideration or death) |
| 118 | NA | Lopez-Otero[24] | Yes | 5-Jun | Spain | Single-center RC | 10 Mar to 6 Apr | EMR | 447979 | Population at risk of COVID-19 | NI | NI | NI | NI | NI | NI | ACEI/ARB | Susceptibility to infection, hospitalization, severity (ICU admission), mortality |
| 119 | NA | Lorente-Ros[133] | Yes | 26-Jun | Spain | Single-center RC | 18 Mar to 23 Mar | EMR | 707 | Consecutive hospitalized adult (≥18 years) COVID-19 patients | NI | 67 ± 16 | 63 | 51 | 20 | NI | ACEi/ARBs | 1-month mortality |
| 120 | NA | Marcos[134] | No | 14-Jul | Spain | Single-center RC | 1 Mar to 23 Apr | NI | 918 | Hospitalized COVID-19 patients | NI | 73 ± 15 | 58 | 53 | 23 | 25 | ACEi/ARB, anticoagulants | Severity (mechanical ventilation or death) |
| 121 | Other Spanish trials | Martínez-López[135] | No | 30-Jun | Spain | Multicenter retrospective case series | 1 Mar to 30 Apr | Patient records | 167 | Hospitalized COVID-19 Patients with Multiple Myeloma | NI | 71 (62–78) | 57 | 40 | 17 | NI | Anticoagulants | Mortality |
| 122 | Other Spanish trials | Pérez-Sáez[136] | Yes | 12-Jul | Spain | Multicentre RC | 18 Mar to 9 May | Societal registry | 80 | Hospitalized tocilizumab-treated kidney transplant patients with severe COVID-19 | 89% Caucasian | 59 ± 12 | 68 | 89 | 29 | 18 | ACEI/ARB | Mortality |

| | | | | | | | | | | | | | | | | | |
|---|---|---|---|---|---|---|---|---|---|---|---|---|---|---|---|---|---|
| 123 | NA | Poblador-Plou[137] | Yes | 17-Jul | Spain | Multicenter RC | 4 Mar to 17 Apr | EHR | 4412 | COVID-19 patients | NI | 68 ± 21 | 41 | 34 | 12 | NI | ACEi/ARBs, antithrombotics, adrenergics, beta-blockers, cardiac glycosides, CCBs, diuretics, LMDS, vasodilators | 30-day mortality |
| 124 | NA | Regina[138] | No | 14-May | Switzerland | Single-centre RC | 1 Mar to 25 Mar | EHR | 200 | Consecutive adult hospitalized COVID-19 patients | NI | 70 (55–81) | 60 | 44 | 22 | 27 | ACEIs, ARBs | Severe disease (mechanical ventilation) |
| 125 | NA | Pongpirul[139] | No | 26-Jun | Thailand | Single-centre RC | 8 Jan to 16 Apr | Medical records | 193 | Consecutive hospitalized adult COVID-19 patients | 91% Thai, 9% Non-Thai | 37 (29–53) | 59 | 16 | 8 | 13 | ACEi/ARB | Severity (asymptomatic/mild/ moderate vs severe/critical - based on WHO-China joint mission/Chinese CDC guidelines) |
| 126 | NA | Selçuk[140] | Yes | 22-Jun | Turkey | Single-center RC | Up to before 31 May 2020 | EMR | 113 | Consecutive hospitalized COVID-19 patients with HTN | NI | 64 ± 11 | 48 | 100 | 42 | NI | ACEi/ARBs, anticoagulants, antiplatelets, beta-blockers, CCBs, diuretics | Hospitalization length, severity (ICU admission or endotracheal intubation), mortality |
| 127 | NA | Senkal[28] | Yes | 3-Jul | Turkey | Single-centre RC | 9 Mar to 11 May | EHR | 611 | Hospitalized adult COVID-19 patients | NI | 57 ± 15 | 59 | 41 | 23 | NI | ACEI/ARB, diuretics | Hospitalization length, severity (hospitalization of ≥14 days, ICU admission or death), mortality |
| 128 | NA | Baker[141] | No | 19-May | UK | Multicentre RC | 8 Jan to 16 Apr | EHR | 286 | Consecutive adult hospitalized COVID-19 patients | 93% White (for 316) | 75 (60–83) for 316 | 55 (for 316) | 42 | 27 | NI | ACEIS, ARBs | 28-day mortality |
| 129 | NA | Bataille[142] | No | 11-Jul | UK | Multicenter RC | 7 May to 22 Jun | COVID Symptom Study app | 27157 | COVID Symptom Study app users tested for COVID-19 | 93% White, 3% Asian, <1% Black, 3% other | 44 ± 18 | 39 | ~13 | 4 | NI | Blood pressure medications | Susceptibility to infection |
| 130 | NA | Bean[143] | Yes | 2 Jun (pre-print posted on 11 Apr) | UK | Multicentre consecutive series/cohort (prospective data collection, retrospective analysis) | 1 Mar to 13 Apr | EHR | 1200 | Adult hospitalized COVID-19 patients | 43% White, 26% Black, 5% Asian, 27% | 68 ± 17 | 57 | 54 | 35 | 15 | ACEIS, ARBs | Severity (death or transfer to ICU within 21-days of symptom onset), mortality |

| | | | | | | | | | | | | | | | | | | |
|---|---|---|---|---|---|---|---|---|---|---|---|---|---|---|---|---|---|---|
| 131 | NA | Fletcher[144] | No | 15-May | UK | Multicentre RC | 1 Jan to 23 Apr | EHR | 1279 | Patients aged ≥16 years at risk of COVID-19 | 37% Asian, 12% Black, 5% White, 45% other (of 2756) | 56 ± 22 (of 2756) | 52 (of 2756) | 19 | 10 | NI | Antihypertensives, anticoagulants, β-blockers, CCBs, antianginal drugs including nitrates, LMDs | Susceptibility to infection |
| 132 | UK Biobank | Ho[145] | No | 2-May | UK | Population-based PC | 16 Mar to 14 Apr | Biobank | 285817 | Participants tested for COVID-19 | 95% White, 1% Black, 2% South Asian, 2% other | 56 ± 8 | 46 | NI | 5 | 23 | Antihypertensives, LMDs | Susceptibility to infection |
| 133 | UK Biobank | Khawaja[146] | No | 11-May | UK | Population-based PC | 16 Mar to 16 Apr | Biobank | 406793 | COVID-19 cases and untested biobank participants as the controls | 94% White, 2% Asian, 2% Black, 2% other | 68 ± 8 | 45 | 33 | 5 | 23 | ACE/ARBs, antihypertensives, β-blockers, CCBs, diuretics | Susceptibility to infection/hospitalization |
| 134 | UK Biobank | Kolin[147] | Preprint | 5-May | UK | Population-based PC | From 16 Mar | Biobank | 1474 | Participants tested for COVID-19 | 89% White, 4% Asian, 5% Black | 58 ± 9 | 53 | NI | 9 | NI | ACEIS, ARBs, CCBs, β-blockers | Susceptibility to infection |
| 135 | UK Biobank | Raisi-Estabragh[148] | No | 15-May | UK | Nation-wide PC | 16 Mar to 14 Apr | Biobank | 1474 | Participants tested for COVID-19 | 89% White | 69 ± 9 | 53 | 49 | 16 | NI | ACEIs, ARBs | Susceptibility to infection |
| 136 | UK Biobank | Raisi-Estabragh[149] | Yes | 14-Jul | UK | Nation-wide RC (prospective data collection in a Biobank) | 16 Mar to 14 Jun | Biobank | 7099 | Hospitalized COVID-19 tested Biobank participants | 92% White, 8% other | 69 ± 9 | 50 | 47 | 15 | NI | ACEi/ARB | Susceptibility to infection |
| 137 | NA | Russell[150] | No | 19-May | UK | Single-centre RC | 29 Feb to 12 May | Research /clinical databases | 156 | Cancer patients with COVID19 | 50% White, 22% Black 4% Asian, 24% other | 65 ± 15 | 58 | 47 | 22 | NI | ACE/ARBs, β-blockers | Severity (mild/moderate vs severe - WHO guidelines) |
| 138 | NA | Sivaloganathan[151] | Yes | 25-Jun | UK | Multicenter case-control | 7 Mar to 9 Apr | Patient Administration System | 93 | ICU COVID-19 patients | NI | 80 ± 9 | NI | NI | NI | NI | Anticoagulants, antiplatelets | Severity (ICU admission) |
| 139 | NA | McKeigue[152] | No | 27-Jul | UK (Scotland) | Population-based case-control | Up to 6 Jun | Scottish Intensive Care Societ and Audit Group (SICSAG) database and National | 36391 | COVID-19 patients. All matches had to be at risk for Covid-19 | NI | NI | NI | NI | NI | NI | ACEIs/ARBs, alpha blockers, antiarrhythmics, antianginals, anticoagulants, antiplatelets, beta-blockers, centrally-acting antihypertensiv | Susceptibility to infection - Severity (entry to critical care or fatal outcome within 28 days) as a proxy |

| | | | | | | | | | | | | | | | | | | |
|---|---|---|---|---|---|---|---|---|---|---|---|---|---|---|---|---|---|---|
| | | | | | | | | Register of Scotland | | | | | | | | | es, cardiac glycosides, CCBs, diuretics, LMDs, vasodilators | |
| 140 | NA | Argenziano[153] | Yes | 29-May | USA | Retrospective case series | 1 Mar to 5 Apr | EHR | 1000 | Consecutive COVID-19 patients | 14% White, 25% Hispano/ Latino, 18% Black, 2% Asian, 41% other | 63 (50–75) | 60 | 54 | 55 | 58 | ACEIS, ARBs, LMDs | Severity (ICU admission) |
| 141 | NA | Auld[154] | Yes | 22 May (posted on medrxiv on 26 Apr) | USA | Multicentre RC | 6 Mar to 17 Apr | EHR | 209 | Adult COVID-19 patients in ICU | 18% White, 70% Black, 3% Asian, 9% other | 64 (54–73) [for 217] | 55 | 62 | 45 | <9 | Vasopressors, inhaled vasodilators | Mortality |
| 142 | UCLA Health System | Bae[155] | Yes | 12-Jul | USA | Single-centre RC | 1 Mar to 15 Apr | EMR | 590 | Consecutive adult COVID-19 patients | NI | 46 (33–60) | 49 | 25 | 26 | 19 | ACEi/ARB | Hospitalization, hospitalization length, severity (mechanical ventilation or ICU admission), mortality |
| 143 | NA | Bramante[156] | No | 20-Jun | USA | Nation-wide RC | 1 Jan to 7 Jun | De-identified claims data | 6256 | Hospitalized adult (≥18 years) COVID-19 Patients with T2DM or obesity | NI | 75 ± 12 | 47 | 59 | 100 | 8 | ACEi/ARB (added) | Mortality |
| 144 | NA | Castro[157] | No | 16-Apr | USA | Multicentre RC | 1 Mar to 7 Apr | EHR | 2271 | COVID-19 patients | 4% Asian, 17% Black, 16% other, 20% unknown, 44% White | 52 ± 20 | 46 | NI | NI | NI | ACEIs, ARBs, antiplatelets, anticoagulants, β-blockers, CCBs, diuretics, sympathomimetics, LMDs, vasodilators | Hospitalization, severity (ventilation) |
| 145 | UCLA Health System | Chang[158] | No | 4-Jul | USA | Multicentre retrospective case-control | 9 Mar to 14 Jun | EHR | 24633 | COVID-19 tested individuals | 55% White, 7% Black, 9% Asian, 29% Other | 49 ± 20 [for 26602] | 44 [for 26602] | NI | NI | NI | ACEi/ARB, anticoagulants | Susceptibility to infection, hospitalization, severity (ICU admission or intubation) |

| | | | | | | | | | | | | | | | | | | |
|---|---|---|---|---|---|---|---|---|---|---|---|---|---|---|---|---|---|---|
| | | | | | | | | | | | [for 26602] | | | | | | | |
| 146 | NA | Dublin[159] | No | 7-Jul | USA | Population cohort study | 29 Feb to 14 June | EHR | 322044 | Adults ≥ 18 years enrolled in Kaiser Permanente Washington in February 2020 (14,547 people tested) | 74% Non-Hispanic White, 5% Non-Hispanic Black, 11% Non-Hispanic Asian, 4% Non-Hispanic mixed race/other, 6% Hispanic | 51 ± 18 | 46 | 21 | 9 | 37 | ACEi/ARB, beta-blockers, CCB, diuretics | Susceptibility to infection, hospitalization |
| 147 | NA | Ebinger[4] | Yes | 23 Jul (posted on medrxiv on 5 May) | USA | Multicentre RC | 8 Mar to 21 Mar | EHR | 442 | COVID-19 patients | 64% White, 13% Black, 8% Asian, 15% other | 53 ± 20 | 58 | 36 | 19 | 16 | ACEIs, ARBs | Hospitalization, severity (severe/critical illness - based on hospitalization, ICU and intubation and mechanical ventilation) |
| 148 | NA | Ferguson [160] | Yes | 14-May | USA | Multicentre retrospective chart review | 13 Mar to 11 Apr | EHR | 72 | Consecutive adult hospitalized COVID-19 patients | 31% Hispanic or Latino, 28% Asian, 26% white, 6% black, 10% other | 60 (43–71) | 53 | 36 | 28 | NI | Any CVD drug, ACEIS, ARBs, β-blockers, CCB, diuretics, LMDs, digoxin | Severity (ICU admission) |
| 149 | NA | Garibaldi [14] | No | 26-May | USA | Multicentre RC | 4 Mar to 24 Apr | COVID-19 registry | 747 | Consecutive hospitalized COVID-19 patients | 33% White, 16% Hispanic, 6% Asian, 39% Black, 5% other | 63 (49–75) for 832 | 53 | 48 | 29 | NI | ACEIs, ARBs, anticoagulants, LMDs | Severity (mild/moderate vs severe/dead - based on who guidelines), mortality |
| 150 | NA | Goshua[161] | Yes | 30-Jun | USA | Single-centre cross-sectional study | 13 Apr to 24 Apr | Medical records | 68 | Hospitalised adult (≥18 years) COVID-19 patients | 24% Black, 51% White, 24% | 62 ± 16 | 60 | 56 | 29 | 37 | ACEi/ARB, statins | Severity (ICU) |

| | | | | | | | | | | | | | | | | | | |
|---|---|---|---|---|---|---|---|---|---|---|---|---|---|---|---|---|---|---|
| | | | | | | | | | | | Hispanic, 1% Asian | | | | | | | |
| 151 | NA | Gu[162] | No | 18-Jun | USA | Multicentre RC | 10 Mar to 22 Apr | EHR | 4412 (subset) | Patients tested for COVID-19 | 59% White, 17% Black, 24% other (for 5698) | 47 ± 21 (for 5698) | 38 (for 5698) | NI | 20 | NI | Anticoagulants, LMDs | Susceptibility to infection, hospitalization, severity (ICU admission), mortality |
| 152 | Other USA studies | Gupta[163] | Yes | 15-Jul | USA | Multicenter RC | 4 Mar to 4 Apr | EMR | 2215 | Consecutive adult patients (≥18 years) ICU COVID-19 patients | 38% White, 30% Black, 6% Asian, 20% Hispanic, 26% Other | 61 ± 15 | 65 | 60 | 39 | NI | ACEI/ARB, anticoagulants, antiplatelets, statins, mineralocorticoid receptor antagonist, vasopressors | 28-day mortality |
| 153 | NA | Hippensteel[164] | Yes | 24-Jun | USA | Single-center RC | 18 Mar to 14 Apr | EHR | 91 | Adult (≥18 years) ICU COVID-19 patients | NI | 56 ± 16 | 58 | NI | 31 | NI | Vasopressor | Mortality |
| 154 | NA | Imam[165] | Yes | 4-Jun | USA | Multicentre RC | 1 Mar to 17 Apr | EMR | 1305 | Hospitalized COVID-19 Patients | 66 African American, 27% Caucasian and 7% other | 61 ± 16 | 54 | 56 | 30 | NI | ACEi/ARB | Mortality |
| 155 | NA | Ip[166] | No | 29-Apr | USA | Multicentre RC | NI | EHR | 1129 (subset) | Hospitalized COVID-19 patients with HTN | NI | NR | NR | 100 | NI | NI | ACEIs, ARBs | Mortality |
| 156 | NA | Jillella[167] | No | 26-May | USA | Multicentre case-series | 24 Mar to 5 May | Patient records | 8 | Adult hospitalized COVID-19 patients with an acute ischemic stroke | 63% Black 25% White, 13% other | 64 ± 6 | 63 | 88 | 75 | NI | ACEIs, ARBs | Severity (ICU admission) |
| 157 | Other USA studies | Khera[168] | No | 19-May | USA | Nation-wide RC | 6 Mar to 3 May | Health insurance database and hospital healthcare records | 2263 | Out-patient adult COVID-19 patients with HTN | 38% White, 19% Black, 3% Hispanic, 2% Asian, 38% other | 69 (59–78) | 47 | 100 | 68 | NI | ACEIS, ARBs | Hospitalization, mortality |
| | | | | | | | | | 7933 | Hospitalized adult COVID-19 patients with HTN | 57% White, 28% Black, 3% | 77 (69–85) | 45 | 100 | 90 | NI | | |

| | | | | | | | | | | | | | | | | | | |
|---|---|---|---|---|---|---|---|---|---|---|---|---|---|---|---|---|---|---|
| | | | | | | | | | | | Hispanic, 2% Asian, 12% other | | | | | | | |
| 158 | Other USA studies | Kim[169] | Yes | 16 Jul (posted on medrxiv on 22 May) | USA | Multicentre RC | 1 Mar to 2 May | Medical chart reviews | 2491 | Hospitalized adult COVID-19 patients | 47% White, 30% Black 12% Hispanic, 10% other | 62 (50–75) | 53 | 57 | 33 | 50 | ACEIs, ARBs | Severity (ICU admission), mortality |
| 159 | NA | King[170] | No | 18-Jul | USA | Multicentre RC | 5 Mar to 26 Apr | EMR | 165 | COVID-19 Patients Managed with Invasive Mechanical Ventilation | 35% White, 24% Black 14% Asian 27% other | 58 (50–68) | 65 | 52 | 35 | 52 | Inhaled Pulmonary Vasodilators | Mortality |
| 160 | NA | Lala[171] | Yes | 8-Jun | USA | Multicentre RC | 27 Feb to 12 Apr | EHR | 2736 | Hospitalized adult (≥18 years) COVID-19 patients | 23% White, 26% African American 4% Asian, 47% other | median 66 | 60 | 39 | 26 | NI | ACEi/ARB, statins | Mortality |
| 161 | NA | Lam[172] | Yes | 23-Jul | USA | Single-centre RC | 7 Feb to 23 May | EMR | 614 | Hospitalized COVID-19 patients with HTN | 61% White, 8% African American 4% Asian, 27% other | 70 ± 12 | 55 | 10 0 | 41 | NI | ACEIs/ARBs | Severity (ICU admission or death), mortality |
| 162 | NA | Lobelo[173] | No | 10-Jul | USA | Multicentre retrospective case series | 3 Mar to 12 May | EHR | 3937 | Participants suspected to have COVID-19 | 55% Black 25% Whit and 20% Other | NI | NI | NI | NI | NI | LMDs | Susceptibility to infection - hospitalization as a proxy |
| 163 | NA | Lubetzky[174] | No | 5-May | USA | Single-centre retrospective chart review | 13 Mar to 20 Apr | EHR | 54 | Adult kidney transplant patients with COVID-19 | 31% White, 31% Hispanic, 24% Black 11% Asian 2% Middle Eastern | 57 (range 29–83) | 70 | 93 | 30 | NI | ACEIs, ARBs, antihyperten sives | Hospitalization |
| 164 | NA | Mehta[25] | Yes | 5-May | USA | Multicentre RC | 8 Mar to 12 Apr | EHR | 18472 | Patients tested for COVID-19 | 69% White, 20% Black 11% other | 49 ± 21 | 40 | 39 | 18 | 27 | ACEIS, ARBs | Susceptibility to infection, hospitalization, severity (ICU admission, ventilator use), mortality |

| | | | | | | | | | | | | | | | | | | |
|---|---|---|---|---|---|---|---|---|---|---|---|---|---|---|---|---|---|---|
| 165 | Veterans Affairs | Morales[175] | No | 12-Jun | USA | Multicentre RC | NI | EHR and claims data | 1392999 | Patients with HTN | NI | NI | NI | 100 | NI | NI | ACEIs, ARBs, CCBs/thiazides | Susceptibility to infection |
| 166 | NA | Nguyen[176] | No | 29-Ju | USA | Single-center RC | 16 Mar to 16 Apr | EMR | 519 | Consecutive COVID-19 Patients | 87% Black, 7% White, 7% Other [for 689] | 55 (40–68) [for 689] | 43 [for 689] | 54 [for 689] | 31 | 44 [for 689] | ACEi/ARB, anticoagulants, antiplatelets, beta-blockers, CCB, diuretics, statins | Hospitalization, mortality |
| 167 | NA | Palaiodimos[177] | Yes | 16 May (posted on medrxiv on 9 May) | USA | Single-centre RC | 9 Mar to 22 Mar | EHR | 200 | Consecutive hospitalized COVID-19 patients | 51% Black, 35% Hispanic/ Latino, 15% other | 64 (50–74) | 49 | 76 | 40 | <23 | ACEIs, ARBs | Severity (oxygen requirements/intubation), mortality |
| 168 | Mount Sinai | Paranjpe[178] | Yes | 29-Jun | USA | Multicenter RC | 14 Mar to 11 Apr | NI | 2773 | Hospitalized COVID-19 patients | NI | NI | NI | NI | NI | NI | Anticoagulants | Mortality |
| 169 | NA | Ramachandran[179] | No | 14-Jul | USA | Single-center RC | 1 Mar to 25 Apr | Medical records | 295 | Hospitalized adult COVID-19 patients | 3% White, 71% African American, 13% Hispanic, 3% Asian, 10% unknown | 65 ± 15 | 55 | 71 | 45 | NI | Statins | Mortality |
| 170 | Veterans Affairs | Rentsch[26] | No | 14-Apr | USA | Multicentre RC | 8 Feb to 30 Mar | Electronic health record data | 3789 | Patients tested for COVID-19 | 56% White, 30% Black, 8% Latino, 6% other | 66 (61-71) | 90 | 65 | 38 | NI | ACEIs, ARBs | Susceptibility to infection, hospitalization, severity (ICU admission) |
| 171 | NA | Reyes[180] | No | 6-May | USA | Single-centre retrospective case study | 20 Mar to 31 Mar | Patient medical records | 217 | Hospitalized adult COVID-19 patients | 39% Black, 18% White, 2% Asian, 41% other | 61 ± 15 | 58 | 65 | 37 | NI | Anticoagulants | Mortality |
| 172 | NA | Reynolds[27] | Yes | 1-May | USA | Multicentre RC | 1 Mar to 15 Apr | EHR | 12594 | Patients tested for COVID-19 | 15% Black, 9% Asian, 47% White, | 49 (34–63) | 42 | 36 | 18 | NI | ACEIS, ARBs, CCBs, β-blockers, thiazides | Susceptibility to infection, severity (intensive care, mechanical ventilation, or death) |

| | | | | | | | | | | | | | | | | | | |
|---|---|---|---|---|---|---|---|---|---|---|---|---|---|---|---|---|---|---|
| | | | | | | | | | | | 30% other | | | | | | | |
| 173 | NA | Richardson[17] | Yes | 22-Apr | USA | Multicentre case series | 1 Mar to 4 Apr | EHR | 1366 (subgroup) | Consecutive hospitalized COVID-19 patients | 23% Black, 9% Asian, 40% White, 29% other (for 5441) | 63 (52–75) [for 5700] | 60 [for 5700] | 57 [for 5700] | 34 [for 5700] | 42 [for 5700] | ACE/ARBs | Hospitalization, severity (ICU admission and mechanical ventilation), mortality |
| 174 | NA | Rodriguez-Nava[181] | Yes | 14-Jul | USA | Single-centre RC | Mar to May | De-identified dataset | 87 | Adult ICU COVID-19 patients | NI | 68 (58–75) | 64 | NI | NI | NI | Statins | Mortality |
| 175 | Other USA studies | Schneeweiss[9] | No | 24-Jul | USA | Population cohort study | 1 Mar to 30 May | EMR and claims data | 24708 | COVID-19 commercially insured patients on ACEis, ARBs or CCBs (aged 40+ and free of chronic kidney disease) | NI | 67 ± 14 | 48 | 36 | 21 | <12 | ARB/ACEI, CCB | Hospitalization, severity (hospitalization, ARDS, intubation or mechanical ventilation) |
| 176 | NA | Shah[182] | No | 6-May | USA | Single-centre RC | 3 Feb to 31 Mar | EHR | 316 | Adult patients tested for COVID-19 | 42% White, 16% Black 26% Asian 16% other | 63 (50–75) for 33; 62 (43–72) for 283 | 52 | 42 | NI | 3 | ACE/ARBs | Susceptibility to infection |
| 177 | NA | Solaimanzadeh[183] | Yes | 12-May | USA | Single-centre RC | 27 Feb to 13 Apr | EHR | 65 | Hospitalized COVID-19 ≥65 years old. | 77% African American, 23% other | mean 75 (range 65–89) | 49 | 86 | 58 | NI | CCB | Severity (intubation and mechanical ventilation), mortality |
| 178 | Mount Sinai | Wang[184] | Yes | 14 Jul (first posted 29 Jun) | USA | Multicentre RC | 1 Mar to 30 Apr | EMR | 58 | COVID-19 patients with multiple myeloma | 63% non-White | median 67 (IQR 13) | 52 | 64 | 28 | 37 | ACEi/ARB, beta blocker, anticoagulation, aspirin, statins | Hospitalization, mortality |

Abbreviations: ACEI = angiotensin-converting enzyme inhibitor, ARB = angiotensin receptor blocker, ARNI = angiotensin receptor-neprilysin inhibitors, CPAP = continuous positive airway pressure, CRRT = continuous renal-replacement therapy, CVD = cardiovascular disease, DM = diabetes mellitus, ECMO = extracorporeal membrane oxygenation, EHR = Electronic health records, HIRA = Health Insurance Review and Assessment Service, HTN = hypertension, LMD = lipid modifying drugs, MENA = Middle East North Africa, NA = not applicable, NI = no information, ICU = intensive care unit, PC = prospective cohort, RC = retrospective cohort, SARS = severe acute respiratory syndrome, TAVR = transcatheter aortic valve replacement. Age is mean ± SD or median (IQR), unless otherwise stated. Obesity is defined as BMI >=30.

**Table S3. Simple meta-regression for the association between testing positive for COVID-19 and taking an angiotensin-converting enzyme inhibitor or angiotensin receptor blocker**

| Variable | Estimate | SE | 95% CI | p-value |
|---|---|---|---|---|
| Publication | | | | |
| Pre-print | Reference | | | |
| Peer-review | 0.1246 | 0.1142 | -0.0993; 0.3485 | 0.2754 |
| Country | | | | |
| China | Reference | | | |
| Other | 0.6019 | 0.2484 | 0.1149; 1.0888 | ***0.0154*** |
| Country | | | | |
| France | Reference | | | |
| Other | -0.3979 | 0.2490 | -0.8860; 0.0902 | 0.1101 |
| Country | | | | |
| Italy | Reference | | | |
| Other | 0.1123 | 0.1785 | -0.2376; 0.4622 | 0.5294 |
| Country | | | | |
| Spain | Reference | | | |
| Other | -0.0758 | 0.2056 | -0.4788; 0.3272 | 0.7124 |
| Country | | | | |
| UK | Reference | | | |
| Other | 0.2387 | 0.1947 | -0.1429; 0.6204 | 0.2202 |
| Country | | | | |
| USA | Reference | | | |
| Other | -0.1534 | 0.1240 | -0.3964; 0.0897 | 0.2161 |
| Number of centres | | | | |
| Multiple | Reference | | | |
| Single | 0.3943 | 0.1398 | 0.1202; 0.6684 | ***0.0048*** |
| Design | | | | |
| Case-control | Reference | | | |
| Cohort | 0.3827 | 0.1252 | 0.1372; 0.6282 | ***0.0022*** |
| Setting | | | | |
| In-patient and other | Reference | | | |
| Only in-patient | -0.0832 | 0.2114 | -0.4976; 0.3311 | 0.6938 |
| Sample size | 0.0000 | 0.0000 | -0.0000; 0.0000 | 0.3907 |
| Age | -0.0062 | 0.0063 | -0.0185; 0.0062 | 0.3270 |
| Male % | -0.0033 | 0.0053 | -0.0137; 0.0070 | 0.5269 |
| Hypertension % | -0.0020 | 0.0024 | -0.0067; 0.0027 | 0.3998 |
| Diabetes mellitus | -0.0080 | 0.0073 | -0.0222; 0.0063 | 0.2745 |
| COVID-19 testing | | | | |
| All controls tested | Reference | | | |
| Not all controls tested | -0.1794 | 0.1304 | -0.4351; 0.0763 | 0.1691 |

**Table S4. Simple meta-regression for the association between testing positive for COVID-19 and taking an angiotensin-converting enzyme inhibitor**

| Variable | Estimate | SE | 95% CI | p-value |
|---|---|---|---|---|
| Publication<br>Pre-print<br>Peer-review | <br>Reference<br>0.2014 | <br><br>0.1189 | <br><br>-0.0315; 0.4344 | <br><br>0.0902 |
| Country<br>France<br>Other | <br>Reference<br>-0.4665 | <br><br>0.3796 | <br><br>-1.2105; 0.2775 | <br><br>0.2191 |
| Country<br>Italy<br>Other | <br>Reference<br>0.0812 | <br><br>0.1694 | <br><br>-0.2509; 0.4132 | <br><br>0.6319 |
| Country<br>Spain<br>Other | <br>Reference<br>-0.0499 | <br><br>0.1860 | <br><br>-0.4143; 0.3146 | <br><br>0.7886 |
| Country<br>UK<br>Other | <br>Reference<br>0.1598 | <br><br>0.1667 | <br><br>-0.1670; 0.4866 | <br><br>0.3379 |
| Country<br>USA<br>Other | <br>Reference<br>-0.0492 | <br><br>0.1312 | <br><br>-0.3064; 0.2081 | <br><br>0.7079 |
| Number of centres<br>Multiple<br>Single | <br>Reference<br>0.3757 | <br><br>0.1568 | <br><br>0.0685; 0.6830 | <br><br>***0.0165*** |
| Design<br>Case-control<br>Cohort | <br>Reference<br>0.2234 | <br><br>0.1309 | <br><br>-0.0332; 0.4799 | <br><br>0.0879 |
| Setting<br>In-patient and other<br>Only in-patient | <br>Reference<br>-0.0910 | <br><br>0.2433 | <br><br>-0.5678; 0.3858 | <br><br>0.7084 |
| Sample size | 0.0000 | 0.0000 | -0.0000; 0.0000 | 0.4872 |
| Age | -0.0089 | 0.0063 | -0.0212; 0.0034 | 0.1553 |
| Male % | -0.0021 | 0.0050 | -0.0118; 0.0077 | 0.6775 |
| Hypertension % | -0.0033 | 0.0022 | -0.0076; 0.0010 | 0.1330 |
| Diabetes mellitus | -0.0081 | 0.0078 | -0.0233; 0.0071 | 0.2961 |
| COVID-19 testing<br>All controls tested<br>Not all controls tested | <br>Reference<br>-0.1787 | <br><br>0.1238 | <br><br>-0.4214; 0.0639 | <br><br>0.1487 |

**Table S5. Simple meta-regression for the association between testing positive for COVID-19 and taking an angiotensin receptor blocker**

| Variable | Estimate | SE | 95% CI | p-value |
|---|---|---|---|---|
| Publication<br>Pre-print<br>Peer-review | <br>Reference<br>0.2593 | <br><br>0.1169 | <br><br>0.0302; 0.4885 | <br><br>***0.0266*** |
| Country<br>France<br>Other | <br>Reference<br>-0.6325 | <br><br>0.3842 | <br><br>-1.3856; 0.1206 | <br><br>0.0998 |
| Country<br>Italy<br>Other | <br>Reference<br>0.0187 | <br><br>0.2030 | <br><br>-0.3792; 0.4166 | <br><br>0.9265 |
| Country<br>Spain<br>Other | <br>Reference<br>-0.1246 | <br><br>0.2249 | <br><br>-0.5653; 0.3161 | <br><br>0.5794 |
| Country<br>UK<br>Other | <br>Reference<br>0.2261 | <br><br>0.2212 | <br><br>-0.2075; 0.6598 | <br><br>0.3067 |
| Country<br>USA<br>Other | <br>Reference<br>-0.1965 | <br><br>0.1503 | <br><br>-0.4912; 0.0981 | <br><br>0.1910 |
| Number of centres<br>Multiple<br>Single | <br>Reference<br>0.3843 | <br><br>0.1847 | <br><br>0.0223; 0.7463 | <br><br>***0.0375*** |
| Design<br>Case-control<br>Cohort | <br>Reference<br>0.3624 | <br><br>0.1318 | <br><br>0.1041; 0.6207 | <br><br>***0.0060*** |
| Setting<br>In-patient and other<br>Only in-patient | <br>Reference<br>-0.0745 | <br><br>0.3157 | <br><br>-0.6933; 0.5443 | <br><br>0.8134 |
| Sample size | 0.0000 | 0.0000 | -0.0000; 0.0000 | 0.7827 |
| Age | -0.0005 | 0.0080 | -0.0161; 0.0151 | 0.9515 |
| Male % | -0.0010 | 0.0064 | -0.0135; 0.0115 | 0.8724 |
| Hypertension % | 0.0012 | 0.0027 | -0.0042; 0.0066 | 0.6697 |
| Diabetes mellitus | -0.0028 | 0.0087 | -0.0199; 0.0143 | 0.7478 |
| COVID-19 testing<br>All controls tested<br>Not all controls tested | <br>Reference<br>-0.1697 | <br><br>0.1638 | <br><br>-0.4907; 0.1514 | <br><br>0.3003 |

**Table S6. Simple meta-regression for the association between testing positive for COVID-19 and being on a lipid modifying drug**

| Variable | Estimate | SE | 95% CI | p-value |
|---|---|---|---|---|
| Publication<br>Pre-print<br>Peer-review | <br>Reference<br>0.1967 | <br><br>0.2532 | <br><br>-0.2995; 0.6930 | <br><br>0.4372 |
| Country<br>France<br>Other | <br>Reference<br>0.1262 | <br><br>0.4213 | <br><br>-0.6996; 0.9519 | <br><br>0.7645 |
| Country<br>UK<br>Other | <br>Reference<br>-0.0418 | <br><br>0.2037 | <br><br>-0.4410; 0.3574 | <br><br>0.8373 |
| Number of centres<br>Multiple<br>Single | <br>Reference<br>-0.1262 | <br><br>0.4213 | <br><br>-0.9519; 0.6996 | <br><br>0.7645 |
| Design<br>Case-control<br>Cohort | <br>Reference<br>-0.0248 | <br><br>0.2072 | <br><br>-0.4310; 0.3813 | <br><br>0.9046 |
| Setting<br>In-patient and other<br>Only in-patient | <br>Reference<br>-0.1188 | <br><br>0.2716 | <br><br>-0.6512; 0.4136 | <br><br>0.6618 |
| Sample size | 0.0000 | 0.0000 | -0.0000; 0.0000 | 0.2909 |
| Age | -0.0036 | 0.0098 | -0.0227; 0.0156 | 0.7149 |
| Male % | 0.0018 | 0.0131 | -0.0238; 0.0275 | 0.8894 |
| Hypertension % | -0.0045 | 0.0070 | -0.0182; 0.0092 | 0.5220 |
| Diabetes mellitus | -0.0306 | 0.0105 | -0.0510; -0.0101 | ***0.0035*** |
| COVID-19 testing<br>All controls tested<br>Not all controls tested | <br>Reference<br>0.3263 | <br><br>0.1405 | <br><br>0.0510; 0.6017 | <br><br>***0.0202*** |

**Table S7. Simple meta-regression for the association between being hospitalized for COVID-19 and taking an angiotensin-converting enzyme inhibitor or angiotensin receptor blocker**

| Variable | Estimate | SE | 95% CI | p-value |
|---|---|---|---|---|
| Publication<br>Pre-print<br>Peer-review | <br>Reference<br>-0.0291 | <br><br>0.2890 | <br><br>-0.5954; 0.5373 | <br><br>0.9199 |
| Country<br>Italy<br>Other | <br>Reference<br>-0.0139 | <br><br>0.4777 | <br><br>-0.9501; 0.9224 | <br><br>0.9769 |
| Country<br>Spain<br>Other | <br>Reference<br>0.7362 | <br><br>0.4618 | <br><br>-0.169; 1.6414 | <br><br>0.1109 |
| Country<br>USA<br>Other | <br>Reference<br>-0.5019 | <br><br>0.2984 | <br><br>-1.0868; 0.0830 | <br><br>0.0926 |
| Number of centres<br>Multiple<br>Single | <br>Reference<br>-0.4466 | <br><br>0.3044 | <br><br>-1.0433; 0.1501 | <br><br>0.1424 |
| Design<br>Case-control<br>Cohort | <br>Reference<br>-0.2974 | <br><br>0.6350 | <br><br>-1.542; 0.9471 | <br><br>0.6395 |
| Sample size | 0.0001 | 0.0001 | 0.0000; 0.0002 | 0.1006 |
| Age | -0.0455 | 0.0185 | -0.0818; -0.0092 | ***0.0140*** |
| Male % | -0.0174 | 0.0100 | -0.0371; 0.0023 | 0.0830 |
| Hypertension % | -0.0197 | 0.0054 | -0.0304; -0.009 | ***0.0003*** |
| Diabetes mellitus | -0.0205 | 0.0113 | -0.0426; 0.0016 | 0.0688 |

**Table S8. Simple meta-regression for the association between being hospitalized for COVID-19 and taking an angiotensin-converting enzyme inhibitor**

| Variable | Estimate | SE | 95% CI | p-value |
|---|---|---|---|---|
| Publication<br>Pre-print<br>Peer-review | <br>Reference<br>0.5204 | <br><br>0.5762 | <br><br>-0.6088; 1.6497 | <br><br>0.3664 |
| Country<br>Italy<br>Other | <br>Reference<br>-0.2993 | <br><br>0.9816 | <br><br>-2.2233; 1.6246 | <br><br>0.7604 |
| Country<br>Spain<br>Other | <br>Reference<br>0.8563 | <br><br>0.7639 | <br><br>-0.6408; 2.3535 | <br><br>0.2623 |
| Country<br>USA<br>Other | <br>Reference<br>-0.9365 | <br><br>0.6293 | <br><br>-2.1699; 0.297 | <br><br>0.1367 |
| Number of centres<br>Multiple<br>Single | <br>Reference<br>-0.3093 | <br><br>0.6099 | <br><br>-1.5046; 0.886 | <br><br>0.6121 |
| Design<br>Case-control<br>Cohort | <br>Reference<br>-0.3291 | <br><br>1.0506 | <br><br>-2.3883; 1.7301 | <br><br>0.7541 |
| Sample size | 0.0004 | 0.0004 | -0.0003; 0.0012 | 0.2890 |
| Age | -0.0474 | 0.0441 | -0.1338; 0.039 | 0.2820 |
| Male % | -0.0046 | 0.0184 | -0.0406; 0.0314 | 0.8031 |
| Hypertension % | -0.0140 | 0.0131 | -0.0398; 0.0117 | 0.2861 |
| Diabetes mellitus | -0.0048 | 0.0308 | -0.0653; 0.0556 | 0.8754 |

**Table S9. Simple meta-regression for the association between being hospitalized for COVID-19 and taking an angiotensin receptor blocker**

| Variable | Estimate | SE | 95% CI | p-value |
|---|---|---|---|---|
| Publication<br>Pre-print<br>Peer-review | 0.1977 | 0.6769 | -1.129; 1.5243 | 0.7703 |
| Country<br>Italy<br>Other | 0.9300 | 1.0227 | -1.0745; 2.9345 | 0.3632 |
| Country<br>Spain<br>Other | 0.6628 | 0.8700 | -1.0423; 2.3678 | 0.4462 |
| Country<br>USA<br>Other | -1.0987 | 0.664 | -2.4001; 0.2027 | 0.0980 |
| Number of centres<br>Multiple<br>Single | -0.7990 | 0.6688 | -2.1099; 0.5119 | 0.2322 |
| Design<br>Case-control<br>Cohort | -0.1529 | 1.1673 | -2.4408; 2.135 | 0.8958 |
| Sample size | 0.0007 | 0.0004 | -0.0001; 0.0016 | 0.0706 |
| Age | -0.1042 | 0.0406 | -0.1838; -0.0245 | ***0.0104*** |
| Male % | -0.0252 | 0.0198 | -0.0641; 0.0137 | 0.2044 |
| Hypertension % | -0.0274 | 0.0132 | -0.0532; -0.0015 | ***0.0379*** |
| Diabetes mellitus | -0.0316 | 0.0330 | -0.0963; 0.0330 | 0.3375 |

**Table S10. Simple meta-regression for the association between hospitalization length and taking an angiotensin-converting enzyme inhibitor or angiotensin receptor blocker**

| Variable | Estimate | SE | 95% CI | p-value |
|---|---|---|---|---|
| Publication<br>Pre-print<br>Peer-review | <br>Reference<br>0.8024 | <br><br>1.9052 | <br><br>-2.9318; 4.5366 | <br><br>0.6736 |
| Country<br>China<br>Other | <br>Reference<br>-0.1177 | <br><br>1.0301 | <br><br>-2.1367; 1.9013 | <br><br>0.9091 |
| Country<br>Turkey<br>Other | <br>Reference<br>1.8230 | <br><br>1.1718 | <br><br>-0.4736; 4.1195 | <br><br>0.1198 |
| Country<br>USA<br>Other | <br>Reference<br>-1.1245 | <br><br>1.0706 | <br><br>-3.2228; 0.9738 | <br><br>0.2936 |
| Number of centres<br>Multiple<br>Single | <br>Reference<br>0.2415 | <br><br>1.2607 | <br><br>-2.2294; 2.7123 | <br><br>0.8481 |
| Sample size | 0.0010 | 0.0015 | -0.0019; 0.0039 | 0.5018 |
| Age | -0.1392 | 0.0533 | -0.2437; -0.0346 | ***0.0091*** |
| Male % | -0.1866 | 0.2633 | -0.7026; 0.3294 | 0.4785 |
| Hypertension % | -0.0316 | 0.0125 | -0.0560; -0.0072 | ***0.0111*** |
| Diabetes mellitus | -0.1443 | 0.0834 | -0.3078; 0.0193 | 0.0838 |

**Table S11. Meta-regression for the association between severity outcomes in COVID-19 patients and taking an angiotensin-converting enzyme inhibitor or angiotensin receptor blocker**

| Variable | Estimate | SE | 95% CI | p-value |
|---|---|---|---|---|
| Publication | | | | |
| Pre-print | Reference | | | |
| Peer-review | 0.0033 | 0.1785 | -0.3466; 0.3532 | 0.9852 |
| Country | | | | |
| Belgium | Reference | | | |
| Other | 0.6952 | 0.5594 | -0.4011; 1.7915 | 0.2139 |
| Country | | | | |
| China | Reference | | | |
| Other | 0.5767 | 0.2011 | 0.1825; 0.9709 | ***0.0041*** |
| Country | | | | |
| France | Reference | | | |
| Other | -0.0835 | 0.2720 | -0.6166; 0.4496 | 0.7588 |
| Country | | | | |
| Italy | Reference | | | |
| Other | 0.0151 | 0.2862 | -0.5458; 0.576 | 0.9579 |
| Country | | | | |
| South Korea | Reference | | | |
| Other | -1.0567 | 0.4810 | -1.9994; -0.114 | ***0.0280*** |
| Country | | | | |
| Spain | Reference | | | |
| Other | 0.3334 | 0.2745 | -0.2046; 0.8715 | 0.2245 |
| Country | | | | |
| Turkey | Reference | | | |
| Other | -0.2765 | 0.4455 | -1.1497; 0.5966 | 0.5348 |
| Country | | | | |
| UK | Reference | | | |
| Other | 0.7367 | 0.4312 | -0.1085; 1.5818 | 0.0876 |
| Country | | | | |
| USA | Reference | | | |
| Other | -0.3762 | 0.2013 | -0.7707; 0.0183 | 0.0616 |
| Number of centres | | | | |
| Multiple | Reference | | | |
| Single | -0.2671 | 0.1625 | -0.5855; 0.0513 | 0.1001 |
| Design | | | | |
| Case-control | Reference | | | |
| Cohort | -0.7034 | 0.5237 | -1.7299; 0.323 | 0.1792 |
| Setting | | | | |
| In-patient and other | Reference | | | |
| Only in-patient | -0.357 | 0.1609 | -0.6725; -0.0416 | ***0.0265*** |
| Sample size | 0.0001 | 0.0000 | 0.0001; 0.0002 | ***0.0011*** |
| Age | -0.0398 | 0.0080 | -0.0555; -0.0242 | ***<.0001*** |
| Male % | -0.0048 | 0.0080 | -0.0204; 0.0109 | 0.5505 |
| Hypertension % | -0.0119 | 0.0028 | -0.0173; -0.0065 | ***<.0001*** |
| Diabetes mellitus | -0.0218 | 0.0072 | -0.0359; -0.0077 | ***0.0025*** |

**Table S12. Meta-regression for the association between severity outcomes in COVID-19 patients and taking an angiotensin-converting enzyme inhibitor**

| Variable | Estimate | SE | 95% CI | p-value |
|---|---|---|---|---|
| Publication<br>Pre-print<br>Peer-review | <br>Reference<br>0.1191 | <br><br>0.2439 | <br><br>-0.3590; 0.5972 | <br><br>0.6253 |
| Country<br>China<br>Other | <br>Reference<br>0.4373 | <br><br>0.3245 | <br><br>-0.1988; 1.0733 | <br><br>0.1778 |
| Country<br>France<br>Other | <br>Reference<br>0.2581 | <br><br>0.4684 | <br><br>-0.6600; 1.1763 | <br><br>0.5816 |
| Country<br>Italy<br>Other | <br>Reference<br>0.1637 | <br><br>0.3413 | <br><br>-0.5052; 0.8326 | <br><br>0.6315 |
| Country<br>Spain<br>Other | <br>Reference<br>0.2465 | <br><br>0.3199 | <br><br>-0.3805; 0.8735 | <br><br>0.4410 |
| Country<br>USA<br>Other | <br>Reference<br>-0.4449 | <br><br>0.2591 | <br><br>-0.9528; 0.0630 | <br><br>0.0860 |
| Number of centres<br>Multiple<br>Single | <br>Reference<br>-0.4398 | <br><br>0.2252 | <br><br>-0.8811; 0.0015 | <br><br>0.0508 |
| Design<br>Case-control<br>Cohort | <br>Reference<br>-0.0277 | <br><br>0.4401 | <br><br>-0.8903; 0.8350 | <br><br>0.9499 |
| Setting<br>In-patient and other<br>Only in-patient | <br>Reference<br>-0.5169 | <br><br>0.2213 | <br><br>-0.9506; -0.0831 | <br><br>***0.0195*** |
| Sample size | 0.0001 | 0.0001 | -0.0000; 0.0003 | 0.1109 |
| Age | -0.0438 | 0.0147 | -0.0725; -0.0150 | ***0.0029*** |
| Male % | -0.0065 | 0.0106 | -0.0273; 0.0142 | 0.5366 |
| Hypertension % | -0.0141 | 0.0040 | -0.0220; -0.0062 | ***0.0004*** |
| Diabetes mellitus | -0.0262 | 0.0122 | -0.0502; -0.0023 | ***0.0319*** |

**Table S13. Meta-regression for the association between severity outcomes in COVID-19 patients and taking an angiotensin receptor blocker**

| Variable | Estimate | SE | 95% CI | p-value |
|---|---|---|---|---|
| Publication<br>Pre-print<br>Peer-review | <br>Reference<br>0.4147 | <br><br>0.2223 | <br><br>-0.0210; 0.8505 | <br><br>0.0621 |
| Country<br>China<br>Other | <br>Reference<br>0.7588 | <br><br>0.2898 | <br><br>0.1908; 1.3267 | <br><br>***0.0088*** |
| Country<br>France<br>Other | <br>Reference<br>-0.1668 | <br><br>0.6909 | <br><br>-1.5210; 1.1873 | <br><br>0.8092 |
| Country<br>Italy<br>Other | <br>Reference<br>-0.0342 | <br><br>0.3231 | <br><br>-0.6676; 0.5992 | <br><br>0.9157 |
| Country<br>Spain<br>Other | <br>Reference<br>0.2667 | <br><br>0.3036 | <br><br>-0.3284; 0.8617 | <br><br>0.3797 |
| Country<br>USA<br>Other | <br>Reference<br>-0.4977 | <br><br>0.2479 | <br><br>-0.9836; -0.0119 | <br><br>***0.0446*** |
| Number of centres<br>Multiple<br>Single | <br>Reference<br>-0.2447 | <br><br>0.2188 | <br><br>-0.6736; 0.1842 | <br><br>0.2635 |
| Design<br>Case-control<br>Cohort | <br>Reference<br>0.1156 | <br><br>0.3738 | <br><br>-0.6170; 0.8482 | <br><br>0.7571 |
| Setting<br>In-patient and other<br>Only in-patient | <br>Reference<br>-0.5165 | <br><br>0.2121 | <br><br>-0.9322; -0.1007 | <br><br>***0.0149*** |
| Sample size | 0.0002 | 0.0001 | 0.0000; 0.0003 | ***0.0449*** |
| Age | -0.0386 | 0.0124 | -0.0629; -0.0143 | ***0.0018*** |
| Male % | -0.0102 | 0.0101 | -0.0300; 0.0096 | 0.3137 |
| Hypertension % | -0.0139 | 0.0037 | -0.0211; -0.0067 | ***0.0002*** |
| Diabetes mellitus | -0.0376 | 0.0124 | -0.0619; -0.0132 | ***0.0025*** |

**Table S14. Meta-regression for the association between severity outcomes in COVID-19 patients and taking an anticoagulant**

| Variable | Estimate | SE | 95% CI | p-value |
|---|---|---|---|---|
| Publication<br>Pre-print<br>Peer-review | <br>Reference<br>-0.4869 | <br><br>0.2976 | <br><br>-1.0702; 0.0964 | <br><br>0.1018 |
| Country<br>China<br>Other | <br>Reference<br>-0.6335 | <br><br>0.4467 | <br><br>-1.5092; 0.2421 | <br><br>0.1562 |
| Country<br>Italy<br>Other | <br>Reference<br>0.5771 | <br><br>0.4899 | <br><br>-0.383; 1.5373 | <br><br>0.2387 |
| Country<br>Spain<br>Other | <br>Reference<br>-0.4371 | <br><br>0.2861 | <br><br>-0.9978; 0.1237 | <br><br>0.1266 |
| Country<br>USA<br>Other | <br>Reference<br>0.3362 | <br><br>0.3697 | <br><br>-0.3884; 1.0608 | <br><br>0.3632 |
| Number of centres<br>Multiple<br>Single | <br>Reference<br>0.3638 | <br><br>0.3004 | <br><br>-0.2249; 0.9526 | <br><br>0.2258 |
| Design<br>Case-control<br>Cohort | <br>Reference<br>0.9263 | <br><br>0.5906 | <br><br>-0.2312; 2.0838 | <br><br>0.1168 |
| Setting<br>In-patient and other<br>Only in-patient | <br>Reference<br>0.1186 | <br><br>0.3027 | <br><br>-0.4746; 0.7119 | <br><br>0.6951 |
| Sample size | 0.0000 | 0.0000 | -0.0001; 0.0001 | 0.8404 |
| Age | 0.0138 | 0.0162 | -0.0178; 0.0455 | 0.3921 |
| Male % | -0.0011 | 0.0141 | -0.0289; 0.0266 | 0.9363 |
| Hypertension % | -0.0019 | 0.0114 | -0.0242; 0.0204 | 0.8698 |
| Diabetes mellitus | 0.0010 | 0.0200 | -0.0383; 0.0403 | 0.9609 |

**Table S15. Meta-regression for the association between severity outcomes in COVID-19 patients and taking an antiplatelet**

| Variable | Estimate | SE | 95% CI | p-value |
|---|---|---|---|---|
| Publication<br>Pre-print<br>Peer-review | <br>Reference<br>-1.2288 | <br><br>0.3165 | <br><br>-1.8492; -0.6085 | <br><br>***0.0001*** |
| Country<br>China<br>Other | <br>Reference<br>0.1919 | <br><br>0.6238 | <br><br>-1.0307; 1.4144 | <br><br>0.7584 |
| Country<br>Italy<br>Other | <br>Reference<br>0.5936 | <br><br>0.5431 | <br><br>-0.4708; 1.6581 | <br><br>0.2744 |
| Number of centres<br>Multiple<br>Single | <br>Reference<br>-0.6765 | <br><br>0.5544 | <br><br>-1.7631; 0.4101 | <br><br>0.2224 |
| Design<br>Case-control<br>Cohort | <br>Reference<br>-0.5256 | <br><br>1.3384 | <br><br>-3.1489; 2.0977 | <br><br>0.6945 |
| Setting<br>In-patient and other<br>Only in-patient | <br>Reference<br>-0.3467 | <br><br>0.5172 | <br><br>-1.3604; 0.6671 | <br><br>0.5027 |
| Sample size | 0.0001 | 0.0001 | 0.0000; 0.0002 | ***0.0166*** |
| Age | -0.0336 | 0.0241 | -0.0808; 0.0136 | 0.1626 |
| Male % | -0.0414 | 0.0256 | -0.0915; 0.0087 | 0.1053 |
| Hypertension % | -0.0292 | 0.0118 | -0.0524; -0.0061 | ***0.0134*** |
| Diabetes mellitus | -0.0381 | 0.0271 | -0.0913; 0.0152 | 0.1610 |

**Table S16. Meta-regression for the association between severity outcomes in COVID-19 patients and taking a beta-blocker**

| Variable | Estimate | SE | 95% CI | p-value |
|---|---|---|---|---|
| <u>Publication</u><br>Pre-print<br>Peer-review | <br>Reference<br>-0.3864 | <br><br>0.3036 | <br><br>-0.9815; 0.2088 | <br><br>0.2032 |
| <u>Country</u><br>China<br>Other | <br>Reference<br>0.1032 | <br><br>0.4816 | <br><br>-0.8407; 1.047 | <br><br>0.8304 |
| <u>Country</u><br>France<br>Other | <br>Reference<br>0.4241 | <br><br>0.3383 | <br><br>-0.2389; 1.0872 | <br><br>0.2099 |
| <u>Country</u><br>USA<br>Other | <br>Reference<br>-0.0626 | <br><br>0.4557 | <br><br>-0.9557; 0.8305 | <br><br>0.8907 |
| <u>Number of centres</u><br>Multiple<br>Single | <br>Reference<br>-0.2425 | <br><br>0.3648 | <br><br>-0.9576; 0.4725 | <br><br>0.5061 |
| <u>Design</u><br>Case-control<br>Cohort | <br>Reference<br>-0.5787 | <br><br>0.7574 | <br><br>-2.0632; 0.9059 | <br><br>0.4449 |
| <u>Setting</u><br>In-patient and other<br>Only in-patient | <br>Reference<br>-0.5095 | <br><br>0.2914 | <br><br>-1.0807; 0.0618 | <br><br>0.0805 |
| Sample size | 0.0001 | 0.0000 | 0.0001; 0.0002 | ***<.0001*** |
| Age | -0.0404 | 0.0098 | -0.0595; -0.0212 | ***<.0001*** |
| Male % | -0.0417 | 0.0112 | -0.0637; -0.0197 | ***0.0002*** |
| Hypertension % | -0.0156 | 0.004 | -0.0234; -0.0078 | ***0.0001*** |
| Diabetes mellitus | -0.0101 | 0.0036 | -0.0172; -0.003 | ***0.0055*** |

**Table S17. Meta-regression for the association between severity outcomes in COVID-19 patients and taking a calcium channel blocker**

| Variable | Estimate | SE | 95% CI | p-value |
|---|---|---|---|---|
| <u>Publication</u><br>Pre-print<br>Peer-review | <br>Reference<br>-0.3272 | <br><br>0.3427 | <br><br>-0.9988; 0.3445 | <br><br>0.3397 |
| <u>Country</u><br>China<br>Other | <br>Reference<br>0.6531 | <br><br>0.2827 | <br><br>0.0991; 1.2071 | <br><br>***0.0209*** |
| <u>Country</u><br>USA<br>Other | <br>Reference<br>-0.0027 | <br><br>0.4073 | <br><br>-0.8009; 0.7956 | <br><br>0.9948 |
| <u>Number of centres</u><br>Multiple<br>Single | <br>Reference<br>-0.273 | <br><br>0.2984 | <br><br>-0.8577; 0.3118 | <br><br>0.3603 |
| <u>Design</u><br>Case-control<br>Cohort | <br>Reference<br>1.0555 | <br><br>0.5396 | <br><br>-0.002; 2.113 | <br><br>0.0504 |
| <u>Setting</u><br>In-patient and other<br>Only in-patient | <br>Reference<br>-0.3419 | <br><br>0.2906 | <br><br>-0.9115; 0.2277 | <br><br>0.2394 |
| Sample size | 0.0001 | 0.0000 | 0.0000; 0.0002 | ***0.0021*** |
| Age | -0.0401 | 0.0124 | -0.0644; -0.0158 | ***0.0012*** |
| Male % | -9.00E-04 | 0.0215 | -0.0430; 0.0412 | 0.9669 |
| Hypertension % | -0.0122 | 0.0039 | -0.0198; -0.0045 | ***0.0019*** |
| Diabetes mellitus | -0.0448 | 0.0139 | -0.0721; -0.0175 | ***0.0013*** |

**Table S18. Meta-regression for the association between severity outcomes in COVID-19 patients and taking a lipid modifying drug**

| Variable | Estimate | SE | 95% CI | p-value |
|---|---|---|---|---|
| Publication<br>Pre-print<br>Peer-review | <br>Reference<br>-0.5547 | <br><br>0.2679 | <br><br>-1.0798; -0.0296 | <br><br>***0.0384*** |
| Country<br>China<br>Other | <br>Reference<br>0.5539 | <br><br>0.4991 | <br><br>-0.4243; 1.532 | <br><br>0.2671 |
| Country<br>USA<br>Other | <br>Reference<br>-0.0965 | <br><br>0.3881 | <br><br>-0.8571; 0.6642 | <br><br>0.8037 |
| Number of centres<br>Multiple<br>Single | <br>Reference<br>-0.0553 | <br><br>0.3921 | <br><br>-0.8238; 0.7132 | <br><br>0.8879 |
| Design<br>Case-control<br>Cohort | <br>Reference<br>-0.0414 | <br><br>0.8018 | <br><br>-1.6129; 1.5301 | <br><br>0.9588 |
| Setting<br>In-patient and other<br>Only in-patient | <br>Reference<br>0.2057 | <br><br>0.3673 | <br><br>-0.5142; 0.9256 | <br><br>0.5754 |
| Sample size | 0.0001 | 0.0000 | 0.0000; 0.0002 | ***0.0424*** |
| Age | -0.0404 | 0.0097 | -0.0594; -0.0214 | ***<.0001*** |
| Male % | -0.0018 | 0.0150 | -0.0311; 0.0275 | 0.9026 |
| Hypertension % | -0.0155 | 0.0066 | -0.0284; -0.0027 | ***0.0180*** |
| Diabetes mellitus | -0.0113 | 0.0045 | -0.0200; -0.0025 | ***0.0121*** |

**Table S19. Meta-regression for the association between mortality outcomes in COVID-19 patients on an angiotensin-converting enzyme inhibitor or angiotensin receptor blocker**

| Variable | Estimate | SE | 95% CI | p-value |
|---|---|---|---|---|
| Publication<br>Pre-print<br>Peer-review | <br>Reference<br>-0.3252 | <br><br>0.2722 | <br><br>-0.8588; 0.2083 | <br><br>0.2322 |
| Country<br>China<br>Other | <br>Reference<br>0.9381 | <br><br>0.2815 | <br><br>0.3864; 1.4899 | <br><br>***0.0009*** |
| Country<br>France<br>Other | <br>Reference<br>-0.4763 | <br><br>0.4645 | <br><br>-1.3868; 0.4341 | <br><br>0.3052 |
| Country<br>Italy<br>Other | <br>Reference<br>-0.1135 | <br><br>0.2746 | <br><br>-0.6517; 0.4247 | <br><br>0.6793 |
| Country<br>Spain<br>Other | <br>Reference<br>-0.2122 | <br><br>0.4000 | <br><br>-0.9961; 0.5717 | <br><br>0.5958 |
| Country<br>Turkey<br>Other | <br>Reference<br>-0.5603 | <br><br>0.6075 | <br><br>-1.7511; 0.6305 | <br><br>0.3564 |
| Country<br>UK<br>Other | <br>Reference<br>0.2352 | <br><br>0.4802 | <br><br>-0.7061; 1.1764 | <br><br>0.6243 |
| Number of centres<br>Multiple<br>Single | <br>Reference<br>-0.3792 | <br><br>0.2506 | <br><br>-0.8703; 0.1119 | <br><br>0.1302 |
| Setting<br>In-patient and other<br>Only in-patient | <br>Reference<br>-0.2315 | <br><br>0.2788 | <br><br>-0.778; 0.315 | <br><br>0.4064 |
| Sample size | 0.0001 | 0.0000 | 0.0000; 0.0002 | ***0.0073*** |
| Age | -0.0559 | 0.0109 | -0.0772; -0.0346 | ***<.0001*** |
| Male % | -0.0117 | 0.0116 | -0.0345; 0.0111 | 0.3141 |
| Hypertension % | -0.0150 | 0.0032 | -0.0213; -0.0088 | ***<.0001*** |
| Diabetes mellitus | -0.0103 | 0.0071 | -0.0241; 0.0035 | 0.1450 |

**Table S20. Simple meta-regression for the association between mortality outcomes in COVID-19 patients on an angiotensin-converting enzyme inhibitor**

| Variable | Estimate | SE | 95% CI | p-value |
|---|---|---|---|---|
| Publication<br>Pre-print<br>Peer-review | <br>Reference<br>-0.0081 | <br><br>0.2896 | <br><br>-0.5757; 0.5595 | <br><br>0.9777 |
| Country<br>China<br>Other | <br>Reference<br>0.2372 | <br><br>0.4372 | <br><br>-0.6197; 1.0941 | <br><br>0.5875 |
| Country<br>Italy<br>Other | <br>Reference<br>0.0353 | <br><br>0.3255 | <br><br>-0.6026; 0.6733 | <br><br>0.9136 |
| Country<br>Spain<br>Other | <br>Reference<br>-0.0162 | <br><br>0.4634 | <br><br>-0.9245; 0.8920 | <br><br>0.9720 |
| Number of centres<br>Multiple<br>Single | <br>Reference<br>-0.4800 | <br><br>0.3088 | <br><br>-1.0852; 0.1253 | <br><br>0.1201 |
| Setting<br>In-patient and other<br>Only in-patient | <br>Reference<br>-0.4199 | <br><br>0.2212 | <br><br>-0.8534; 0.0136 | <br><br>0.0576 |
| Sample size | 0.0001 | 0.0001 | -0.0001; 0.0002 | 0.2739 |
| Age | -0.0479 | 0.0093 | -0.0662; -0.0297 | ***<.0001*** |
| Male % | -0.0091 | 0.0135 | -0.0354; 0.0173 | 0.4999 |
| Hypertension % | -0.0117 | 0.0014 | -0.0144; -0.009 | ***<.0001*** |
| Diabetes mellitus | -0.0063 | 0.0033 | -0.0128; 0.0003 | 0.0607 |

**Table S21. Simple meta-regression for the association between mortality outcomes in COVID-19 patients on an angiotensin receptor blocker**

| Variable | Estimate | SE | 95% CI | p-value |
|---|---|---|---|---|
| Publication<br>Pre-print<br>Peer-review | <br>Reference<br>-0.5314 | <br><br>0.3673 | <br><br>-1.2513; 0.1885 | <br><br>0.1479 |
| Country<br>China<br>Other | <br>Reference<br>0.8462 | <br><br>0.5009 | <br><br>-0.1356; 1.828 | <br><br>0.0912 |
| Country<br>Italy<br>Other | <br>Reference<br>0.0961 | <br><br>0.3393 | <br><br>-0.5689; 0.7611 | <br><br>0.7771 |
| Country<br>Spain<br>Other | <br>Reference<br>-0.0508 | <br><br>0.4654 | <br><br>-0.9629; 0.8614 | <br><br>0.9131 |
| Number of centres<br>Multiple<br>Single | <br>Reference<br>-0.3106 | <br><br>0.3197 | <br><br>-0.9372; 0.316 | <br><br>0.3313 |
| Setting<br>In-patient and other<br>Only in-patient | <br>Reference<br>-0.0675 | <br><br>0.3472 | <br><br>-0.748; 0.6129 | <br><br>0.8457 |
| Sample size | 0.0001 | 0.0001 | 0.0000; 0.0002 | ***0.0471*** |
| Age | -0.0539 | 0.0122 | -0.0778; -0.0299 | ***<.0001*** |
| Male % | -0.0251 | 0.0146 | -0.0538; 0.0035 | 0.0856 |
| Hypertension % | -0.0136 | 0.0038 | -0.0211; -0.0061 | ***0.0004*** |
| Diabetes mellitus | -0.0056 | 0.0052 | -0.0157; 0.0045 | 0.2797 |

**Table S22. Simple meta-regression for the association between mortality outcomes in COVID-19 patients on anticoagulants**

| Variable | Estimate | SE | 95% CI | p-value |
|---|---|---|---|---|
| Publication<br>Pre-print<br>Peer-review | <br>Reference<br>-0.3663 | <br><br>0.7139 | <br><br>-1.7655; 1.0329 | <br><br>0.6079 |
| Country<br>China<br>Other | <br>Reference<br>0.0470 | <br><br>1.1653 | <br><br>-2.2371; 2.3310 | <br><br>0.9678 |
| Country<br>Italy<br>Other | <br>Reference<br>0.8549 | <br><br>0.8241 | <br><br>-0.7602; 2.4701 | <br><br>0.2995 |
| Country<br>USA<br>Other | <br>Reference<br>-0.0185 | <br><br>0.7082 | <br><br>-1.4065; 1.3695 | <br><br>0.9791 |
| Number of centres<br>Multiple<br>Single | <br>Reference<br>0.0028 | <br><br>0.7448 | <br><br>-1.457; 1.4626 | <br><br>0.9970 |
| Setting<br>In-patient and other<br>Only in-patient | <br>Reference<br>-1.4134 | <br><br>0.4613 | <br><br>-2.3175; -0.5092 | <br><br>***0.0022*** |
| Sample size | 0.0002 | 0.0001 | 0.0001; 0.0004 | ***0.0004*** |
| Age | -0.0556 | 0.0297 | -0.1137; 0.0026 | 0.0611 |
| Male % | -0.0482 | 0.0239 | -0.0951; -0.0014 | ***0.0435*** |
| Hypertension % | -0.0161 | 0.014 | -0.0435; 0.0113 | 0.2488 |
| Diabetes mellitus | -0.0344 | 0.0223 | -0.0781; 0.0093 | 0.1232 |

**Table S23. Meta-regression for the association between mortality outcomes in COVID-19 patients on antiplatelets**

| Variable | Estimate | SE | 95% CI | p-value |
|---|---|---|---|---|
| Publication<br>Pre-print<br>Peer-review | <br>Reference<br>-1.2404 | <br><br>0.4228 | <br><br>-2.0691; -0.4116 | <br><br>***0.0034*** |
| Country<br>China<br>Other | <br>Reference<br>1.4171 | <br><br>1.0413 | <br><br>-0.6237; 3.458 | <br><br>0.1735 |
| Country<br>Italy<br>Other | <br>Reference<br>0.2754 | <br><br>0.5955 | <br><br>-0.8918; 1.4426 | <br><br>0.6438 |
| Number of centres<br>Multiple<br>Single | <br>Reference<br>-0.3326 | <br><br>0.5650 | <br><br>-1.4399; 0.7748 | <br><br>0.5561 |
| Design<br>Case-control<br>Cohort | <br>Reference<br>2.4768 | <br><br>1.6707 | <br><br>-0.7976; 5.7512 | <br><br>0.1382 |
| Setting<br>In-patient and other<br>Only in-patient | <br>Reference<br>-0.1224 | <br><br>0.5312 | <br><br>-1.1636; 0.9187 | <br><br>0.8177 |
| Sample size | 0.0002 | 0.0000 | 0.0001; 0.0003 | ***0.0001*** |
| Age | -0.0681 | 0.0184 | -0.1043; -0.0320 | ***0.0002*** |
| Male % | 0.0002 | 0.0201 | -0.0391; 0.0396 | 0.9903 |
| Hypertension % | -0.0202 | 0.0097 | -0.0392; -0.0013 | ***0.0365*** |
| Diabetes mellitus | -0.0125 | 0.0205 | -0.0526; 0.0277 | 0.5421 |

**Table S24. Meta-regression for the association between mortality outcomes in COVID-19 patients on beta-blockers**

| Variable | Estimate | SE | 95% CI | p-value |
|---|---|---|---|---|
| Publication<br>Pre-print<br>Peer-review | <br>Reference<br>-0.3126 | <br><br>0.3981 | <br><br>-1.093; 0.4677 | <br><br>0.4323 |
| Country<br>France<br>Other | <br>Reference<br>0.1907 | <br><br>0.5086 | <br><br>-0.8062; 1.1875 | <br><br>0.7077 |
| Country<br>Italy<br>Other | <br>Reference<br>-0.1402 | <br><br>0.4691 | <br><br>-1.0595; 0.7792 | <br><br>0.7651 |
| Country<br>USA<br>Other | <br>Reference<br>0.9966 | <br><br>0.6146 | <br><br>-0.208; 2.2012 | <br><br>0.1049 |
| Number of centres<br>Multiple<br>Single | <br>Reference<br>-0.5298 | <br><br>0.3783 | <br><br>-1.2713; 0.2118 | <br><br>0.1614 |
| Setting<br>In-patient and other<br>Only in-patient | <br>Reference<br>-0.6419 | <br><br>0.3641 | <br><br>-1.3556; 0.0717 | <br><br>0.0779 |
| Sample size | 0.0001 | 0.0000 | 0.0001; 0.0002 | ***<.0001*** |
| Age | -0.0443 | 0.0207 | -0.0849; -0.0037 | ***0.0325*** |
| Male % | -0.0044 | 0.0151 | -0.0339; 0.0252 | 0.7715 |
| Hypertension % | -0.0180 | 0.0060 | -0.0297; -0.0062 | ***0.0028*** |
| Diabetes mellitus | -0.0095 | 0.0072 | -0.0236; 0.0046 | 0.1862 |

**Table S25. Meta-regression for the association between mortality outcomes in COVID-19 patients on calcium channel blockers**

| Variable | Estimate | SE | 95% CI | p-value |
|---|---|---|---|---|
| Publication<br>Pre-print<br>Peer-review | <br>Reference<br>-0.2755 | <br><br>0.4154 | <br><br>-1.0898; 0.5388 | <br><br>0.5072 |
| Country<br>China<br>Other | <br>Reference<br>0.3707 | <br><br>0.3797 | <br><br>-0.3735; 1.1148 | <br><br>0.3290 |
| Country<br>Italy<br>Other | <br>Reference<br>-0.3082 | <br><br>0.3920 | <br><br>-1.0766; 0.4602 | <br><br>0.4318 |
| Number of centres<br>Multiple<br>Single | <br>Reference<br>-0.4443 | <br><br>0.3000 | <br><br>-1.0323; 0.1437 | <br><br>0.1386 |
| Setting<br>In-patient and other<br>Only in-patient | <br>Reference<br>-0.5260 | <br><br>0.2955 | <br><br>-1.1053; 0.0533 | <br><br>0.0751 |
| Sample size | 0.0001 | 0.0000 | 0.0000; 0.0002 | ***0.0130*** |
| Age | -0.0482 | 0.0207 | -0.0887; -0.0077 | ***0.0198*** |
| Male % | -0.0028 | 0.0168 | -0.0357; 0.0301 | 0.8673 |
| Hypertension % | -0.0114 | 0.0055 | -0.0221; -0.0007 | ***0.0361*** |
| Diabetes mellitus | -0.032 | 0.0104 | -0.0524; -0.0115 | ***0.0022*** |

**Table S26. Meta-regression for the association between mortality outcomes in COVID-19 patients on diuretics**

| Variable | Estimate | SE | 95% CI | p-value |
|---|---|---|---|---|
| <u>Publication</u><br>Pre-print<br>Peer-review | <br>Reference<br>-1.0024 | <br><br>0.4672 | <br><br>-1.9182; -0.0866 | <br><br>***0.0319*** |
| <u>Country</u><br>France<br>Other | <br>Reference<br>1.0617 | <br><br>0.4869 | <br><br>0.1074; 2.016 | <br><br>***0.0292*** |
| <u>Country</u><br>Italy<br>Other | <br>Reference<br>-0.1579 | <br><br>0.4849 | <br><br>-1.1082; 0.7924 | <br><br>0.7446 |
| <u>Number of centres</u><br>Multiple<br>Single | <br>Reference<br>-0.6744 | <br><br>0.4266 | <br><br>-1.5104; 0.1617 | <br><br>0.1139 |
| <u>Setting</u><br>In-patient and other<br>Only in-patient | <br>Reference<br>-0.8976 | <br><br>0.3571 | <br><br>-1.5975; -0.1978 | <br><br>***0.0119*** |
| Sample size | 0.0000 | 0.0000 | 0.0000; 0.0001 | 0.1918 |
| Age | -0.0782 | 0.0212 | -0.1197; -0.0366 | ***0.0002*** |
| Male % | -0.0285 | 0.0155 | -0.0589; 0.0019 | 0.0657 |
| Hypertension % | -0.0198 | 0.009 | -0.0374; -0.0022 | ***0.0274*** |
| Diabetes mellitus | -0.0088 | 0.007 | -0.0225; 0.005 | 0.2119 |

**Table S27. Meta-regression for the association between mortality outcomes in COVID-19 patients on lipid modifying drugs**

| Variable | Estimate | SE | 95% CI | p-value |
|---|---|---|---|---|
| <u>Publication</u><br>Pre-print<br>Peer-review | <br>Reference<br>-0.5946 | <br><br>0.3574 | <br><br>-1.295; 0.1059 | <br><br>0.0962 |
| <u>Country</u><br>Italy<br>Other | <br>Reference<br>-0.3565 | <br><br>0.4022 | <br><br>-1.1448; 0.4317 | <br><br>0.3754 |
| <u>Number of centres</u><br>Multiple<br>Single | <br>Reference<br>-0.1152 | <br><br>0.3990 | <br><br>-0.8972; 0.6668 | <br><br>0.7728 |
| <u>Setting</u><br>In-patient and other<br>Only in-patient | <br>Reference<br>-0.5139 | <br><br>0.4081 | <br><br>-1.3138; 0.2860 | <br><br>0.2080 |
| Sample size | 0.0000 | 0.0000 | -0.0001; 0.0001 | 0.8888 |
| Age | -0.0217 | 0.0252 | -0.0711; 0.0277 | 0.3887 |
| Male % | -0.0109 | 0.0147 | -0.0397; 0.0178 | 0.4569 |
| Hypertension % | -0.0016 | 0.0091 | -0.0194; 0.0161 | 0.8559 |
| Diabetes mellitus | -0.0048 | 0.007 | -0.0186; 0.009 | 0.4943 |

### Supplementary Figures

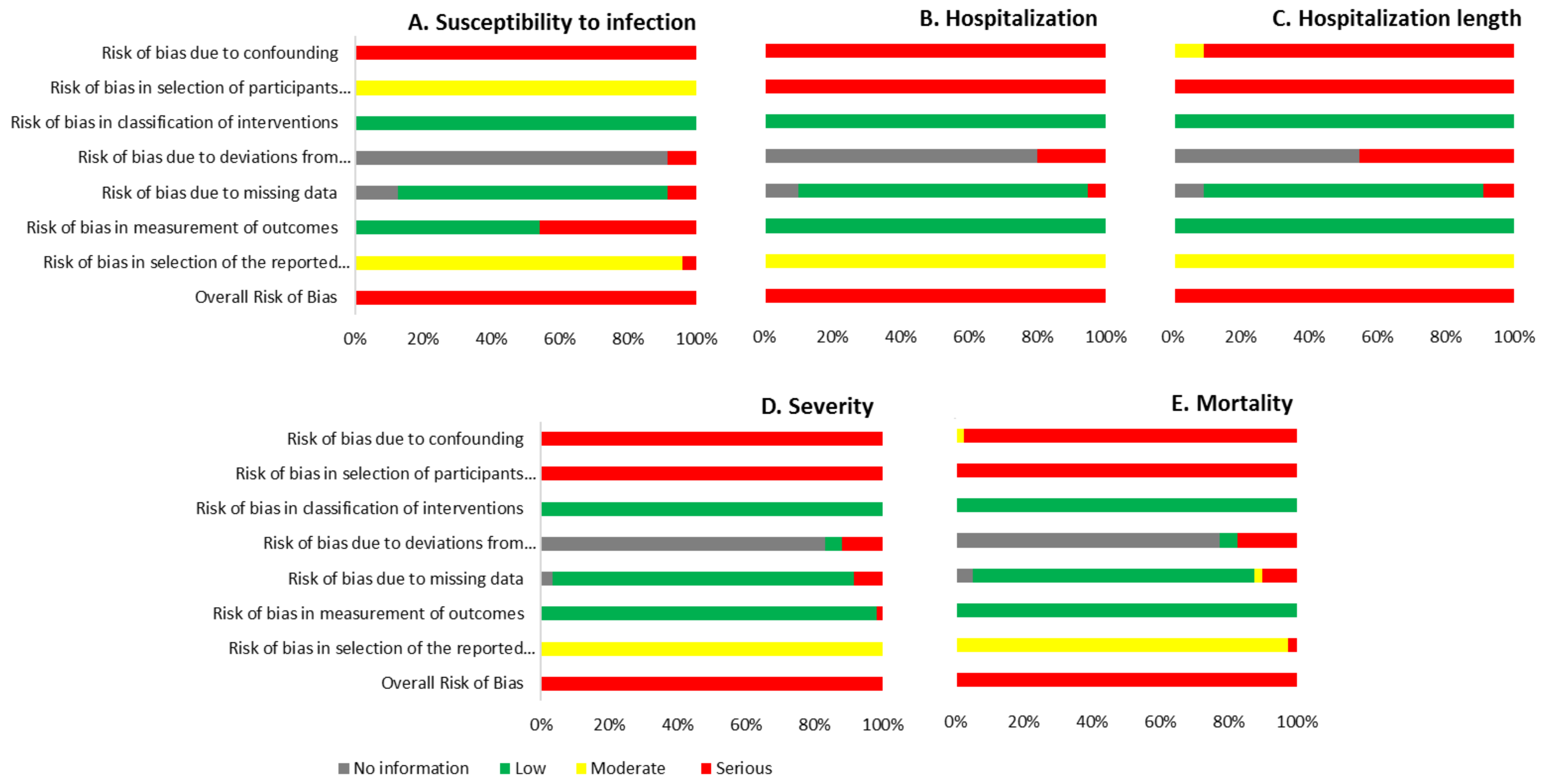


**Figure S1. Risk of bias summaries for the associations between COVID-19 outcomes and being on an angiotensin-converting enzyme inhibitor or angiotensin receptor blocker.**

**A. Studies from the same cluster included**

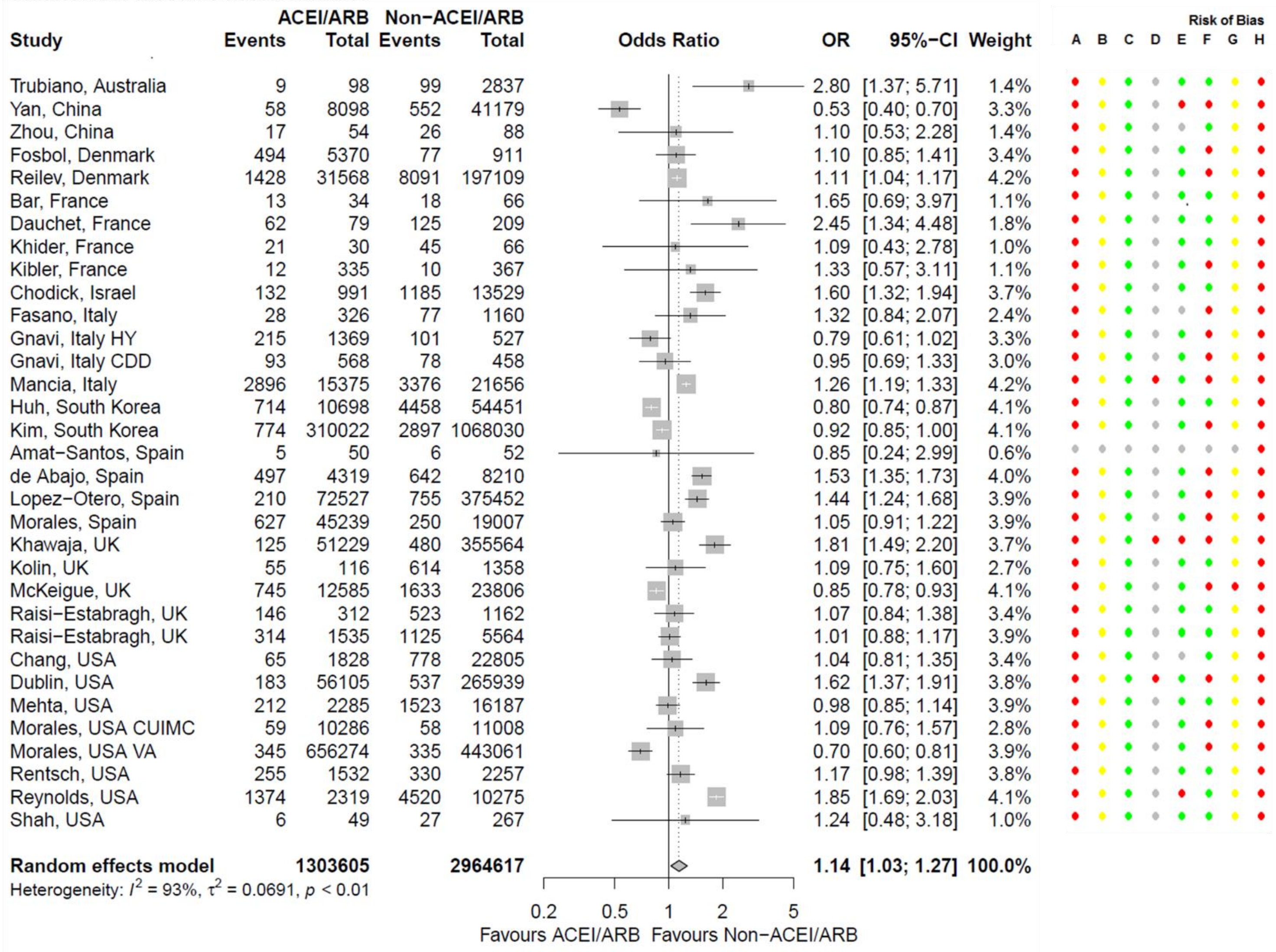


**Figure S2. Forest plot for association between testing positive for COVID-19 and being on an angiotensin-converting enzyme inhibitor (ACEI) or angiotensin receptor blocker (ARB).** Bar,[74] Chang,[158] Chodick,[86] Fasano,[93] Huh,[121] Kim,[123] Mancia,[102] McKeigue[152] and Raisi-Estabragh[149] estimates assume that none of the patients are taking both ACEIs and ARBs. The Amat-Santos[126] study was assessed using the revised Cochrane risk-of-bias tool for randomized trials[185] with a final rating of 'some concerns'. Gnavi[98] and Morales[175] studies each provided two separate cohorts. Risk of bias legend. A = risk of bias due to confounding, B = risk of bias in selection of participants into the study, C = risk of bias in classification of interventions, D = risk of bias due to deviations from intended interventions, E = risk of bias due to missing data, F = risk of bias in measurement of outcomes, G = risk of bias in selection of the reported result, H = overall risk of bias. Color codes. Colour codes. Red = serious, yellow = moderate, green = low, grey = unclear. Abbreviations. CDD = circulatory diseases/diabetes population, CUIMC = Columbia University Irving Medical Center, HY = hypertension population, VA = Department of Veterans Affairs database.

**B. China-only studies**

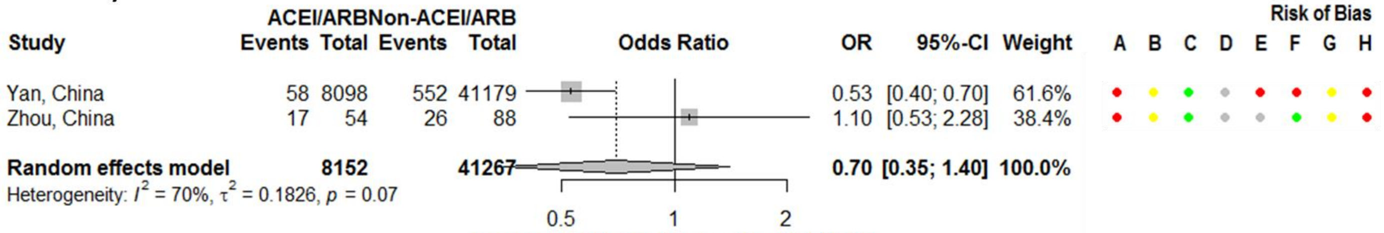


**C. Non-Chinese studies**

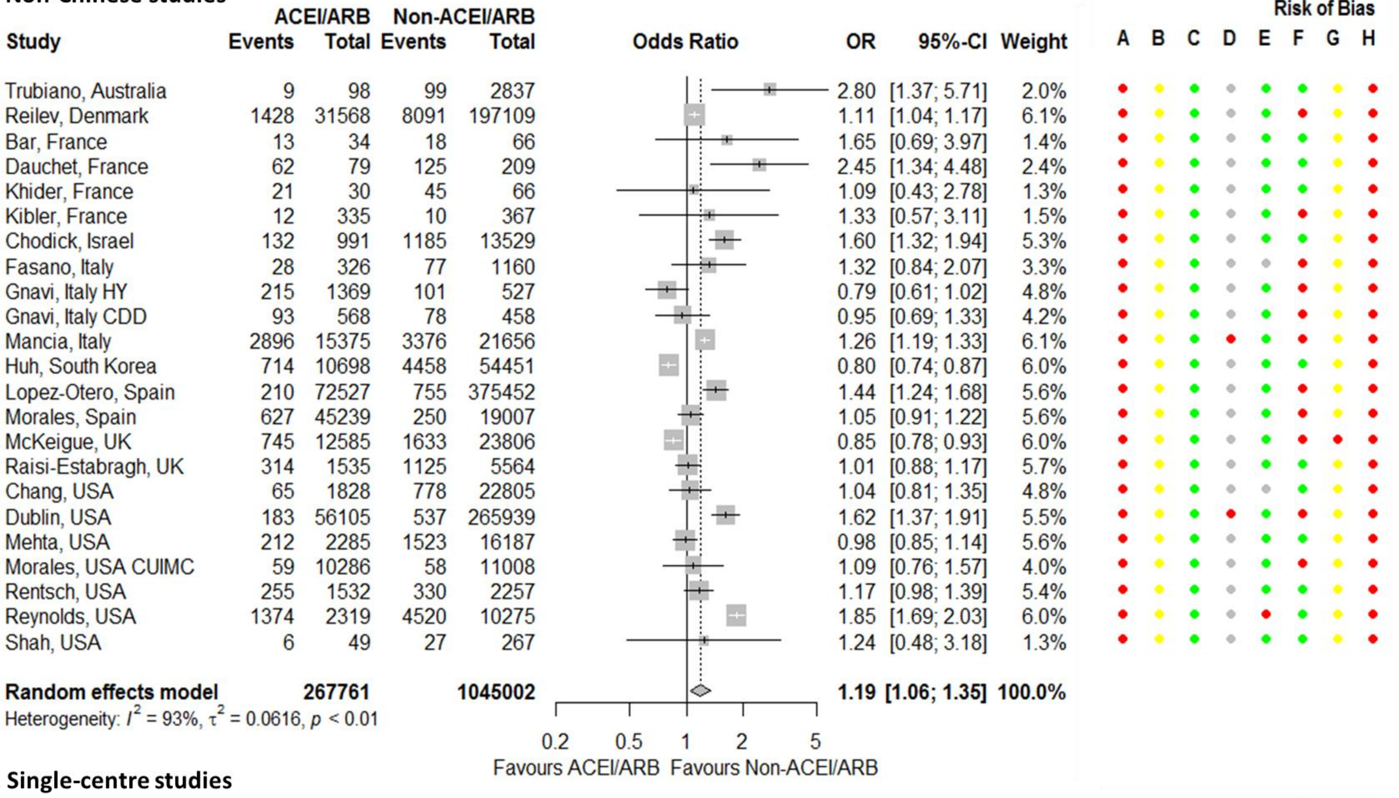


**D. Single-centre studies**

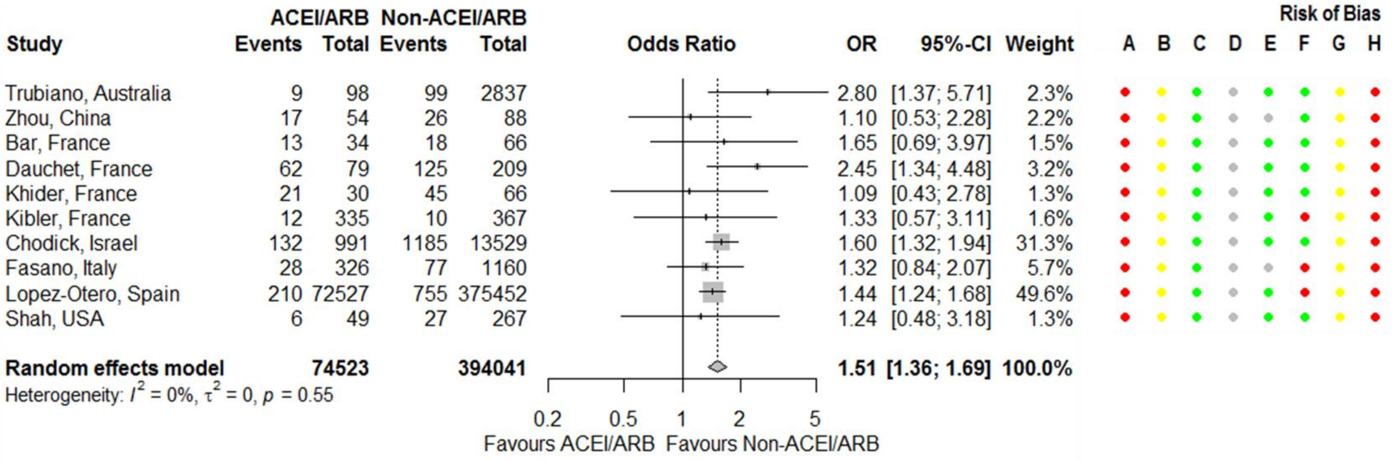


**Figure S2 Continued.**

**E. Multicentre studies**

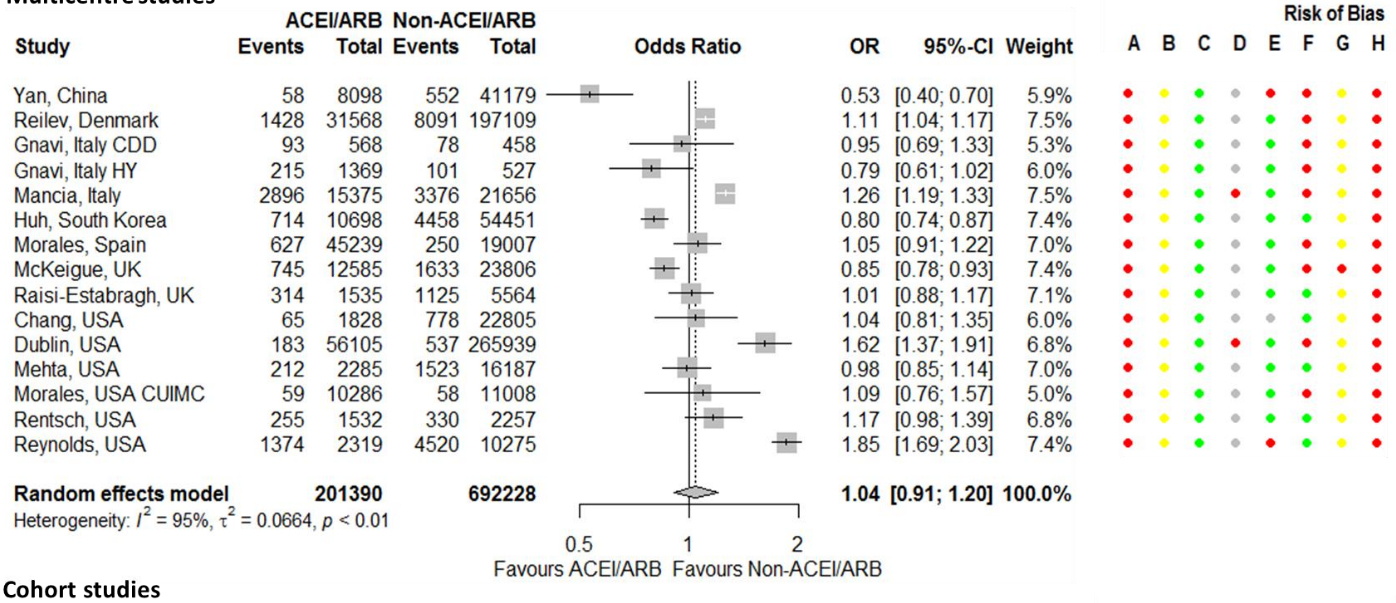


**F. Cohort studies**

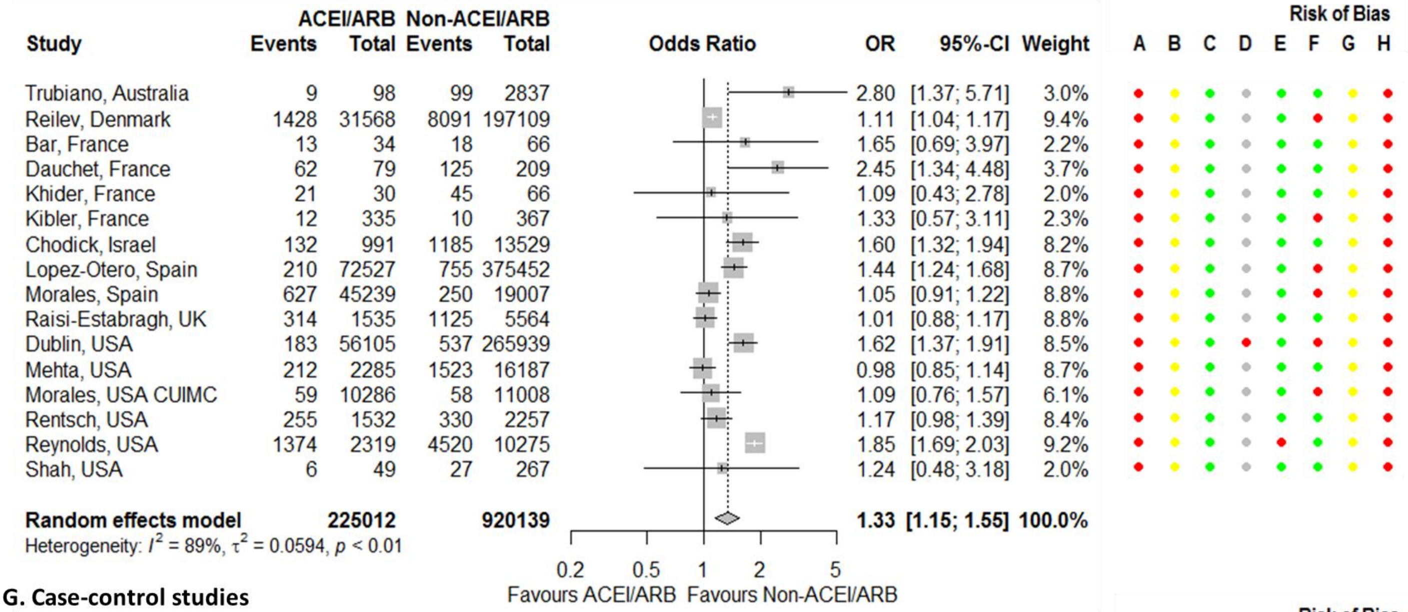


**G. Case-control studies**

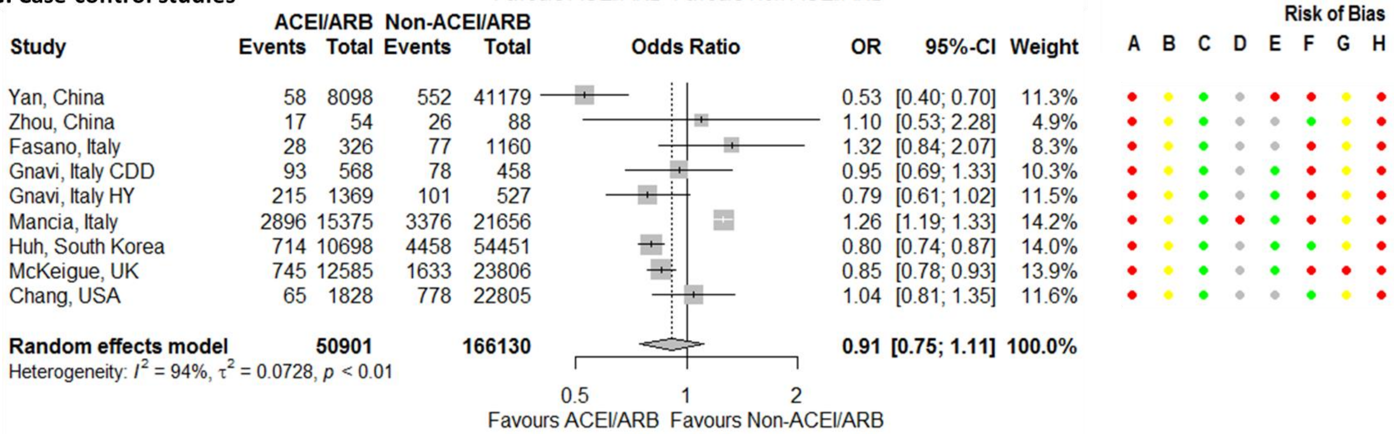


**Figure S2 Continued.**

### H. Adjusted estimates

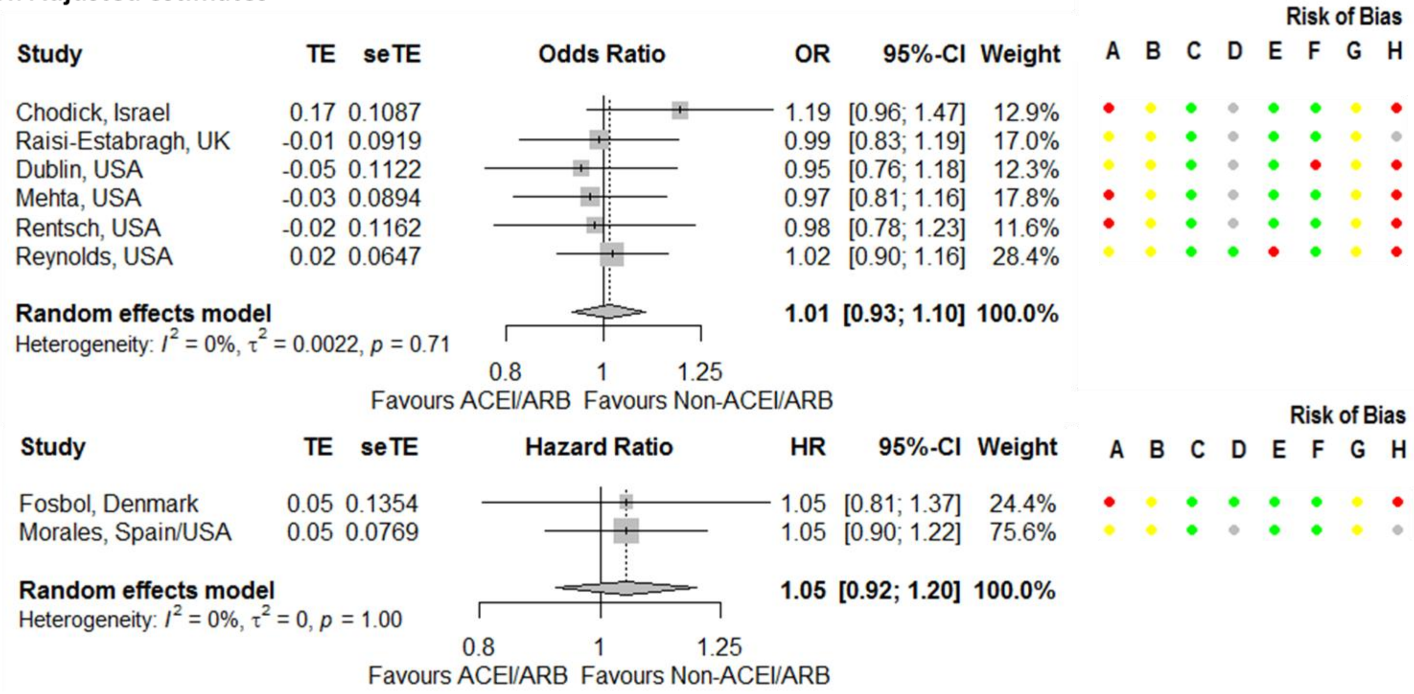


**Figure S2 Continued.**

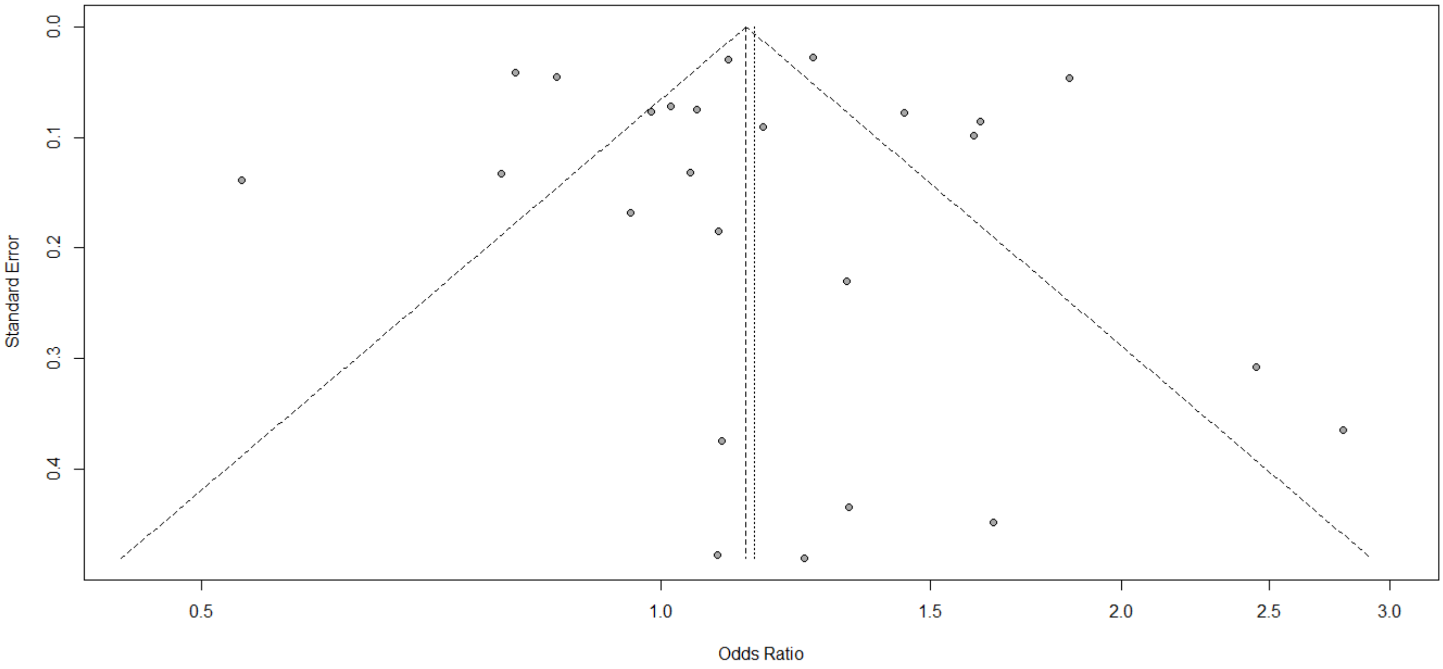


**Figure S3. Funnel plot for association between testing positive for COVID-19 and being on an angiotensin-converting enzyme inhibitor or angiotensin receptor blocker.**

**A. Studies from the same cluster included**

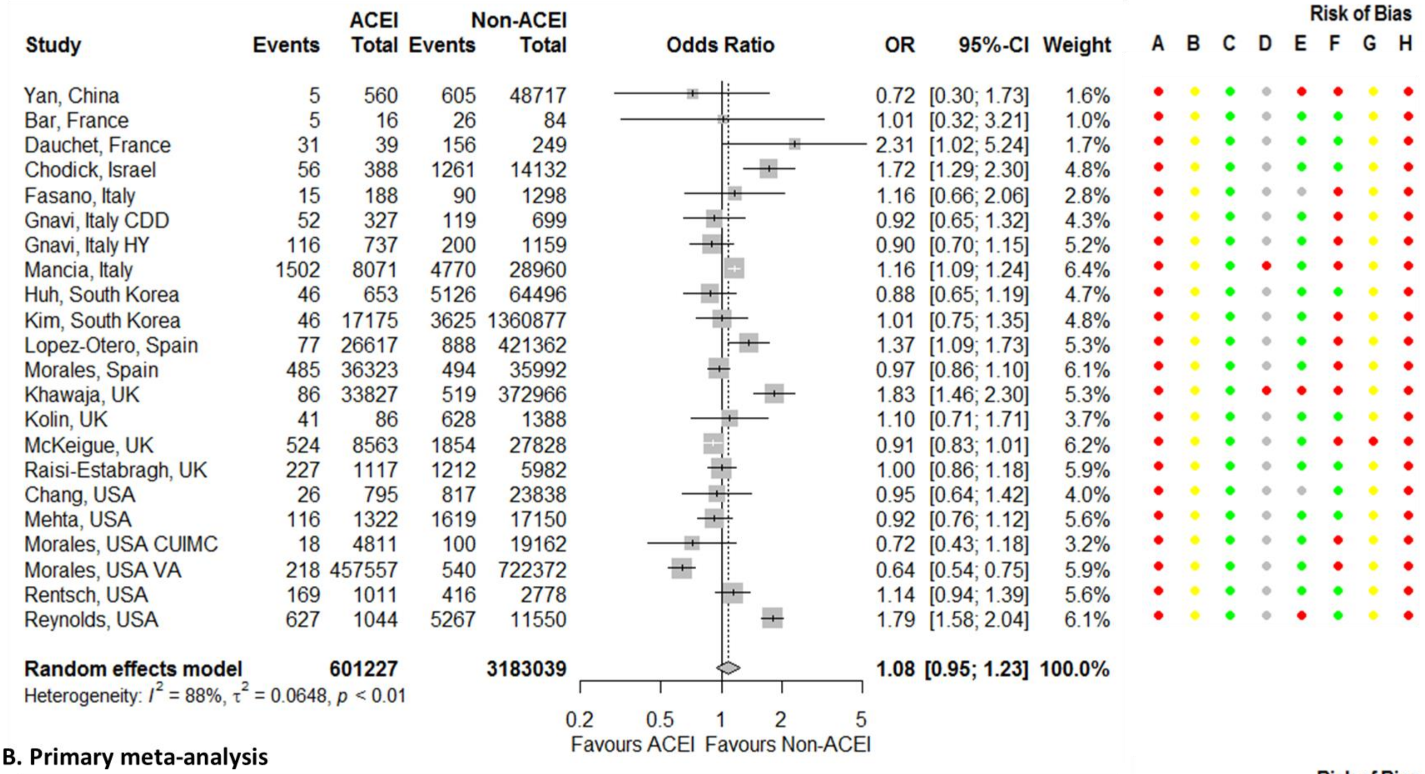


**B. Primary meta-analysis**

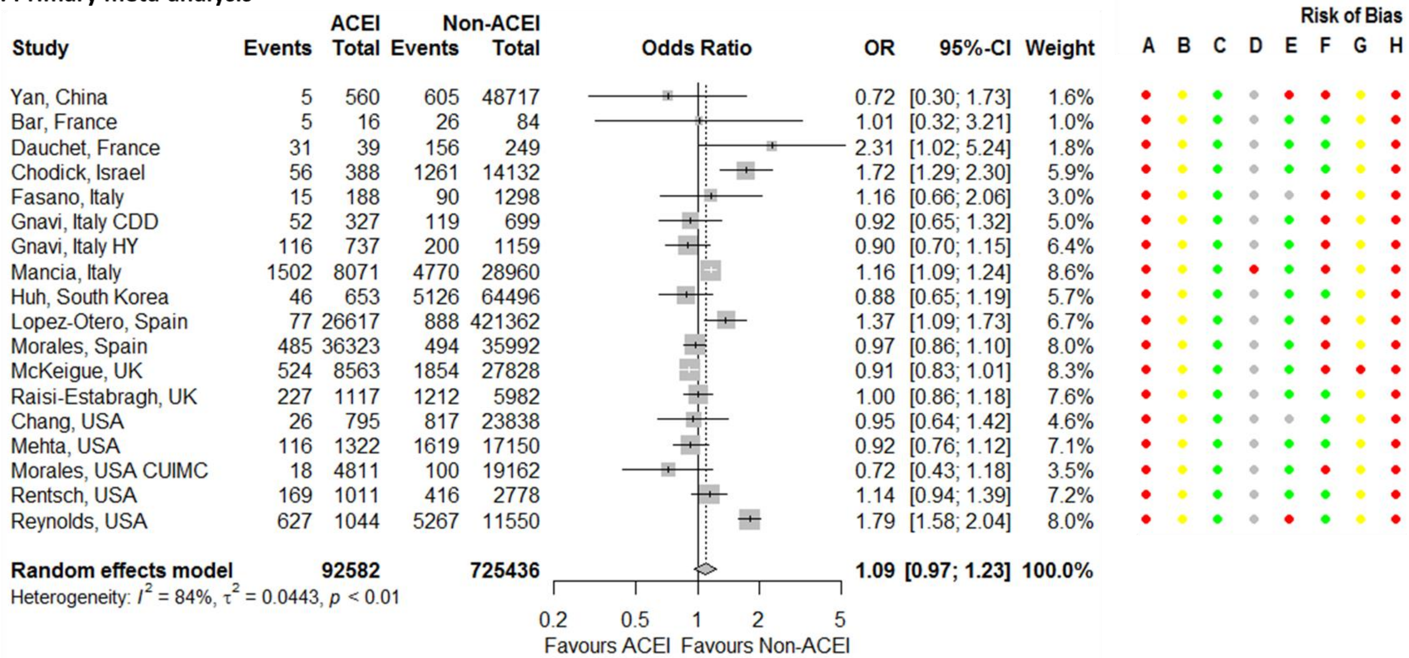


**Figure S4. Forest plot for association between testing positive for COVID-19 and being on an angiotensin-converting enzyme inhibitor (ACEI).** Risk of bias legend. A = risk of bias due to confounding, B = risk of bias in selection of participants into the study, C = risk of bias in classification of interventions, D = risk of bias due to deviations from intended interventions, E = risk of bias due to missing data, F = risk of bias in measurement of outcomes, G = risk of bias in selection of the reported result, H = overall risk of bias. Color codes. Colour codes. Red = serious, yellow = moderate, green = low, grey = unclear. Abbreviations. CDD = circulatory diseases/diabetes population, CUIMC = Columbia University Irving Medical Center, HY = hypertension population, VA = Department of Veterans Affairs database.

**C. Single-centre studies**

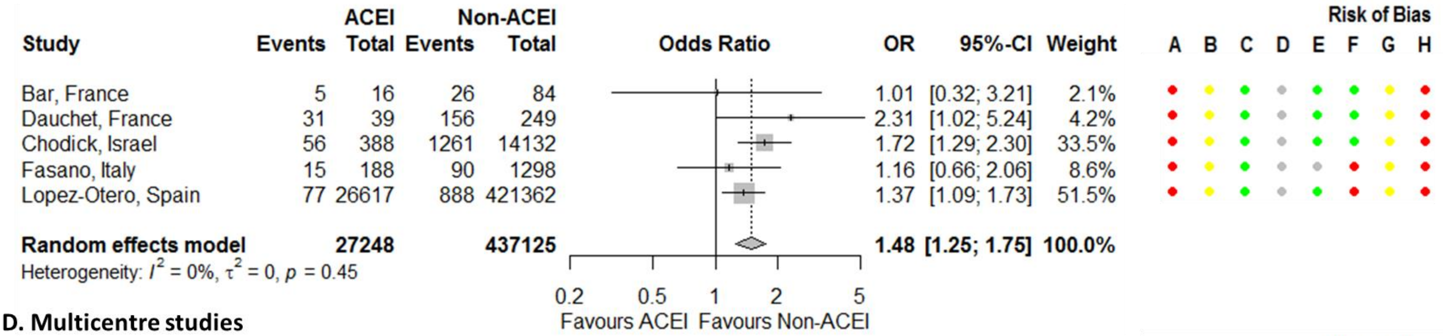


**D. Multicentre studies**

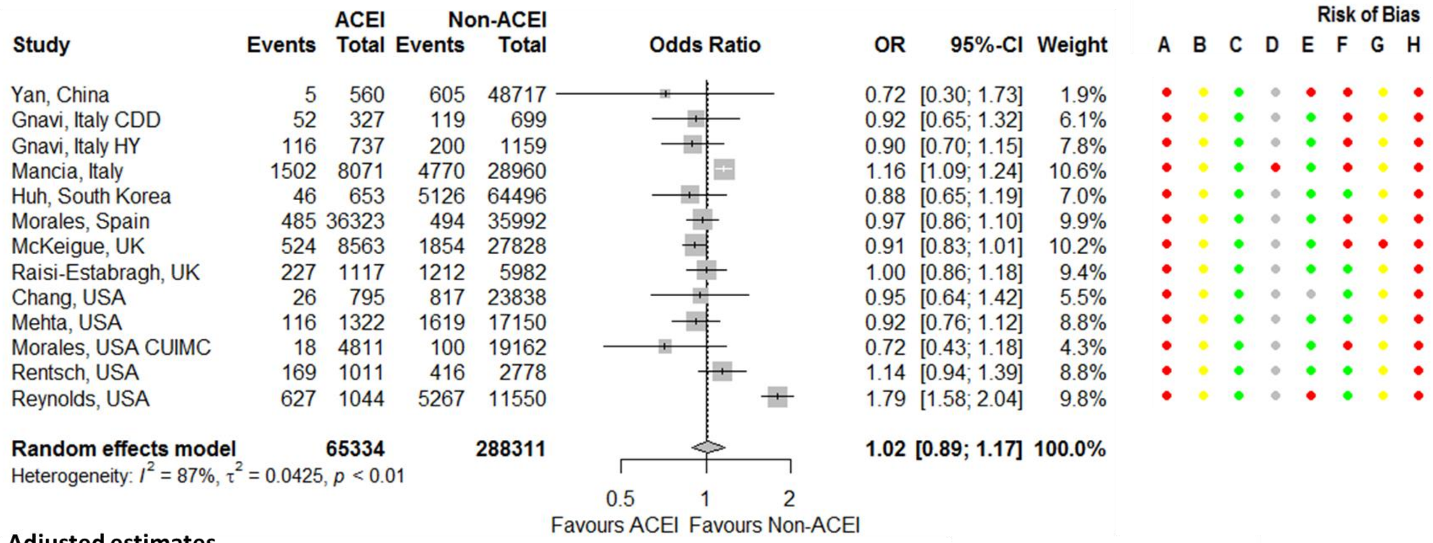


**E. Adjusted estimates**

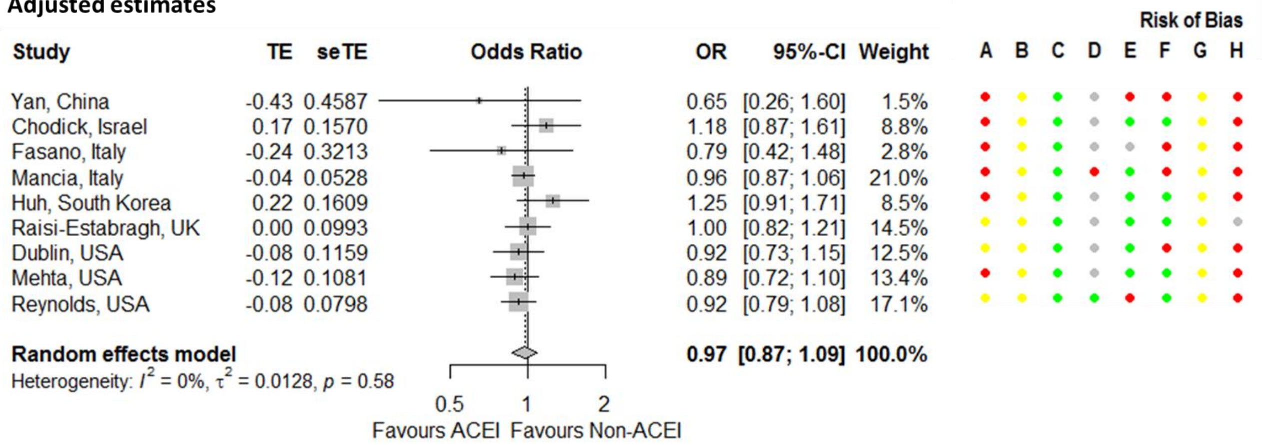


**Figure S4 continued**.

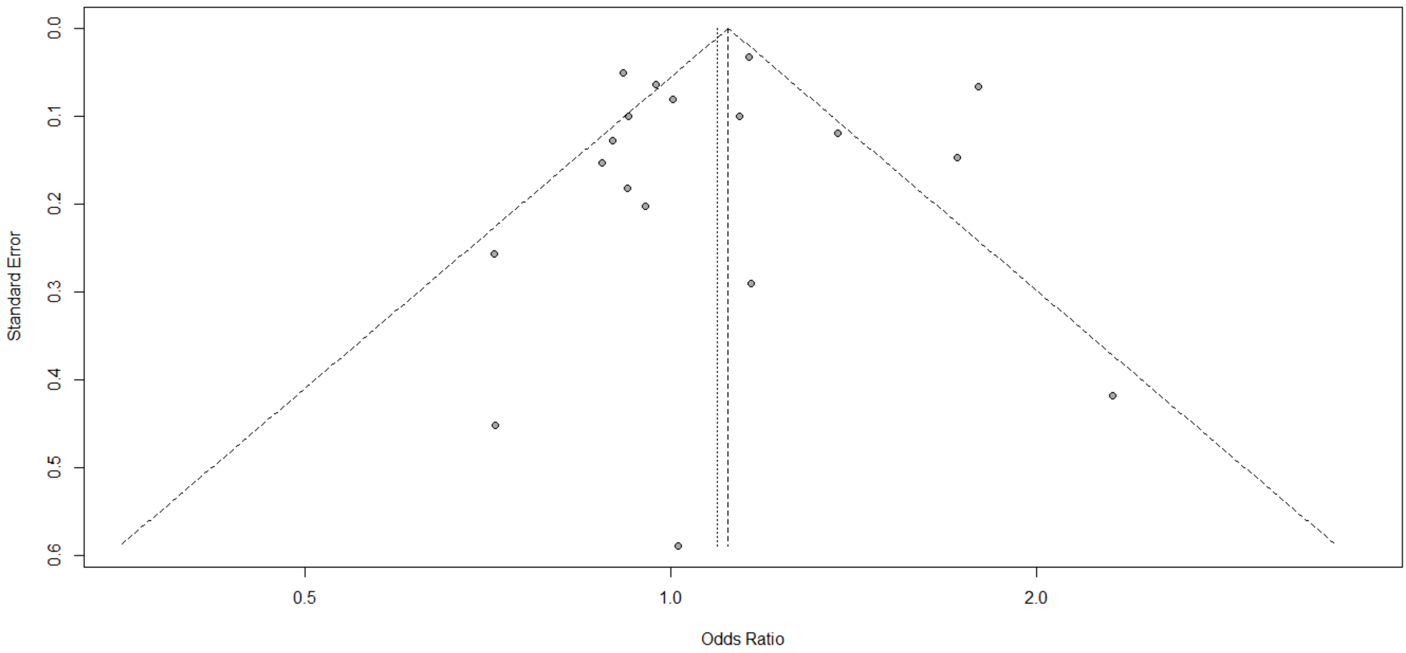


**Figure S5. Funnel plot for association between testing positive for COVID-19 and being on an angiotensin-converting enzyme inhibitor.**

**A. Studies from the same cluster included**

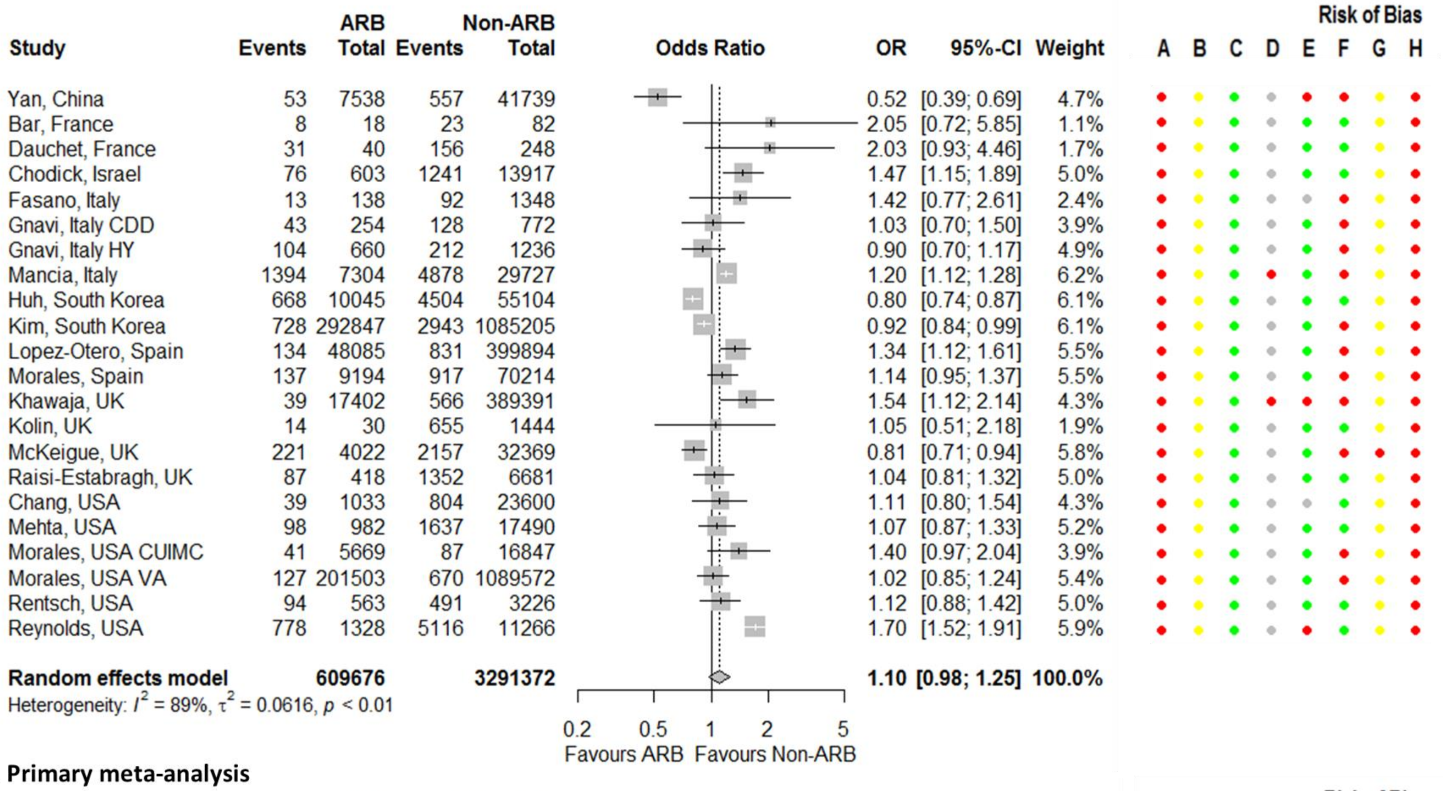


**B. Primary meta-analysis**

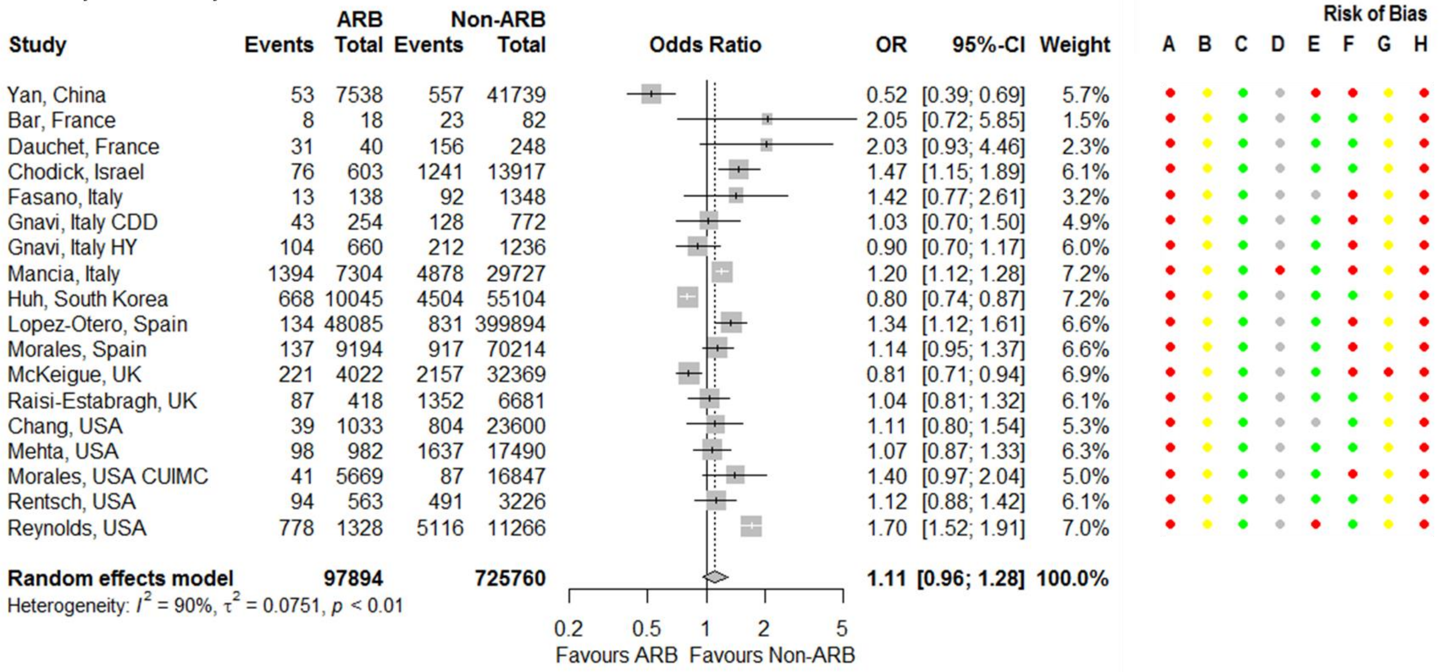


**Figure S6. Forest plot for association between testing positive for COVID-19 and being on an angiotensin receptor blocker (ARB) with studies from the same cluster all included.** Risk of bias legend. A = risk of bias due to confounding, B = risk of bias in selection of participants into the study, C = risk of bias in classification of interventions, D = risk of bias due to deviations from intended interventions, E = risk of bias due to missing data, F = risk of bias in measurement of outcomes, G = risk of bias in selection of the reported result, H = overall risk of bias. Color codes. Colour codes. Red = serious, yellow = moderate, green = low, grey = unclear. Abbreviations. CDD = circulatory diseases/diabetes population, CUIMC = Columbia University Irving Medical Center, HY = hypertension population, VA = Department of Veterans Affairs database.

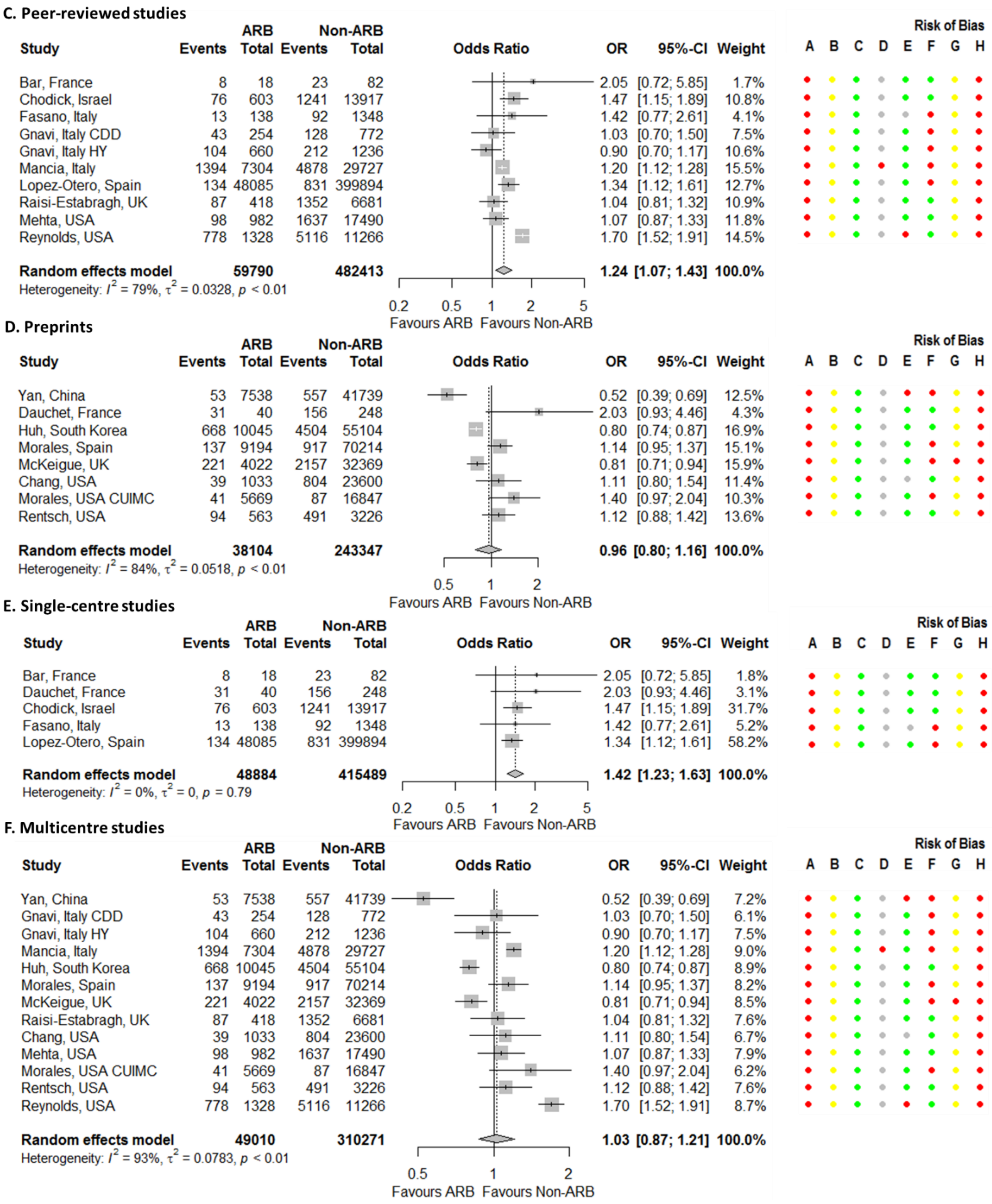


**Figure S6 Continued.**

**G. Cohort studies**

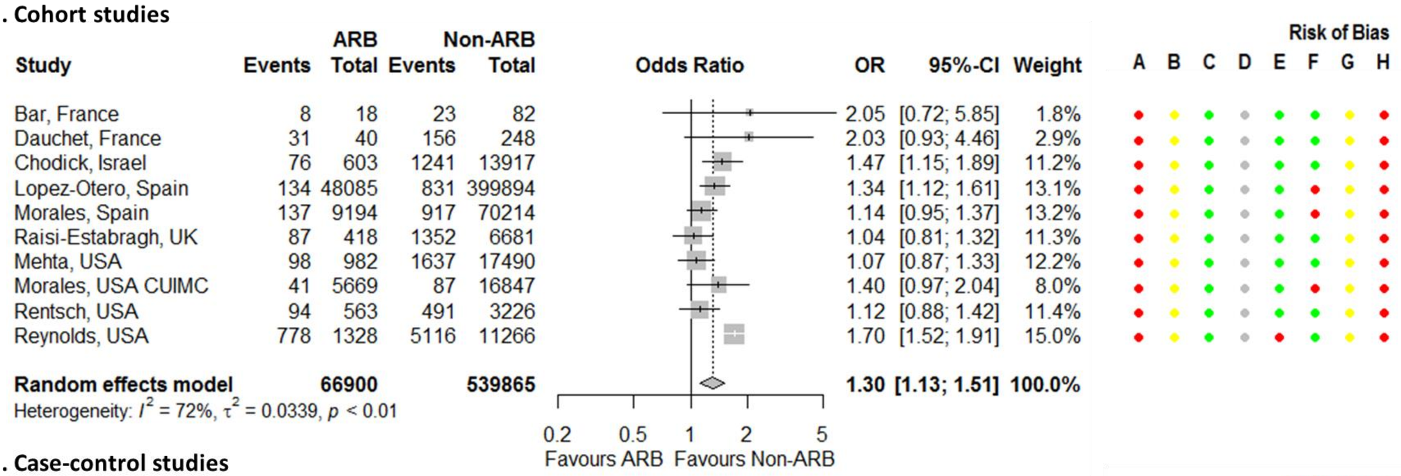


**H. Case-control studies**

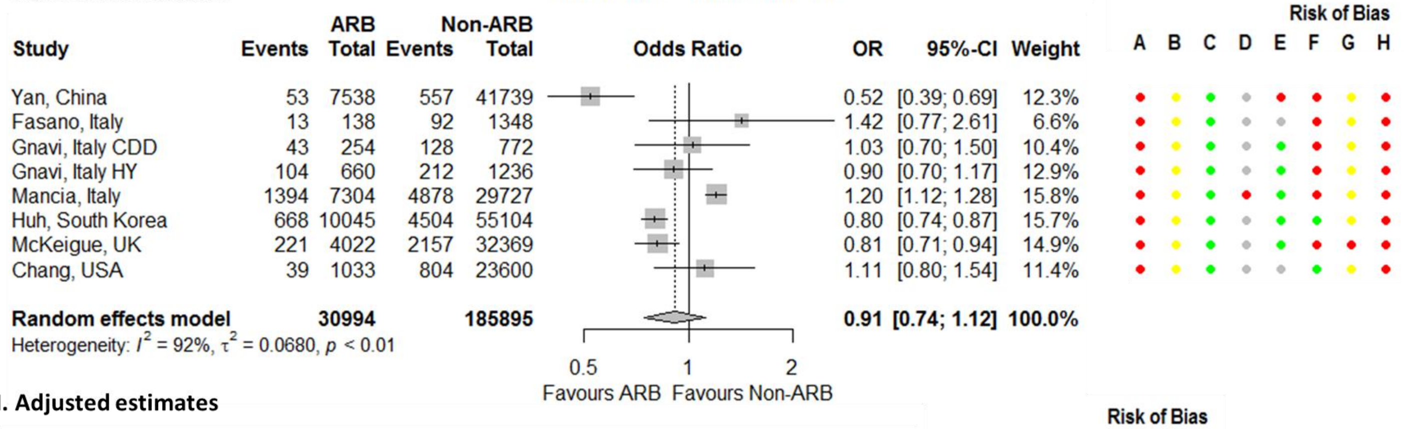


**I. Adjusted estimates**

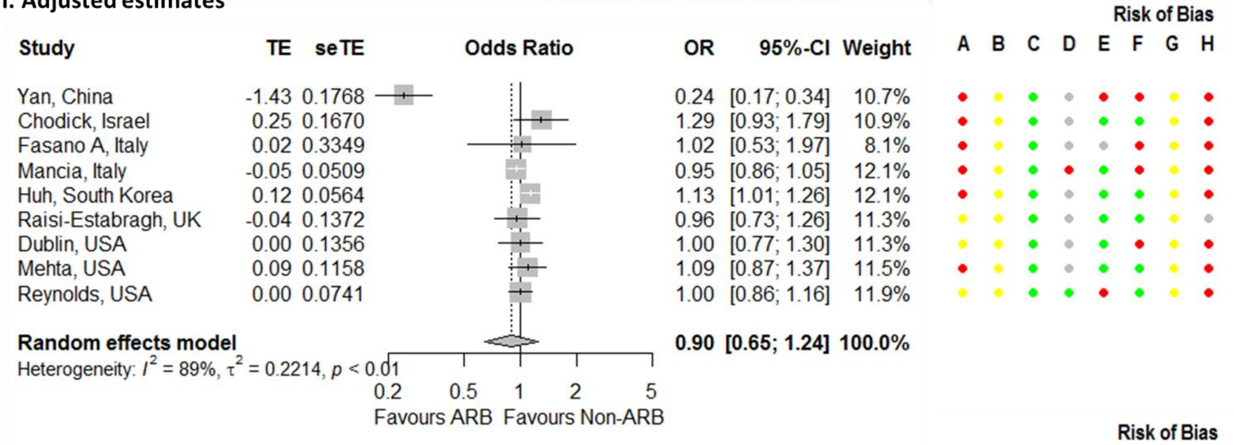


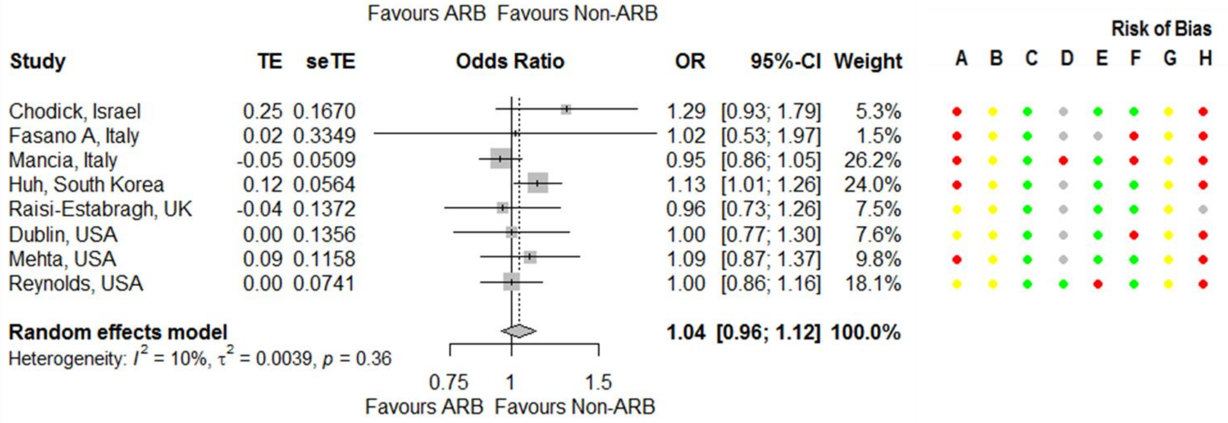


**Figure S6 Continued.**

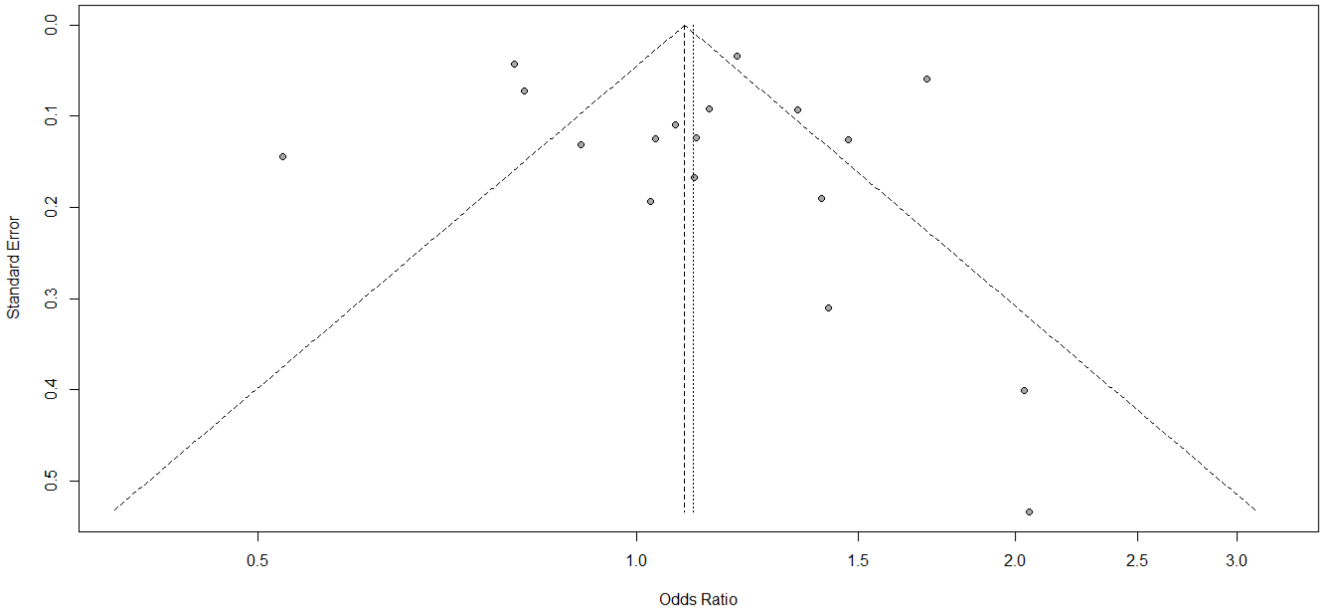


**Figure S7. Funnel plot for association between testing positive for COVID-19 and being on an angiotensin receptor blocker.**

**A. Anticoagulants**

| Study | Anticoag Events | Anticoag Total | Non-Anticoag Events | Non-Anticoag Total | OR | 95%-CI | Weight |
|---|---|---|---|---|---|---|---|
| Reilev, Denmark | 517 | 10644 | 9002 | 218033 | 1.19 | [1.08; 1.30] | 17.8% |
| Kibler, France | 10 | 319 | 12 | 383 | 1.00 | [0.43; 2.35] | 3.8% |
| Rieder, Germany | 6 | 32 | 43 | 158 | 0.62 | [0.24; 1.60] | 3.1% |
| Mancia, Italy | 643 | 2816 | 5629 | 34215 | 1.50 | [1.37; 1.65] | 17.7% |
| Amat-Santos, Spain | 6 | 34 | 5 | 68 | 2.70 | [0.76; 9.59] | 1.9% |
| Fletcher, UK | 206 | 402 | 446 | 877 | 1.02 | [0.80; 1.29] | 14.3% |
| McKeigue, UK | 410 | 4283 | 1968 | 32108 | 1.62 | [1.45; 1.81] | 17.4% |
| Chang, USA | 19 | 561 | 824 | 24072 | 0.99 | [0.62; 1.57] | 8.6% |
| Gu, USA | 148 | 1025 | 578 | 3387 | 0.82 | [0.67; 1.00] | 15.4% |
| **Random effects model** | | **20116** | | **313301** | **1.17** | **[0.98; 1.41]** | **100.0%** |

Heterogeneity: $I^2 = 86\%$, $\tau^2 = 0.0485$, $p < 0.01$

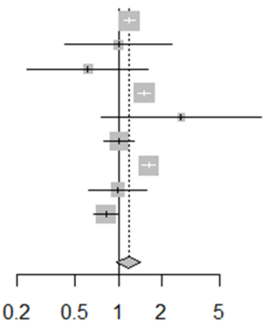


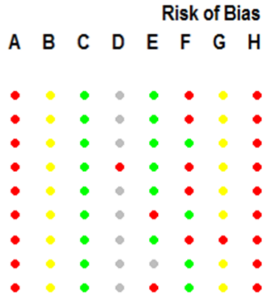


**B. Antiplatelets**

| Study | Antiplatelets Events | Antiplatelets Total | Non-Antiplatelets Events | Non-Antiplatelets Total | OR | 95%-CI | Weight |
|---|---|---|---|---|---|---|---|
| Yan, China | 11 | 649 | 599 | 48628 | 1.38 | [0.76; 2.52] | 9.5% |
| Reilev, Denmark | 777 | 17673 | 8742 | 211004 | 1.06 | [0.99; 1.15] | 25.8% |
| Kibler, France | 13 | 365 | 9 | 337 | 1.35 | [0.57; 3.19] | 5.7% |
| Mancia, Italy | 1363 | 6231 | 4909 | 30800 | 1.48 | [1.38; 1.58] | 25.9% |
| Huh, South Korea | 8 | 543 | 5164 | 64606 | 0.17 | [0.09; 0.35] | 7.8% |
| McKeigue, UK | 691 | 9523 | 1687 | 26868 | 1.17 | [1.07; 1.28] | 25.4% |
| **Random effects model** | | **34984** | | **382243** | **1.07** | **[0.85; 1.35]** | **100.0%** |

Heterogeneity: $I^2 = 93\%$, $\tau^2 = 0.0527$, $p < 0.01$

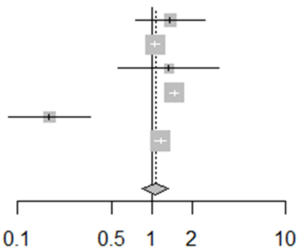


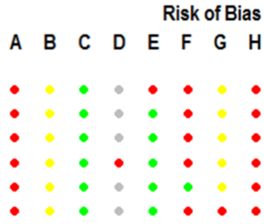


**C. Beta-blockers**

| Study | ß-blockers Events | ß-blockers Total | Non-ß-blockers Events | Non-ß-blockers Total | OR | 95%-CI | Weight |
|---|---|---|---|---|---|---|---|
| Yan, China | 14 | 1236 | 596 | 48041 | 0.91 | [0.54; 1.55] | 3.8% |
| Reilev, Denmark | 774 | 17929 | 8745 | 210748 | 1.04 | [0.97; 1.12] | 16.5% |
| Khider, France | 8 | 13 | 58 | 83 | 0.69 | [0.21; 2.32] | 0.9% |
| Mancia, Italy | 1826 | 8949 | 4446 | 28082 | 1.36 | [1.28; 1.45] | 16.9% |
| Kim, South Korea | 225 | 80898 | 3446 | 1297154 | 1.05 | [0.91; 1.20] | 14.3% |
| Fletcher, UK | 129 | 267 | 523 | 1012 | 0.87 | [0.67; 1.14] | 9.1% |
| Kolin, UK | 65 | 144 | 604 | 1330 | 0.99 | [0.70; 1.40] | 6.9% |
| McKeigue, UK | 688 | 8979 | 1690 | 27412 | 1.26 | [1.15; 1.38] | 16.0% |
| Reynolds, USA | 912 | 1686 | 4982 | 10908 | 1.40 | [1.26; 1.55] | 15.6% |
| **Random effects model** | | **120101** | | **1624770** | **1.15** | **[1.02; 1.29]** | **100.0%** |

Heterogeneity: $I^2 = 84\%$, $\tau^2 = 0.0200$, $p < 0.01$

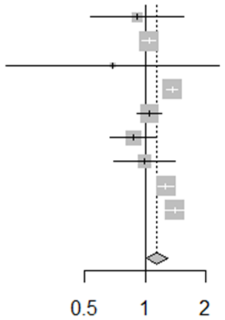


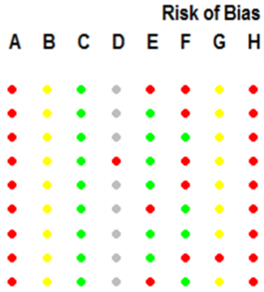


**D. Calcium channel blockers (CCBs)**

| Study | CCBs Events | CCBs Total | Non-CCBs Events | Non-CCBs Total | OR | 95%-CI | Weight |
|---|---|---|---|---|---|---|---|
| Yan, China | 89 | 4829 | 521 | 44448 | 1.58 | [1.26; 1.99] | 11.3% |
| Reilev, Denmark | 743 | 17661 | 8776 | 211016 | 1.01 | [0.94; 1.09] | 13.3% |
| Khider, France | 13 | 20 | 53 | 76 | 0.81 | [0.28; 2.28] | 2.5% |
| Mancia, Italy | 1446 | 7372 | 4826 | 29659 | 1.26 | [1.18; 1.34] | 13.4% |
| Kim, South Korea | 659 | 274252 | 3012 | 1103800 | 0.88 | [0.81; 0.96] | 13.3% |
| Fletcher, UK | 152 | 304 | 500 | 975 | 0.95 | [0.73; 1.23] | 10.7% |
| Kolin, UK | 65 | 145 | 604 | 1329 | 0.98 | [0.69; 1.38] | 9.2% |
| McKeigue, UK | 526 | 8804 | 1852 | 27587 | 0.88 | [0.80; 0.98] | 13.1% |
| Reynolds, USA | 992 | 1672 | 4902 | 10922 | 1.79 | [1.61; 1.99] | 13.1% |
| **Random effects model** | | **315059** | | **1429812** | **1.12** | **[0.93; 1.35]** | **100.0%** |

Heterogeneity: $I^2 = 95\%$, $\tau^2 = 0.0636$, $p < 0.01$

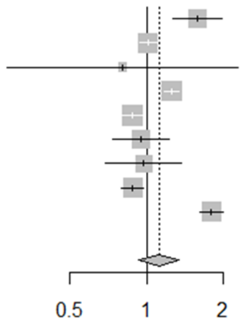


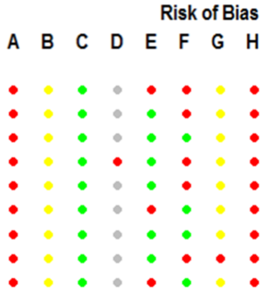


**Figure S8. Forest plots for association between testing positive for COVID-19 and being on cardiovascular drugs.** Risk of bias legend. A = risk of bias due to confounding, B = risk of bias in selection of participants into the study, C = risk of bias in classification of interventions, D = risk of bias due to deviations from intended interventions, E = risk of bias due to missing data, F = risk of bias in measurement of outcomes, G = risk of bias in selection of the reported result, H = overall risk of bias. Color codes. Colour codes. Red = serious, yellow = moderate, green = low, grey = unclear.

**E. Class III antiarrythmics (Amiodarone)**

| Study | Amiodarone Events | Amiodarone Total | Non-Amiodarone Events | Non-Amiodarone Total | OR | 95%-CI | Weight |
|---|---|---|---|---|---|---|---|
| Kibler, France | 2 | 101 | 20 | 601 | 0.59 | [0.14; 2.55] | 26.4% |
| Huh, South Korea | 17 | 714 | 5155 | 64435 | 0.28 | [0.17; 0.45] | 36.8% |
| McKeigue, UK | 19 | 190 | 2359 | 36201 | 1.59 | [0.99; 2.57] | 36.8% |
| **Random effects model** | | **1005** | | **101237** | **0.65** | **[0.17; 2.47]** | **100.0%** |

Heterogeneity: $I^2 = 92\%$, $\tau^2 = 1.2091$, $p < 0.01$

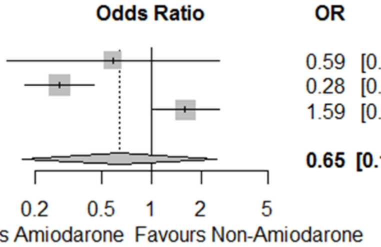


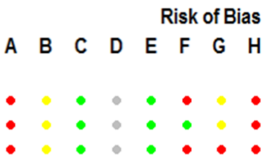


**F. Diuretics**

| Study | Diuretics Events | Diuretics Total | Non-Diuretics Events | Non-Diuretics Total | OR | 95%-CI | Weight |
|---|---|---|---|---|---|---|---|
| Yan, China | 13 | 2480 | 597 | 46797 | 0.41 | [0.24; 0.71] | 5.6% |
| Reilev, Denmark | 538 | 10990 | 8981 | 217687 | 1.20 | [1.09; 1.31] | 13.7% |
| Khider, France | 6 | 9 | 60 | 87 | 0.90 | [0.21; 3.87] | 1.2% |
| Mancia, Italy | 1902 | 9322 | 4370 | 27709 | 1.37 | [1.29; 1.45] | 14.0% |
| Davies, South Africa | 5272 | 569180 | 17036 | 2891752 | 1.58 | [1.53; 1.63] | 14.2% |
| Kim, South Korea | 423 | 161417 | 3248 | 1216635 | 0.98 | [0.89; 1.09] | 13.6% |
| Khawaja, UK | 71 | 27137 | 534 | 379656 | 1.86 | [1.45; 2.39] | 10.9% |
| McKeigue, UK | 560 | 4529 | 1818 | 31862 | 2.33 | [2.11; 2.58] | 13.6% |
| Reynolds, USA | 551 | 989 | 5343 | 11605 | 1.47 | [1.29; 1.68] | 13.1% |
| **Random effects model** | | **786053** | | **4823790** | **1.37** | **[1.16; 1.62]** | **100.0%** |

Heterogeneity: $I^2 = 96\%$, $\tau^2 = 0.0514$, $p < 0.01$

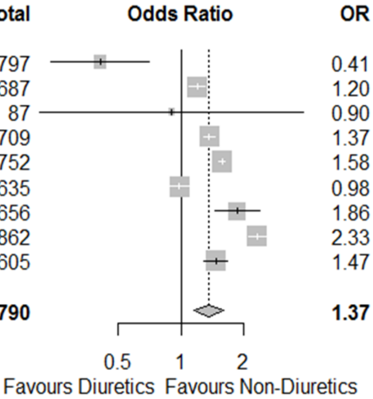


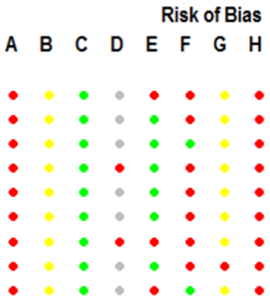


**G. Loop diuretics**

| Study | Loop diuretics Events | Loop diuretics Total | Non-Loop diuretics Events | Non-Loop diuretics Total | OR | 95%-CI | Weight |
|---|---|---|---|---|---|---|---|
| Reilev, Denmark | 538 | 10990 | 8981 | 217687 | 1.20 | [1.09; 1.31] | 27.7% |
| Mancia, Italy | 871 | 3282 | 5401 | 33749 | 1.90 | [1.75; 2.06] | 27.8% |
| Khawaja, UK | 14 | 3145 | 591 | 403648 | 3.05 | [1.79; 5.19] | 17.0% |
| McKeigue, UK | 560 | 4529 | 1818 | 31862 | 2.33 | [2.11; 2.58] | 27.6% |
| **Random effects model** | | **21946** | | **686946** | **1.92** | **[1.35; 2.71]** | **100.0%** |

Heterogeneity: $I^2 = 97\%$, $\tau^2 = 0.1106$, $p < 0.01$

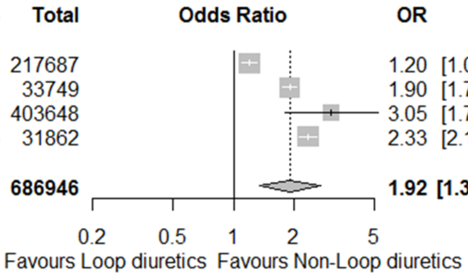


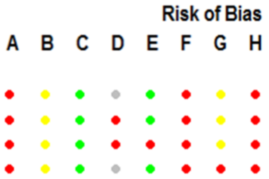


**H. Thiazide and related diuretics**

| Study | Thiazides Events | Thiazides Total | Non-Thiazides Events | Non-Thiazides Total | OR | 95%-CI | Weight |
|---|---|---|---|---|---|---|---|
| Reilev, Denmark | 408 | 9136 | 9111 | 219541 | 1.08 | [0.98; 1.19] | 20.8% |
| Mancia, Italy | 1104 | 6178 | 5168 | 30853 | 1.08 | [1.01; 1.16] | 21.2% |
| Khawaja, UK | 71 | 27137 | 534 | 379656 | 1.86 | [1.45; 2.39] | 17.8% |
| McKeigue P, UK | 177 | 4046 | 2201 | 32345 | 0.63 | [0.54; 0.73] | 19.9% |
| Reynolds, USA | 551 | 989 | 5343 | 11605 | 1.47 | [1.29; 1.68] | 20.3% |
| **Random effects model** | | **47486** | | **674000** | **1.14** | **[0.88; 1.47]** | **100.0%** |

Heterogeneity: $I^2 = 95\%$, $\tau^2 = 0.0772$, $p < 0.01$

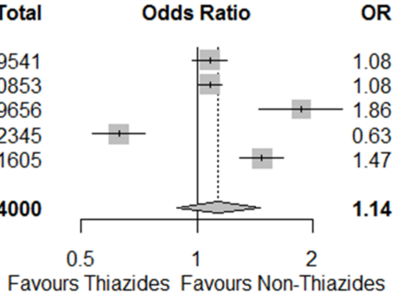


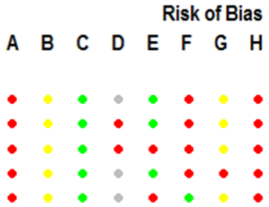


**I. Potassium-sparing diuretics and aldosterone antagonists (PSDAA)**

| Study | PSDAAs Events | PSDAAs Total | Non-PSDAAs Events | Non-PSDAAs Total | OR | 95%-CI | Weight |
|---|---|---|---|---|---|---|---|
| Mancia, Italy | 239 | 977 | 6033 | 36054 | 1.61 | [1.39; 1.87] | 54.4% |
| Khawaja, UK | 5 | 1193 | 600 | 405600 | 2.84 | [1.18; 6.86] | 4.9% |
| McKeigue, UK | 96 | 802 | 2282 | 35589 | 1.98 | [1.60; 2.47] | 40.7% |
| **Random effects model** | | **2972** | | **477243** | **1.80** | **[1.47; 2.21]** | **100.0%** |

Heterogeneity: $I^2 = 45\%$, $\tau^2 = 0.0137$, $p = 0.16$

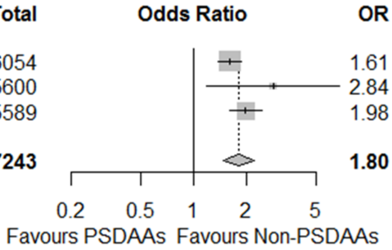


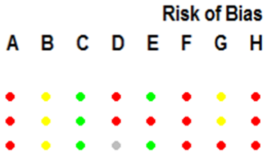


**Figure S8. Continued.**

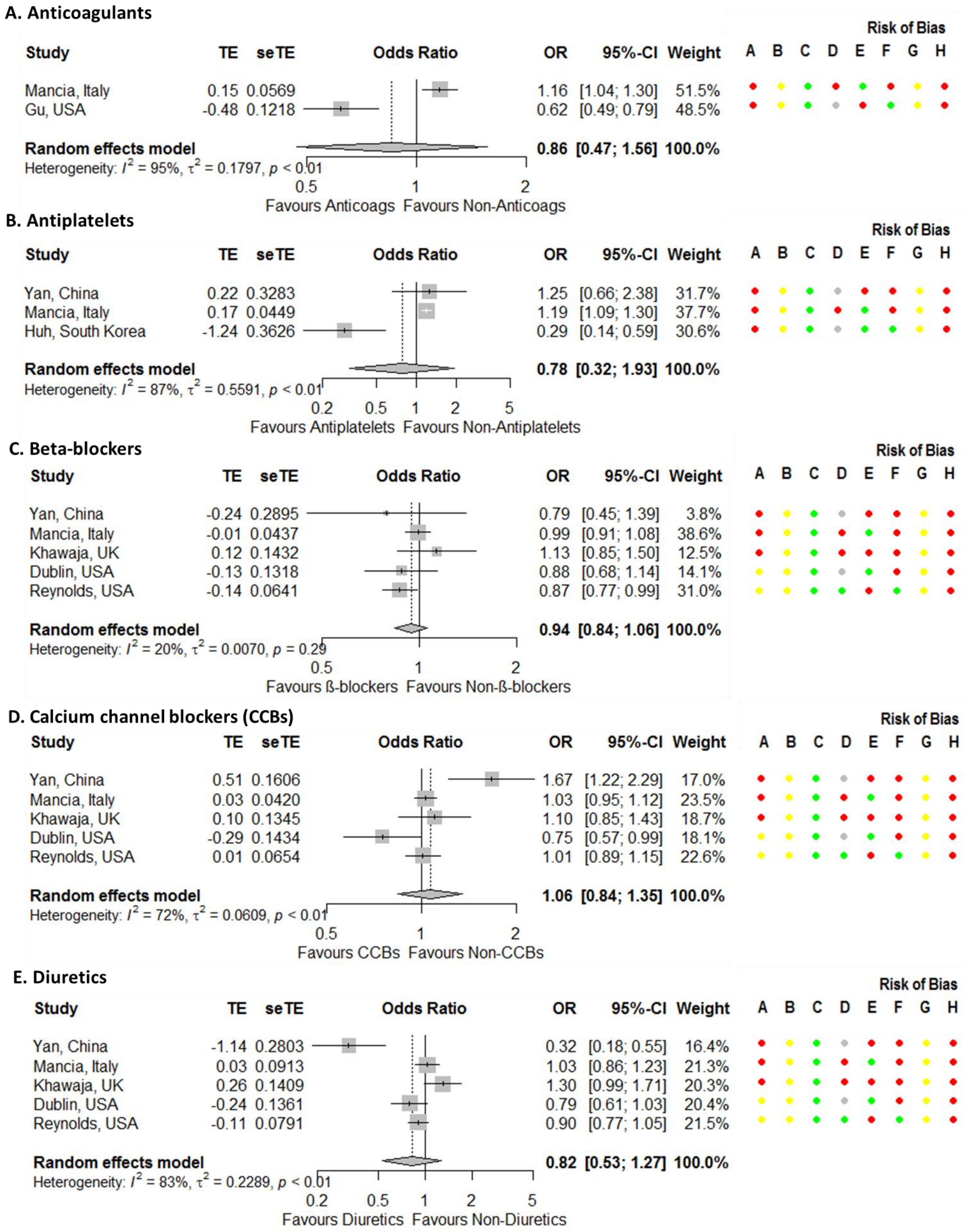


**Figure S9. Forest plots for association between testing positive for COVID-19 and being on cardiovascular drugs (adjusted odds ratios).** Risk of bias legend. A = risk of bias due to confounding, B = risk of bias in selection of participants into the study, C = risk of bias in classification of interventions, D = risk of bias due to deviations from intended interventions, E = risk of bias due to missing data, F = risk of bias in measurement of outcomes, G = risk of bias in selection of the reported result, H = overall risk of bias. Color codes. Colour codes. Red = serious, yellow = moderate, green = low, grey = unclear.

**F. Loop diuretics**

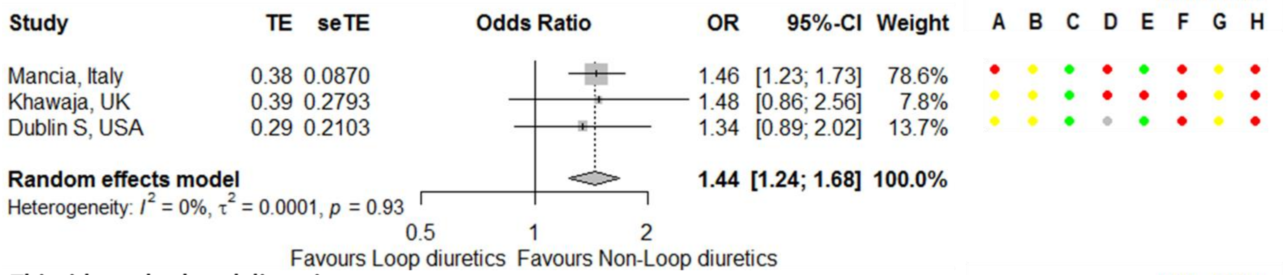


**G. Thiazide and related diuretics**

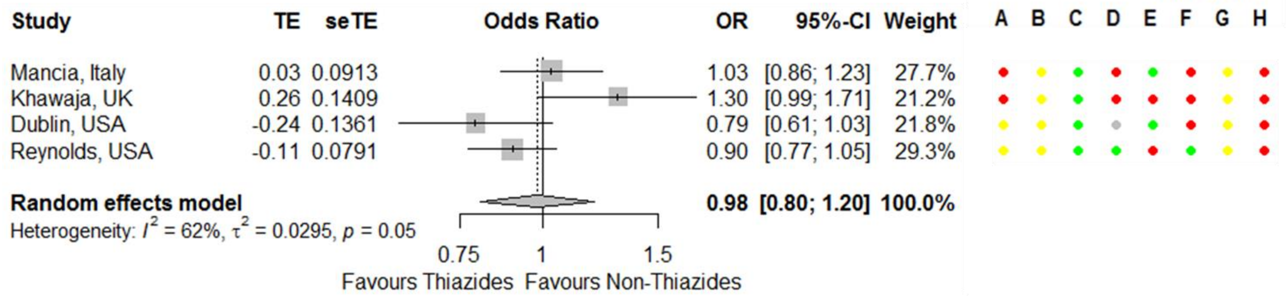


**H. Potassium-sparing diuretics and aldosterone antagonists (PSDAA)**

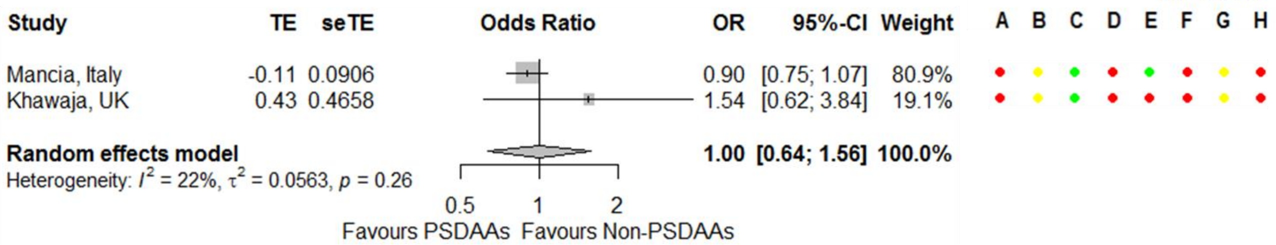


**Figure S9. Continued.**

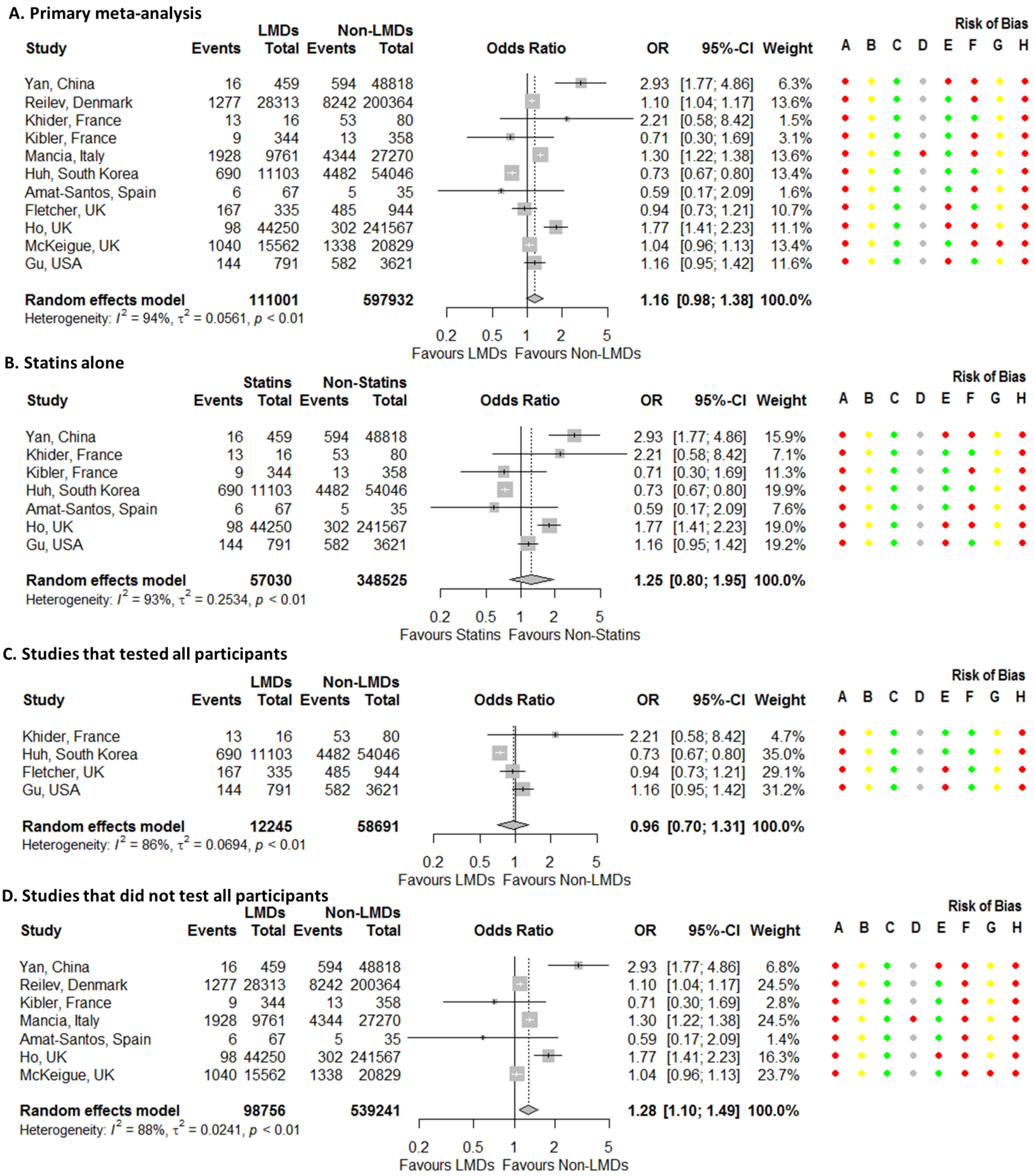


**Figure S10. Forest plots for association between testing positive for COVID-19 and being on a lipid modifying drug.** Risk of bias legend. A = risk of bias due to confounding, B = risk of bias in selection of participants into the study, C = risk of bias in classification of interventions, D = risk of bias due to deviations from intended interventions, E = risk of bias due to missing data, F = risk of bias in measurement of outcomes, G = risk of bias in selection of the reported result, H = overall risk of bias. Color codes. Colour codes. Red = serious, yellow = moderate, green = low, grey = unclear.

**E. Adjusted estimates**

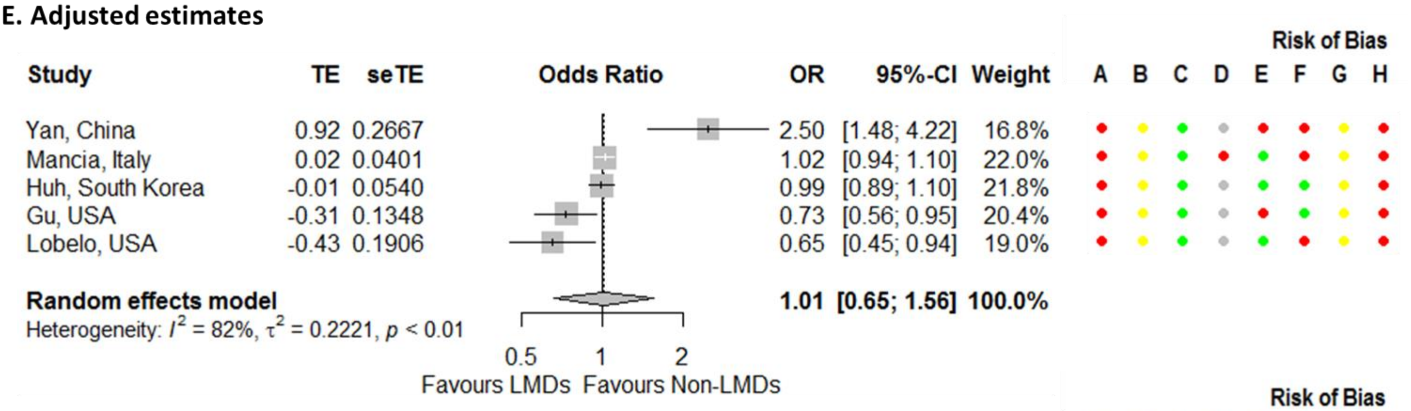


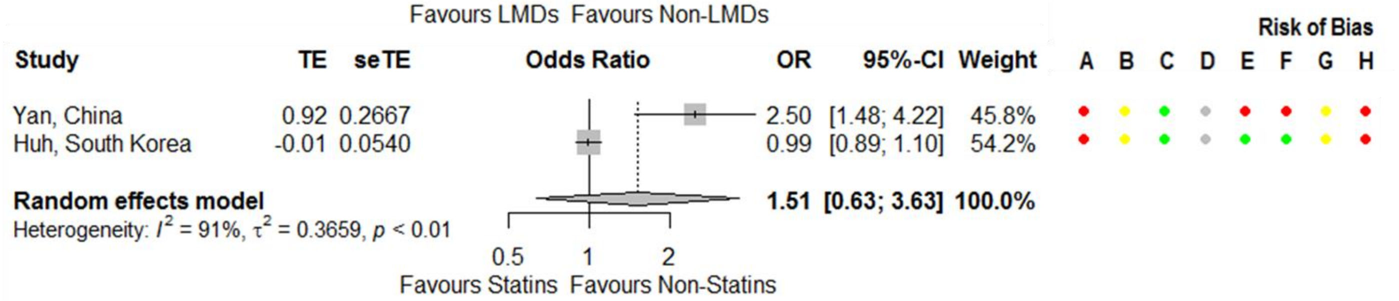


**Figure S10. Continued.**

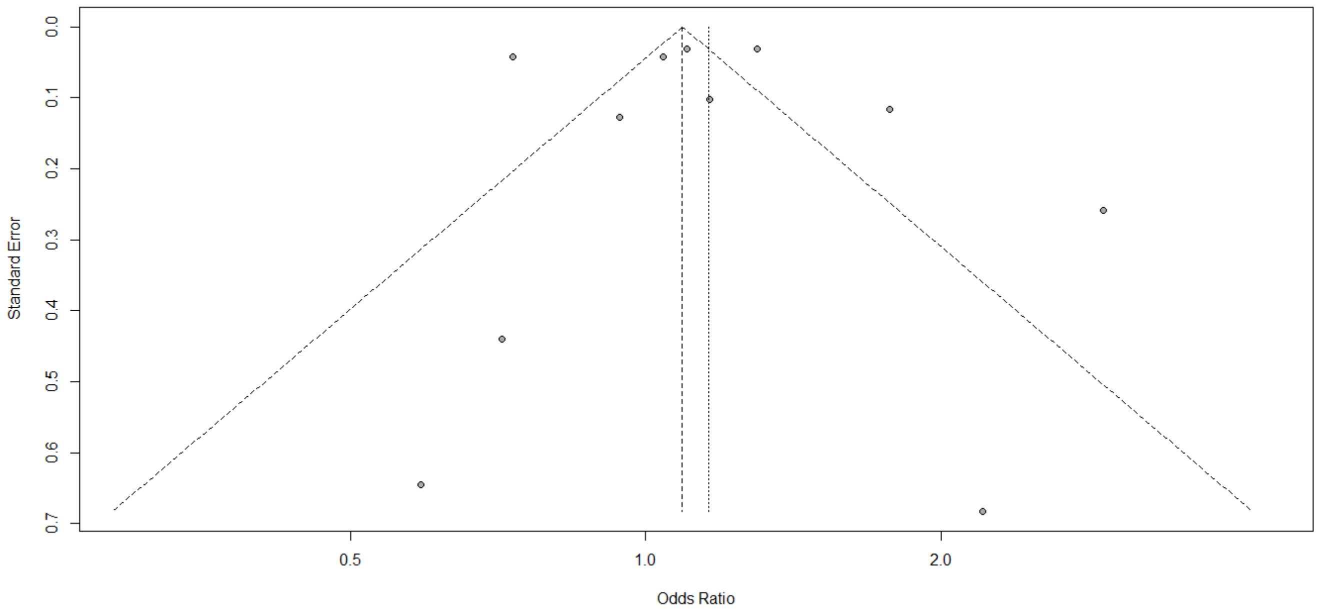


**Figure S11. Funnel plot for association between testing positive for COVID-19 and being on a lipid modifying drug.**

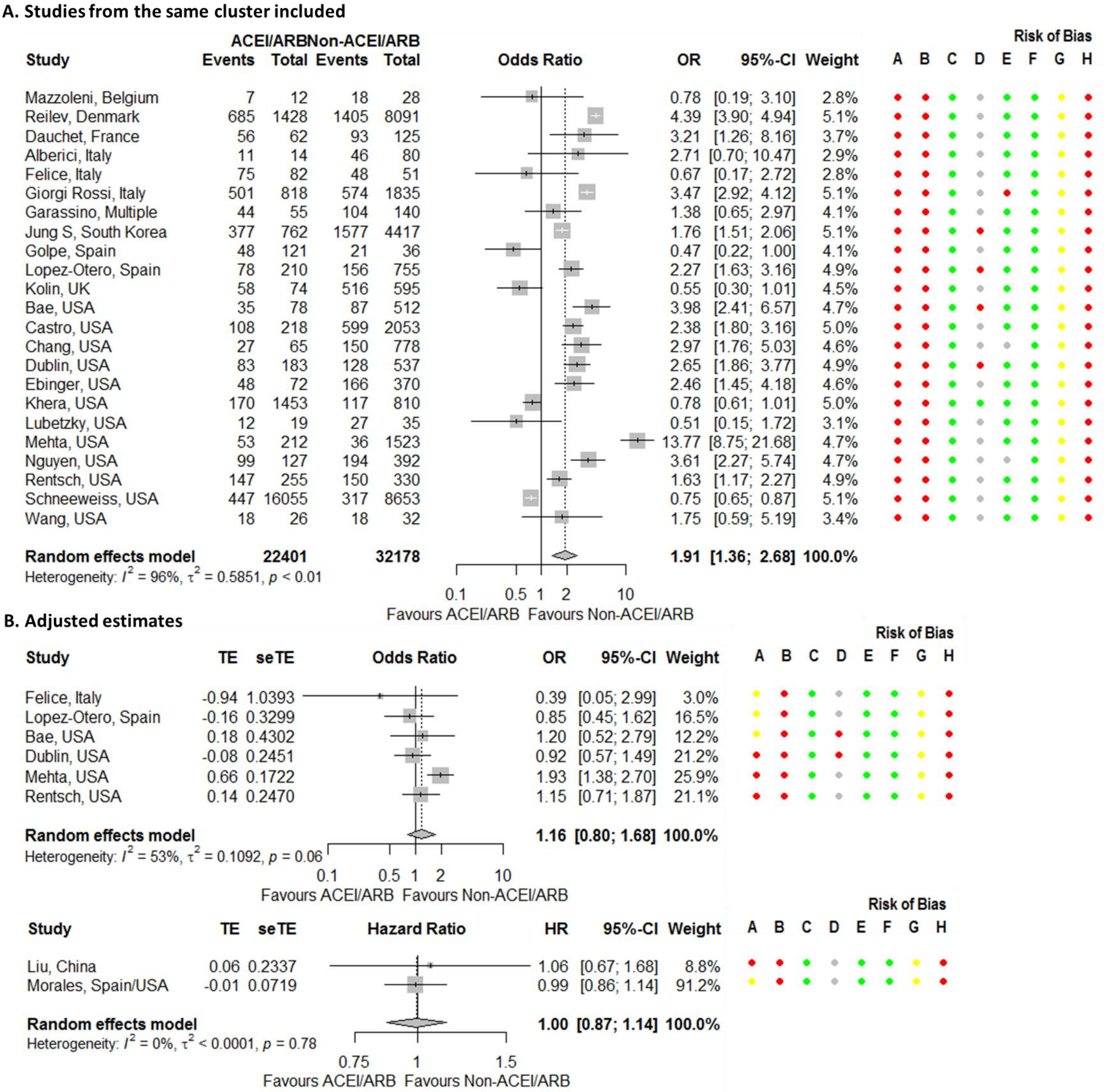


**Figure S12. Forest plot for association between being hospitalized for COVID-19 and being on an angiotensin-converting enzyme inhibitor (ACEI) or angiotensin receptor blocker (ARB).** Castro,[157] Chang,[158] Ebinger,[4] Garassino,[111] Giorgi Rossi,[97] Golpe,[131] and Nguyen[176] assume that none of the patients are taking both ACEis and ARBs. Risk of bias legend. A = risk of bias due to confounding, B = risk of bias in selection of participants into the study, C = risk of bias in classification of interventions, D = risk of bias due to deviations from intended interventions, E = risk of bias due to missing data, F = risk of bias in measurement of outcomes, G = risk of bias in selection of the reported result, H = overall risk of bias. Color codes. Colour codes. Red = serious, yellow = moderate, green = low, grey = unclear.

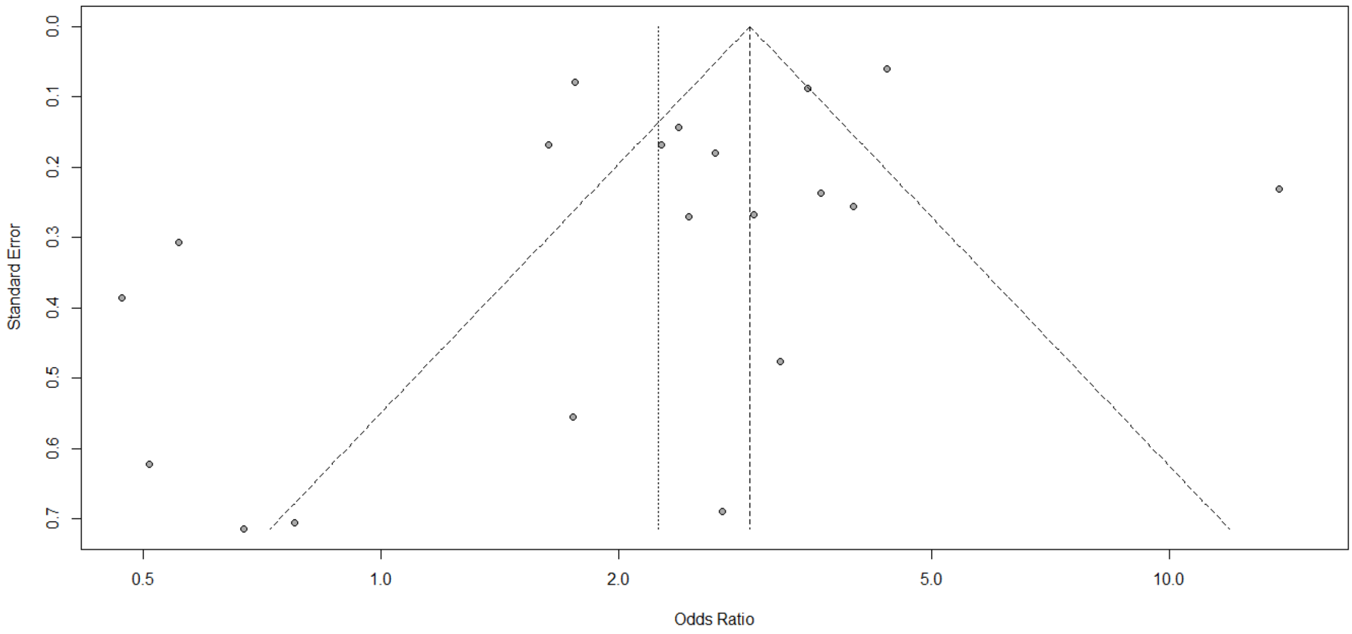


**Figure S13. Funnel plot for association between being hospitalized for COVID-19 and being on an angiotensin-converting enzyme inhibitor or angiotensin receptor blocker.**

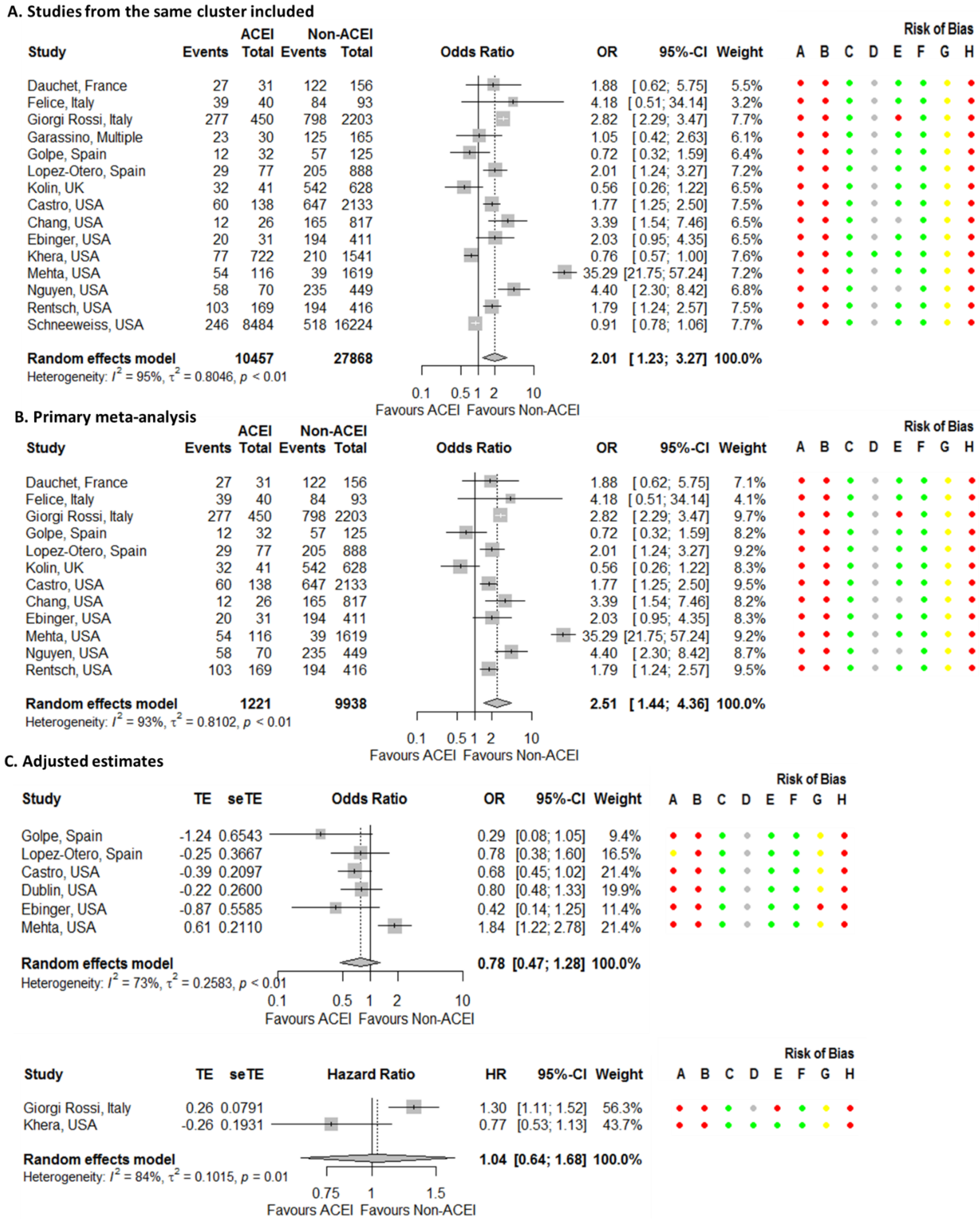


**Figure S14. Forest plot for association between being hospitalized for COVID-19 and being on an angiotensin-converting enzyme inhibitor (ACEI).** Risk of bias legend. A = risk of bias due to confounding, B = risk of bias in selection of participants into the study, C = risk of bias in classification of interventions, D = risk of bias due to deviations from intended interventions, E = risk of bias due to missing data, F = risk of bias in measurement of outcomes, G = risk of bias in selection of the reported result, H = overall risk of bias. Color codes. Colour codes. Red = serious, yellow = moderate, green = low, grey = unclear.

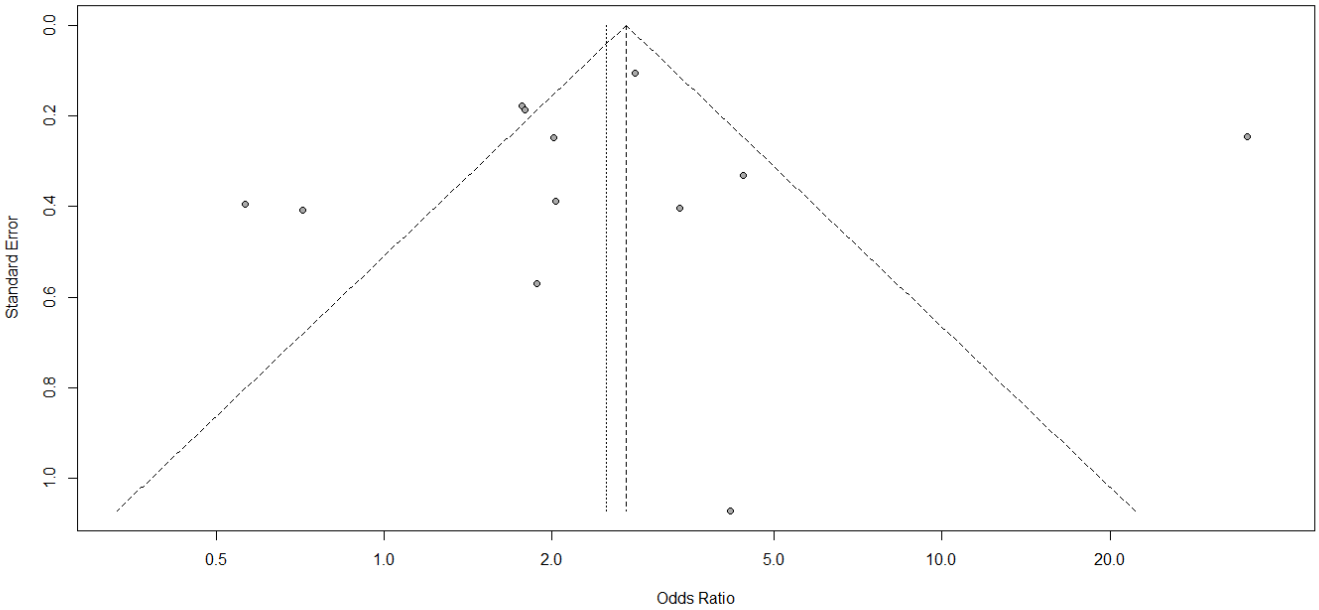


**Figure S15. Funnel plot for association between being hospitalized for COVID-19 and being on an angiotensin-converting enzyme inhibitor.**

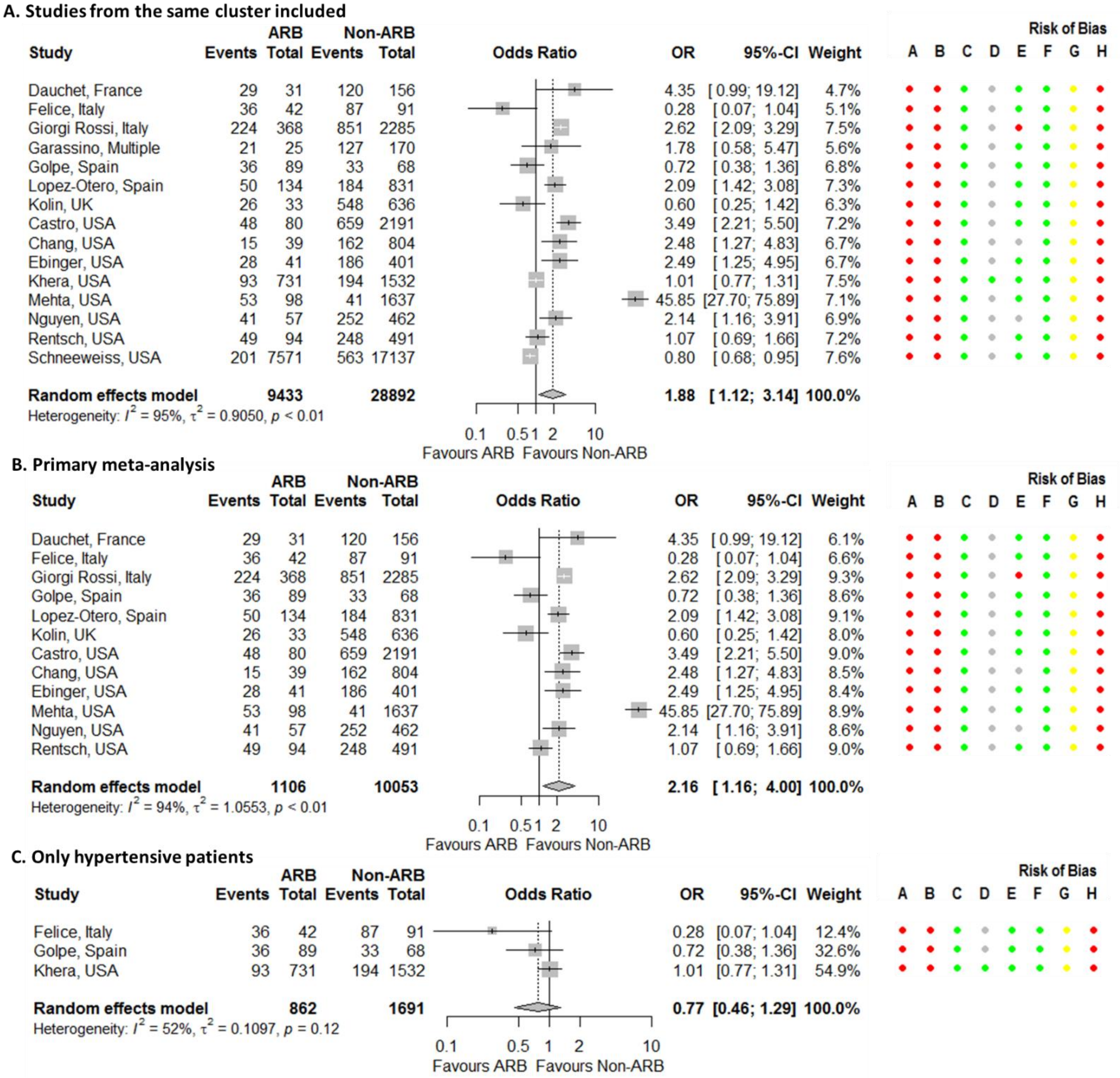


**Figure S16. Forest plot for association between being hospitalized for COVID-19 and being on an angiotensin receptor blocker (ARB) with studies from the same cluster all included.** Risk of bias legend. A = risk of bias due to confounding, B = risk of bias in selection of participants into the study, C = risk of bias in classification of interventions, D = risk of bias due to deviations from intended interventions, E = risk of bias due to missing data, F = risk of bias in measurement of outcomes, G = risk of bias in selection of the reported result, H = overall risk of bias. Color codes. Colour codes. Red = serious, yellow = moderate, green = low, grey = unclear.

**D. Adjusted estimates**

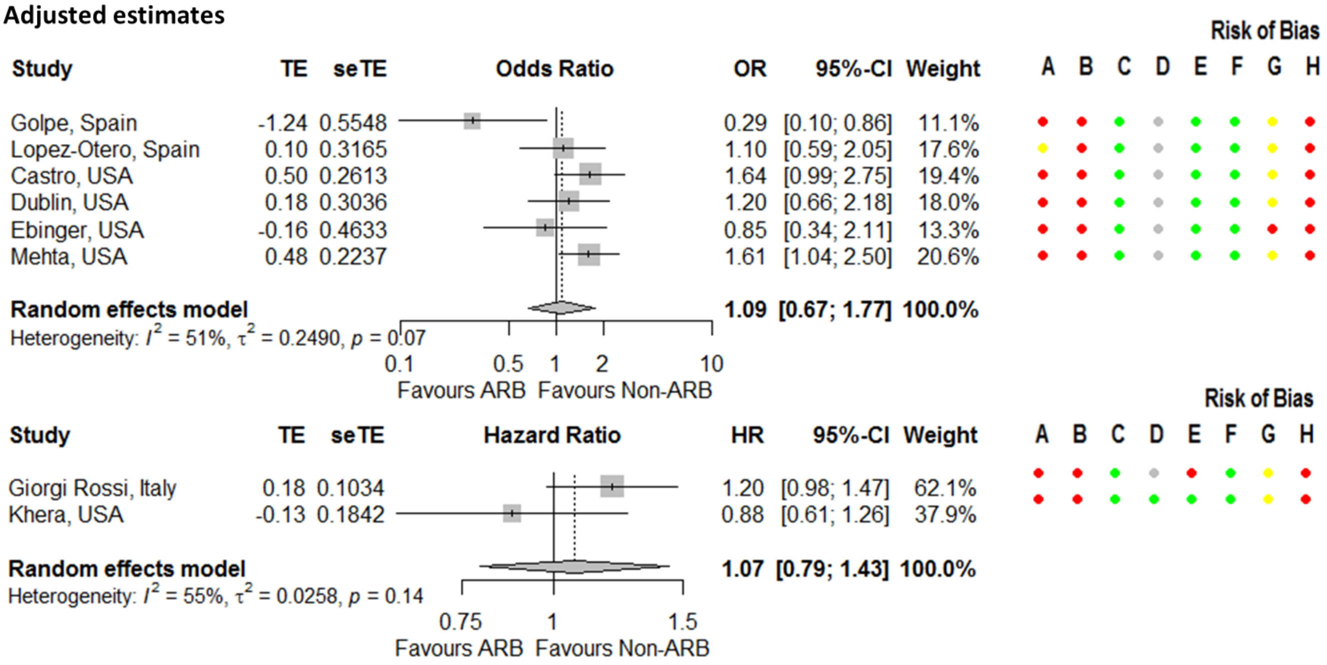


**Figure S16. Continued.**

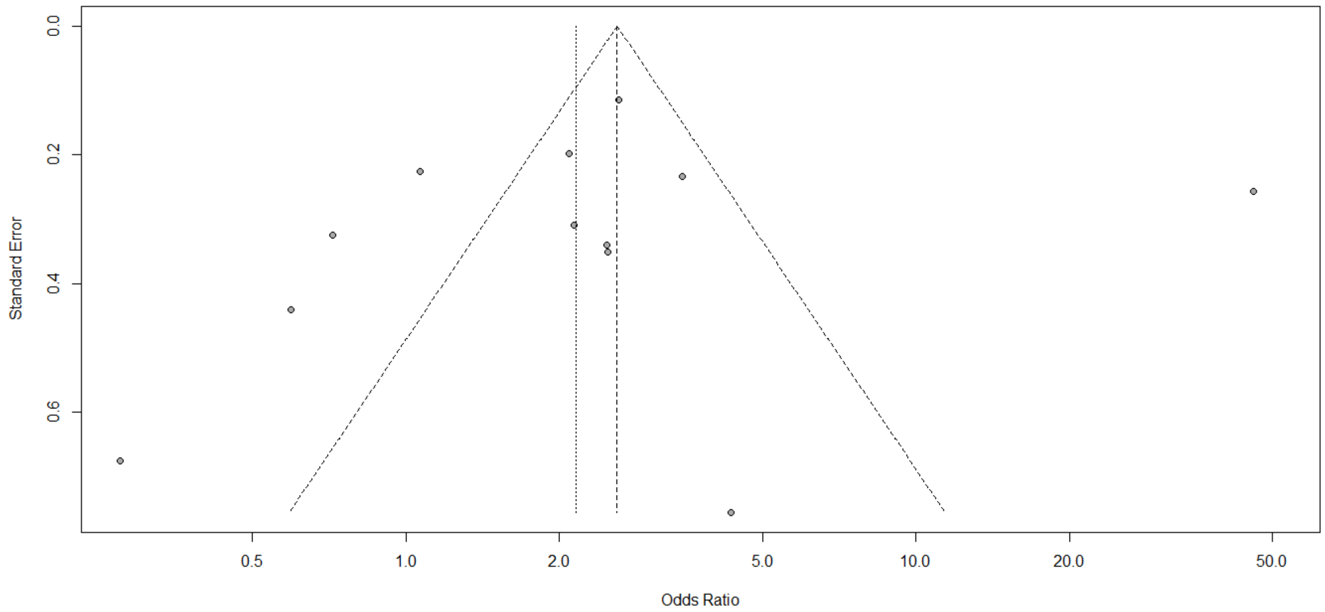


**Figure S17. Funnel plot for association between being hospitalized for COVID-19 and being on angiotensin receptor blocker.**

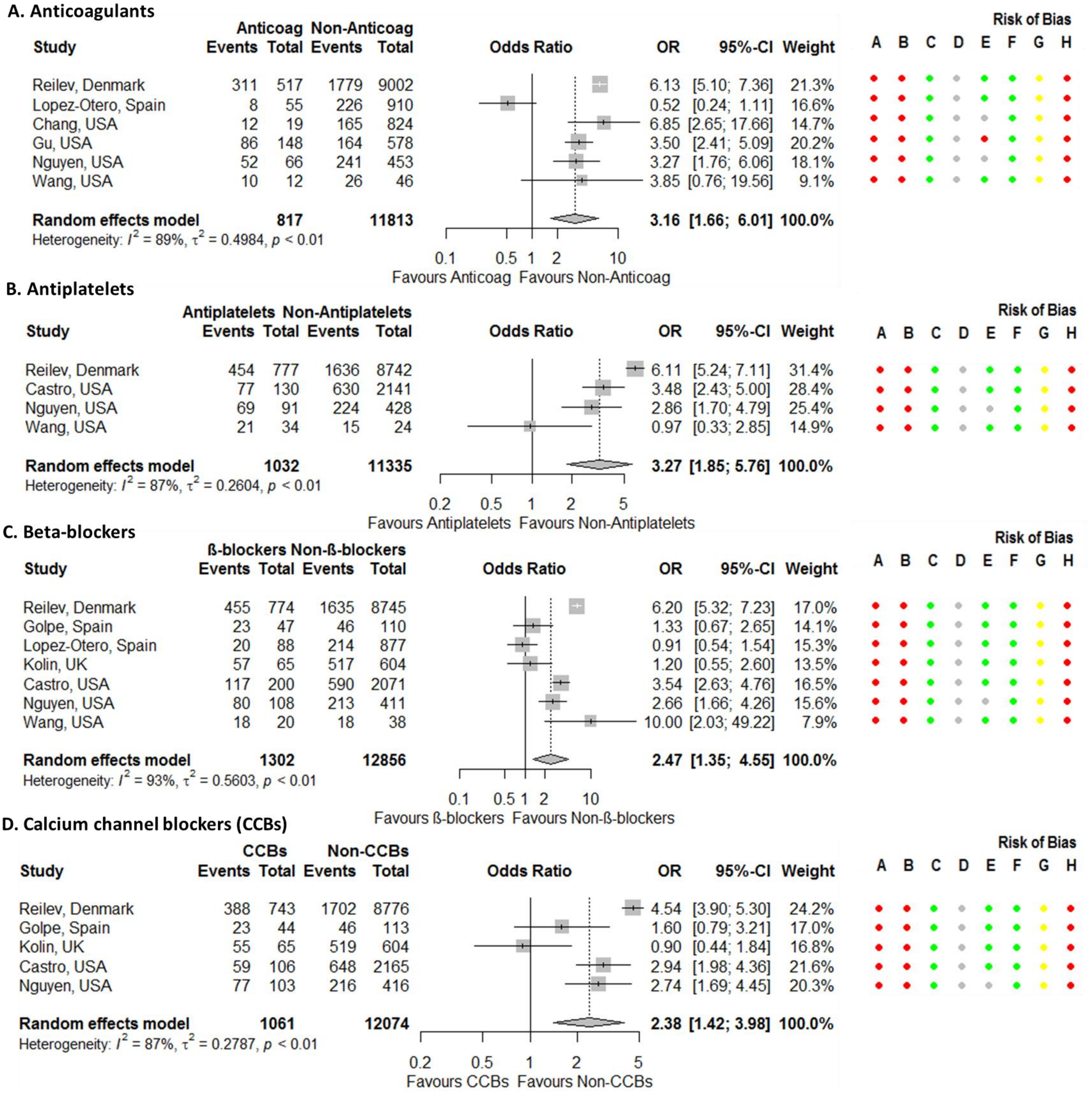


**Figure S18. Forest plots for association between being hospitalized for COVID-19 and being on cardiovascular drugs.** Castro[157] assumes none of the patients are taking two beta-blockers (atenolol, labetalol or metoprolol) or two statins (atorvastatin, rosuvastatin or simvastatin). Risk of bias legend. A = risk of bias due to confounding, B = risk of bias in selection of participants into the study, C = risk of bias in classification of interventions, D = risk of bias due to deviations from intended interventions, E = risk of bias due to missing data, F = risk of bias in measurement of outcomes, G = risk of bias in selection of the reported result, H = overall risk of bias. Color codes. Colour codes. Red = serious, yellow = moderate, green = low, grey = unclear.

**E. Diuretics**

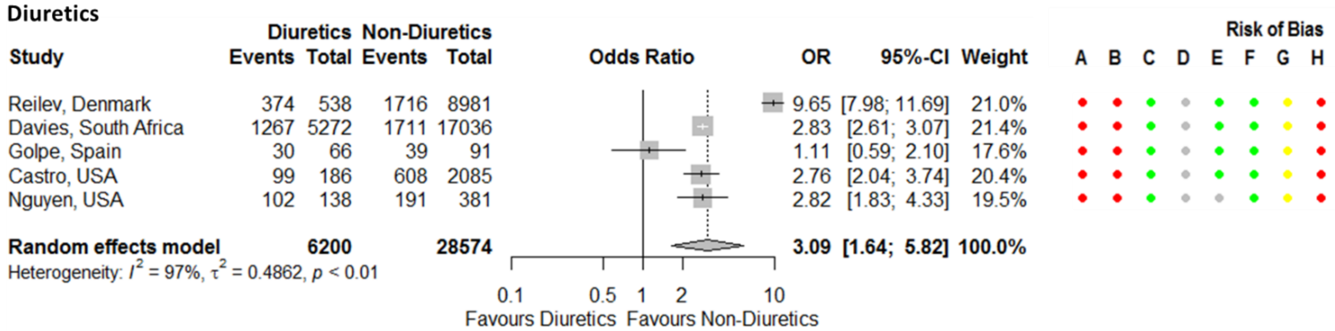


**F. Lipid modifying drugs (LMDs)**

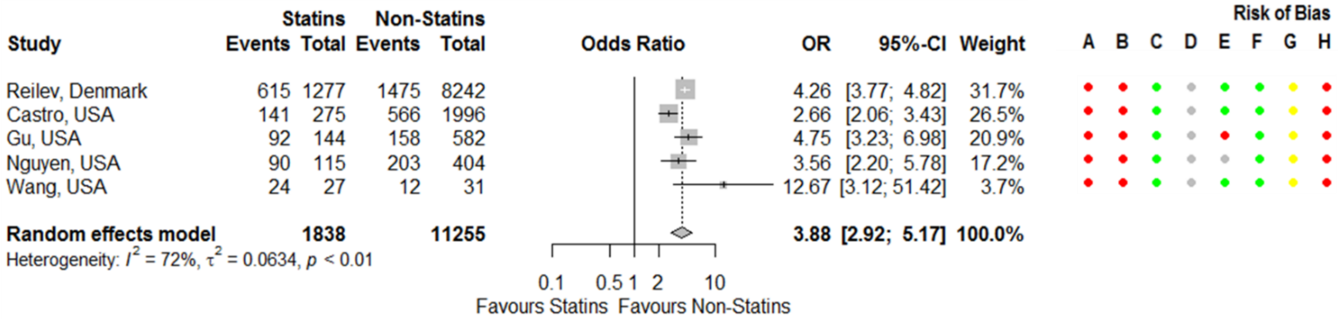


**Figure S18. Continued.**

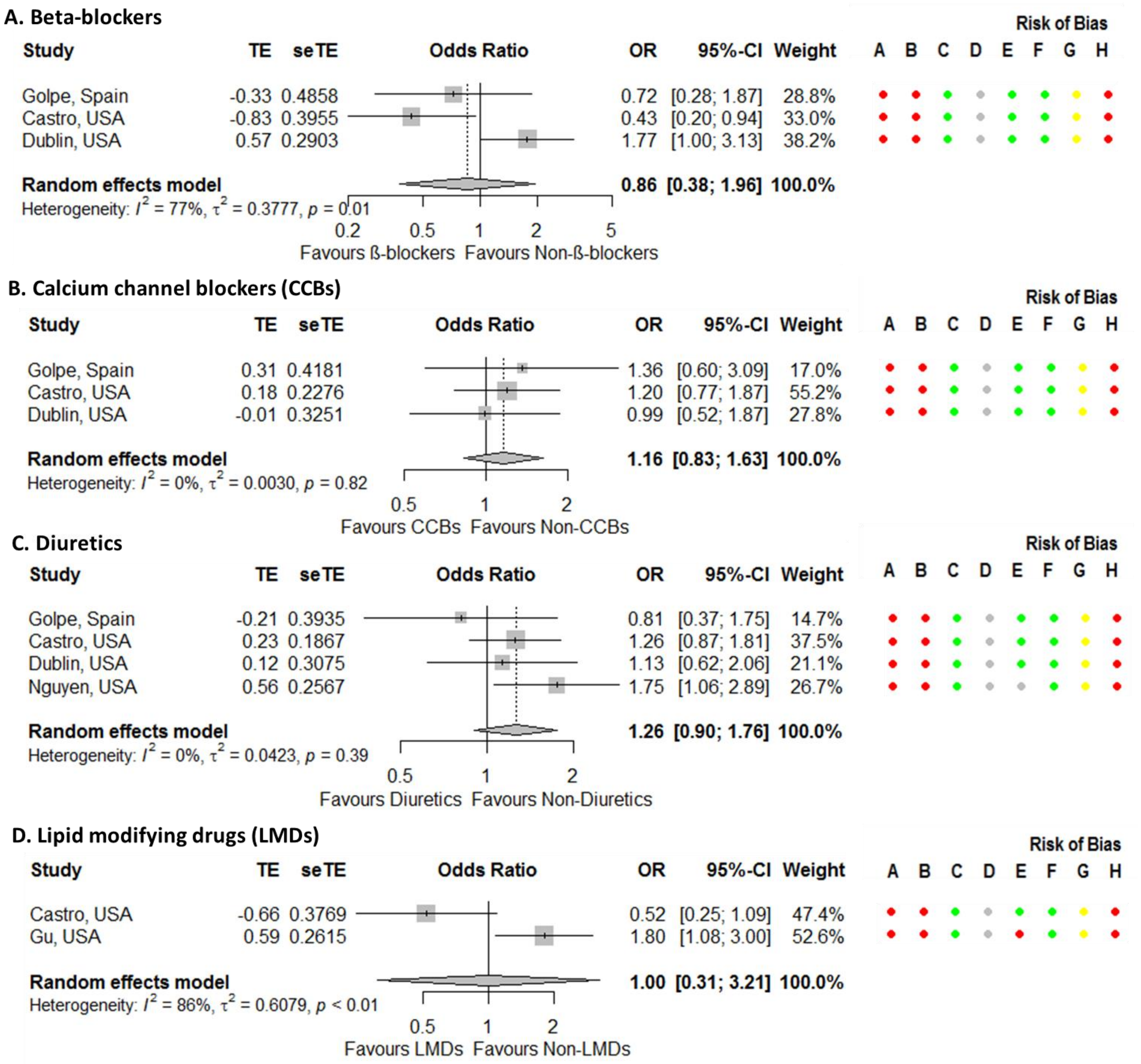


**Figure S19. Forest plots for association between being hospitalized for COVID-19 and being on cardiovascular drugs – adjusted estimates.** Risk of bias legend. A = risk of bias due to confounding, B = risk of bias in selection of participants into the study, C = risk of bias in classification of interventions, D = risk of bias due to deviations from intended interventions, E = risk of bias due to missing data, F = risk of bias in measurement of outcomes, G = risk of bias in selection of the reported result, H = overall risk of bias. Color codes. Colour codes. Red = serious, yellow = moderate, green = low, grey = unclear.

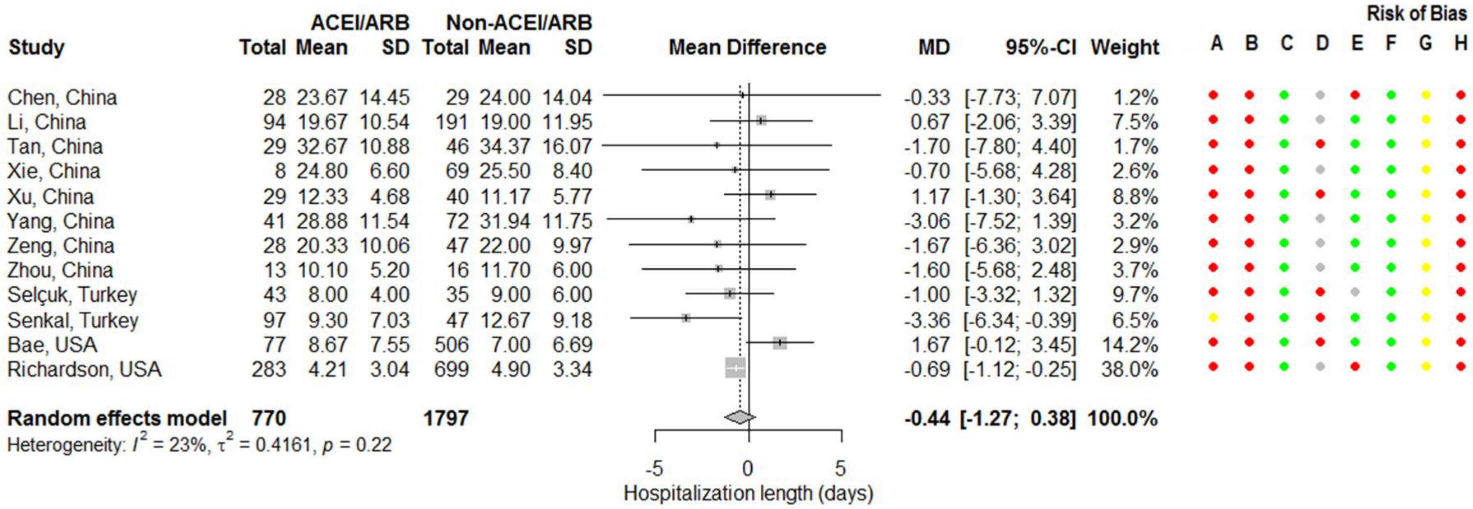


**Figure S20. Forest plot for association between length of hospitalization for COVID-19 and being on an angiotensin-converting enzyme inhibitor (ACEI) or angiotensin receptor blocker (ARB) with studies from the same cluster all included.** Risk of bias legend. A = risk of bias due to confounding, B = risk of bias in selection of participants into the study, C = risk of bias in classification of interventions, D = risk of bias due to deviations from intended interventions, E = risk of bias due to missing data, F = risk of bias in measurement of outcomes, G = risk of bias in selection of the reported result, H = overall risk of bias. Color codes. Colour codes. Red = serious, yellow = moderate, green = low, grey = unclear.

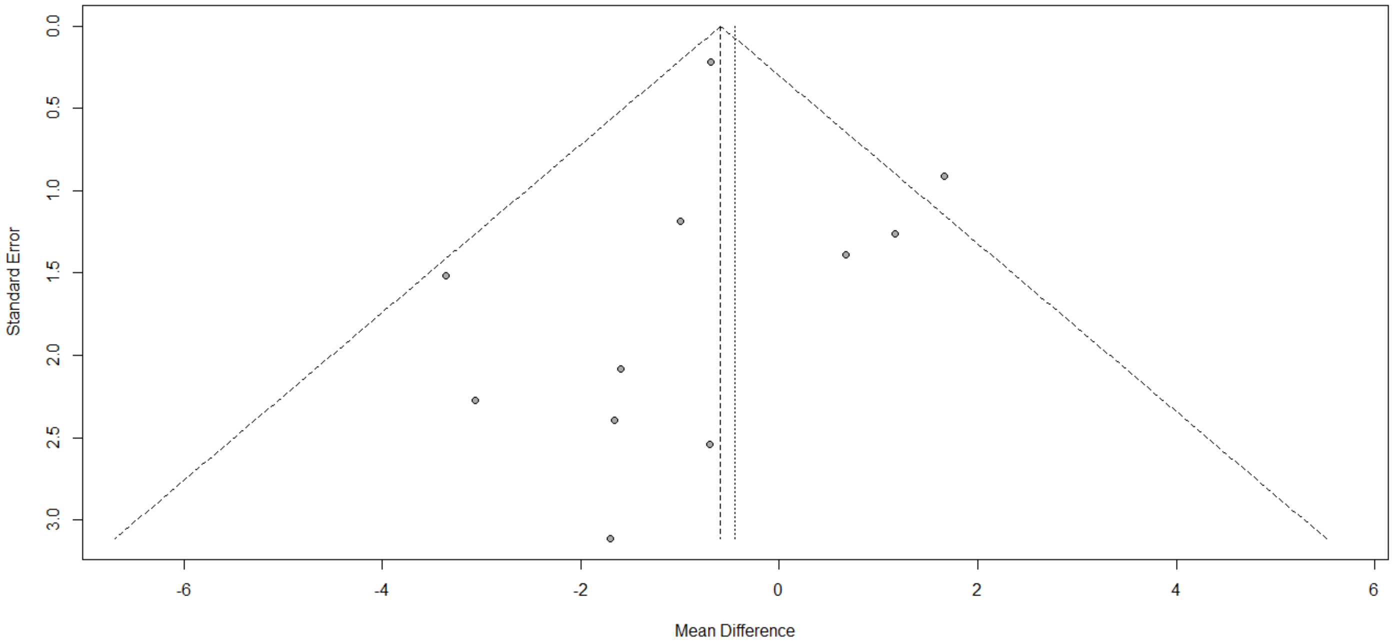


**Figure S21. Funnel plot for association between length of hospitalization for COVID-19 and being on an angiotensin-converting enzyme inhibitor or angiotensin receptor blocker.**

**A. Studies from the same cluster all included**

| Study | ACEI/ARB Events | ACEI/ARB Total | Non−ACEI/ARB Events | Non−ACEI/ARB Total | OR | 95%−CI | Weight |
|---|---|---|---|---|---|---|---|
| De Spiegeleer, Belgium | 6 | 30 | 31 | 124 | 0.75 | [0.28; 2.00] | 1.1% |
| Mazzoleni, Belgium | 7 | 12 | 18 | 28 | 0.78 | [0.19; 3.10] | 0.7% |
| Feng Y, China | 4 | 33 | 36 | 80 | 0.17 | [0.05; 0.52] | 0.9% |
| Feng Z, China | 1 | 16 | 16 | 49 | 0.14 | [0.02; 1.13] | 0.4% |
| Gao, China | 74 | 183 | 221 | 667 | 1.37 | [0.98; 1.92] | 1.9% |
| Hu, China | 28 | 65 | 33 | 84 | 1.17 | [0.61; 2.26] | 1.5% |
| Huang, China | 13 | 20 | 24 | 30 | 0.46 | [0.13; 1.67] | 0.8% |
| Li J, China | 57 | 115 | 116 | 247 | 1.11 | [0.71; 1.73] | 1.7% |
| Li T, China | 37 | 97 | 68 | 215 | 1.33 | [0.81; 2.20] | 1.7% |
| Li X, China | 19 | 42 | 247 | 503 | 0.86 | [0.45; 1.61] | 1.5% |
| Liu Y, China | 7 | 22 | 31 | 56 | 0.38 | [0.13; 1.07] | 1.0% |
| Liu Xiaofan, China | 4 | 5 | 16 | 29 | 3.25 | [0.32; 32.75] | 0.3% |
| Liu Xiulan, China | 36 | 74 | 39 | 83 | 1.07 | [0.57; 2.00] | 1.5% |
| Meng, China | 4 | 17 | 12 | 25 | 0.33 | [0.08; 1.31] | 0.8% |
| Peng, China | 3 | 22 | 13 | 90 | 0.94 | [0.24; 3.61] | 0.8% |
| Tan, China | 27 | 31 | 60 | 69 | 1.01 | [0.29; 3.58] | 0.8% |
| Xie Yang, China | 10 | 35 | 140 | 584 | 1.27 | [0.59; 2.71] | 1.3% |
| Xie Yangjing, China | 3 | 3 | 21 | 59 | 12.53 | [0.62; 254.25] | 0.2% |
| Xiong, China | 16 | 45 | 14 | 86 | 2.84 | [1.23; 6.55] | 1.2% |
| Xu, China | 11 | 40 | 22 | 61 | 0.67 | [0.28; 1.60] | 1.2% |
| Yan, China | 23 | 58 | 34 | 79 | 0.87 | [0.44; 1.73] | 1.4% |
| Yang, China | 15 | 43 | 35 | 83 | 0.73 | [0.34; 1.58] | 1.3% |
| Ye, China | 27 | 62 | 32 | 80 | 1.16 | [0.59; 2.27] | 1.4% |
| Zeng, China | 15 | 28 | 15 | 47 | 2.46 | [0.94; 6.45] | 1.1% |
| Zhang, China | 24 | 188 | 75 | 940 | 1.69 | [1.04; 2.75] | 1.7% |
| Zhou, Hong Kong (China) | 14 | 184 | 4 | 792 | 16.22 | [5.27; 49.90] | 0.9% |
| Fosbol, Denmark | 292 | 895 | 526 | 3585 | 2.82 | [2.38; 3.33] | 2.0% |
| Reilev, Denmark | 118 | 1428 | 182 | 8091 | 3.91 | [3.08; 4.97] | 1.9% |
| Feuth, Finland | 3 | 9 | 4 | 19 | 1.88 | [0.32; 11.02] | 0.5% |
| Allenbach, France | 6 | 25 | 41 | 122 | 0.62 | [0.23; 1.68] | 1.1% |
| Basse, France | 9 | 29 | 26 | 112 | 1.49 | [0.60; 3.66] | 1.2% |
| Benotmane, France | 8 | 15 | 11 | 25 | 1.45 | [0.40; 5.26] | 0.8% |
| Cariou, France | 232 | 737 | 150 | 580 | 1.32 | [1.03; 1.68] | 1.9% |
| Dauchet, France | 56 | 62 | 93 | 125 | 3.21 | [1.26; 8.16] | 1.1% |
| Liabeuf, France | 52 | 96 | 64 | 172 | 1.99 | [1.20; 3.31] | 1.7% |
| Meszaros, France | 32 | 68 | 12 | 35 | 1.70 | [0.73; 3.97] | 1.2% |
| Oussalah, France | 26 | 43 | 50 | 103 | 1.62 | [0.79; 3.34] | 1.4% |
| Sacco, Germany | 7 | 22 | 32 | 143 | 1.62 | [0.61; 4.31] | 1.1% |
| Ashraf, Iran | 4 | 19 | 11 | 81 | 1.70 | [0.48; 6.06] | 0.8% |
| Benelli, Italy | 42 | 110 | 70 | 301 | 2.04 | [1.28; 3.26] | 1.7% |
| Bravi, Italy | 267 | 450 | 379 | 1153 | 2.98 | [2.38; 3.73] | 2.0% |
| Felice, Italy | 61 | 82 | 44 | 51 | 0.46 | [0.18; 1.18] | 1.1% |
| Parigi, Italy | 57 | 114 | 91 | 211 | 1.32 | [0.83; 2.08] | 1.7% |
| Sardu, Italy | 23 | 45 | 9 | 17 | 0.93 | [0.30; 2.84] | 1.0% |
| Higuchi, Japan | 1 | 8 | 6 | 49 | 1.02 | [0.11; 9.84] | 0.4% |
| Choi HK, South Korea | 77 | 892 | 116 | 693 | 0.47 | [0.35; 0.64] | 1.9% |
| Choi MH, South Korea | 6 | 16 | 30 | 277 | 4.94 | [1.68; 14.56] | 1.0% |
| Jung, South Korea | 14 | 762 | 22 | 4417 | 3.74 | [1.90; 7.34] | 1.4% |
| Rhee, South Korea | 13 | 327 | 21 | 505 | 0.95 | [0.47; 1.93] | 1.4% |
| de Abajo, Spain | 215 | 497 | 178 | 642 | 1.99 | [1.55; 2.55] | 1.9% |
| Golpe, Spain | 48 | 121 | 21 | 36 | 0.47 | [0.22; 1.00] | 1.3% |
| Jurado, Spain | 56 | 191 | 45 | 111 | 0.61 | [0.37; 0.99] | 1.7% |
| Lopez−Otero, Spain | 13 | 210 | 20 | 755 | 2.43 | [1.19; 4.96] | 1.4% |
| Marcos, Spain | 122 | 298 | 241 | 620 | 1.09 | [0.82; 1.44] | 1.9% |
| Regina, Switzerland | 11 | 51 | 26 | 149 | 1.30 | [0.59; 2.87] | 1.3% |
| Pongpirul, Thailand | 4 | 17 | 28 | 176 | 1.63 | [0.49; 5.35] | 0.9% |
| Selçuk, Turkey | 37 | 74 | 7 | 39 | 4.57 | [1.79; 11.66] | 1.1% |
| Senkal, Turkey | 48 | 165 | 117 | 446 | 1.15 | [0.78; 1.72] | 1.8% |
| Bean, UK | 127 | 399 | 288 | 801 | 0.83 | [0.64; 1.07] | 1.9% |
| Russell, UK | 4 | 33 | 24 | 123 | 0.57 | [0.18; 1.77] | 0.9% |
| Argenziano, USA | 71 | 284 | 165 | 716 | 1.11 | [0.81; 1.53] | 1.9% |
| Bae, USA | 13 | 78 | 32 | 512 | 3.00 | [1.50; 6.01] | 1.4% |
| Castro, USA | 32 | 218 | 181 | 2053 | 1.78 | [1.19; 2.67] | 1.8% |
| Chang, USA | 6 | 27 | 50 | 150 | 0.57 | [0.22; 1.51] | 1.1% |
| Ebinger, USA | 48 | 72 | 166 | 370 | 2.46 | [1.45; 4.18] | 1.6% |
| Ferguson, USA | 7 | 13 | 14 | 59 | 3.75 | [1.08; 13.02] | 0.8% |
| Garibaldi, USA | 55 | 151 | 174 | 596 | 1.39 | [0.95; 2.02] | 1.8% |
| Goshua, USA | 15 | 18 | 33 | 50 | 2.58 | [0.65; 10.14] | 0.8% |
| Jillella, USA | 2 | 3 | 4 | 5 | 0.50 | [0.02; 12.90] | 0.2% |
| Kim, USA | 201 | 573 | 417 | 1322 | 1.17 | [0.95; 1.44] | 2.0% |
| Mehta, USA | 22 | 212 | 15 | 1523 | 11.64 | [5.94; 22.83] | 1.4% |
| Palaiodimos, USA | 26 | 62 | 64 | 138 | 0.84 | [0.46; 1.53] | 1.5% |
| Rentsch, USA | 69 | 255 | 53 | 330 | 1.94 | [1.30; 2.90] | 1.8% |
| Reynolds, USA | 336 | 1374 | 666 | 4520 | 1.87 | [1.62; 2.17] | 2.0% |
| Richardson, USA | 87 | 413 | 141 | 953 | 1.54 | [1.14; 2.07] | 1.9% |
| Schneeweiss, USA | 271 | 16055 | 163 | 8653 | 0.89 | [0.74; 1.09] | 2.0% |
| **Random effects model** | | **29588** | | **51905** | **1.39** | **[1.20; 1.61]** | **100.0%** |

Heterogeneity: $I^2 = 84\%$, $\tau^2 = 0.2822$, $p < 0.01$

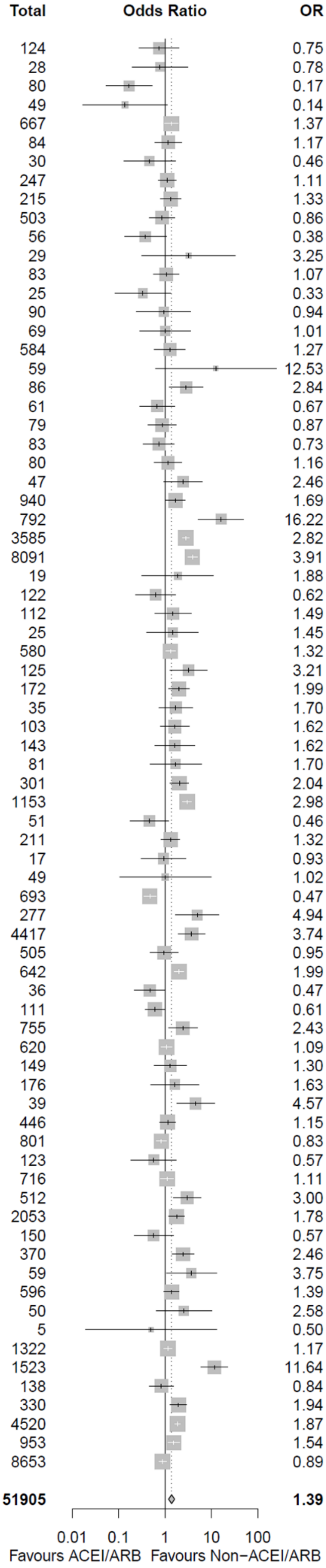


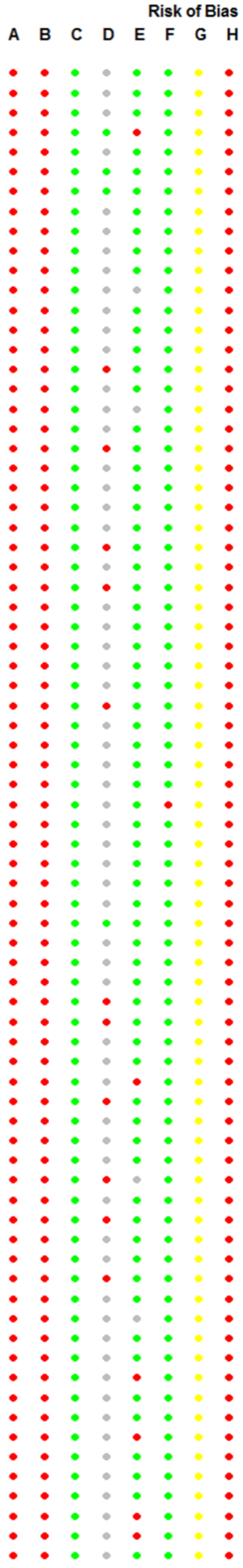

**B. China-only studies**

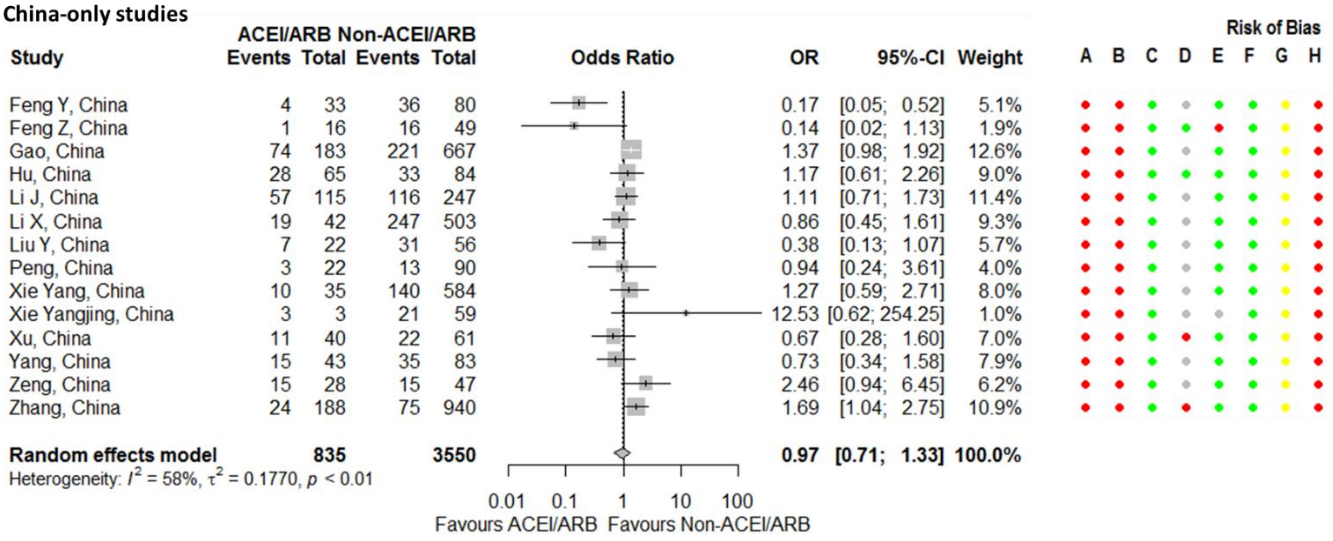


**Figure S22. Forest plot for association between severity outcomes in COVID-19 patients and being on an angiotensin-converting enzyme inhibitor (ACEI) or angiotensin receptor blocker (ARB).** Benelli,[89] Castro,[157] Chang,[158] Dauchet,[5] Ebinger,[4] Ferguson,[160] Feuth,[72] Golpe,[131] Kim,[123] Liu Y,[46] Marcos,[134] Pongpirul,[139] and Yan[56] estimates assume that none of the patients are taking both ACEis and ARBs. Risk of bias legend. A = risk of bias due to confounding, B = risk of bias in selection of participants into the study, C = risk of bias in classification of interventions, D = risk of bias due to deviations from intended interventions, E = risk of bias due to missing data, F = risk of bias in measurement of outcomes, G = risk of bias in selection of the reported result, H = overall risk of bias. Color codes. Colour codes. Red = serious, yellow = moderate, green = low, grey = unclear.

### C. Not in China

| Study | ACEI/ARB Events | ACEI/ARB Total | Non-ACEI/ARB Events | Non-ACEI/ARB Total | OR | 95%-CI | Weight |
|---|---|---|---|---|---|---|---|
| De Spiegeleer, Belgium | 6 | 30 | 31 | 124 | 0.75 | [0.28; 2.00] | 1.7% |
| Mazzoleni, Belgium | 7 | 12 | 18 | 28 | 0.78 | [0.19; 3.10] | 1.2% |
| Zhou, Hong Kong (China) | 14 | 184 | 4 | 792 | 16.22 | [5.27; 49.90] | 1.5% |
| Reilev, Denmark | 118 | 1428 | 182 | 8091 | 3.91 | [3.08; 4.97] | 3.3% |
| Feuth, Finland | 3 | 9 | 4 | 19 | 1.88 | [0.32; 11.02] | 0.8% |
| Allenbach, France | 6 | 25 | 41 | 122 | 0.62 | [0.23; 1.68] | 1.7% |
| Basse, France | 9 | 29 | 26 | 112 | 1.49 | [0.60; 3.66] | 1.9% |
| Benotmane, France | 8 | 15 | 11 | 25 | 1.45 | [0.40; 5.26] | 1.3% |
| Dauchet, France | 56 | 62 | 93 | 125 | 3.21 | [1.26; 8.16] | 1.8% |
| Liabeuf, France | 52 | 96 | 64 | 172 | 1.99 | [1.20; 3.31] | 2.7% |
| Meszaros, France | 32 | 68 | 12 | 35 | 1.70 | [0.73; 3.97] | 2.0% |
| Oussalah, France | 26 | 43 | 50 | 103 | 1.62 | [0.79; 3.34] | 2.3% |
| Sacco, Germany | 7 | 22 | 32 | 143 | 1.62 | [0.61; 4.31] | 1.7% |
| Ashraf, Iran | 4 | 19 | 11 | 81 | 1.70 | [0.48; 6.06] | 1.3% |
| Benelli, Italy | 42 | 110 | 70 | 301 | 2.04 | [1.28; 3.26] | 2.8% |
| Bravi, Italy | 267 | 450 | 379 | 1153 | 2.98 | [2.38; 3.73] | 3.3% |
| Felice, Italy | 61 | 82 | 44 | 51 | 0.46 | [0.18; 1.18] | 1.8% |
| Parigi, Italy | 57 | 114 | 91 | 211 | 1.32 | [0.83; 2.08] | 2.8% |
| Sardu, Italy | 23 | 45 | 9 | 17 | 0.93 | [0.30; 2.84] | 1.5% |
| Higuchi, Japan | 1 | 8 | 6 | 49 | 1.02 | [0.11; 9.84] | 0.6% |
| Choi MH, South Korea | 6 | 16 | 30 | 277 | 4.94 | [1.68; 14.56] | 1.6% |
| Jung, South Korea | 14 | 762 | 22 | 4417 | 3.74 | [1.90; 7.34] | 2.4% |
| de Abajo, Spain | 215 | 497 | 178 | 642 | 1.99 | [1.55; 2.55] | 3.3% |
| Golpe, Spain | 48 | 121 | 21 | 36 | 0.47 | [0.22; 1.00] | 2.2% |
| Jurado, Spain | 56 | 191 | 45 | 111 | 0.61 | [0.37; 0.99] | 2.8% |
| Lopez−Otero, Spain | 13 | 210 | 20 | 755 | 2.43 | [1.19; 4.96] | 2.3% |
| Marcos, Spain | 122 | 298 | 241 | 620 | 1.09 | [0.82; 1.44] | 3.2% |
| Regina, Switzerland | 11 | 51 | 26 | 149 | 1.30 | [0.59; 2.87] | 2.1% |
| Pongpirul, Thailand | 4 | 17 | 28 | 176 | 1.63 | [0.49; 5.35] | 1.4% |
| Selçuk, Turkey | 37 | 74 | 7 | 39 | 4.57 | [1.79; 11.66] | 1.8% |
| Senkal, Turkey | 48 | 165 | 117 | 446 | 1.15 | [0.78; 1.72] | 3.0% |
| Bean, UK | 127 | 399 | 288 | 801 | 0.83 | [0.64; 1.07] | 3.2% |
| Russell, UK | 4 | 33 | 24 | 123 | 0.57 | [0.18; 1.77] | 1.5% |
| Argenziano, USA | 71 | 284 | 165 | 716 | 1.11 | [0.81; 1.53] | 3.1% |
| Bae, USA | 13 | 78 | 32 | 512 | 3.00 | [1.50; 6.01] | 2.3% |
| Castro, USA | 32 | 218 | 181 | 2053 | 1.78 | [1.19; 2.67] | 3.0% |
| Ebinger, USA | 48 | 72 | 166 | 370 | 2.46 | [1.45; 4.18] | 2.7% |
| Ferguson, USA | 7 | 13 | 14 | 59 | 3.75 | [1.08; 13.02] | 1.3% |
| Garibaldi, USA | 55 | 151 | 174 | 596 | 1.39 | [0.95; 2.02] | 3.0% |
| Goshua, USA | 15 | 18 | 33 | 50 | 2.58 | [0.65; 10.14] | 1.2% |
| Jillella, USA | 2 | 3 | 4 | 5 | 0.50 | [0.02; 12.90] | 0.3% |
| Mehta, USA | 22 | 212 | 15 | 1523 | 11.64 | [5.94; 22.83] | 2.4% |
| Palaiodimos, USA | 26 | 62 | 64 | 138 | 0.84 | [0.46; 1.53] | 2.5% |
| Rentsch, USA | 69 | 255 | 53 | 330 | 1.94 | [1.30; 2.90] | 3.0% |
| Reynolds, USA | 336 | 1374 | 666 | 4520 | 1.87 | [1.62; 2.17] | 3.4% |
| Richardson, USA | 87 | 413 | 141 | 953 | 1.54 | [1.14; 2.07] | 3.2% |
| **Random effects model** | | **8838** | | **32171** | **1.70** | **[1.41; 2.04]** | **100.0%** |

Heterogeneity: $I^2 = 82\%$, $\tau^2 = 0.2543$, $p < 0.01$

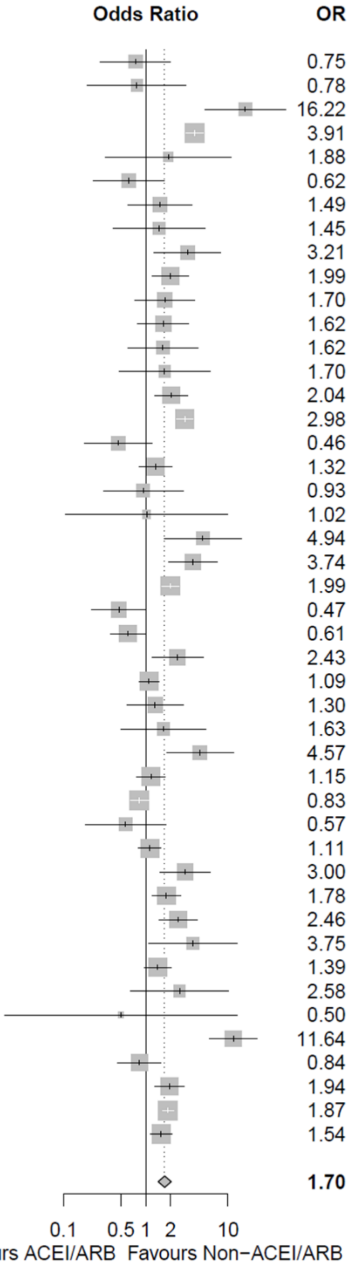


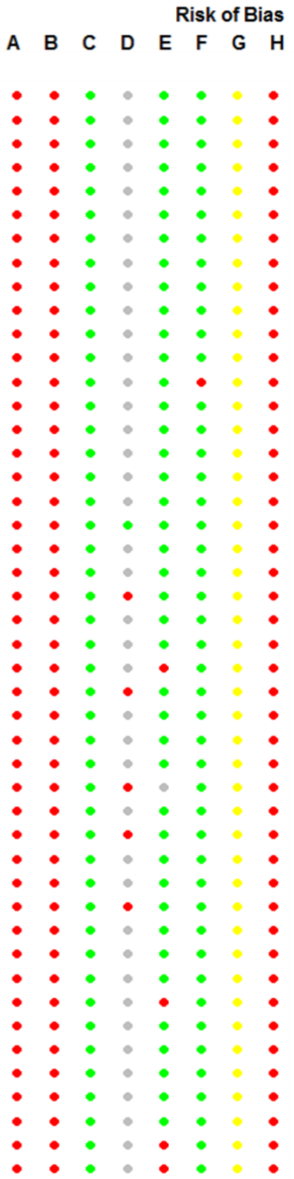


### D. South Korea-only studies

| Study | ACEI/ARB Events | ACEI/ARB Total | Non-ACEI/ARB Events | Non-ACEI/ARB Total | OR | 95%-CI | Weight |
|---|---|---|---|---|---|---|---|
| Choi MH, South Korea | 6 | 16 | 30 | 277 | 4.94 | [1.68; 14.56] | 28.0% |
| Jung, South Korea | 14 | 762 | 22 | 4417 | 3.74 | [1.90; 7.34] | 72.0% |
| **Random effects model** | | **778** | | **4694** | **4.04** | **[2.28; 7.16]** | **100.0%** |

Heterogeneity: $I^2 = 0\%$, $\tau^2 = 0$, $p = 0.67$

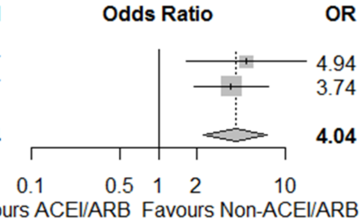


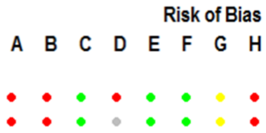


**Figure S22. Continued.**

### E. Not in South Korea

| Study | ACEI/ARB Events | ACEI/ARB Total | Non−ACEI/ARB Events | Non−ACEI/ARB Total | OR | 95%−CI | Weight |
|---|---|---|---|---|---|---|---|
| De Spiegeleer, Belgium | 6 | 30 | 31 | 124 | 0.75 | [0.28; 2.00] | 1.4% |
| Mazzoleni, Belgium | 7 | 12 | 18 | 28 | 0.78 | [0.19; 3.10] | 1.0% |
| Feng Y, China | 4 | 33 | 36 | 80 | 0.17 | [0.05; 0.52] | 1.2% |
| Feng Z, China | 1 | 16 | 16 | 49 | 0.14 | [0.02; 1.13] | 0.5% |
| Gao, China | 74 | 183 | 221 | 667 | 1.37 | [0.98; 1.92] | 2.5% |
| Hu, China | 28 | 65 | 33 | 84 | 1.17 | [0.61; 2.26] | 1.9% |
| Li J, China | 57 | 115 | 116 | 247 | 1.11 | [0.71; 1.73] | 2.3% |
| Li X, China | 19 | 42 | 247 | 503 | 0.86 | [0.45; 1.61] | 2.0% |
| Liu Y, China | 7 | 22 | 31 | 56 | 0.38 | [0.13; 1.07] | 1.3% |
| Peng, China | 3 | 22 | 13 | 90 | 0.94 | [0.24; 3.61] | 1.0% |
| Xie Yang, China | 10 | 35 | 140 | 584 | 1.27 | [0.59; 2.71] | 1.8% |
| Xie Yangjing, China | 3 | 3 | 21 | 59 | 12.53 | [0.62; 254.25] | 0.3% |
| Xu, China | 11 | 40 | 22 | 61 | 0.67 | [0.28; 1.60] | 1.6% |
| Yang, China | 15 | 43 | 35 | 83 | 0.73 | [0.34; 1.58] | 1.8% |
| Zeng, China | 15 | 28 | 15 | 47 | 2.46 | [0.94; 6.45] | 1.4% |
| Zhang, China | 24 | 188 | 75 | 940 | 1.69 | [1.04; 2.75] | 2.3% |
| Zhou, Hong Kong (China) | 14 | 184 | 4 | 792 | 16.22 | [5.27; 49.90] | 1.2% |
| Reilev, Denmark | 118 | 1428 | 182 | 8091 | 3.91 | [3.08; 4.97] | 2.6% |
| Feuth, Finland | 3 | 9 | 4 | 19 | 1.88 | [0.32; 11.02] | 0.7% |
| Allenbach, France | 6 | 25 | 41 | 122 | 0.62 | [0.23; 1.68] | 1.4% |
| Basse, France | 9 | 29 | 26 | 112 | 1.49 | [0.60; 3.66] | 1.5% |
| Benotmane, France | 8 | 15 | 11 | 25 | 1.45 | [0.40; 5.26] | 1.0% |
| Dauchet, France | 56 | 62 | 93 | 125 | 3.21 | [1.26; 8.16] | 1.5% |
| Liabeuf, France | 52 | 96 | 64 | 172 | 1.99 | [1.20; 3.31] | 2.2% |
| Meszaros, France | 32 | 68 | 12 | 35 | 1.70 | [0.73; 3.97] | 1.6% |
| Oussalah, France | 26 | 43 | 50 | 103 | 1.62 | [0.79; 3.34] | 1.8% |
| Sacco, Germany | 7 | 22 | 32 | 143 | 1.62 | [0.61; 4.31] | 1.4% |
| Ashraf, Iran | 4 | 19 | 11 | 81 | 1.70 | [0.48; 6.06] | 1.1% |
| Benelli, Italy | 42 | 110 | 70 | 301 | 2.04 | [1.28; 3.26] | 2.3% |
| Bravi, Italy | 267 | 450 | 379 | 1153 | 2.98 | [2.38; 3.73] | 2.7% |
| Felice, Italy | 61 | 82 | 44 | 51 | 0.46 | [0.18; 1.18] | 1.5% |
| Parigi, Italy | 57 | 114 | 91 | 211 | 1.32 | [0.83; 2.08] | 2.3% |
| Sardu, Italy | 23 | 45 | 9 | 17 | 0.93 | [0.30; 2.84] | 1.2% |
| Higuchi, Japan | 1 | 8 | 6 | 49 | 1.02 | [0.11; 9.84] | 0.5% |
| de Abajo, Spain | 215 | 497 | 178 | 642 | 1.99 | [1.55; 2.55] | 2.6% |
| Golpe, Spain | 48 | 121 | 21 | 36 | 0.47 | [0.22; 1.00] | 1.8% |
| Jurado, Spain | 56 | 191 | 45 | 111 | 0.61 | [0.37; 0.99] | 2.3% |
| Lopez−Otero, Spain | 13 | 210 | 20 | 755 | 2.43 | [1.19; 4.96] | 1.8% |
| Marcos, Spain | 122 | 298 | 241 | 620 | 1.09 | [0.82; 1.44] | 2.6% |
| Regina, Switzerland | 11 | 51 | 26 | 149 | 1.30 | [0.59; 2.87] | 1.7% |
| Pongpirul, Thailand | 4 | 17 | 28 | 176 | 1.63 | [0.49; 5.35] | 1.1% |
| Selçuk, Turkey | 37 | 74 | 7 | 39 | 4.57 | [1.79; 11.66] | 1.5% |
| Senkal, Turkey | 48 | 165 | 117 | 446 | 1.15 | [0.78; 1.72] | 2.4% |
| Bean, UK | 127 | 399 | 288 | 801 | 0.83 | [0.64; 1.07] | 2.6% |
| Russell, UK | 4 | 33 | 24 | 123 | 0.57 | [0.18; 1.77] | 1.2% |
| Argenziano, USA | 71 | 284 | 165 | 716 | 1.11 | [0.81; 1.53] | 2.5% |
| Bae, USA | 13 | 78 | 32 | 512 | 3.00 | [1.50; 6.01] | 1.9% |
| Castro, USA | 32 | 218 | 181 | 2053 | 1.78 | [1.19; 2.67] | 2.4% |
| Ebinger, USA | 48 | 72 | 166 | 370 | 2.46 | [1.45; 4.18] | 2.2% |
| Ferguson, USA | 7 | 13 | 14 | 59 | 3.75 | [1.08; 13.02] | 1.1% |
| Garibaldi, USA | 55 | 151 | 174 | 596 | 1.39 | [0.95; 2.02] | 2.4% |
| Goshua, USA | 15 | 18 | 33 | 50 | 2.58 | [0.65; 10.14] | 1.0% |
| Jillella, USA | 2 | 3 | 4 | 5 | 0.50 | [0.02; 12.90] | 0.2% |
| Mehta, USA | 22 | 212 | 15 | 1523 | 11.64 | [5.94; 22.83] | 1.9% |
| Palaiodimos, USA | 26 | 62 | 64 | 138 | 0.84 | [0.46; 1.53] | 2.0% |
| Rentsch, USA | 69 | 255 | 53 | 330 | 1.94 | [1.30; 2.90] | 2.4% |
| Reynolds, USA | 336 | 1374 | 666 | 4520 | 1.87 | [1.62; 2.17] | 2.7% |
| Richardson, USA | 87 | 413 | 141 | 953 | 1.54 | [1.14; 2.07] | 2.6% |
| **Random effects model** | | **8895** | | **31027** | **1.45** | **[1.23; 1.72]** | **100.0%** |

Heterogeneity: $I^2 = 81\%$, $\tau^2 = 0.2578$, $p < 0.01$

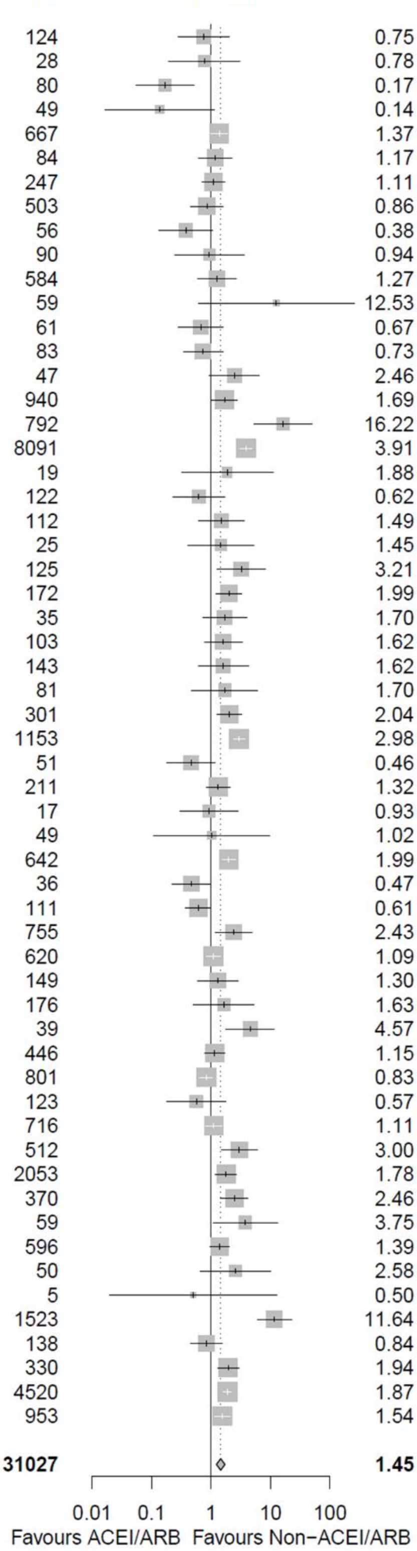


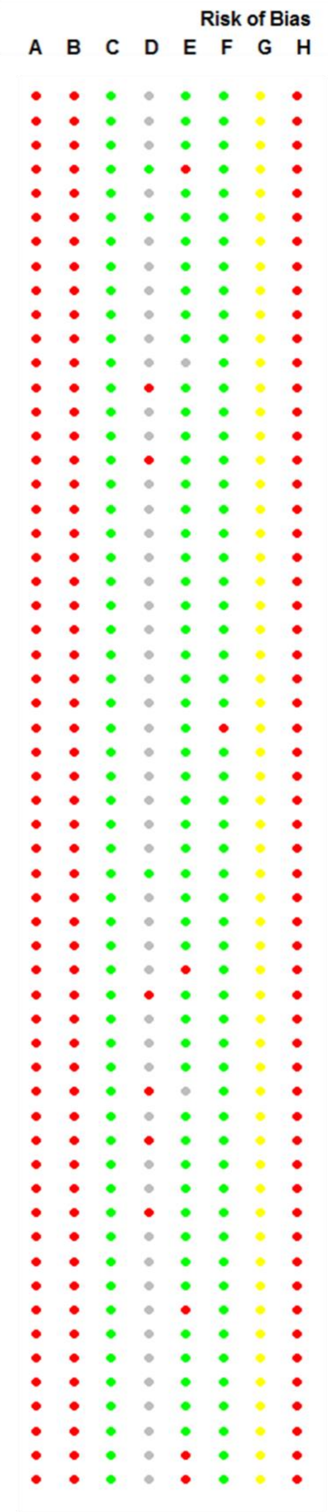


**Figure S22. Continued.**

### F. Only diabetic patients

| Study | ACEI/ARB Events | ACEI/ARB Total | Non-ACEI/ARB Events | Non-ACEI/ARB Total | OR | 95%-CI | Weight |
|---|---|---|---|---|---|---|---|
| Cariou, France | 232 | 737 | 150 | 580 | 1.32 | [1.03; 1.68] | 89.5% |
| Rhee, South Korea | 13 | 327 | 21 | 505 | 0.95 | [0.47; 1.93] | 10.5% |
| **Random effects model** | | **1064** | | **1085** | **1.27** | **[1.01; 1.60]** | **100.0%** |

Heterogeneity: $I^2 = 0\%$, $\tau^2 = 0$, $p = 0.40$

Odds Ratio axis: 0.5, 1, 2 — Favours ACEI/ARB | Favours Non-ACEI/ARB

Risk of Bias: A B C D E F G H

### G. Only in-patients (hospitalized patients)

| Study | ACEI/ARB Events | ACEI/ARB Total | Non-ACEI/ARB Events | Non-ACEI/ARB Total | OR | 95%-CI | Weight |
|---|---|---|---|---|---|---|---|
| Feng Y, China | 4 | 33 | 36 | 80 | 0.17 | [0.05; 0.52] | 1.2% |
| Feng Z, China | 1 | 16 | 16 | 49 | 0.14 | [0.02; 1.13] | 0.4% |
| Gao, China | 74 | 183 | 221 | 667 | 1.37 | [0.98; 1.92] | 3.6% |
| Li J, China | 57 | 115 | 116 | 247 | 1.11 | [0.71; 1.73] | 3.2% |
| Li X, China | 19 | 42 | 247 | 503 | 0.86 | [0.45; 1.61] | 2.4% |
| Liu Y, China | 7 | 22 | 31 | 56 | 0.38 | [0.13; 1.07] | 1.3% |
| Peng, China | 3 | 22 | 13 | 90 | 0.94 | [0.24; 3.61] | 0.9% |
| Xie Yang, China | 10 | 35 | 140 | 584 | 1.27 | [0.59; 2.71] | 2.0% |
| Xie Yangjing, China | 3 | 3 | 21 | 59 | 12.53 | [0.62; 254.25] | 0.2% |
| Xu, China | 11 | 40 | 22 | 61 | 0.67 | [0.28; 1.60] | 1.7% |
| Yang, China | 15 | 43 | 35 | 83 | 0.73 | [0.34; 1.58] | 2.0% |
| Zeng, China | 15 | 28 | 15 | 47 | 2.46 | [0.94; 6.45] | 1.5% |
| Zhang, China | 24 | 188 | 75 | 940 | 1.69 | [1.04; 2.75] | 3.0% |
| Reilev, Denmark | 118 | 685 | 182 | 1405 | 1.40 | [1.09; 1.80] | 3.9% |
| Feuth, Finland | 3 | 9 | 4 | 19 | 1.88 | [0.32; 11.02] | 0.6% |
| Allenbach, France | 6 | 25 | 41 | 122 | 0.62 | [0.23; 1.68] | 1.4% |
| Benotmane, France | 8 | 15 | 11 | 25 | 1.45 | [0.40; 5.26] | 1.0% |
| Dauchet, France | 34 | 56 | 54 | 93 | 1.12 | [0.57; 2.19] | 2.3% |
| Liabeuf, France | 52 | 96 | 64 | 172 | 1.99 | [1.20; 3.31] | 2.9% |
| Meszaros, France | 32 | 68 | 12 | 35 | 1.70 | [0.73; 3.97] | 1.8% |
| Oussalah, France | 26 | 43 | 50 | 103 | 1.62 | [0.79; 3.34] | 2.1% |
| Zhou, Hong Kong (China) | 14 | 184 | 4 | 792 | 16.22 | [5.27; 49.90] | 1.2% |
| Ashraf, Iran | 4 | 19 | 11 | 81 | 1.70 | [0.48; 6.06] | 1.0% |
| Benelli, Italy | 42 | 110 | 70 | 301 | 2.04 | [1.28; 3.26] | 3.0% |
| Parigi, Italy | 57 | 114 | 91 | 211 | 1.32 | [0.83; 2.08] | 3.1% |
| Sardu, Italy | 23 | 45 | 9 | 17 | 0.93 | [0.30; 2.84] | 1.2% |
| Higuchi, Japan | 1 | 8 | 6 | 49 | 1.02 | [0.11; 9.84] | 0.4% |
| Jung, South Korea | 14 | 377 | 22 | 1577 | 2.73 | [1.38; 5.38] | 2.2% |
| de Abajo, Spain | 215 | 497 | 178 | 642 | 1.99 | [1.55; 2.55] | 4.0% |
| Jurado, Spain | 56 | 191 | 45 | 111 | 0.61 | [0.37; 0.99] | 3.0% |
| Lopez−Otero, Spain | 13 | 78 | 20 | 156 | 1.36 | [0.64; 2.90] | 2.0% |
| Marcos, Spain | 122 | 298 | 241 | 620 | 1.09 | [0.82; 1.44] | 3.8% |
| Regina, Switzerland | 11 | 51 | 26 | 149 | 1.30 | [0.59; 2.87] | 1.9% |
| Pongpirul, Thailand | 4 | 17 | 28 | 176 | 1.63 | [0.49; 5.35] | 1.1% |
| Selçuk, Turkey | 37 | 74 | 7 | 39 | 4.57 | [1.79; 11.66] | 1.6% |
| Senkal, Turkey | 48 | 165 | 117 | 446 | 1.15 | [0.78; 1.72] | 3.3% |
| Bean, UK | 127 | 399 | 288 | 801 | 0.83 | [0.64; 1.07] | 3.9% |
| Bae, USA | 13 | 35 | 32 | 87 | 1.02 | [0.45; 2.29] | 1.9% |
| Castro, USA | 32 | 108 | 181 | 599 | 0.97 | [0.62; 1.52] | 3.1% |
| Ebinger, USA | 18 | 48 | 59 | 166 | 1.09 | [0.56; 2.12] | 2.3% |
| Ferguson, USA | 7 | 13 | 14 | 59 | 3.75 | [1.08; 13.02] | 1.0% |
| Garibaldi, USA | 55 | 151 | 174 | 596 | 1.39 | [0.95; 2.02] | 3.4% |
| Goshua, USA | 15 | 18 | 33 | 50 | 2.58 | [0.65; 10.14] | 0.9% |
| Jillella, USA | 2 | 3 | 4 | 5 | 0.50 | [0.02; 12.90] | 0.2% |
| Mehta, USA | 22 | 53 | 15 | 36 | 0.99 | [0.42; 2.35] | 1.7% |
| Palaiodimos, USA | 26 | 62 | 64 | 138 | 0.84 | [0.46; 1.53] | 2.5% |
| Rentsch, USA | 69 | 147 | 53 | 150 | 1.62 | [1.02; 2.58] | 3.1% |
| Richardson, USA | 87 | 413 | 141 | 953 | 1.54 | [1.14; 2.07] | 3.8% |
| **Random effects model** | | **5445** | | **14447** | **1.29** | **[1.12; 1.49]** | **100.0%** |

Heterogeneity: $I^2 = 61\%$, $\tau^2 = 0.1207$, $p < 0.01$

Odds Ratio axis: 0.01, 0.1, 1, 10, 100 — Favours ACEI/ARB | Favours Non−ACEI/ARB

Risk of Bias: A B C D E F G H

**Figure S22. Continued.**

**H. Adjusted estimates**

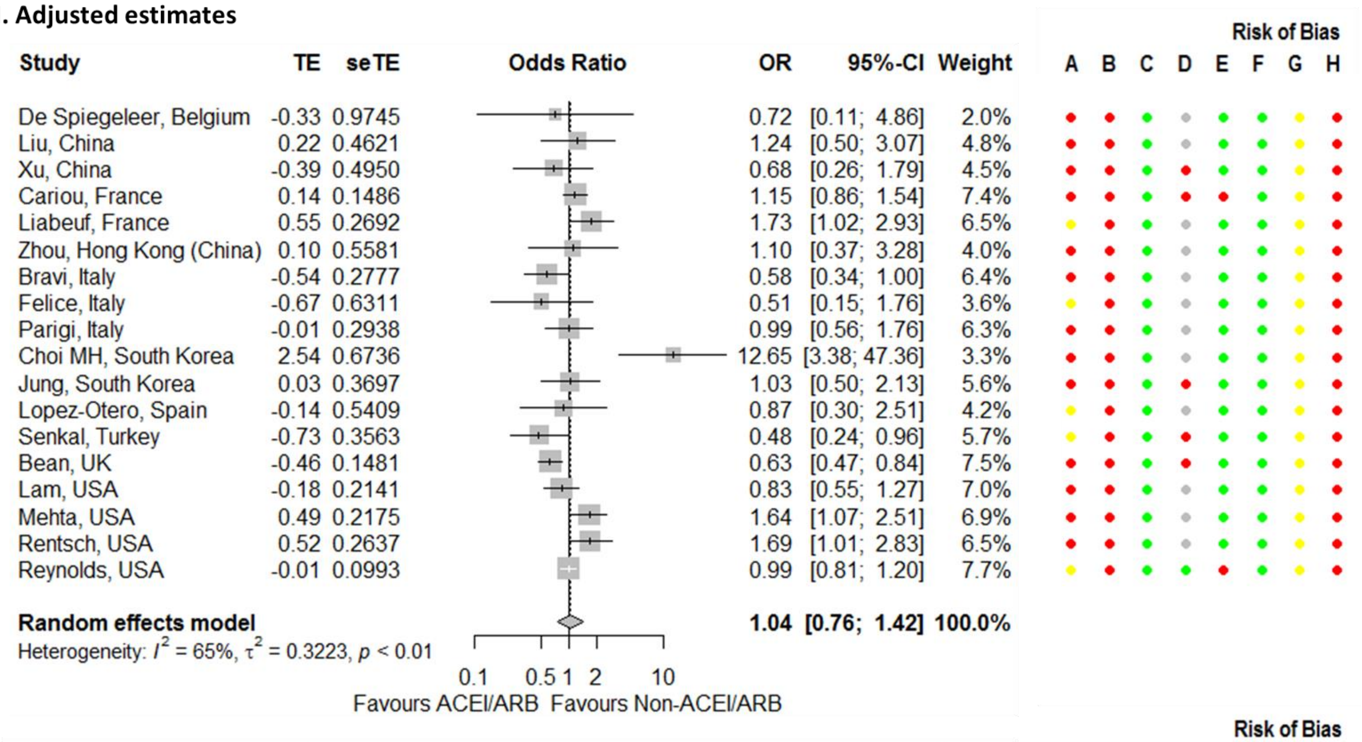


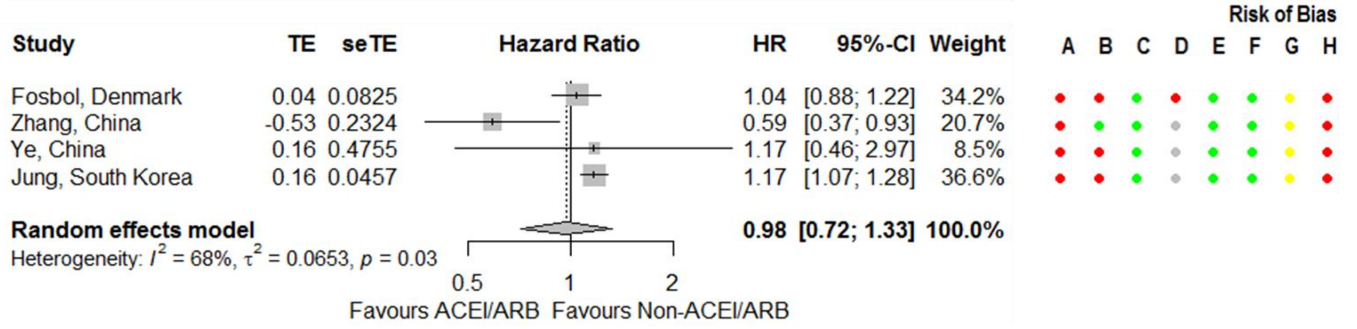


**Figure S22. Continued.**

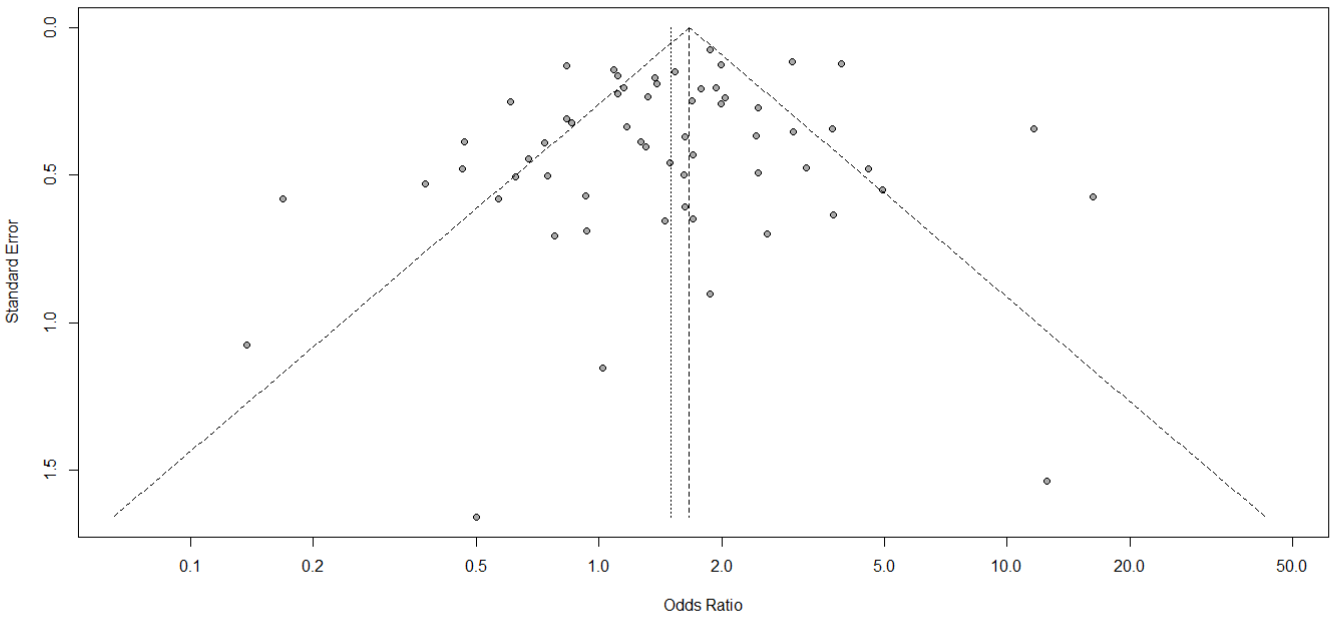


**Figure S23. Funnel plot for the association between severity outcomes in COVID-19 patients and being on an angiotensin-converting enzyme inhibitor or angiotensin receptor blocker.**

**A. Studies from the same cluster all included**

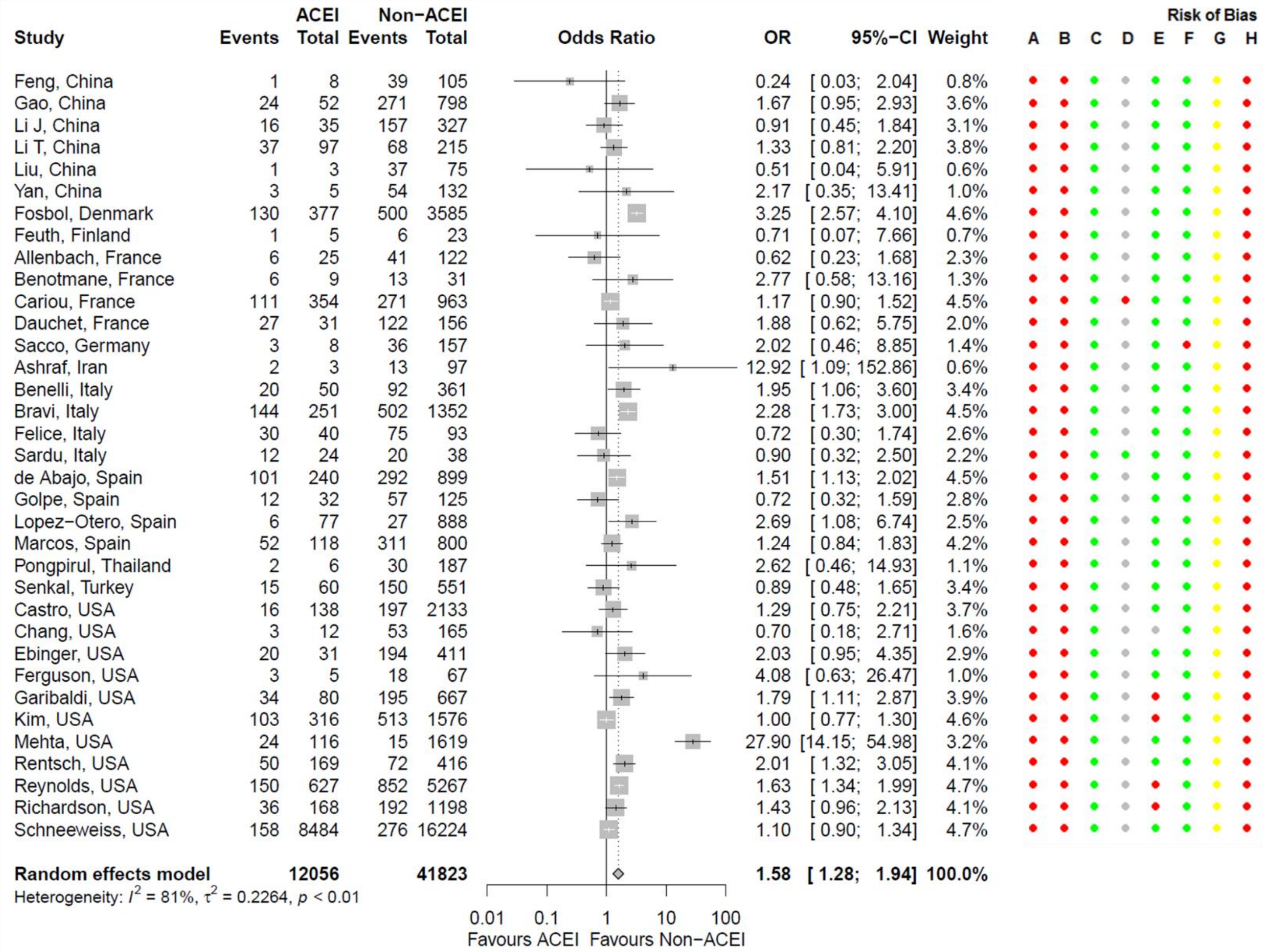


**Figure S24. Forest plot for association between severity outcomes in COVID-19 patients and being on an angiotensin-converting enzyme inhibitor (ACEI).** Risk of bias legend. A = risk of bias due to confounding, B = risk of bias in selection of participants into the study, C = risk of bias in classification of interventions, D = risk of bias due to deviations from intended interventions, E = risk of bias due to missing data, F = risk of bias in measurement of outcomes, G = risk of bias in selection of the reported result, H = overall risk of bias. Color codes. Colour codes. Red = serious, yellow = moderate, green = low, grey = unclear.

### B. Primary meta-analysis

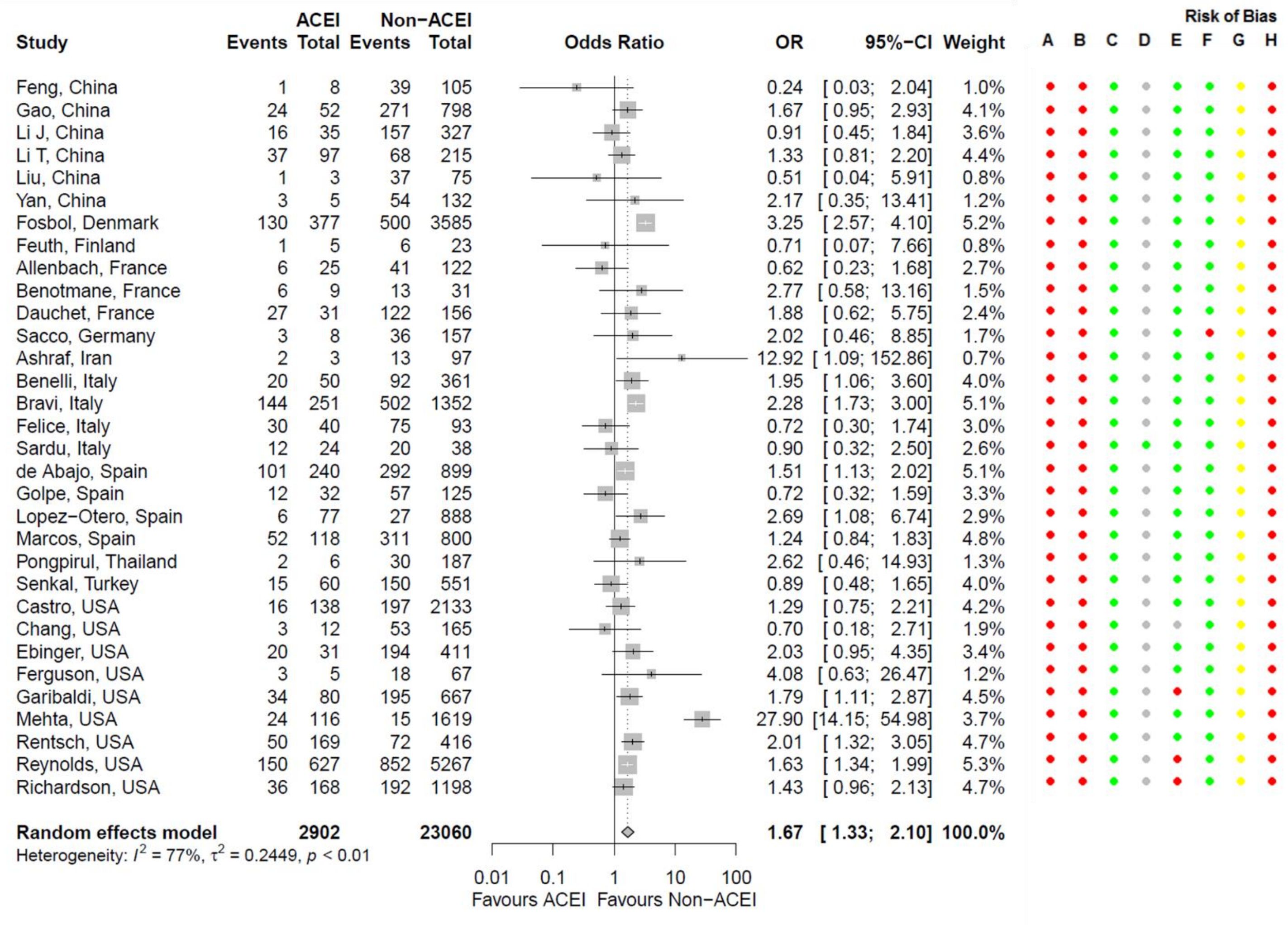


**Figure S24. Continued.**

**C. Only hospitalized patients**

| Study | ACEI Events | ACEI Total | Non-ACEI Events | Non-ACEI Total | OR | 95%-CI | Weight |
|---|---|---|---|---|---|---|---|
| Feng Y, China | 1 | 8 | 39 | 105 | 0.24 | [0.03; 2.04] | 0.6% |
| Gao, China | 24 | 52 | 271 | 798 | 1.67 | [0.95; 2.93] | 6.0% |
| Li J, China | 16 | 35 | 157 | 327 | 0.91 | [0.45; 1.84] | 4.4% |
| Li T, China | 37 | 97 | 68 | 215 | 1.33 | [0.81; 2.20] | 7.1% |
| Liu Y, China | 1 | 3 | 37 | 75 | 0.51 | [0.04; 5.91] | 0.4% |
| Feuth, Finland | 1 | 5 | 6 | 23 | 0.71 | [0.07; 7.66] | 0.5% |
| Allenbach, France | 6 | 25 | 41 | 122 | 0.62 | [0.23; 1.68] | 2.4% |
| Benotmane, France | 6 | 9 | 13 | 31 | 2.77 | [0.58; 13.16] | 1.0% |
| Dauchet, France | 13 | 27 | 75 | 122 | 0.58 | [0.25; 1.35] | 3.2% |
| Ashraf, Iran | 2 | 3 | 13 | 97 | 12.92 | [1.09; 152.86] | 0.4% |
| Benelli, Italy | 20 | 50 | 92 | 361 | 1.95 | [1.06; 3.60] | 5.3% |
| Sardu, Italy | 12 | 24 | 20 | 38 | 0.90 | [0.32; 2.50] | 2.3% |
| de Abajo, Spain | 101 | 240 | 292 | 899 | 1.51 | [1.13; 2.02] | 12.8% |
| Marcos, Spain | 52 | 118 | 311 | 800 | 1.24 | [0.84; 1.83] | 9.7% |
| Pongpirul, Thailand | 2 | 6 | 30 | 187 | 2.62 | [0.46; 14.93] | 0.8% |
| Senkal, Turkey | 15 | 60 | 150 | 551 | 0.89 | [0.48; 1.65] | 5.3% |
| Castro, USA | 16 | 60 | 197 | 647 | 0.83 | [0.46; 1.51] | 5.6% |
| Chang, USA | 3 | 12 | 53 | 165 | 0.70 | [0.18; 2.71] | 1.4% |
| Ebinger, USA | 5 | 20 | 72 | 194 | 0.56 | [0.20; 1.62] | 2.2% |
| Ferguson, USA | 3 | 5 | 18 | 67 | 4.08 | [0.63; 26.47] | 0.7% |
| Garibaldi, USA | 34 | 80 | 195 | 667 | 1.79 | [1.11; 2.87] | 7.7% |
| Mehta, USA | 24 | 54 | 15 | 39 | 1.28 | [0.55; 2.96] | 3.2% |
| Rentsch, USA | 50 | 103 | 72 | 194 | 1.60 | [0.99; 2.59] | 7.5% |
| Richardson, USA | 36 | 168 | 192 | 1198 | 1.43 | [0.96; 2.13] | 9.4% |
| **Random effects model** | | **1264** | | **7922** | **1.27** | **[1.08; 1.50]** | **100.0%** |

Heterogeneity: $I^2 = 22\%$, $\tau^2 = 0.0322$, $p = 0.16$

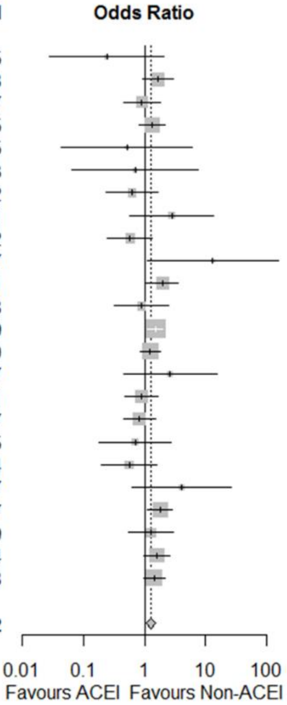


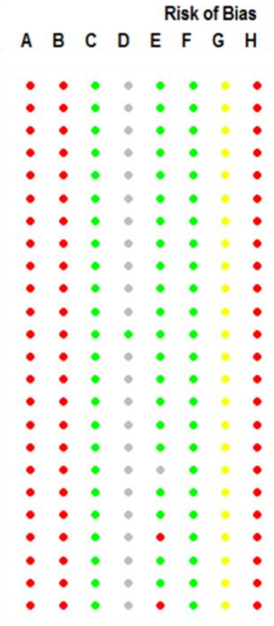


**D. Only hypertensive patients**

| Study | ACEI Events | ACEI Total | Non-ACEI Events | Non-ACEI Total | OR | 95%-CI | Weight |
|---|---|---|---|---|---|---|---|
| Feng Y, China | 1 | 8 | 39 | 105 | 0.24 | [0.03; 2.04] | 1.2% |
| Gao, China | 24 | 52 | 271 | 798 | 1.67 | [0.95; 2.93] | 10.7% |
| Li J, China | 16 | 35 | 157 | 327 | 0.91 | [0.45; 1.84] | 8.1% |
| Liu Y, China | 1 | 3 | 37 | 75 | 0.51 | [0.04; 5.91] | 0.9% |
| Bravi, Italy | 144 | 251 | 192 | 292 | 0.70 | [0.50; 0.99] | 17.3% |
| Felice, Italy | 30 | 40 | 75 | 93 | 0.72 | [0.30; 1.74] | 5.7% |
| Sardu, Italy | 12 | 24 | 20 | 38 | 0.90 | [0.32; 2.50] | 4.5% |
| Golpe, Spain | 12 | 32 | 57 | 125 | 0.72 | [0.32; 1.59] | 6.7% |
| Senkal, Turkey | 15 | 60 | 72 | 188 | 0.54 | [0.28; 1.03] | 8.9% |
| Reynolds, USA | 139 | 584 | 158 | 583 | 0.84 | [0.65; 1.09] | 20.5% |
| Richardson, USA | 36 | 168 | 192 | 1198 | 1.43 | [0.96; 2.13] | 15.5% |
| **Random effects model** | | **1257** | | **3822** | **0.89** | **[0.70; 1.13]** | **100.0%** |

Heterogeneity: $I^2 = 40\%$, $\tau^2 = 0.0530$, $p = 0.08$

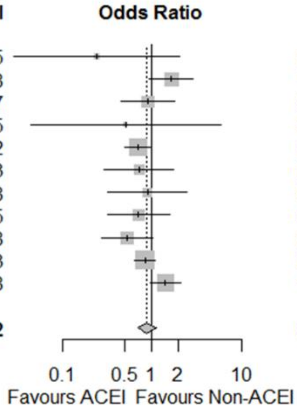


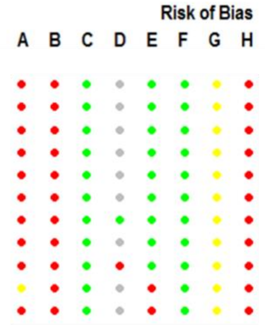


**E. Adjusted estimates**

| Study | TE | seTE | OR | 95%-CI | Weight |
|---|---|---|---|---|---|
| Yan, China | 0.21 | 0.9519 | 1.23 | [0.19; 7.95] | 4.3% |
| Cheung, Hong Kong (China) | -1.97 | 0.9624 | 0.14 | [0.02; 0.92] | 4.2% |
| Bravi, Italy | -0.36 | 0.2406 | 0.70 | [0.44; 1.12] | 14.6% |
| Golpe, Spain | -1.24 | 0.6543 | 0.29 | [0.08; 1.05] | 7.1% |
| Lopez-Otero, Spain | -0.03 | 0.7499 | 0.97 | [0.22; 4.22] | 6.0% |
| Senkal, Turkey | -0.76 | 0.4007 | 0.47 | [0.21; 1.03] | 11.3% |
| Castro, USA | -0.11 | 0.3279 | 0.90 | [0.47; 1.71] | 12.8% |
| Ebinger, USA | -0.87 | 0.5585 | 0.42 | [0.14; 1.25] | 8.5% |
| Mehta, USA | 0.57 | 0.2561 | 1.77 | [1.07; 2.92] | 14.3% |
| Reynolds, USA | -0.11 | 0.1185 | 0.90 | [0.71; 1.14] | 16.7% |
| **Random effects model** | | | **0.72** | **[0.46; 1.13]** | **100.0%** |

Heterogeneity: $I^2 = 53\%$, $\tau^2 = 0.2955$, $p = 0.02$

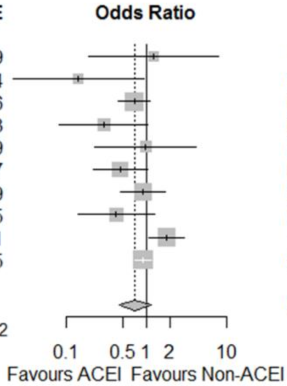


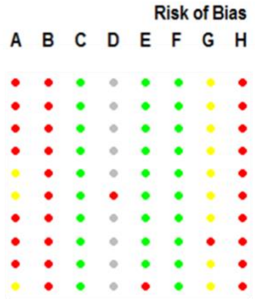


| Study | TE | seTE | HR | 95%-CI | Weight |
|---|---|---|---|---|---|
| Fosbol, Denmark | 0.14 | 0.1315 | 1.15 | [0.89; 1.49] | 14.5% |
| Morales, USA | 0.17 | 0.0540 | 1.18 | [1.06; 1.31] | 85.5% |
| **Random effects model** | | | **1.18** | **[1.07; 1.30]** | **100.0%** |

Heterogeneity: $I^2 = 0\%$, $\tau^2 < 0.0001$, $p = 0.86$

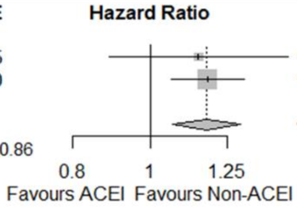


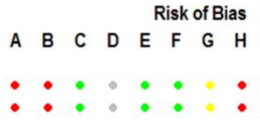


**Figure S24. Continued.**

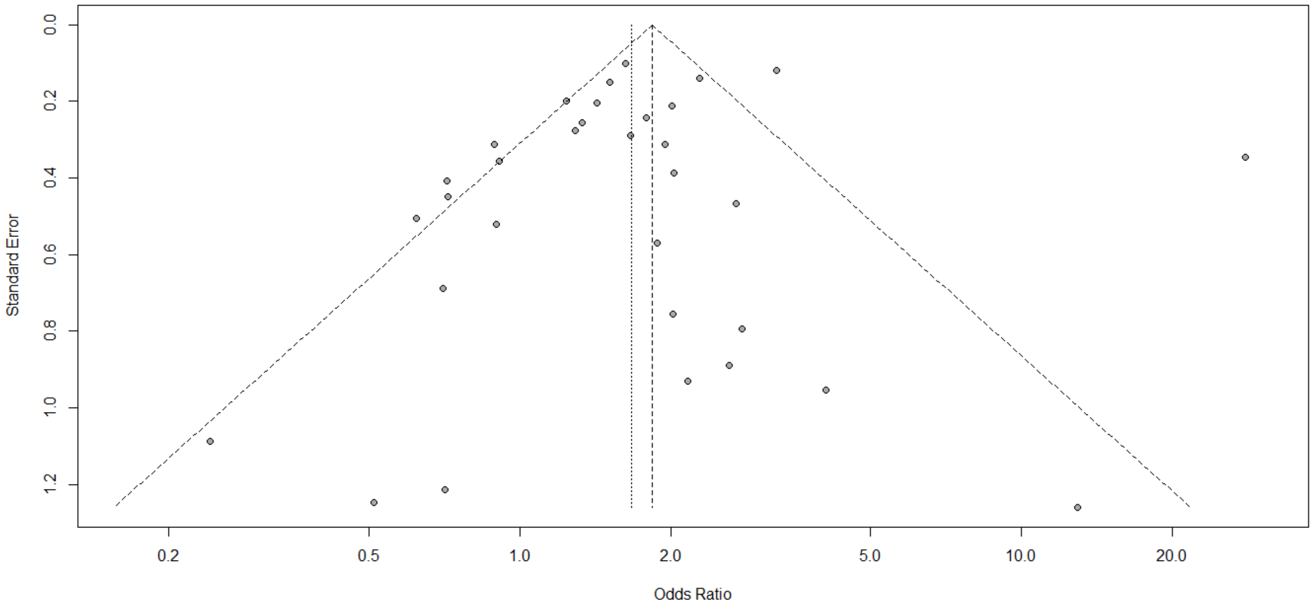


**Figure S25. Funnel plot for the association between severity outcomes in COVID-19 patients and being on an angiotensin-converting enzyme inhibitor.**

**A. Studies from the same cluster all included**

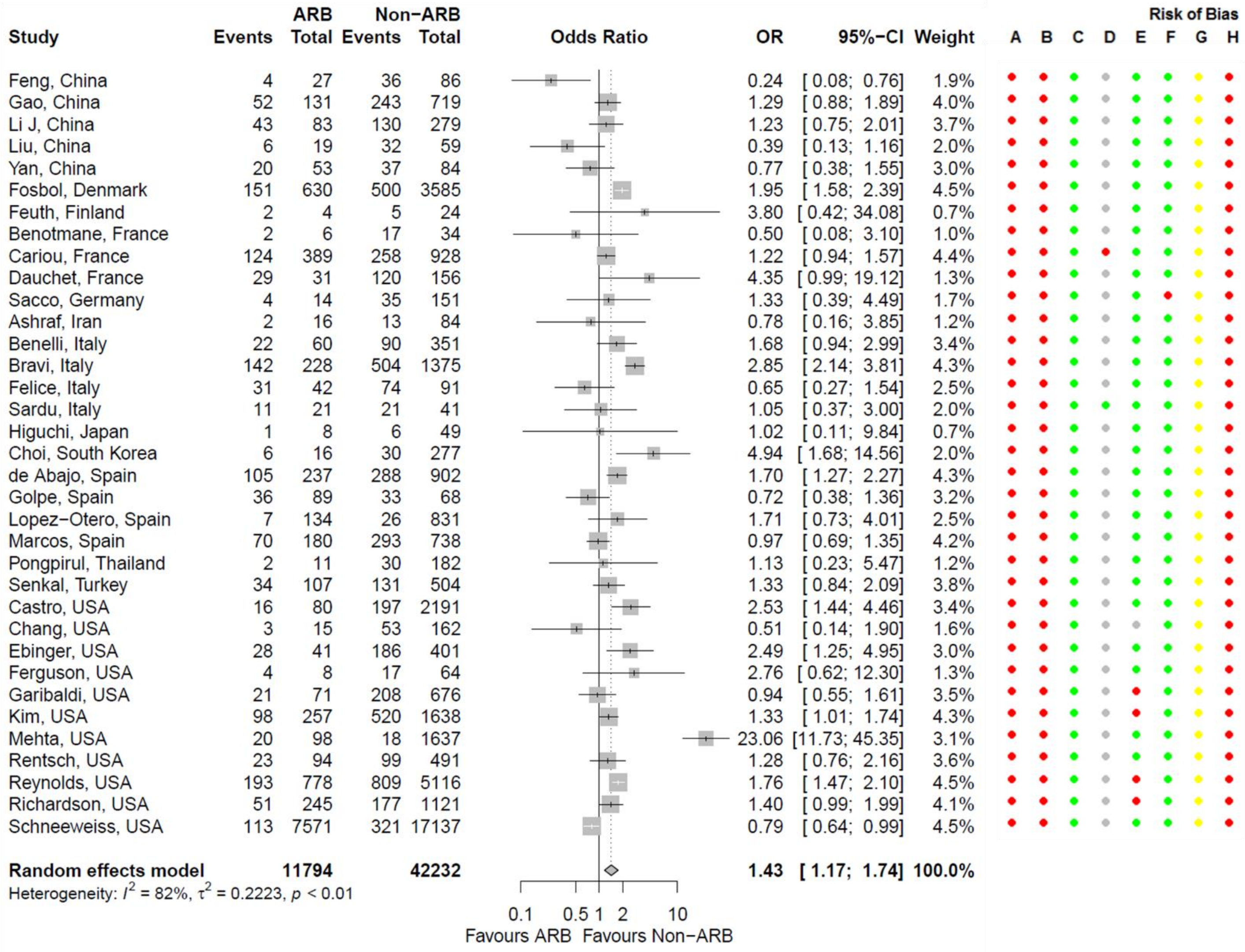


**Figure S26. Forest plot for association between severity outcomes in COVID-19 patients and being on an angiotensin receptor blocker (ARB).** Risk of bias legend. A = risk of bias due to confounding, B = risk of bias in selection of participants into the study, C = risk of bias in classification of interventions, D = risk of bias due to deviations from intended interventions, E = risk of bias due to missing data, F = risk of bias in measurement of outcomes, G = risk of bias in selection of the reported result, H = overall risk of bias. Color codes. Colour codes. Red = serious, yellow = moderate, green = low, grey = unclear.

**B. Primary meta-analysis**

| Study | ARB Events | ARB Total | Non-ARB Events | Non-ARB Total | OR | 95%-CI | Weight |
|---|---|---|---|---|---|---|---|
| Feng, China | 4 | 27 | 36 | 86 | 0.24 | [ 0.08; 0.76] | 2.2% |
| Gao, China | 52 | 131 | 243 | 719 | 1.29 | [ 0.88; 1.89] | 4.6% |
| Li J, China | 43 | 83 | 130 | 279 | 1.23 | [ 0.75; 2.01] | 4.2% |
| Liu, China | 6 | 19 | 32 | 59 | 0.39 | [ 0.13; 1.16] | 2.3% |
| Yan, China | 20 | 53 | 37 | 84 | 0.77 | [ 0.38; 1.55] | 3.4% |
| Fosbol, Denmark | 151 | 630 | 500 | 3585 | 1.95 | [ 1.58; 2.39] | 5.1% |
| Feuth, Finland | 2 | 4 | 5 | 24 | 3.80 | [ 0.42; 34.08] | 0.8% |
| Benotmane, France | 2 | 6 | 17 | 34 | 0.50 | [ 0.08; 3.10] | 1.1% |
| Dauchet, France | 29 | 31 | 120 | 156 | 4.35 | [ 0.99; 19.12] | 1.5% |
| Sacco, Germany | 4 | 14 | 35 | 151 | 1.33 | [ 0.39; 4.49] | 2.0% |
| Ashraf, Iran | 2 | 16 | 13 | 84 | 0.78 | [ 0.16; 3.85] | 1.4% |
| Benelli, Italy | 22 | 60 | 90 | 351 | 1.68 | [ 0.94; 2.99] | 3.9% |
| Bravi, Italy | 142 | 228 | 504 | 1375 | 2.85 | [ 2.14; 3.81] | 4.9% |
| Felice, Italy | 31 | 42 | 74 | 91 | 0.65 | [ 0.27; 1.54] | 2.9% |
| Sardu, Italy | 11 | 21 | 21 | 41 | 1.05 | [ 0.37; 3.00] | 2.4% |
| Higuchi, Japan | 1 | 8 | 6 | 49 | 1.02 | [ 0.11; 9.84] | 0.8% |
| Choi, South Korea | 6 | 16 | 30 | 277 | 4.94 | [ 1.68; 14.56] | 2.3% |
| de Abajo, Spain | 105 | 237 | 288 | 902 | 1.70 | [ 1.27; 2.27] | 4.9% |
| Golpe, Spain | 36 | 89 | 33 | 68 | 0.72 | [ 0.38; 1.36] | 3.7% |
| Lopez−Otero, Spain | 7 | 134 | 26 | 831 | 1.71 | [ 0.73; 4.01] | 2.9% |
| Marcos, Spain | 70 | 180 | 293 | 738 | 0.97 | [ 0.69; 1.35] | 4.7% |
| Pongpirul, Thailand | 2 | 11 | 30 | 182 | 1.13 | [ 0.23; 5.47] | 1.4% |
| Senkal, Turkey | 34 | 107 | 131 | 504 | 1.33 | [ 0.84; 2.09] | 4.3% |
| Castro, USA | 16 | 80 | 197 | 2191 | 2.53 | [ 1.44; 4.46] | 3.9% |
| Chang, USA | 3 | 15 | 53 | 162 | 0.51 | [ 0.14; 1.90] | 1.8% |
| Ebinger, USA | 28 | 41 | 186 | 401 | 2.49 | [ 1.25; 4.95] | 3.5% |
| Ferguson, USA | 4 | 8 | 17 | 64 | 2.76 | [ 0.62; 12.30] | 1.5% |
| Garibaldi, USA | 21 | 71 | 208 | 676 | 0.94 | [ 0.55; 1.61] | 4.0% |
| Mehta, USA | 20 | 98 | 18 | 1637 | 23.06 | [11.73; 45.35] | 3.5% |
| Rentsch, USA | 23 | 94 | 99 | 491 | 1.28 | [ 0.76; 2.16] | 4.1% |
| Reynolds, USA | 193 | 778 | 809 | 5116 | 1.76 | [ 1.47; 2.10] | 5.2% |
| Richardson, USA | 51 | 245 | 177 | 1121 | 1.40 | [ 0.99; 1.99] | 4.7% |
| **Random effects model** | | **3577** | | **22529** | **1.48** | **[ 1.19; 1.85]** | **100.0%** |

Heterogeneity: $I^2 = 79\%$, $\tau^2 = 0.2304$, $p < 0.01$

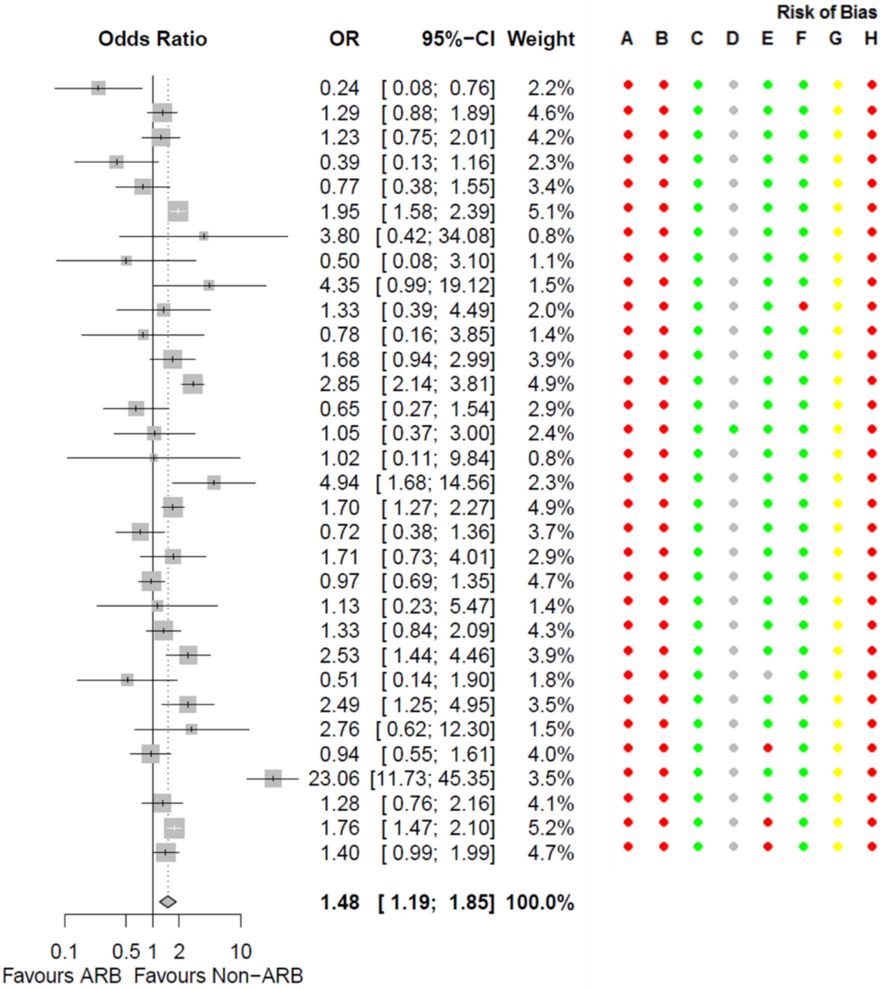


**Figure S26. Continued.**

**C. China-only studies**

| Study | ARB Events | ARB Total | Non-ARB Events | Non-ARB Total | OR | 95%-CI | Weight |
|---|---|---|---|---|---|---|---|
| Feng Y, China | 4 | 27 | 36 | 86 | 0.24 | [0.08; 0.76] | 12.5% |
| Gao, China | 52 | 131 | 243 | 719 | 1.29 | [0.88; 1.89] | 28.2% |
| Li J, China | 43 | 83 | 130 | 279 | 1.23 | [0.75; 2.01] | 25.6% |
| Liu Y, China | 6 | 19 | 32 | 59 | 0.39 | [0.13; 1.16] | 13.2% |
| Yan, China | 20 | 53 | 37 | 84 | 0.77 | [0.38; 1.55] | 20.5% |
| **Random effects model** | | **313** | | **1227** | **0.79** | **[0.48; 1.32]** | **100.0%** |

Heterogeneity: $I^2$ = 66%, $\tau^2$ = 0.2026, $p$ = 0.02

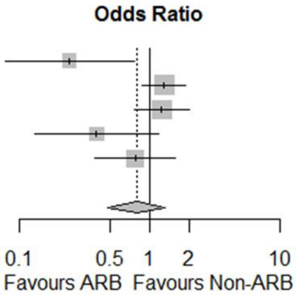


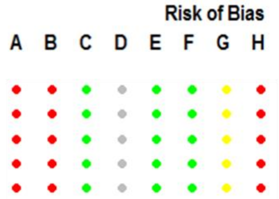


**D. Chinese studies excluded**

| Study | ARB Events | ARB Total | Non-ARB Events | Non-ARB Total | OR | 95%-CI | Weight |
|---|---|---|---|---|---|---|---|
| Fosbol, Denmark | 151 | 630 | 500 | 3585 | 1.95 | [ 1.58; 2.39] | 6.2% |
| Feuth, Finland | 2 | 4 | 5 | 24 | 3.80 | [ 0.42; 34.08] | 1.0% |
| Benotmane, France | 2 | 6 | 17 | 34 | 0.50 | [ 0.08; 3.10] | 1.3% |
| Dauchet, France | 29 | 31 | 120 | 156 | 4.35 | [ 0.99; 19.12] | 1.8% |
| Sacco, Germany | 4 | 14 | 35 | 151 | 1.33 | [ 0.39; 4.49] | 2.3% |
| Ashraf, Iran | 2 | 16 | 13 | 84 | 0.78 | [ 0.16; 3.85] | 1.6% |
| Benelli, Italy | 22 | 60 | 90 | 351 | 1.68 | [ 0.94; 2.99] | 4.7% |
| Bravi, Italy | 142 | 228 | 504 | 1375 | 2.85 | [ 2.14; 3.81] | 6.0% |
| Felice, Italy | 31 | 42 | 74 | 91 | 0.65 | [ 0.27; 1.54] | 3.4% |
| Sardu, Italy | 11 | 21 | 21 | 41 | 1.05 | [ 0.37; 3.00] | 2.8% |
| Higuchi, Japan | 1 | 8 | 6 | 49 | 1.02 | [ 0.11; 9.84] | 0.9% |
| Choi MH, South Korea | 6 | 16 | 30 | 277 | 4.94 | [ 1.68; 14.56] | 2.7% |
| de Abajo, Spain | 105 | 237 | 288 | 902 | 1.70 | [ 1.27; 2.27] | 6.0% |
| Golpe, Spain | 36 | 89 | 33 | 68 | 0.72 | [ 0.38; 1.36] | 4.4% |
| Lopez-Otero, Spain | 7 | 134 | 26 | 831 | 1.71 | [ 0.73; 4.01] | 3.5% |
| Marcos, Spain | 70 | 180 | 293 | 738 | 0.97 | [ 0.69; 1.35] | 5.8% |
| Pongpirul, Thailand | 2 | 11 | 30 | 182 | 1.13 | [ 0.23; 5.47] | 1.6% |
| Senkal, Turkey | 34 | 107 | 131 | 504 | 1.33 | [ 0.84; 2.09] | 5.3% |
| Castro, USA | 16 | 80 | 197 | 2191 | 2.53 | [ 1.44; 4.46] | 4.7% |
| Chang, USA | 3 | 15 | 53 | 162 | 0.51 | [ 0.14; 1.90] | 2.1% |
| Ebinger, USA | 28 | 41 | 186 | 401 | 2.49 | [ 1.25; 4.95] | 4.2% |
| Ferguson, USA | 4 | 8 | 17 | 64 | 2.76 | [ 0.62; 12.30] | 1.8% |
| Garibaldi, USA | 21 | 71 | 208 | 676 | 0.94 | [ 0.55; 1.61] | 4.9% |
| Mehta, USA | 20 | 98 | 18 | 1637 | 23.06 | [11.73; 45.35] | 4.2% |
| Rentsch, USA | 23 | 94 | 99 | 491 | 1.28 | [ 0.76; 2.16] | 4.9% |
| Reynolds, USA | 193 | 778 | 809 | 5116 | 1.76 | [ 1.47; 2.10] | 6.3% |
| Richardson, USA | 51 | 245 | 177 | 1121 | 1.40 | [ 0.99; 1.99] | 5.7% |
| **Random effects model** | | **3264** | | **21302** | **1.69** | **[ 1.34; 2.13]** | **100.0%** |

Heterogeneity: $I^2$ = 78%, $\tau^2$ = 0.2130, $p$ < 0.01

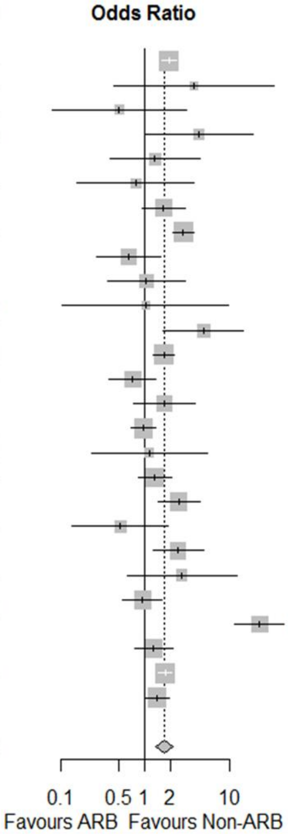


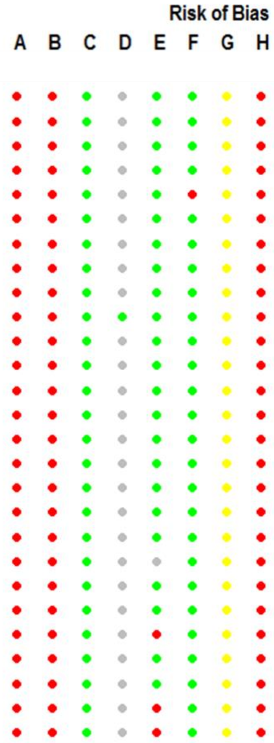


**E. USA-only studies**

| Study | ARB Events | ARB Total | Non-ARB Events | Non-ARB Total | OR | 95%-CI | Weight |
|---|---|---|---|---|---|---|---|
| Castro, USA | 16 | 80 | 197 | 2191 | 2.53 | [ 1.44; 4.46] | 12.0% |
| Chang, USA | 3 | 15 | 53 | 162 | 0.51 | [ 0.14; 1.90] | 7.2% |
| Ebinger, USA | 28 | 41 | 186 | 401 | 2.49 | [ 1.25; 4.95] | 11.2% |
| Ferguson, USA | 4 | 8 | 17 | 64 | 2.76 | [ 0.62; 12.30] | 6.3% |
| Garibaldi, USA | 21 | 71 | 208 | 676 | 0.94 | [ 0.55; 1.61] | 12.3% |
| Mehta, USA | 20 | 98 | 18 | 1637 | 23.06 | [11.73; 45.35] | 11.3% |
| Rentsch, USA | 23 | 94 | 99 | 491 | 1.28 | [ 0.76; 2.16] | 12.4% |
| Reynolds, USA | 193 | 778 | 809 | 5116 | 1.76 | [ 1.47; 2.10] | 14.0% |
| Richardson, USA | 51 | 245 | 177 | 1121 | 1.40 | [ 0.99; 1.99] | 13.3% |
| **Random effects model** | | **1430** | | **11859** | **2.08** | **[ 1.26; 3.43]** | **100.0%** |

Heterogeneity: $I^2$ = 89%, $\tau^2$ = 0.4561, $p$ < 0.01

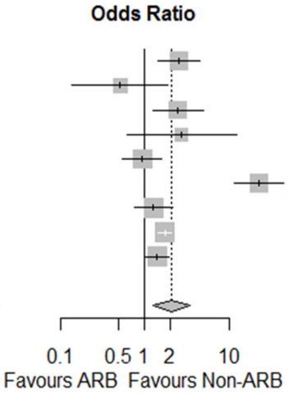


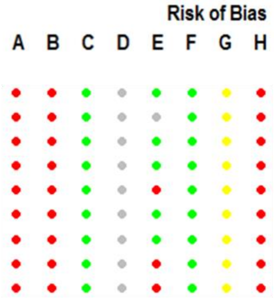


**Figure S26. Continued.**

**F. USA studies excluded**

| Study | ARB Events | ARB Total | Non-ARB Events | Non-ARB Total | OR | 95%-CI | Weight |
|---|---|---|---|---|---|---|---|
| Feng Y, China | 4 | 27 | 36 | 86 | 0.24 | [0.08; 0.76] | 2.9% |
| Gao, China | 52 | 131 | 243 | 719 | 1.29 | [0.88; 1.89] | 7.2% |
| Li J, China | 43 | 83 | 130 | 279 | 1.23 | [0.75; 2.01] | 6.4% |
| Liu Y, China | 6 | 19 | 32 | 59 | 0.39 | [0.13; 1.16] | 3.1% |
| Yan, China | 20 | 53 | 37 | 84 | 0.77 | [0.38; 1.55] | 5.0% |
| Fosbol, Denmark | 151 | 630 | 500 | 3585 | 1.95 | [1.58; 2.39] | 8.2% |
| Feuth, Finland | 2 | 4 | 5 | 24 | 3.80 | [0.42; 34.08] | 1.0% |
| Benotmane, France | 2 | 6 | 17 | 34 | 0.50 | [0.08; 3.10] | 1.4% |
| Dauchet, France | 29 | 31 | 120 | 156 | 4.35 | [0.99; 19.12] | 2.0% |
| Sacco, Germany | 4 | 14 | 35 | 151 | 1.33 | [0.39; 4.49] | 2.7% |
| Ashraf, Iran | 2 | 16 | 13 | 84 | 0.78 | [0.16; 3.85] | 1.8% |
| Benelli, Italy | 22 | 60 | 90 | 351 | 1.68 | [0.94; 2.99] | 5.8% |
| Bravi, Italy | 142 | 228 | 504 | 1375 | 2.85 | [2.14; 3.81] | 7.8% |
| Felice, Italy | 31 | 42 | 74 | 91 | 0.65 | [0.27; 1.54] | 4.1% |
| Sardu, Italy | 11 | 21 | 21 | 41 | 1.05 | [0.37; 3.00] | 3.2% |
| Higuchi, Japan | 1 | 8 | 6 | 49 | 1.02 | [0.11; 9.84] | 1.0% |
| Choi MH, South Korea | 6 | 16 | 30 | 277 | 4.94 | [1.68; 14.56] | 3.1% |
| de Abajo, Spain | 105 | 237 | 288 | 902 | 1.70 | [1.27; 2.27] | 7.8% |
| Golpe, Spain | 36 | 89 | 33 | 68 | 0.72 | [0.38; 1.36] | 5.4% |
| Lopez-Otero, Spain | 7 | 134 | 26 | 831 | 1.71 | [0.73; 4.01] | 4.1% |
| Marcos, Spain | 70 | 180 | 293 | 738 | 0.97 | [0.69; 1.35] | 7.5% |
| Pongpirul, Thailand | 2 | 11 | 30 | 182 | 1.13 | [0.23; 5.47] | 1.8% |
| Senkal, Turkey | 34 | 107 | 131 | 504 | 1.33 | [0.84; 2.09] | 6.7% |
| **Random effects model** | | **2147** | | **10670** | **1.28** | **[1.01; 1.63]** | **100.0%** |

Heterogeneity: $I^2 = 69\%$, $\tau^2 = 0.1687$, $p < 0.01$

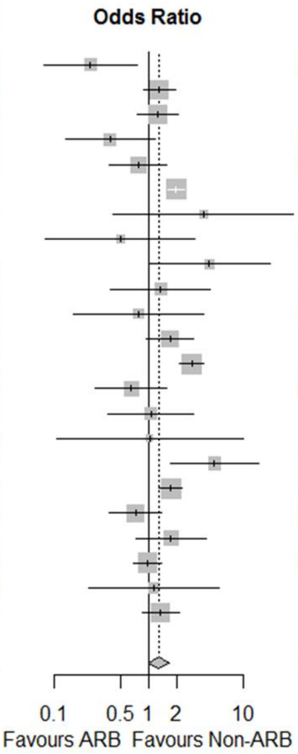


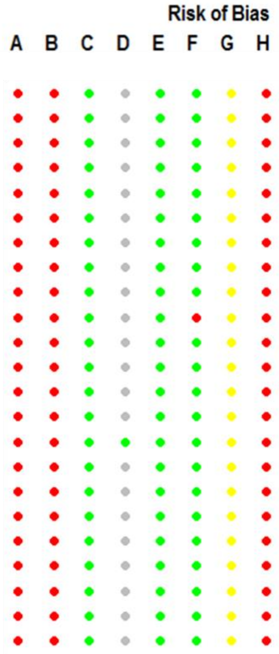


**G. Only hospitalized patients**

| Study | ARB Events | ARB Total | Non-ARB Events | Non-ARB Total | OR | 95%-CI | Weight |
|---|---|---|---|---|---|---|---|
| Feng Y, China | 4 | 27 | 36 | 86 | 0.24 | [0.08; 0.76] | 1.9% |
| Gao, China | 52 | 131 | 243 | 719 | 1.29 | [0.88; 1.89] | 8.4% |
| Li J, China | 43 | 83 | 130 | 279 | 1.23 | [0.75; 2.01] | 6.6% |
| Liu Y, China | 6 | 19 | 32 | 59 | 0.39 | [0.13; 1.16] | 2.1% |
| Feuth, Finland | 2 | 4 | 5 | 24 | 3.80 | [0.42; 34.08] | 0.6% |
| Benotmane, France | 2 | 6 | 17 | 34 | 0.50 | [0.08; 3.10] | 0.8% |
| Dauchet, France | 21 | 29 | 67 | 120 | 2.08 | [0.85; 5.06] | 2.9% |
| Ashraf, Iran | 2 | 16 | 13 | 84 | 0.78 | [0.16; 3.85] | 1.1% |
| Benelli, Italy | 22 | 60 | 90 | 351 | 1.68 | [0.94; 2.99] | 5.5% |
| Sardu, Italy | 11 | 21 | 21 | 41 | 1.05 | [0.37; 3.00] | 2.2% |
| Higuchi, Japan | 1 | 8 | 6 | 49 | 1.02 | [0.11; 9.84] | 0.5% |
| Choi MH, South Korea | 6 | 16 | 30 | 277 | 4.94 | [1.68; 14.56] | 2.1% |
| de Abajo, Spain | 105 | 237 | 288 | 902 | 1.70 | [1.27; 2.27] | 10.3% |
| Lopez-Otero, Spain | 7 | 50 | 26 | 184 | 0.99 | [0.40; 2.43] | 2.9% |
| Marcos, Spain | 70 | 180 | 293 | 738 | 0.97 | [0.69; 1.35] | 9.4% |
| Pongpirul, Thailand | 2 | 11 | 30 | 182 | 1.13 | [0.23; 5.47] | 1.1% |
| Senkal, Turkey | 34 | 107 | 131 | 504 | 1.33 | [0.84; 2.09] | 7.2% |
| Castro, USA | 16 | 48 | 197 | 659 | 1.17 | [0.63; 2.19] | 4.9% |
| Chang, USA | 3 | 15 | 53 | 162 | 0.51 | [0.14; 1.90] | 1.5% |
| Ebinger, USA | 13 | 28 | 64 | 186 | 1.65 | [0.74; 3.68] | 3.4% |
| Ferguson, USA | 4 | 8 | 17 | 64 | 2.76 | [0.62; 12.30] | 1.2% |
| Garibaldi, USA | 21 | 71 | 208 | 676 | 0.94 | [0.55; 1.61] | 6.0% |
| Mehta, USA | 20 | 53 | 18 | 41 | 0.77 | [0.34; 1.78] | 3.3% |
| Rentsch, USA | 23 | 49 | 99 | 248 | 1.33 | [0.72; 2.46] | 5.0% |
| Richardson, USA | 51 | 245 | 177 | 1121 | 1.40 | [0.99; 1.99] | 9.1% |
| **Random effects model** | | **1522** | | **7790** | **1.23** | **[1.04; 1.46]** | **100.0%** |

Heterogeneity: $I^2 = 34\%$, $\tau^2 = 0.0511$, $p = 0.05$

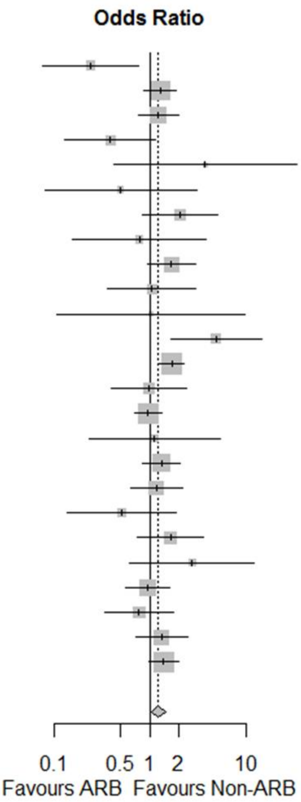


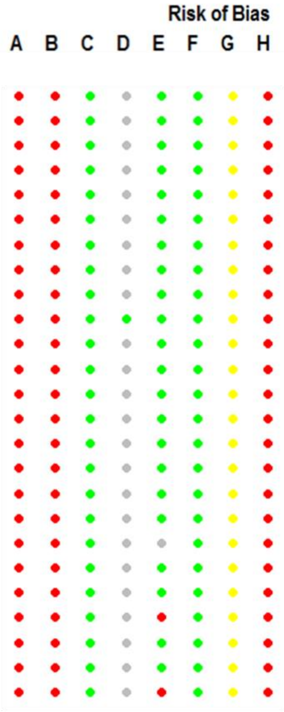


**Figure S26. Continued.**

**H. Only hypertensive patients**

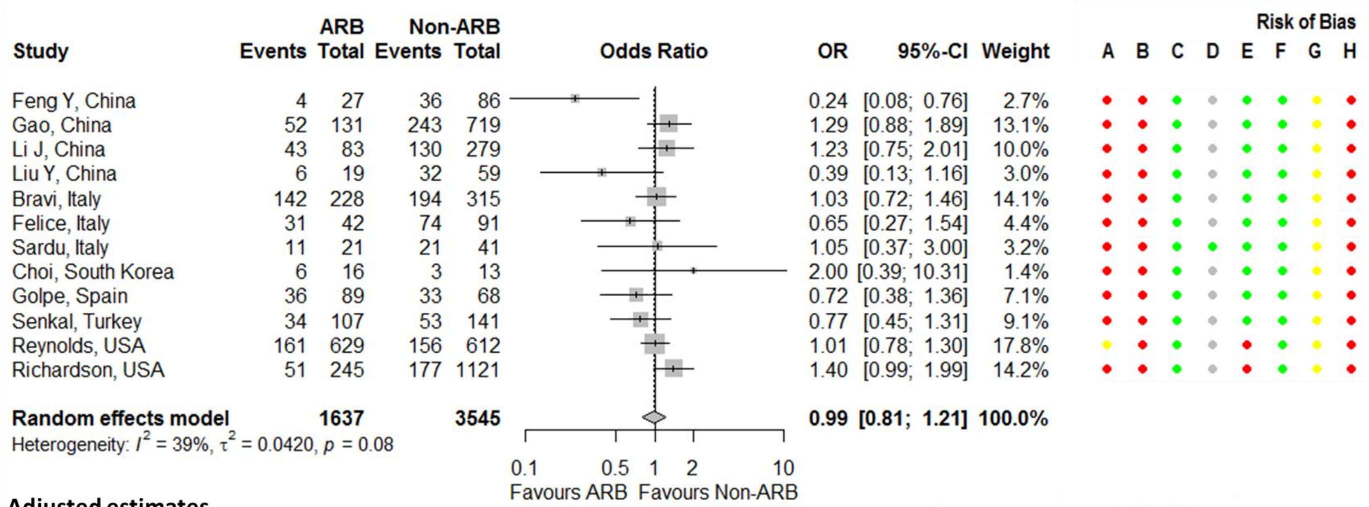


**I. Adjusted estimates**

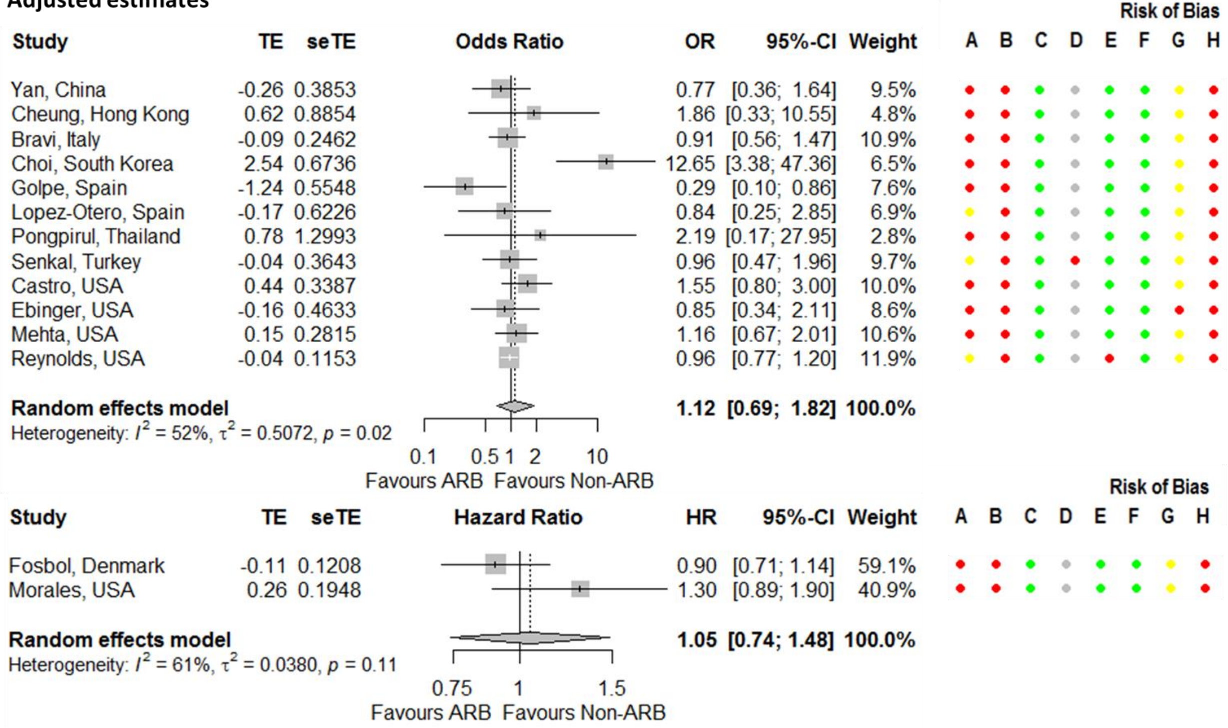


**Figure S26. Continued.**

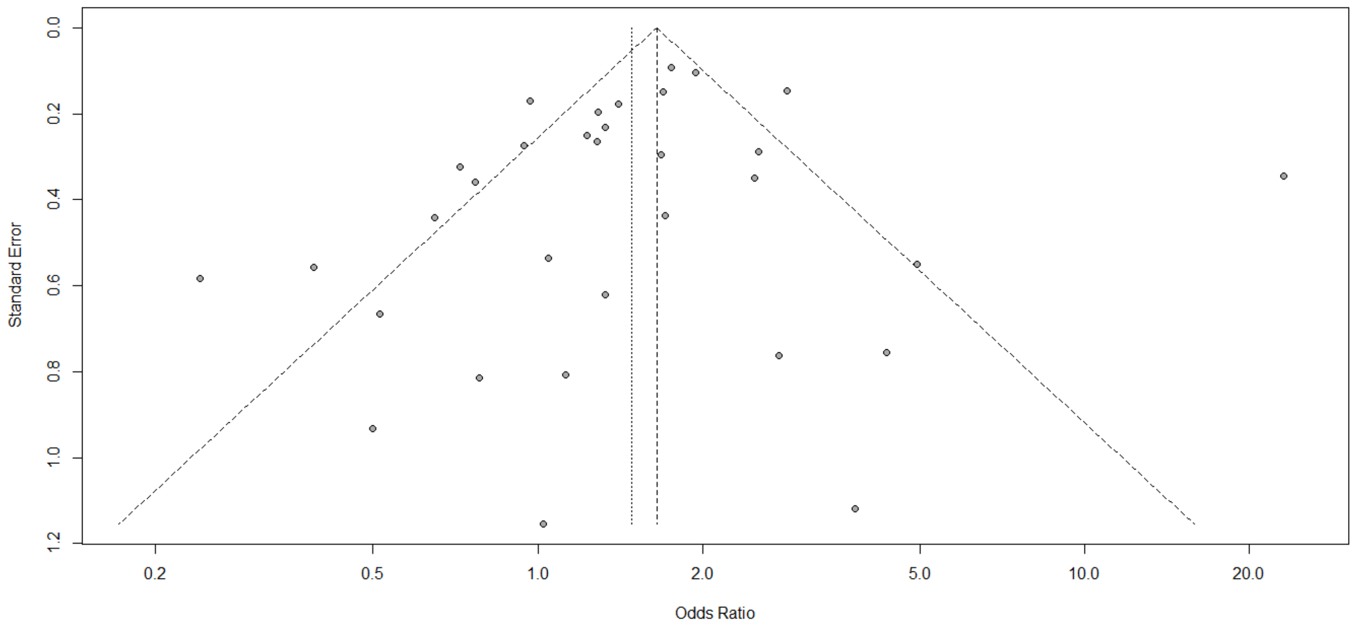


**Figure S27. Funnel plot for the association between severity outcomes in COVID-19 patients and being on an angiotensin receptor blocker.**

**A. Primary meta-analysis**

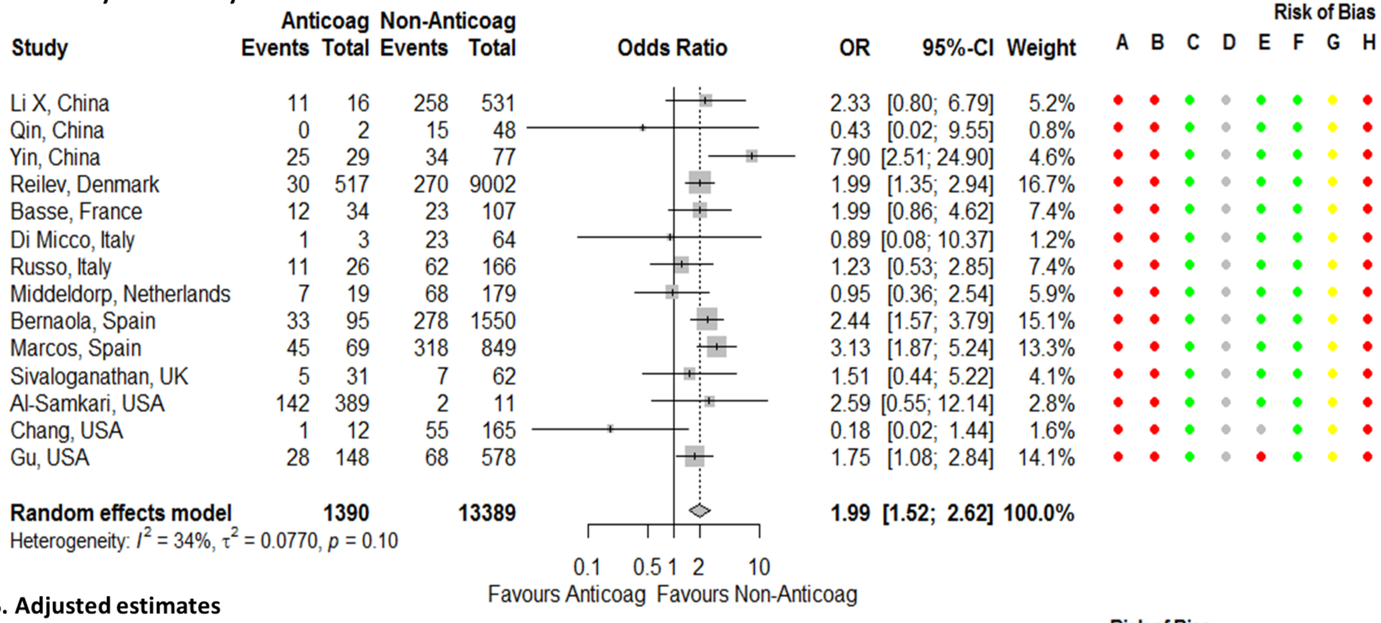


**B. Adjusted estimates**

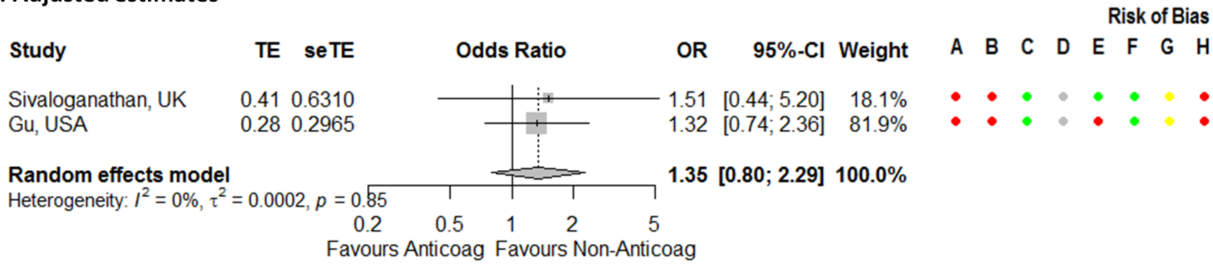


**Figure S28. Forest plot for association between severity outcomes in COVID-19 patients and being on an anticoagulant.** Risk of bias legend. A = risk of bias due to confounding, B = risk of bias in selection of participants into the study, C = risk of bias in classification of interventions, D = risk of bias due to deviations from intended interventions, E = risk of bias due to missing data, F = risk of bias in measurement of outcomes, G = risk of bias in selection of the reported result, H = overall risk of bias. Color codes. Colour codes. Red = serious, yellow = moderate, green = low, grey = unclear.

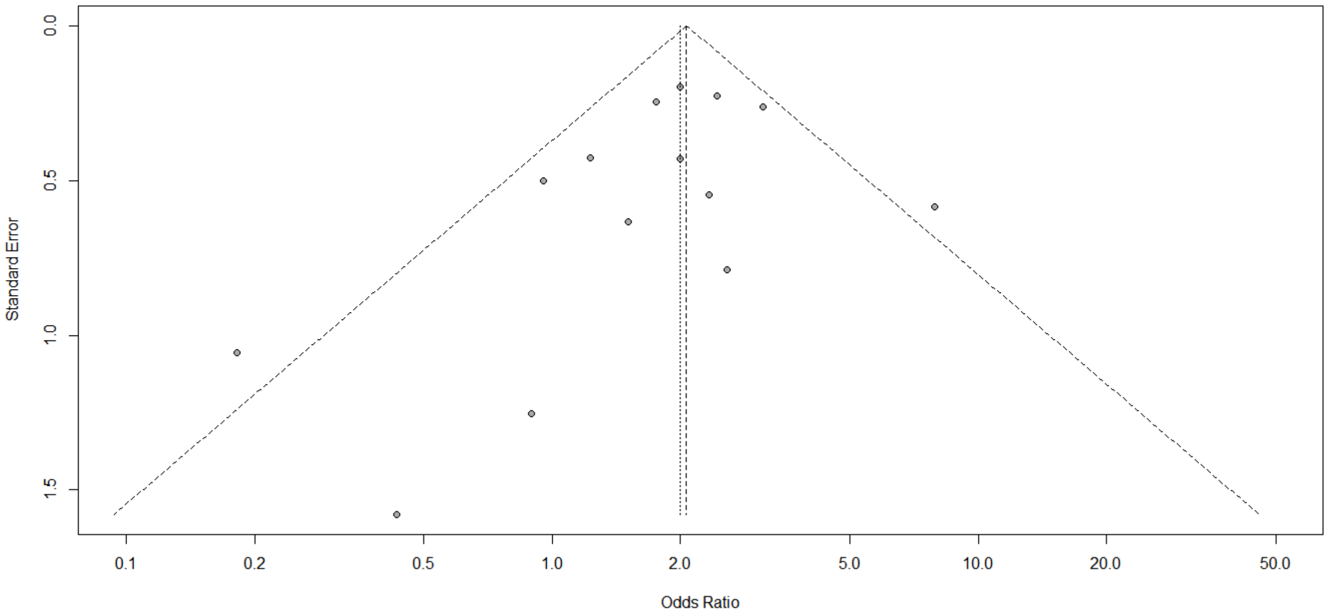


**Figure S29. Funnel plot for association between severity outcomes in COVID-19 patients and being on an anticoagulant.**

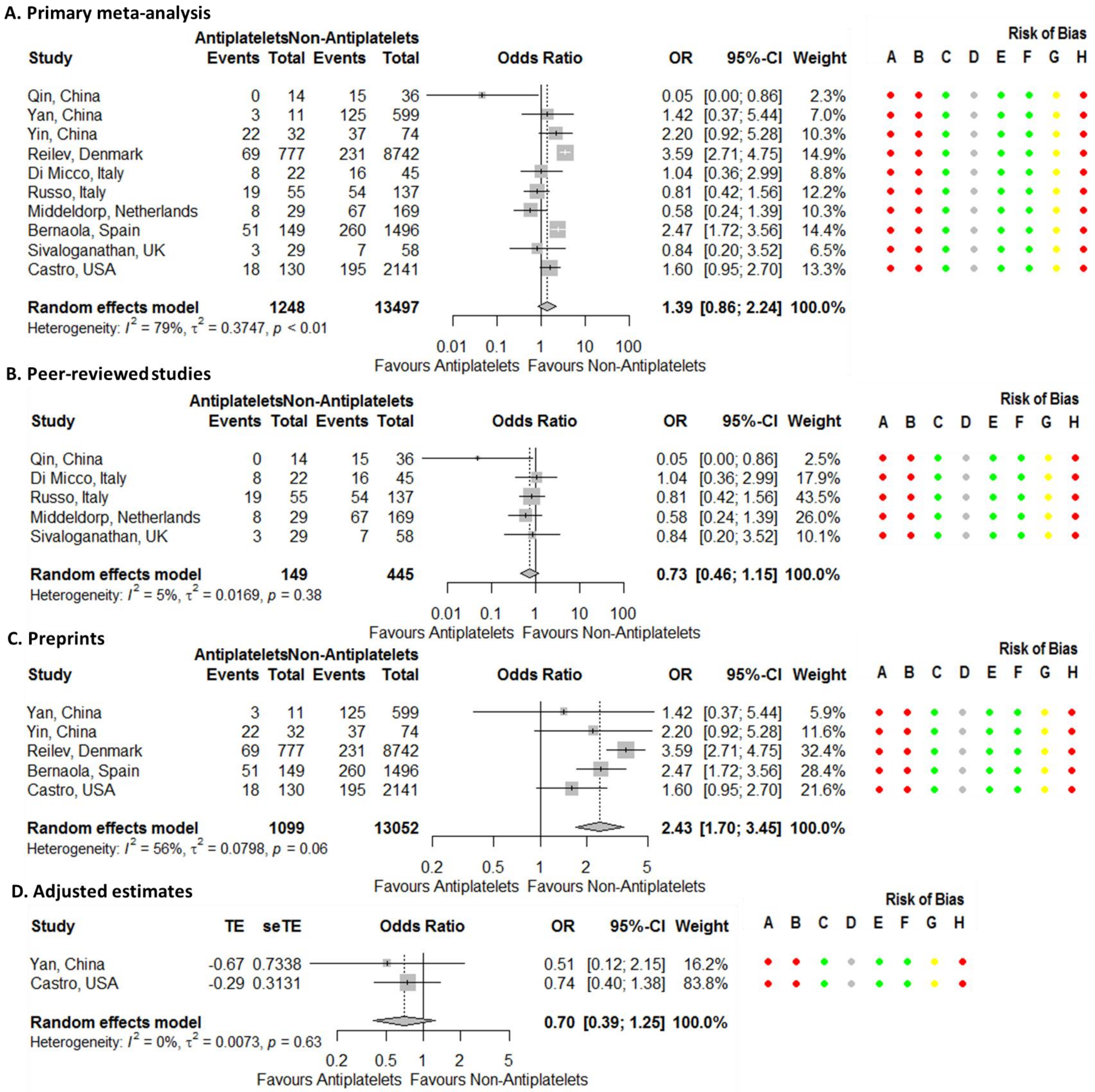


**Figure S30. Forest plots for association between severity outcomes in COVID-19 patients and being on an antiplatelet.** Risk of bias legend. A = risk of bias due to confounding, B = risk of bias in selection of participants into the study, C = risk of bias in classification of interventions, D = risk of bias due to deviations from intended interventions, E = risk of bias due to missing data, F = risk of bias in measurement of outcomes, G = risk of bias in selection of the reported result, H = overall risk of bias. Color codes. Colour codes. Red = serious, yellow = moderate, green = low, grey = unclear.

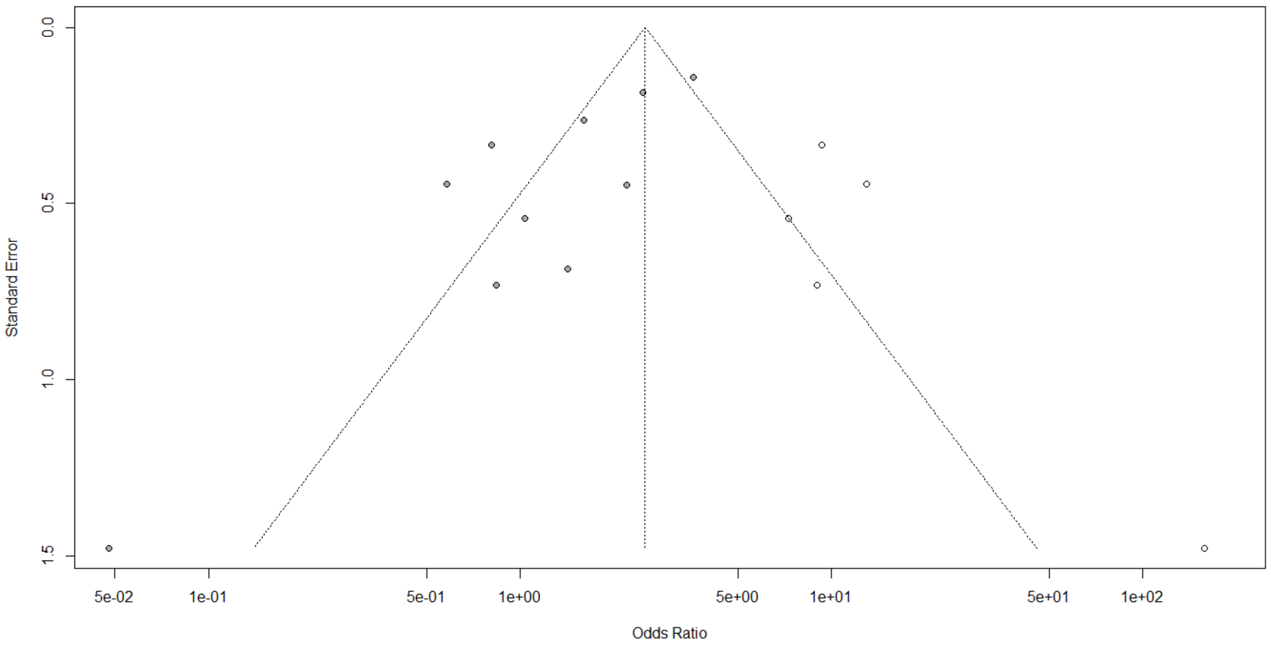


**Figure S31. Funnel plot for association between severity outcomes in COVID-19 patients and being on an antiplatelet.** Solid dots represent the 10 studies included in the primary meta-analysis whereas the open dots, the 5 'filled' studies.

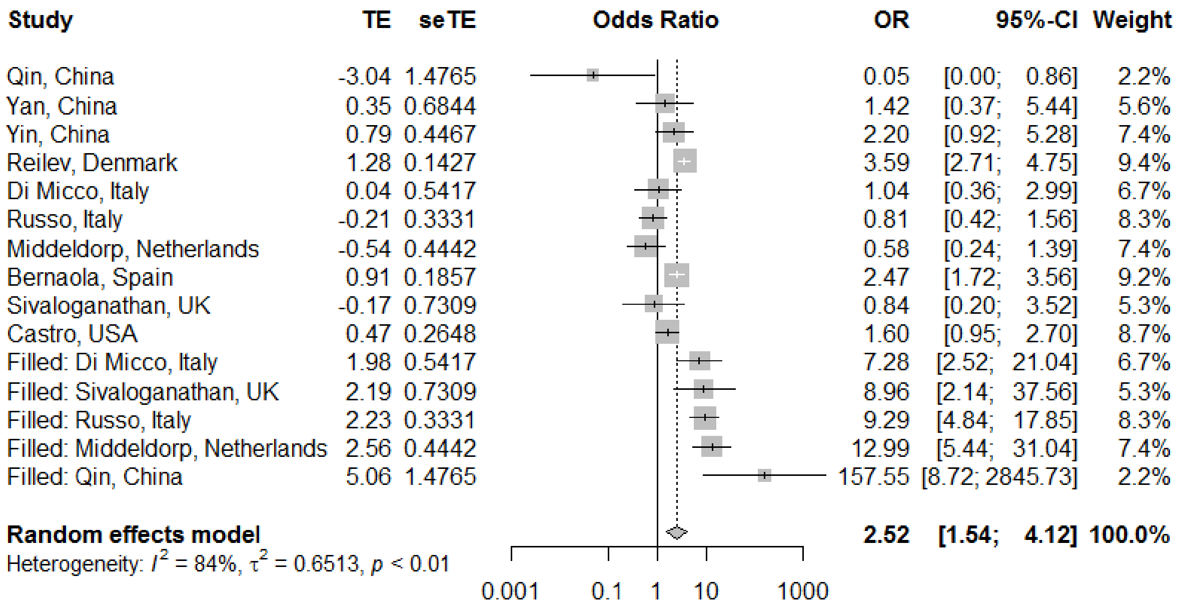


**Figure S32. Forest plot for the association between severity outcomes in COVID-19 patients and being an antiplatelet after adjusting for publication bias.**

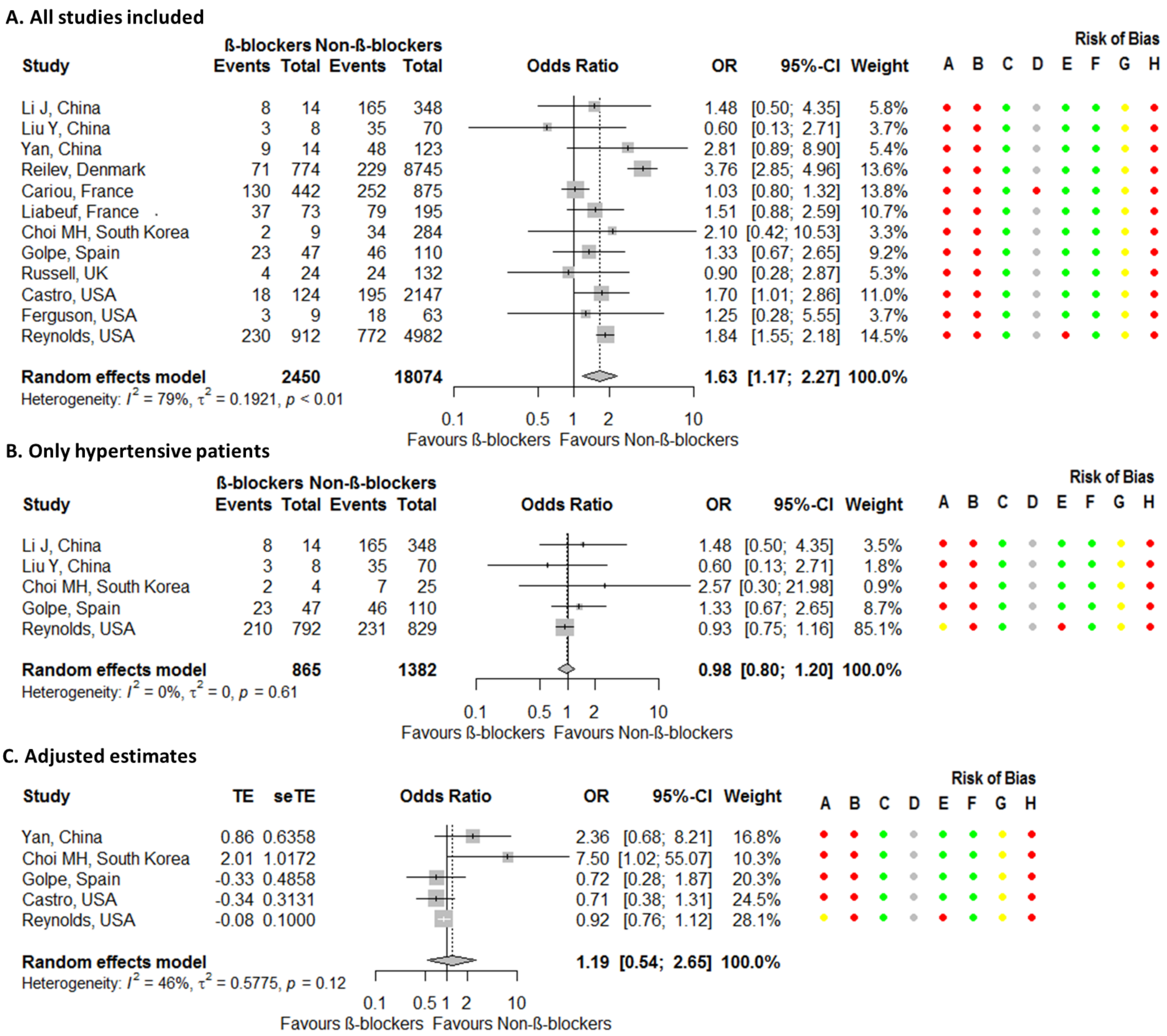


**Figure S33. Forest plots for association between severity outcomes in COVID-19 patients and being on a beta-blocker.** Risk of bias legend. A = risk of bias due to confounding, B = risk of bias in selection of participants into the study, C = risk of bias in classification of interventions, D = risk of bias due to deviations from intended interventions, E = risk of bias due to missing data, F = risk of bias in measurement of outcomes, G = risk of bias in selection of the reported result, H = overall risk of bias. Color codes. Colour codes. Red = serious, yellow = moderate, green = low, grey = unclear.

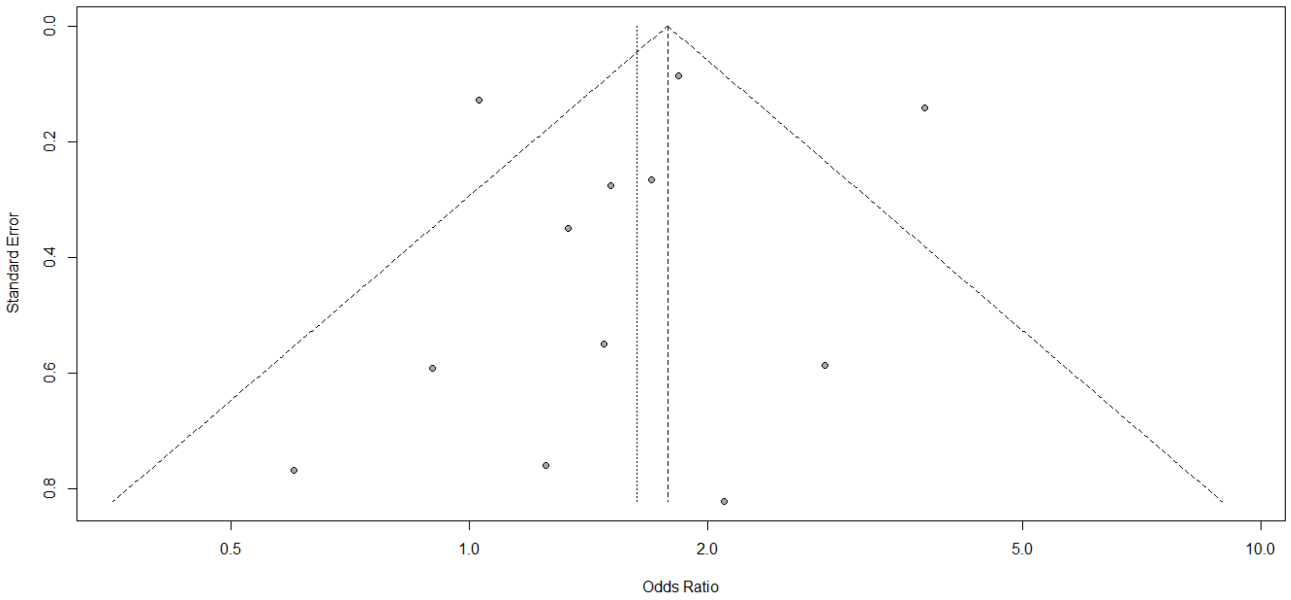


**Figure S34. Funnel plot for association between severity outcomes in COVID-19 patients and being on a beta-blocker.**

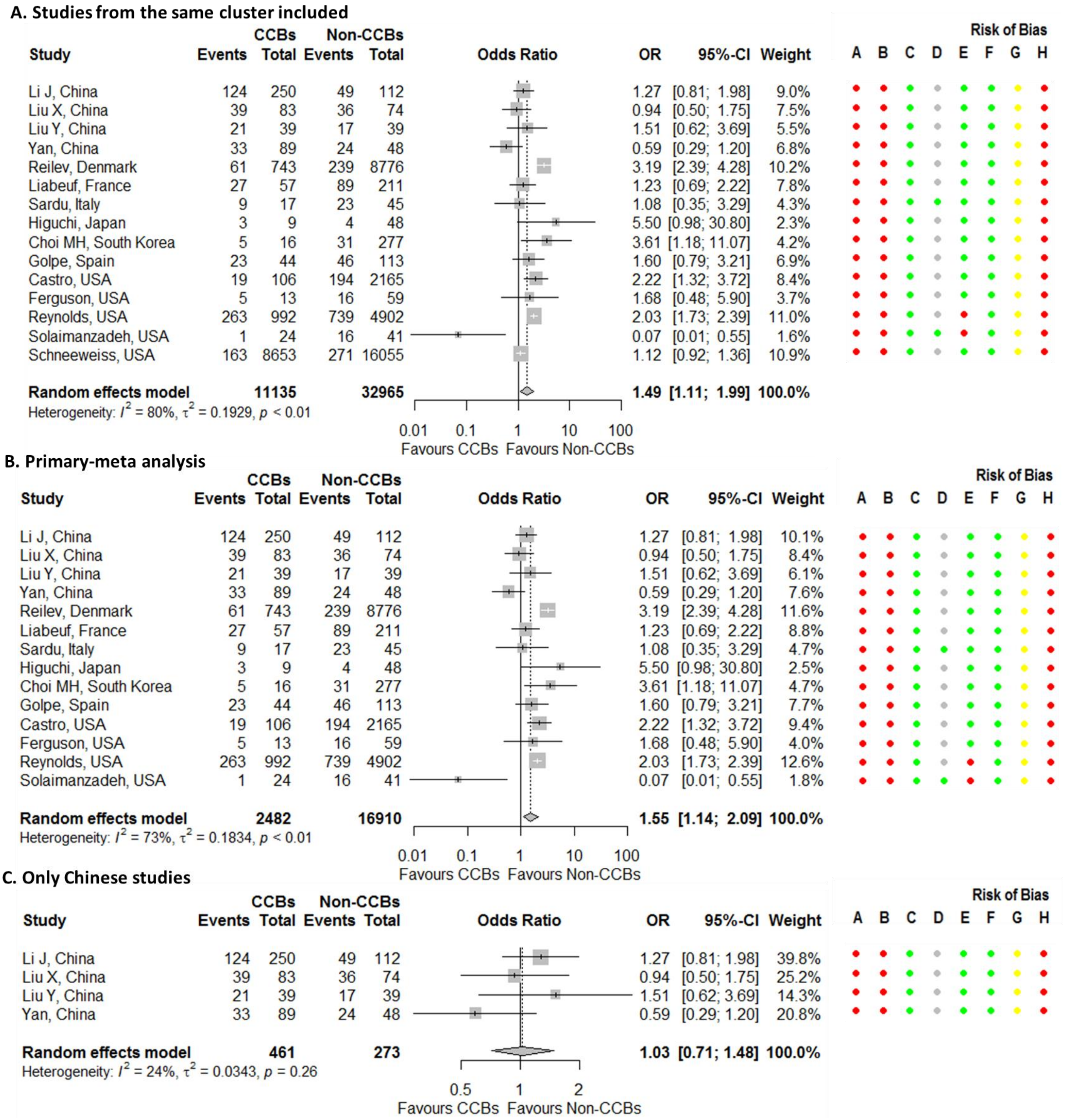


**Figure S35. Forest plots for association between severity outcomes in COVID-19 patients and being on a calcium channel blocker (CCB).** Risk of bias legend. A = risk of bias due to confounding, B = risk of bias in selection of participants into the study, C = risk of bias in classification of interventions, D = risk of bias due to deviations from intended interventions, E = risk of bias due to missing data, F = risk of bias in measurement of outcomes, G = risk of bias in selection of the reported result, H = overall risk of bias. Color codes. Colour codes. Red = serious, yellow = moderate, green = low, grey = unclear.

**D. Chinese studies excluded**

| Study | CCBs Events | CCBs Total | Non-CCBs Events | Non-CCBs Total | OR | 95%-CI | Weight |
|---|---|---|---|---|---|---|---|
| Reilev, Denmark | 61 | 743 | 239 | 8776 | 3.19 | [2.39; 4.28] | 18.7% |
| Liabeuf, France | 27 | 57 | 89 | 211 | 1.23 | [0.69; 2.22] | 12.8% |
| Sardu, Italy | 9 | 17 | 23 | 45 | 1.08 | [0.35; 3.29] | 6.1% |
| Higuchi, Japan | 3 | 9 | 4 | 48 | 5.50 | [0.98; 30.80] | 3.1% |
| Choi MH, South Korea | 5 | 16 | 31 | 277 | 3.61 | [1.18; 11.07] | 6.1% |
| Golpe, Spain | 23 | 44 | 46 | 113 | 1.60 | [0.79; 3.21] | 10.9% |
| Castro, USA | 19 | 106 | 194 | 2165 | 2.22 | [1.32; 3.72] | 14.1% |
| Ferguson, USA | 5 | 13 | 16 | 59 | 1.68 | [0.48; 5.90] | 5.1% |
| Reynolds, USA | 263 | 992 | 739 | 4902 | 2.03 | [1.73; 2.39] | 20.9% |
| Solaimanzadeh, USA | 1 | 24 | 16 | 41 | 0.07 | [0.01; 0.55] | 2.2% |
| **Random effects model** | | **2021** | | **16637** | **1.93** | **[1.40; 2.67]** | **100.0%** |

Heterogeneity: $I^2 = 65\%$, $\tau^2 = 0.1241$, $p < 0.01$

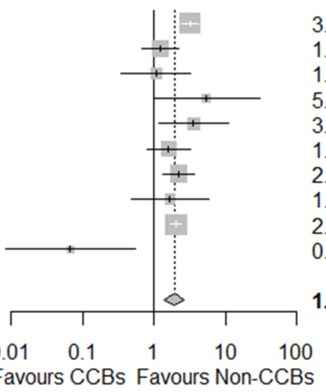


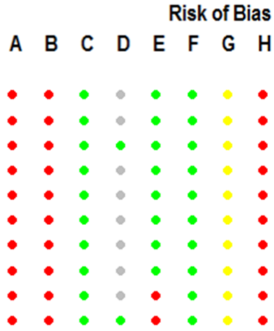


**E. Only hypertensive patients**

| Study | CCBs Events | CCBs Total | Non-CCBs Events | Non-CCBs Total | OR | 95%-CI | Weight |
|---|---|---|---|---|---|---|---|
| Li J, China | 124 | 250 | 49 | 112 | 1.27 | [0.81; 1.98] | 14.5% |
| Liu X, China | 39 | 83 | 36 | 74 | 0.94 | [0.50; 1.75] | 7.4% |
| Liu Y, China | 21 | 39 | 17 | 39 | 1.51 | [0.62; 3.69] | 3.6% |
| Sardu, Italy | 9 | 17 | 23 | 45 | 1.08 | [0.35; 3.29] | 2.3% |
| Choi MH, South Korea | 5 | 16 | 4 | 13 | 1.02 | [0.21; 4.98] | 1.2% |
| Golpe, Spain | 23 | 44 | 46 | 113 | 1.60 | [0.79; 3.21] | 5.9% |
| Reynolds, USA | 253 | 950 | 207 | 930 | 1.27 | [1.03; 1.57] | 65.1% |
| **Random effects model** | | **1399** | | **1326** | **1.26** | **[1.06; 1.49]** | **100.0%** |

Heterogeneity: $I^2 = 0\%$, $\tau^2 = 0$, $p = 0.95$

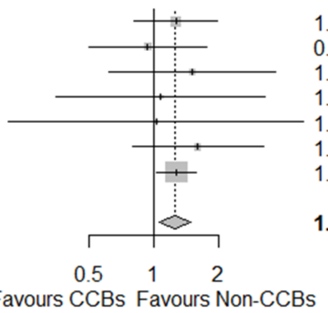


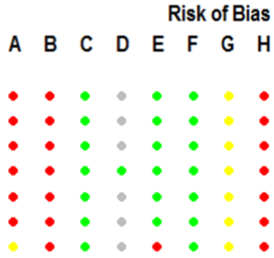


**F. Adjusted estimates**

| Study | TE | seTE | OR | 95%-CI | Weight |
|---|---|---|---|---|---|
| Liu X, China | -0.21 | 0.4596 | 0.81 | [0.33; 1.99] | 15.9% |
| Yan, China | -0.65 | 0.3952 | 0.52 | [0.24; 1.13] | 16.9% |
| Castro, USA | 0.20 | 0.3115 | 1.22 | [0.66; 2.24] | 18.1% |
| Reynolds, USA | 0.22 | 0.0984 | 1.24 | [1.02; 1.50] | 20.3% |
| Choi MH, South Korea | 2.32 | 0.6974 | 10.20 | [2.60; 40.01] | 12.3% |
| Golpe, Spain | 0.31 | 0.4181 | 1.36 | [0.60; 3.09] | 16.5% |
| **Random effects model** | | | **1.31** | **[0.62; 2.80]** | **100.0%** |

Heterogeneity: $I^2 = 66\%$, $\tau^2 = 0.7255$, $p = 0.01$

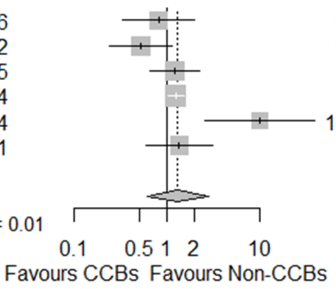


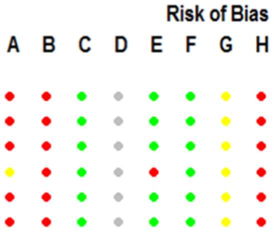


**Figure S35. Continued.**

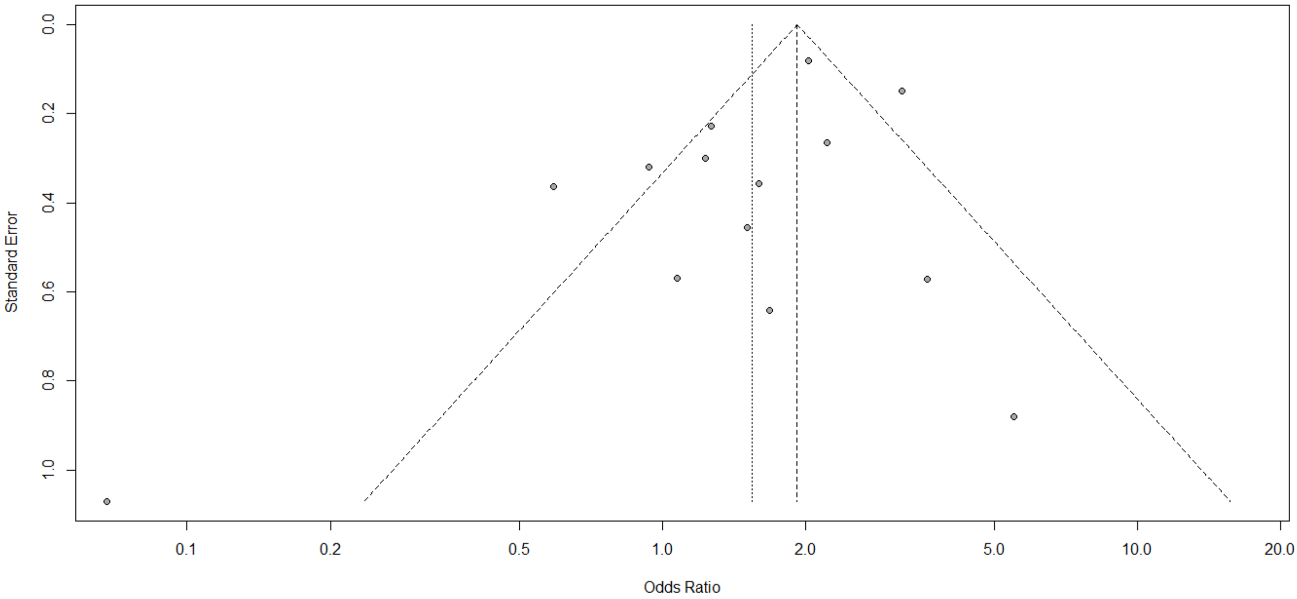


**Figure S36. Funnel plot for association between severity outcomes in COVID-19 patients and being on a calcium channel blocker.**

**A. Primary meta-analysis**

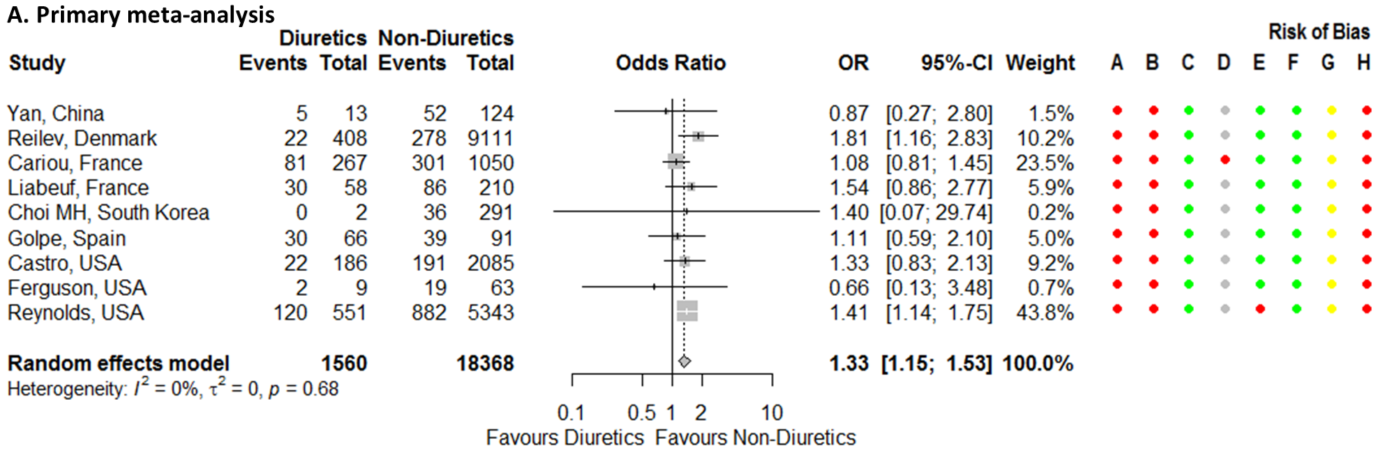


**B. Loop diuretics**

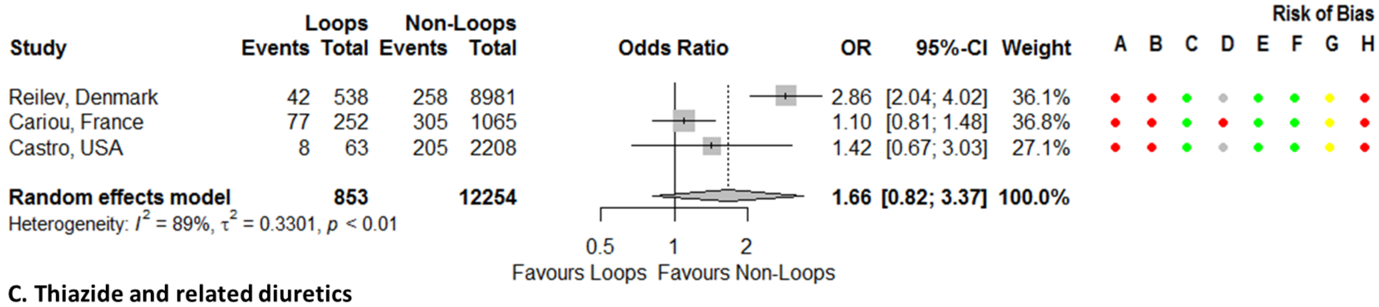


**C. Thiazide and related diuretics**

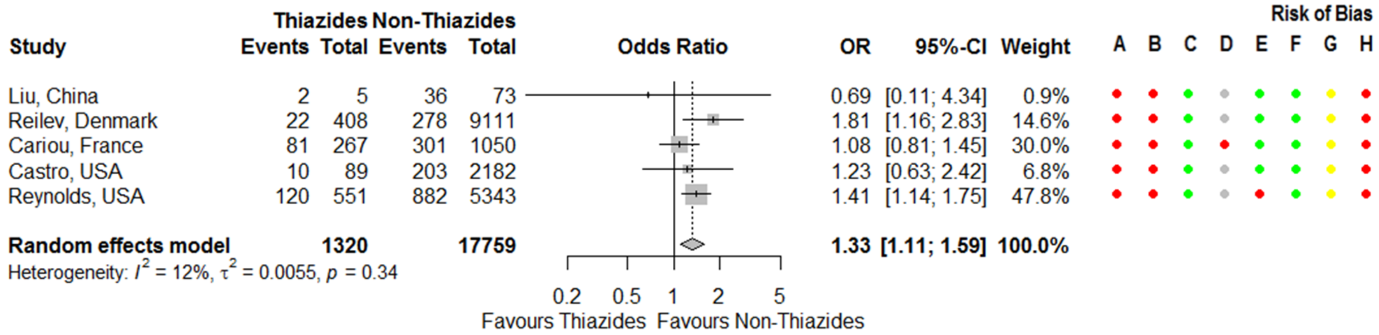


**D. Potassium-sparing diuretics and aldosterone antagonists (PSDAAs)**

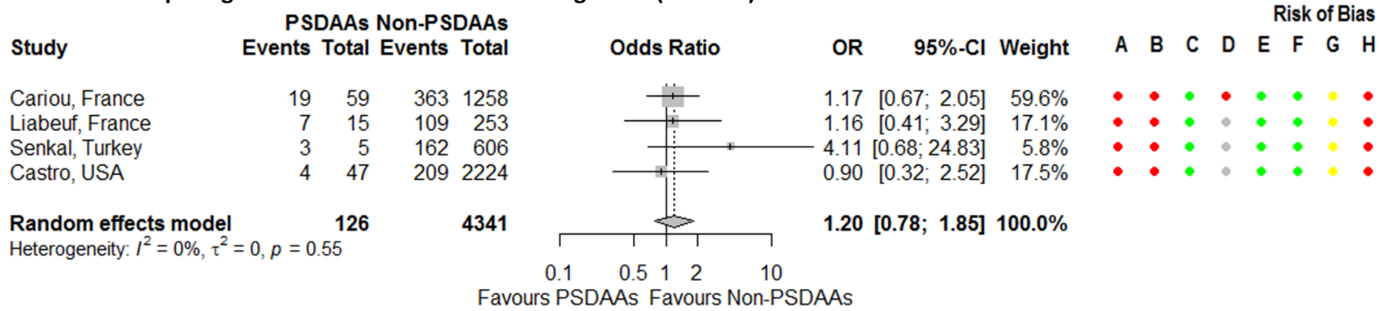


**E. Adjusted estimates**

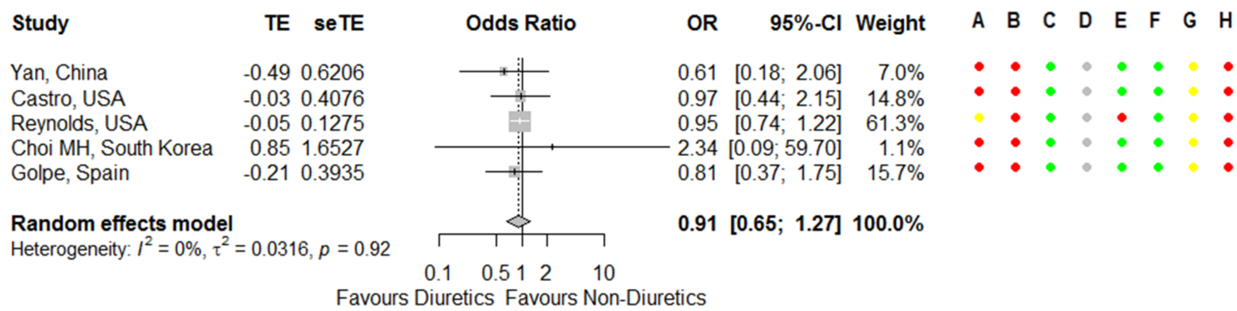

**Figure S37. Forest plots for association between severity outcomes in COVID-19 patients and being on a diuretic.** Risk of bias legend. A = risk of bias due to confounding, B = risk of bias in selection of participants into the study, C = risk of bias in classification of interventions, D = risk of bias due to deviations from intended interventions, E = risk of bias due to missing data, F = risk of bias in measurement of outcomes, G = risk of bias in selection of the reported result, H = overall risk of bias. Color codes. Colour codes. Red = serious, yellow = moderate, green = low, grey = unclear.

**A. Studies from the same cluster included**

| Study | LMDs Events | LMDs Total | Non-LMDs Events | Non-LMDs Total | OR | 95%-CI | Weight |
|---|---|---|---|---|---|---|---|
| De Spiegeleer, Belgium | 6 | 31 | 31 | 123 | 0.71 | [0.27; 1.90] | 5.8% |
| Jiang, China | 0 | 4 | 27 | 127 | 0.41 | [0.02; 7.77] | 1.2% |
| Qin, China | 2 | 12 | 13 | 38 | 0.38 | [0.07; 2.02] | 3.1% |
| Yan, China | 5 | 16 | 123 | 594 | 1.74 | [0.59; 5.10] | 5.3% |
| Yin, China | 19 | 33 | 40 | 73 | 1.12 | [0.49; 2.57] | 6.6% |
| Reilev, Denmark | 106 | 1277 | 194 | 8242 | 3.76 | [2.94; 4.80] | 10.1% |
| Cariou, France | 184 | 627 | 198 | 690 | 1.03 | [0.81; 1.31] | 10.1% |
| Sacco, Germany | 1 | 10 | 38 | 155 | 0.34 | [0.04; 2.79] | 2.2% |
| Higuchi, Japan | 4 | 12 | 3 | 45 | 7.00 | [1.31; 37.45] | 3.1% |
| Choi MH, South Korea | 6 | 16 | 30 | 277 | 4.94 | [1.68; 14.56] | 5.2% |
| Argenziano, USA | 93 | 361 | 143 | 639 | 1.20 | [0.89; 1.63] | 9.8% |
| Castro, USA | 25 | 180 | 188 | 2091 | 1.63 | [1.04; 2.56] | 9.0% |
| Ferguson, USA | 7 | 17 | 14 | 55 | 2.05 | [0.66; 6.41] | 5.0% |
| Garibaldi, USA | 111 | 271 | 118 | 476 | 2.10 | [1.53; 2.90] | 9.8% |
| Gu, USA | 33 | 144 | 63 | 582 | 2.45 | [1.53; 3.91] | 8.9% |
| Goshua, USA | 15 | 20 | 33 | 48 | 1.36 | [0.42; 4.45] | 4.8% |
| **Random effects model** | | **3031** | | **14255** | **1.65** | **[1.16; 2.33]** | **100.0%** |

Heterogeneity: $I^2 = 82\%$, $\tau^2 = 0.2967$, $p < 0.01$

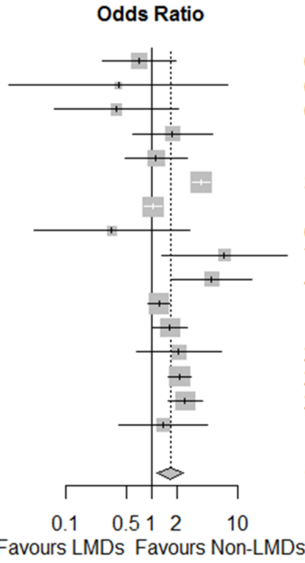


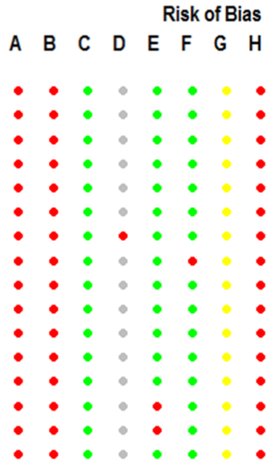


**B. Primary meta-analysis**

| Study | LMDs Events | LMDs Total | Non-LMDs Events | Non-LMDs Total | OR | 95%-CI | Weight |
|---|---|---|---|---|---|---|---|
| De Spiegeleer, Belgium | 6 | 31 | 31 | 123 | 0.71 | [0.27; 1.90] | 5.8% |
| Qin, China | 2 | 12 | 13 | 38 | 0.38 | [0.07; 2.02] | 3.2% |
| Yan, China | 5 | 16 | 123 | 594 | 1.74 | [0.59; 5.10] | 5.3% |
| Yin, China | 19 | 33 | 40 | 73 | 1.12 | [0.49; 2.57] | 6.7% |
| Reilev, Denmark | 106 | 1277 | 194 | 8242 | 3.76 | [2.94; 4.80] | 10.2% |
| Cariou, France | 184 | 627 | 198 | 690 | 1.03 | [0.81; 1.31] | 10.2% |
| Sacco, Germany | 1 | 10 | 38 | 155 | 0.34 | [0.04; 2.79] | 2.2% |
| Higuchi, Japan | 4 | 12 | 3 | 45 | 7.00 | [1.31; 37.45] | 3.1% |
| Choi MH, South Korea | 6 | 16 | 30 | 277 | 4.94 | [1.68; 14.56] | 5.3% |
| Argenziano, USA | 93 | 361 | 143 | 639 | 1.20 | [0.89; 1.63] | 10.0% |
| Castro, USA | 25 | 180 | 188 | 2091 | 1.63 | [1.04; 2.56] | 9.1% |
| Ferguson, USA | 7 | 17 | 14 | 55 | 2.05 | [0.66; 6.41] | 5.0% |
| Garibaldi, USA | 111 | 271 | 118 | 476 | 2.10 | [1.53; 2.90] | 9.9% |
| Gu, USA | 33 | 144 | 63 | 582 | 2.45 | [1.53; 3.91] | 9.0% |
| Goshua, USA | 15 | 20 | 33 | 48 | 1.36 | [0.42; 4.45] | 4.8% |
| **Random effects model** | | **3027** | | **14128** | **1.68** | **[1.18; 2.38]** | **100.0%** |

Heterogeneity: $I^2 = 83\%$, $\tau^2 = 0.2976$, $p < 0.01$

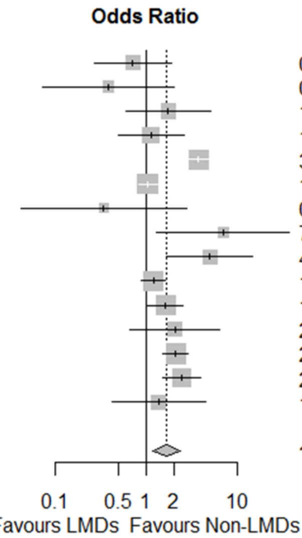


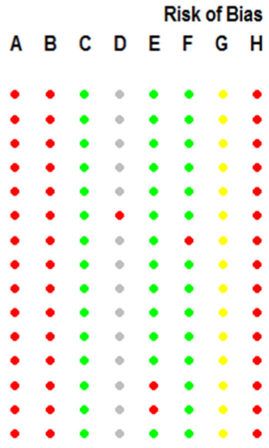


**C. Peer-reviewed studies**

| Study | LMDs Events | LMDs Total | Non-LMDs Events | Non-LMDs Total | OR | 95%-CI | Weight |
|---|---|---|---|---|---|---|---|
| De Spiegeleer, Belgium | 6 | 31 | 31 | 123 | 0.71 | [0.27; 1.90] | 9.4% |
| Qin, China | 2 | 12 | 13 | 38 | 0.38 | [0.07; 2.02] | 3.8% |
| Cariou, France | 184 | 627 | 198 | 690 | 1.03 | [0.81; 1.31] | 33.7% |
| Choi MH, South Korea | 6 | 16 | 30 | 277 | 4.94 | [1.68; 14.56] | 8.0% |
| Argenziano, USA | 93 | 361 | 143 | 639 | 1.20 | [0.89; 1.63] | 30.8% |
| Ferguson, USA | 7 | 17 | 14 | 55 | 2.05 | [0.66; 6.41] | 7.3% |
| Goshua, USA | 15 | 20 | 33 | 48 | 1.36 | [0.42; 4.45] | 6.9% |
| **Random effects model** | | **1084** | | **1870** | **1.22** | **[0.87; 1.72]** | **100.0%** |

Heterogeneity: $I^2 = 48\%$, $\tau^2 = 0.0753$, $p = 0.07$

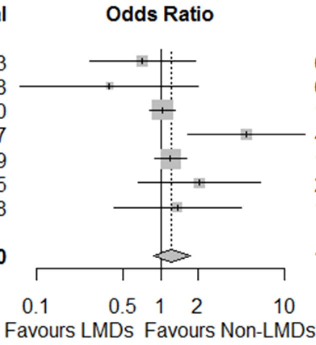


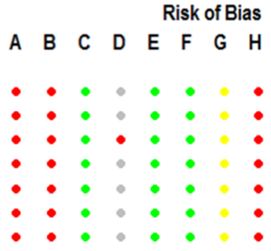


**Figure S38. Forest plots for association between severity outcomes in COVID-19 patients and being on a lipid modifying drug (LMD).** Risk of bias legend. A = risk of bias due to confounding, B = risk of bias in selection of participants into the study, C = risk of bias in classification of interventions, D = risk of bias due to deviations from intended interventions, E = risk of bias due to missing data, F = risk of bias in measurement of outcomes, G = risk of bias in selection of the reported result, H = overall risk of bias. Color codes. Colour codes. Red = serious, yellow = moderate, green = low, grey = unclear.

**D. Preprints**

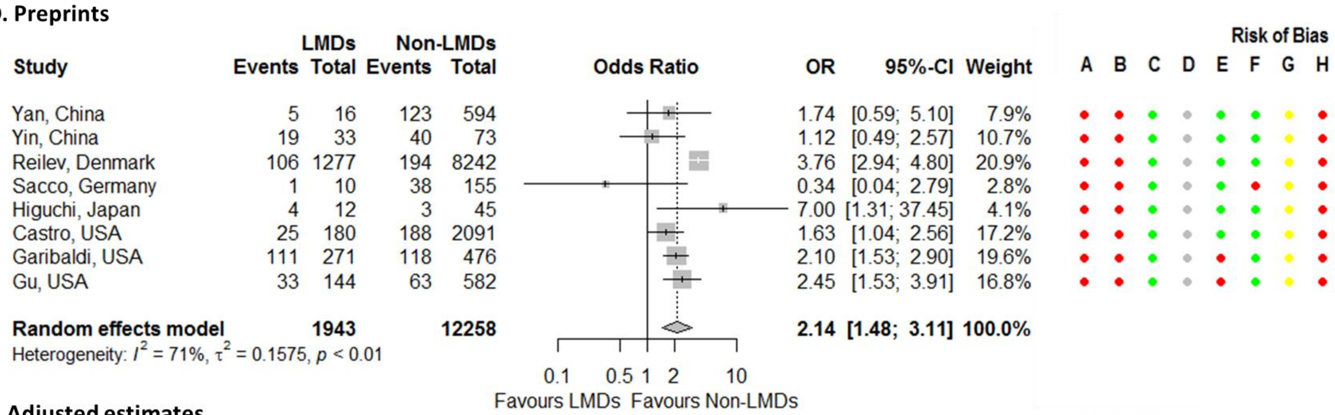


**E. Adjusted estimates**

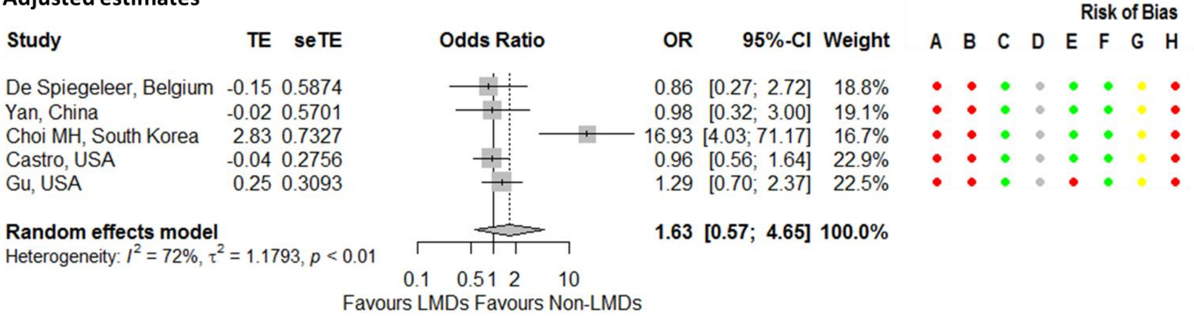


**Figure S38. Continued.**

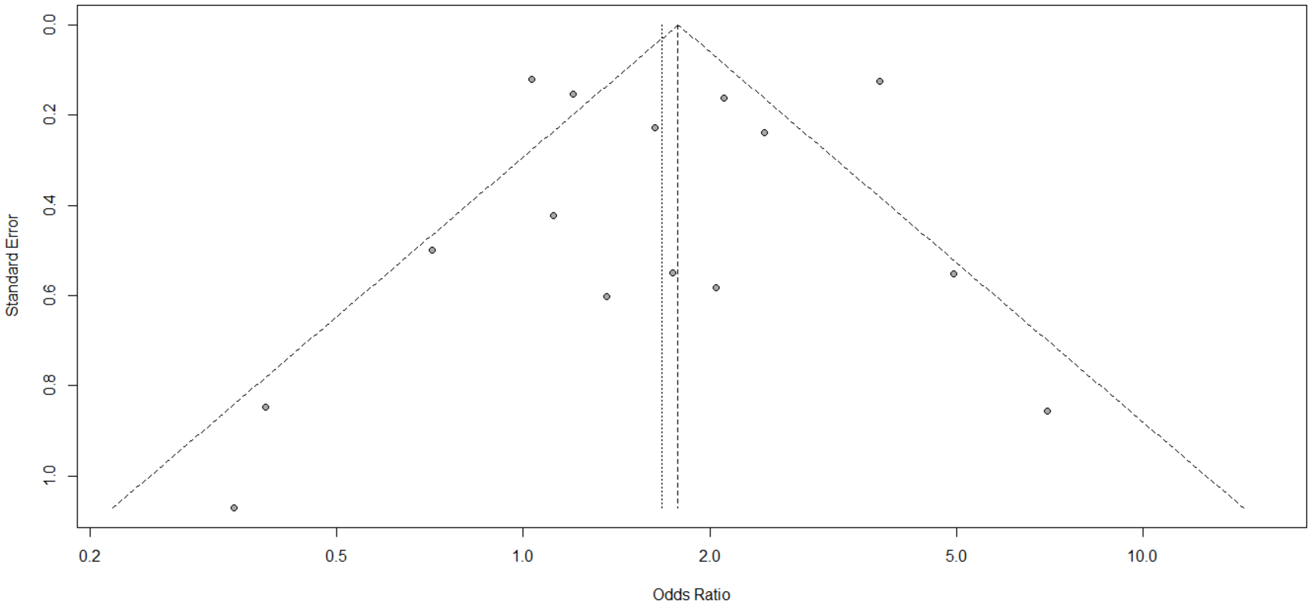


**Figure S39. Funnel plot for association between severity outcomes in COVID-19 patients and being on a lipid modifying drug.**

**A. Studies from the same cluster included**

| Study | ACEI/ARB Events | ACEI/ARB Total | Non−ACEI/ARB Events | Non−ACEI/ARB Total | OR | 95%−CI | Weight |
|---|---|---|---|---|---|---|---|
| Mazzoleni, Belgium | 2 | 12 | 9 | 28 | 0.42 | [0.08; 2.34] | 0.7% |
| Chen M, China | 3 | 11 | 28 | 112 | 1.12 | [0.28; 4.54] | 0.9% |
| Chen Y, China | 4 | 32 | 10 | 39 | 0.41 | [0.12; 1.48] | 1.0% |
| Gao, China | 4 | 183 | 30 | 667 | 0.47 | [0.16; 1.36] | 1.2% |
| Guo, China | 7 | 19 | 36 | 168 | 2.14 | [0.79; 5.83] | 1.2% |
| Hu, China | 1 | 65 | 0 | 84 | 3.93 | [0.16; 98.07] | 0.2% |
| Huang, China | 0 | 20 | 3 | 30 | 0.19 | [0.01; 3.92] | 0.3% |
| Li J, China | 21 | 115 | 56 | 247 | 0.76 | [0.44; 1.33] | 1.8% |
| Liu X, China | 5 | 74 | 1 | 83 | 5.94 | [0.68; 52.08] | 0.5% |
| Meng, China | 0 | 17 | 1 | 25 | 0.47 | [0.02; 12.14] | 0.2% |
| Peng, China | 4 | 22 | 13 | 90 | 1.32 | [0.38; 4.51] | 1.0% |
| Tan, China | 0 | 31 | 11 | 69 | 0.08 | [0.00; 1.42] | 0.3% |
| Xie Yang, China | 2 | 35 | 26 | 584 | 1.30 | [0.30; 5.72] | 0.8% |
| Xu, China | 11 | 40 | 21 | 61 | 0.72 | [0.30; 1.73] | 1.4% |
| Yang G, China | 2 | 43 | 11 | 83 | 0.32 | [0.07; 1.51] | 0.7% |
| Yang W, China | 30 | 62 | 103 | 282 | 1.63 | [0.94; 2.84] | 1.8% |
| Ye, China | 1 | 62 | 0 | 80 | 3.93 | [0.16; 98.06] | 0.2% |
| Zeng H, China | 8 | 62 | 157 | 969 | 0.77 | [0.36; 1.64] | 1.5% |
| Zeng Z, China | 2 | 28 | 5 | 47 | 0.65 | [0.12; 3.58] | 0.7% |
| Zhang P, China | 7 | 188 | 92 | 940 | 0.36 | [0.16; 0.78] | 1.5% |
| Zhang XJ , China | 7 | 204 | 20 | 204 | 0.33 | [0.14; 0.79] | 1.4% |
| Zhou X, China | 2 | 15 | 5 | 21 | 0.49 | [0.08; 2.97] | 0.6% |
| Fosbol, Denmark | 181 | 895 | 297 | 3585 | 2.81 | [2.29; 3.43] | 2.2% |
| Reilev, Denmark | 187 | 1428 | 337 | 8091 | 3.47 | [2.87; 4.19] | 2.2% |
| Cariou, France | 92 | 737 | 48 | 580 | 1.58 | [1.09; 2.28] | 2.1% |
| Liabeuf, France | 17 | 96 | 30 | 172 | 1.02 | [0.53; 1.96] | 1.7% |
| Meszaros, France | 15 | 68 | 3 | 35 | 3.02 | [0.81; 11.24] | 0.9% |
| Oussalah, France | 10 | 43 | 9 | 104 | 3.20 | [1.20; 8.55] | 1.3% |
| Rath, Germany | 8 | 60 | 8 | 63 | 1.06 | [0.37; 3.02] | 1.2% |
| Ashraf, Iran | 2 | 14 | 10 | 68 | 0.97 | [0.19; 4.99] | 0.7% |
| Alberici, Italy | 3 | 9 | 2 | 11 | 2.25 | [0.29; 17.76] | 0.5% |
| Benelli, Italy | 25 | 110 | 47 | 301 | 1.59 | [0.92; 2.74] | 1.8% |
| Cannata, Italy | 39 | 173 | 39 | 224 | 1.38 | [0.84; 2.27] | 1.9% |
| Conversano, Italy | 21 | 69 | 21 | 122 | 2.10 | [1.05; 4.22] | 1.6% |
| Felice, Italy | 15 | 82 | 18 | 51 | 0.41 | [0.18; 0.92] | 1.5% |
| Giacomelli, Italy | 23 | 62 | 25 | 171 | 3.44 | [1.77; 6.71] | 1.7% |
| Giorgi Rossi, Italy | 108 | 818 | 109 | 1835 | 2.41 | [1.82; 3.19] | 2.1% |
| Iaccarino, Italy | 106 | 655 | 82 | 936 | 2.01 | [1.48; 2.73] | 2.1% |
| Inciardi, Italy | 9 | 30 | 17 | 69 | 1.31 | [0.51; 3.40] | 1.3% |
| Sardu, Italy | 7 | 45 | 2 | 17 | 1.38 | [0.26; 7.42] | 0.7% |
| Trecarichi, Italy | 3 | 16 | 11 | 32 | 0.44 | [0.10; 1.88] | 0.8% |
| Violi, Italy | 19 | 111 | 45 | 208 | 0.75 | [0.41; 1.35] | 1.8% |
| Ayed, Kuwait | 5 | 10 | 40 | 93 | 1.32 | [0.36; 4.89] | 0.9% |
| Garassino, Multiple | 17 | 53 | 47 | 135 | 0.88 | [0.45; 1.74] | 1.6% |
| Jung, Multiple | 62 | 157 | 85 | 167 | 0.63 | [0.41; 0.98] | 2.0% |
| Choi HK, South Korea | 49 | 892 | 79 | 693 | 0.45 | [0.31; 0.65] | 2.0% |
| Hwang, South Korea | 5 | 13 | 21 | 90 | 2.05 | [0.61; 6.95] | 1.0% |
| Jung, South Korea | 33 | 762 | 51 | 4417 | 3.88 | [2.48; 6.05] | 2.0% |
| Lee, South Korea | 50 | 977 | 62 | 7289 | 6.29 | [4.30; 9.18] | 2.0% |
| Amat−Santos, Spain | 2 | 5 | 2 | 6 | 1.33 | [0.11; 15.70] | 0.4% |
| Fernández−Ruiz, Spain | 1 | 7 | 4 | 11 | 0.29 | [0.03; 3.37] | 0.4% |
| Lopez−Otero, Spain | 11 | 210 | 27 | 755 | 1.49 | [0.73; 3.06] | 1.6% |
| Lorente−Ros, Spain | 60 | 225 | 80 | 482 | 1.83 | [1.25; 2.67] | 2.0% |
| Pérez−Sáez, Spain | 10 | 26 | 16 | 54 | 1.48 | [0.56; 3.97] | 1.3% |
| Poblador−Plou, Spain | 159 | 631 | 612 | 3781 | 1.74 | [1.43; 2.13] | 2.2% |
| Selçuk, Turkey | 31 | 74 | 4 | 39 | 6.31 | [2.03; 19.58] | 1.1% |
| Senkal, Turkey | 7 | 104 | 5 | 52 | 0.68 | [0.20; 2.25] | 1.0% |
| Baker, UK | 17 | 70 | 63 | 216 | 0.78 | [0.42; 1.45] | 1.7% |
| Bean, UK | 106 | 399 | 182 | 801 | 1.23 | [0.93; 1.62] | 2.2% |
| Bae, USA | 1 | 78 | 5 | 512 | 1.32 | [0.15; 11.42] | 0.5% |
| Garibaldi, USA | 11 | 151 | 102 | 596 | 0.38 | [0.20; 0.73] | 1.7% |
| Gupta, USA | 311 | 766 | 473 | 1449 | 1.41 | [1.18; 1.69] | 2.2% |
| Imam, USA | 105 | 565 | 95 | 740 | 1.55 | [1.15; 2.10] | 2.1% |
| Ip, USA | 137 | 460 | 262 | 669 | 0.66 | [0.51; 0.85] | 2.2% |
| Khera, USA | 664 | 4587 | 466 | 3346 | 1.05 | [0.92; 1.19] | 2.3% |
| Kim (ACEI), USA | 53 | 316 | 264 | 1576 | 1.00 | [0.72; 1.38] | 2.1% |
| Kim (ARB), USA | 52 | 257 | 266 | 1638 | 1.31 | [0.94; 1.82] | 2.1% |
| Lam, USA | 58 | 335 | 62 | 279 | 0.73 | [0.49; 1.09] | 2.0% |
| Mehta, USA | 8 | 211 | 34 | 1494 | 1.69 | [0.77; 3.71] | 1.5% |
| Nguyen, USA | 11 | 99 | 19 | 194 | 1.15 | [0.52; 2.53] | 1.5% |
| Palaiodimos, USA | 13 | 62 | 35 | 138 | 0.78 | [0.38; 1.61] | 1.6% |
| Richardson, USA | 130 | 413 | 254 | 953 | 1.26 | [0.98; 1.63] | 2.2% |
| Wang, USA | 9 | 26 | 5 | 32 | 2.86 | [0.82; 9.98] | 1.0% |
| **Random effects model** | | **19802** | | **54285** | **1.23** | **[1.04; 1.44]** | **100.0%** |

Heterogeneity: $I^2 = 85\%$, $\tau^2 = 0.3030$, $p < 0.01$

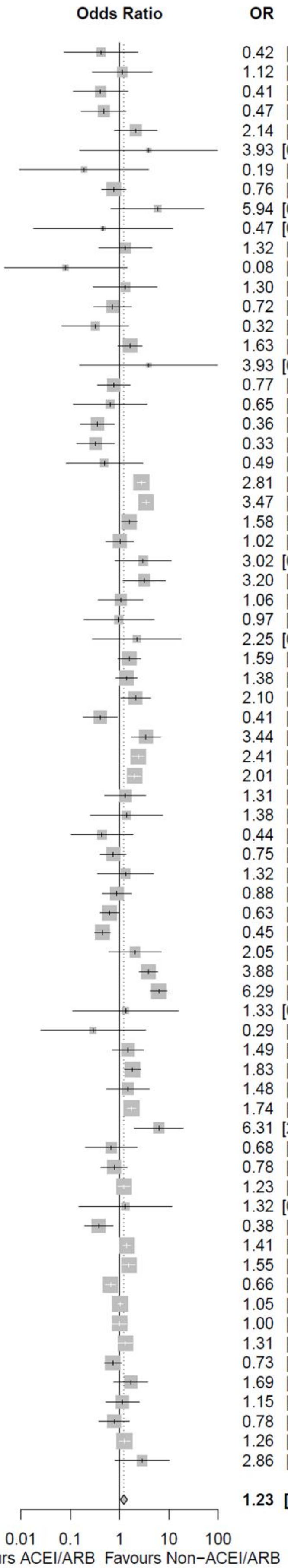


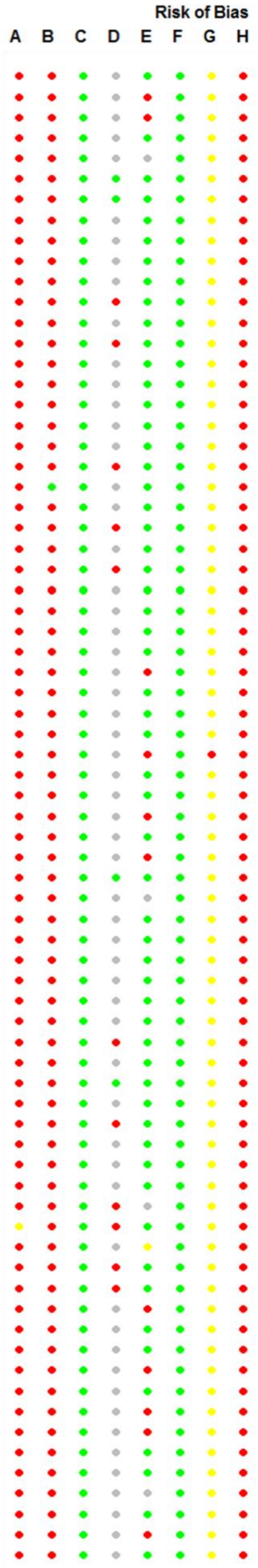

**B. China-only studies**

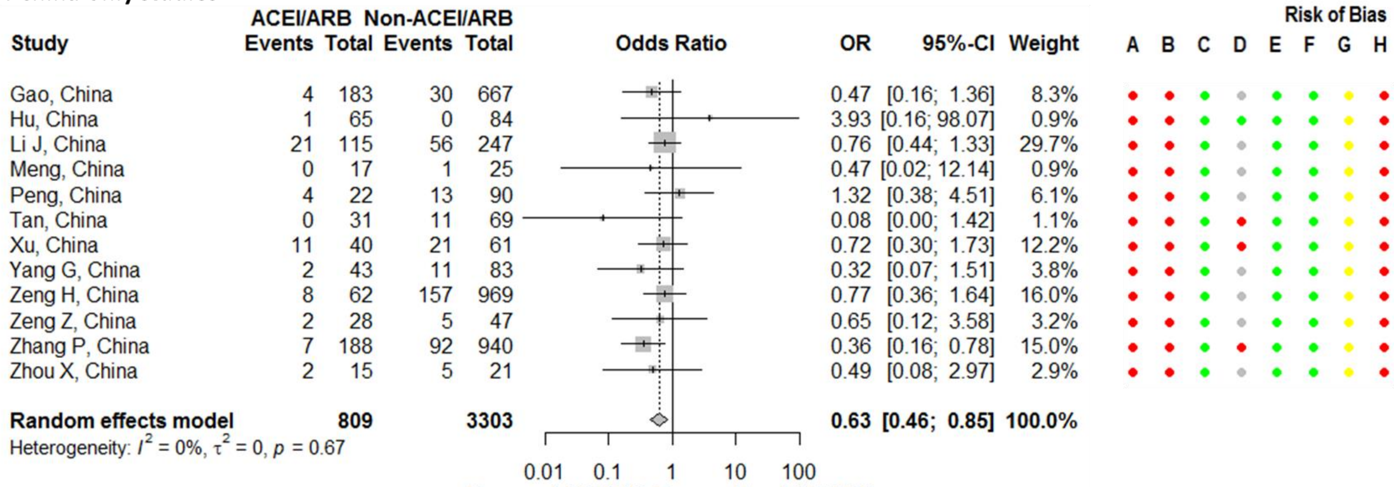


**C. Chinese studies excluded**

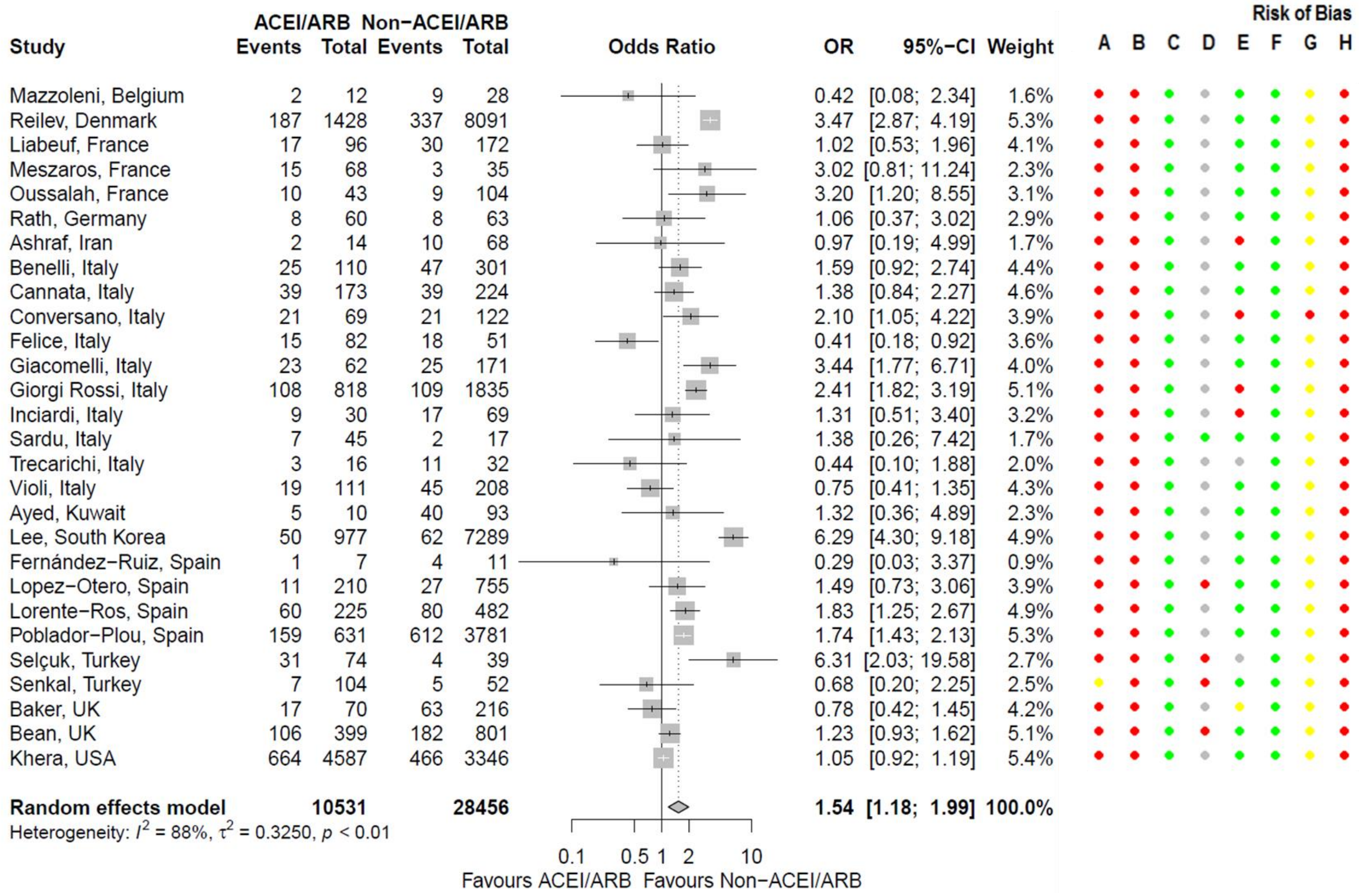


**Figure S40. Forest plots for association between mortality outcomes in COVID-19 patients and being on an angiotensin-converting enzyme inhibitor (ACEI) or angiotensin receptor blocker (ARB).** Benelli,[89] Fernández-Ruiz,[130] Garassino,[111] Giacomelli,[96] Giorgi Rossi,[97] Gupta,[163] Iaccarino,[99] Lam,[172] Nguyen,[176] Poblador-Plou,[137] and Violi[110] estimates assume that none of the patients are taking both ACEis and ARBs. Risk of bias legend. A = risk of bias due to confounding, B = risk of bias in selection of participants into the study, C = risk of bias in classification of interventions, D = risk of bias due to deviations from intended interventions, E = risk of bias due to missing data, F = risk of bias in measurement of outcomes, G = risk of bias in selection of the reported result, H = overall risk of bias. Color codes. Colour codes. Red = serious, yellow = moderate, green = low, grey = unclear.

**D. Adjusted estimates**

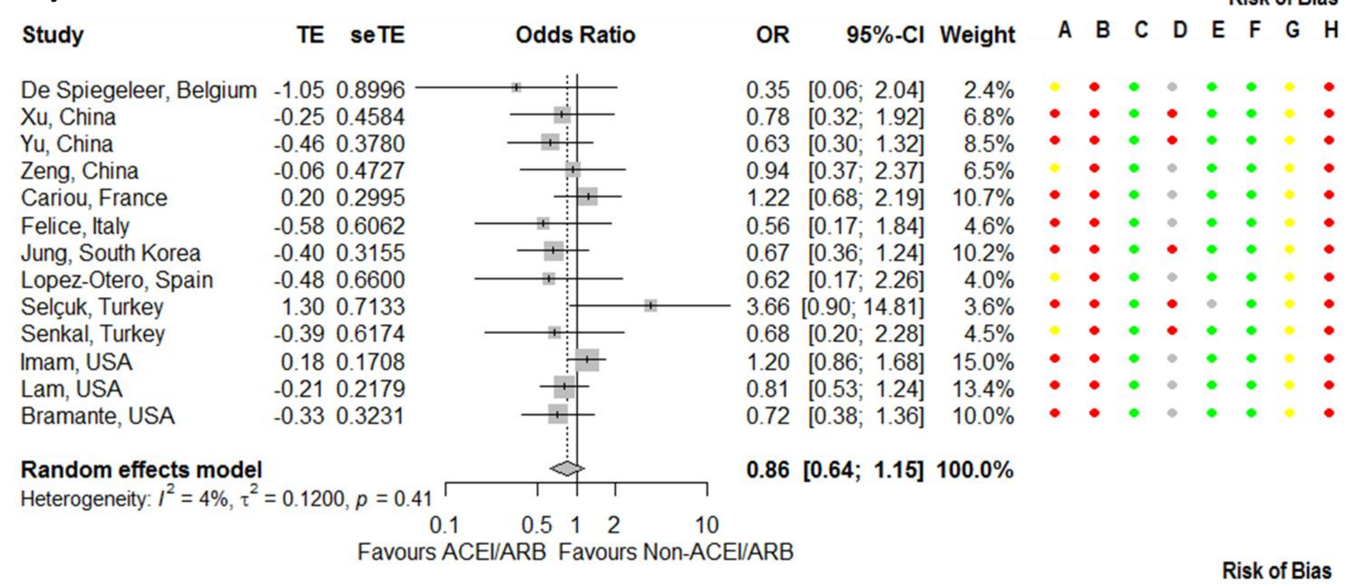


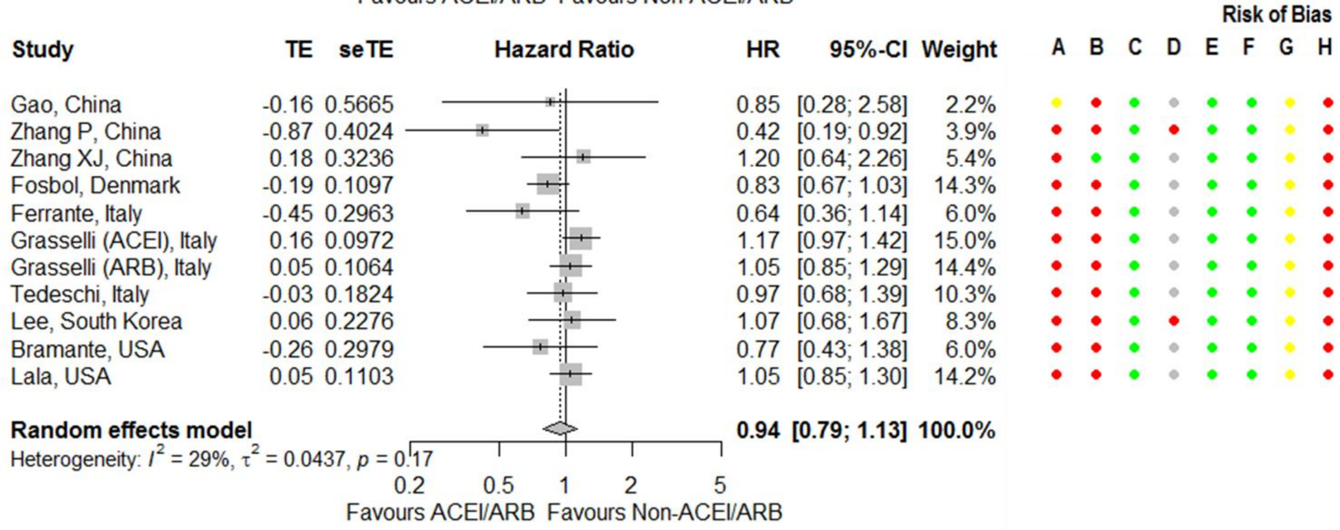


**Figure S40. Continued.**

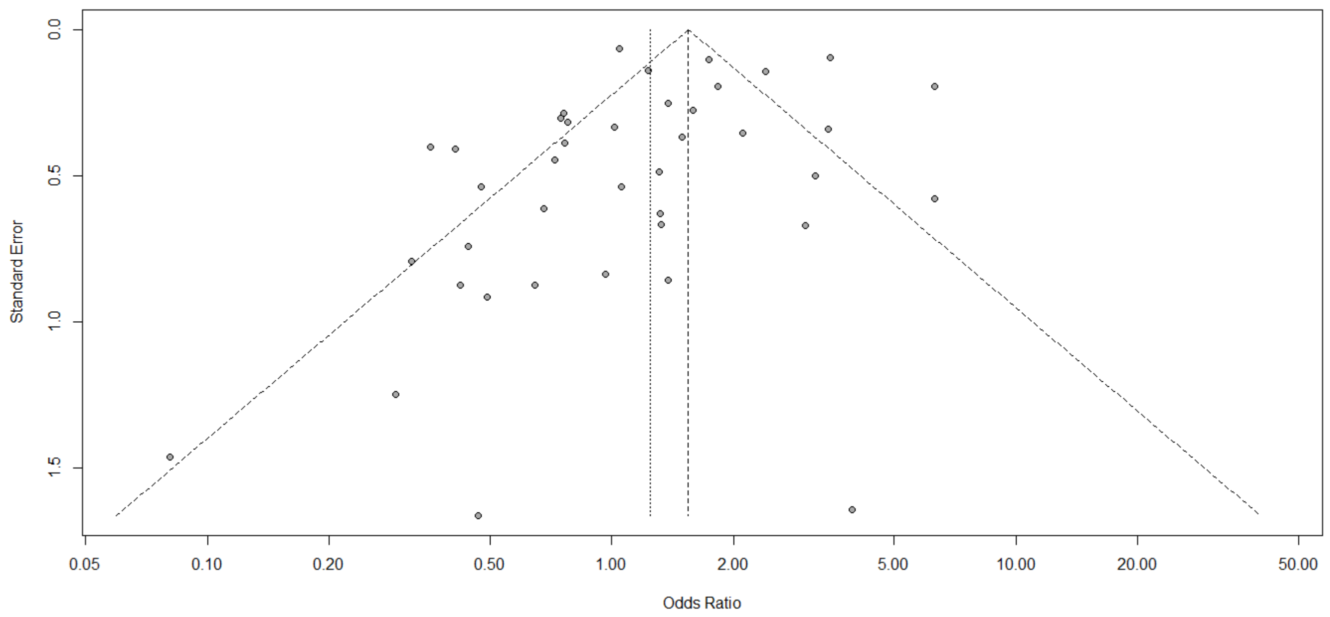


**Figure S41. Funnel plot for association between mortality outcomes in COVID-19 patients on an angiotensin-converting enzyme inhibitor or angiotensin receptor blocker.**

**A. Studies from the same cluster included**

| Study | ACEI Events | ACEI Total | Non−ACEI Events | Non−ACEI Total | OR | 95%−CI | Weight |
|---|---|---|---|---|---|---|---|
| Gao, China | 1 | 52 | 33 | 798 | 0.45 | [0.06; 3.39] | 0.9% |
| Li J, China | 7 | 35 | 70 | 327 | 0.92 | [0.38; 2.19] | 3.0% |
| Yang W, China | 30 | 62 | 103 | 282 | 1.63 | [0.94; 2.84] | 4.3% |
| Fosbol, Denmark | 76 | 377 | 256 | 3585 | 3.28 | [2.48; 4.35] | 5.5% |
| Cariou, France | 47 | 354 | 93 | 963 | 1.43 | [0.99; 2.08] | 5.2% |
| Ashraf, Iran | 0 | 3 | 12 | 79 | 0.77 | [0.04; 15.87] | 0.5% |
| Alberici, Italy | 2 | 6 | 3 | 14 | 1.83 | [0.22; 15.33] | 0.9% |
| Benelli, Italy | 11 | 50 | 61 | 361 | 1.39 | [0.67; 2.86] | 3.6% |
| Felice, Italy | 8 | 40 | 25 | 93 | 0.68 | [0.28; 1.67] | 2.9% |
| Giacomelli, Italy | 9 | 31 | 39 | 202 | 1.71 | [0.73; 4.00] | 3.1% |
| Giorgi Rossi, Italy | 56 | 450 | 161 | 2203 | 1.80 | [1.31; 2.49] | 5.4% |
| Iaccarino, Italy | 63 | 348 | 125 | 1243 | 1.98 | [1.42; 2.75] | 5.4% |
| Sardu, Italy | 4 | 24 | 5 | 38 | 1.32 | [0.32; 5.50] | 1.6% |
| Violi, Italy | 15 | 69 | 49 | 250 | 1.14 | [0.59; 2.19] | 3.9% |
| Ayed, Kuwait | 5 | 10 | 40 | 93 | 1.32 | [0.36; 4.89] | 1.8% |
| Garassino, Multiple | 10 | 28 | 54 | 160 | 1.09 | [0.47; 2.53] | 3.1% |
| Jung C, Multiple | 19 | 62 | 128 | 262 | 0.46 | [0.26; 0.84] | 4.1% |
| Lee, South Korea | 2 | 36 | 110 | 8230 | 4.34 | [1.03; 18.30] | 1.6% |
| Amat−Santos, Spain | 2 | 5 | 2 | 6 | 1.33 | [0.11; 15.70] | 0.7% |
| Fernández−Ruiz, Spain | 1 | 4 | 4 | 14 | 0.83 | [0.07; 10.60] | 0.6% |
| Lopez−Otero, Spain | 2 | 77 | 36 | 888 | 0.63 | [0.15; 2.67] | 1.6% |
| Poblador−Plou, Spain | 78 | 275 | 693 | 4137 | 1.97 | [1.50; 2.59] | 5.6% |
| Senkal, Turkey | 2 | 52 | 10 | 104 | 0.38 | [0.08; 1.78] | 1.4% |
| Garibaldi, USA | 7 | 80 | 106 | 667 | 0.51 | [0.23; 1.13] | 3.3% |
| Gupta, USA | 154 | 401 | 630 | 1814 | 1.17 | [0.94; 1.46] | 5.8% |
| Ip, USA | 70 | 257 | 329 | 872 | 0.62 | [0.45; 0.84] | 5.4% |
| Khera, USA | 319 | 2361 | 811 | 5572 | 0.92 | [0.80; 1.05] | 6.0% |
| Kim (ACEI), USA | 53 | 316 | 264 | 1576 | 1.00 | [0.72; 1.38] | 5.4% |
| Nguyen, USA | 4 | 58 | 26 | 235 | 0.60 | [0.20; 1.78] | 2.3% |
| Richardson, USA | 55 | 168 | 329 | 1198 | 1.29 | [0.91; 1.82] | 5.3% |
| **Random effects model** | | **6091** | | **36266** | **1.18** | **[0.96; 1.46]** | **100.0%** |

Heterogeneity: $I^2 = 79\%$, $\tau^2 = 0.1882$, $p < 0.01$

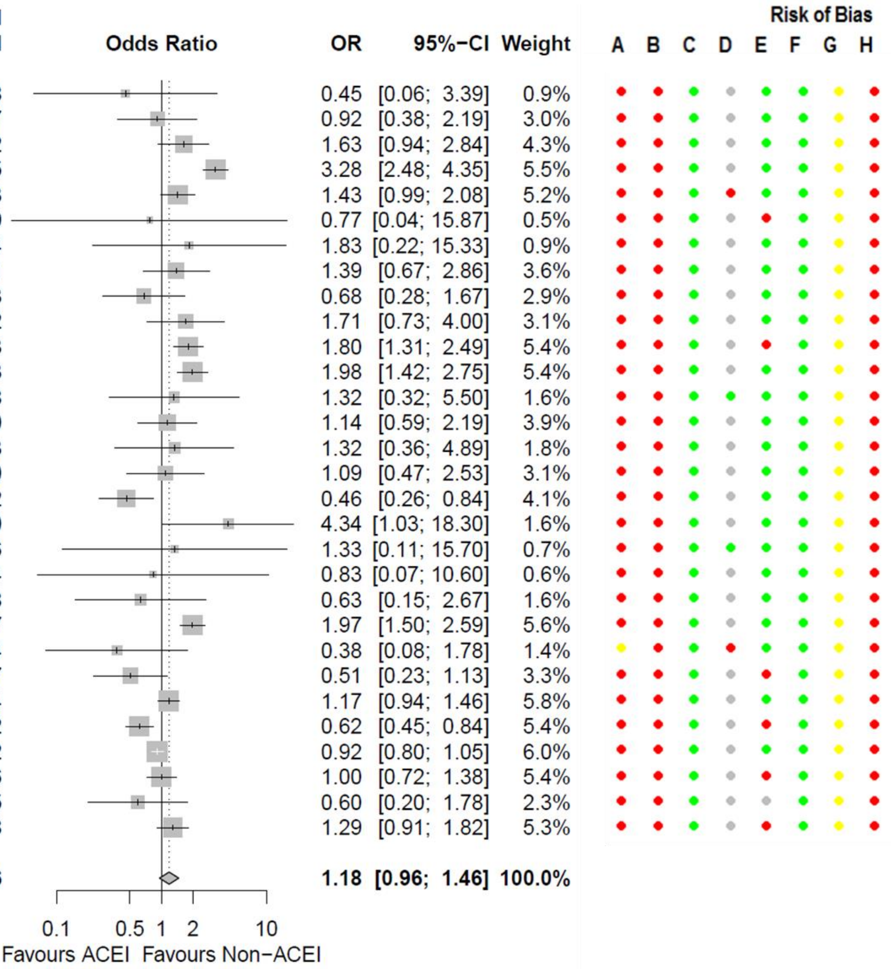


**Figure S42. Forest plot for association between mortality outcomes in COVID-19 patients on an angiotensin-converting enzyme inhibitor (ACEI).** Risk of bias legend. A = risk of bias due to confounding, B = risk of bias in selection of participants into the study, C = risk of bias in classification of interventions, D = risk of bias due to deviations from intended interventions, E = risk of bias due to missing data, F = risk of bias in measurement of outcomes, G = risk of bias in selection of the reported result, H = overall risk of bias. Color codes. Colour codes. Red = serious, yellow = moderate, green = low, grey = unclear.

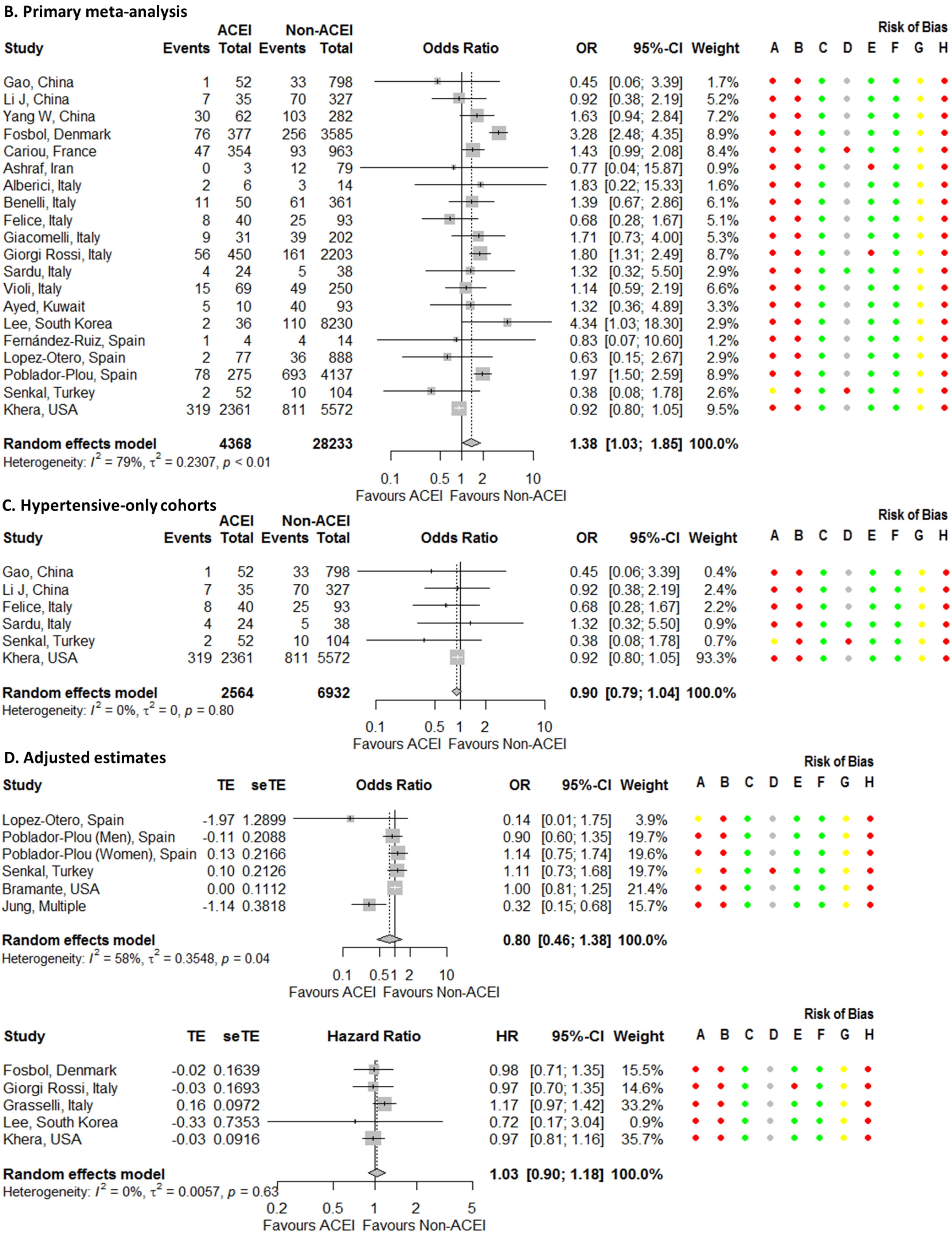


**Figure S42. Continued.**

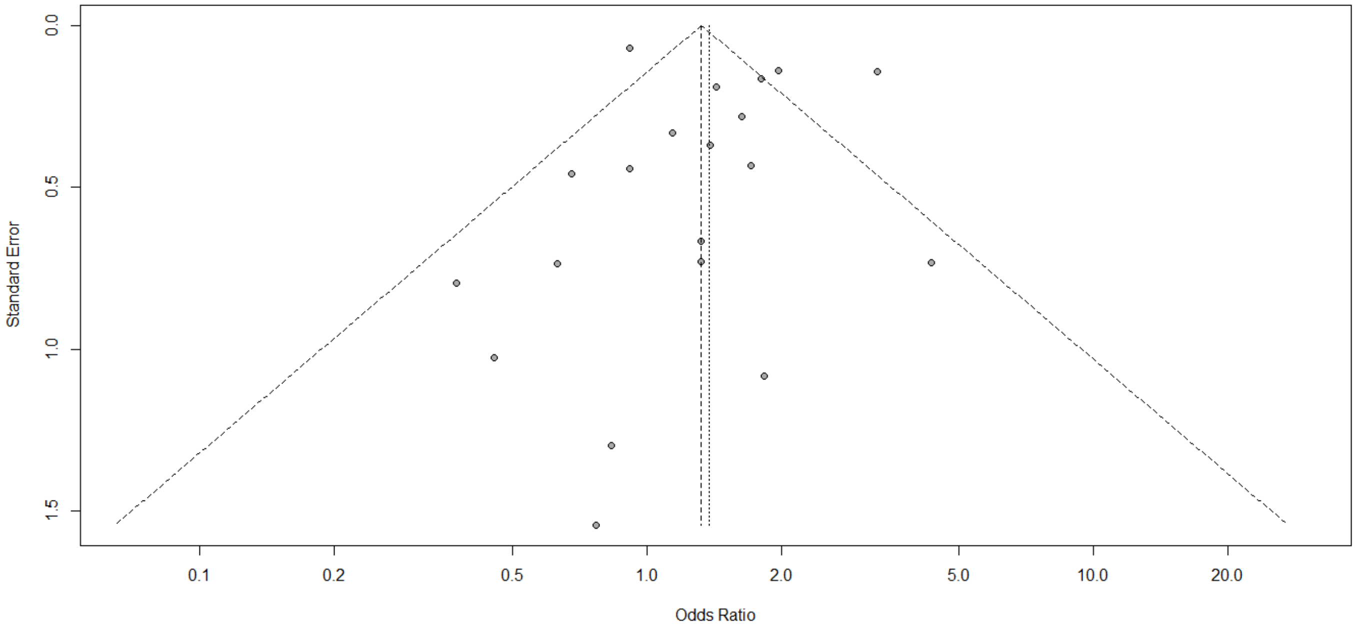


**Figure S43. Funnel plot for association between mortality outcomes in COVID-19 patients on an angiotensin-converting enzyme inhibitor.**

**A. Studies from the same cluster included**

| Study | ARB Events | ARB Total | Non-ARB Events | Non-ARB Total | OR | 95%-CI | Weight |
|---|---|---|---|---|---|---|---|
| Gao, China | 3 | 131 | 31 | 719 | 0.52 | [0.16; 1.73] | 2.0% |
| Li J, China | 15 | 83 | 62 | 279 | 0.77 | [0.41; 1.44] | 3.9% |
| Fosbol, Denmark | 84 | 630 | 256 | 3585 | 2.00 | [1.54; 2.60] | 5.7% |
| Cariou, France | 45 | 389 | 95 | 928 | 1.15 | [0.79; 1.67] | 5.2% |
| Ashraf, Iran | 2 | 17 | 10 | 65 | 0.73 | [0.14; 3.71] | 1.2% |
| Alberici, Italy | 1 | 3 | 4 | 17 | 1.62 | [0.11; 22.98] | 0.5% |
| Benelli, Italy | 14 | 60 | 58 | 351 | 1.54 | [0.79; 2.98] | 3.8% |
| Felice, Italy | 7 | 42 | 26 | 91 | 0.50 | [0.20; 1.27] | 2.7% |
| Giacomelli, Italy | 14 | 31 | 34 | 202 | 4.07 | [1.83; 9.03] | 3.2% |
| Giorgi Rossi, Italy | 52 | 368 | 165 | 2285 | 2.11 | [1.51; 2.95] | 5.4% |
| Iaccarino, Italy | 43 | 307 | 145 | 1284 | 1.28 | [0.89; 1.84] | 5.2% |
| Sardu, Italy | 3 | 21 | 6 | 41 | 0.97 | [0.22; 4.35] | 1.4% |
| Violi, Italy | 4 | 42 | 60 | 277 | 0.38 | [0.13; 1.11] | 2.3% |
| Garassino, Multiple | 7 | 25 | 57 | 163 | 0.72 | [0.29; 1.83] | 2.7% |
| Lee, South Korea | 48 | 941 | 64 | 7325 | 6.10 | [4.17; 8.92] | 5.2% |
| Jung C, Multiple | 43 | 95 | 104 | 229 | 0.99 | [0.61; 1.61] | 4.7% |
| Fernández−Ruiz, Spain | 0 | 3 | 5 | 15 | 0.27 | [0.01; 6.29] | 0.4% |
| Lopez−Otero, Spain | 9 | 134 | 29 | 831 | 1.99 | [0.92; 4.31] | 3.3% |
| Poblador−Plou, Spain | 81 | 356 | 690 | 4056 | 1.44 | [1.11; 1.87] | 5.7% |
| Senkal, Turkey | 5 | 52 | 7 | 104 | 1.47 | [0.44; 4.89] | 2.0% |
| Garibaldi, USA | 4 | 71 | 109 | 676 | 0.31 | [0.11; 0.87] | 2.4% |
| Gupta, USA | 157 | 365 | 627 | 1850 | 1.47 | [1.17; 1.85] | 5.9% |
| Ip, USA | 67 | 203 | 332 | 926 | 0.88 | [0.64; 1.22] | 5.5% |
| Khera, USA | 345 | 2226 | 785 | 5707 | 1.15 | [1.00; 1.32] | 6.2% |
| Kim, USA | 52 | 257 | 266 | 1638 | 1.31 | [0.94; 1.82] | 5.4% |
| Nguyen, USA | 7 | 41 | 23 | 252 | 2.05 | [0.82; 5.14] | 2.7% |
| Richardson, USA | 75 | 245 | 309 | 1121 | 1.16 | [0.86; 1.57] | 5.6% |
| **Random effects model** | | **7138** | | **35017** | **1.30** | **[1.07; 1.59]** | **100.0%** |

Heterogeneity: $I^2 = 80\%$, $\tau^2 = 0.1669$, $p < 0.01$

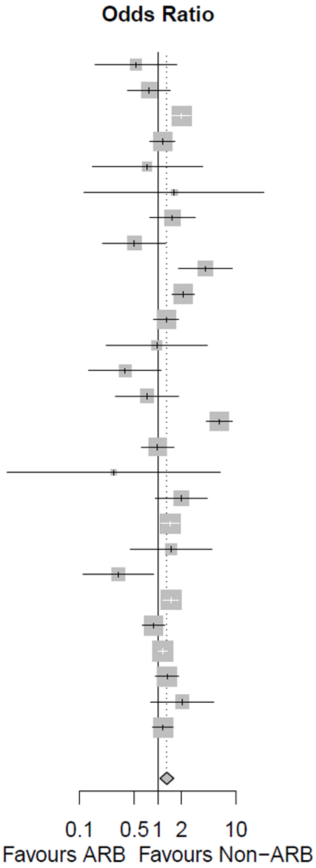


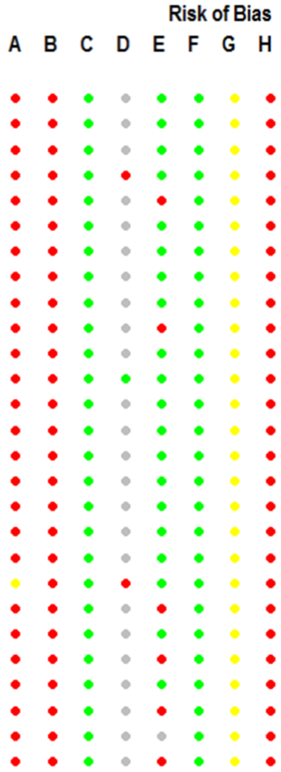


**B. Primary meta-analysis**

| Study | ARB Events | ARB Total | Non-ARB Events | Non-ARB Total | OR | 95%-CI | Weight |
|---|---|---|---|---|---|---|---|
| Gao, China | 3 | 131 | 31 | 719 | 0.52 | [0.16; 1.73] | 3.8% |
| Li J, China | 15 | 83 | 62 | 279 | 0.77 | [0.41; 1.44] | 6.7% |
| Fosbol, Denmark | 84 | 630 | 256 | 3585 | 2.00 | [1.54; 2.60] | 8.7% |
| Cariou, France | 45 | 389 | 95 | 928 | 1.15 | [0.79; 1.67] | 8.1% |
| Ashraf, Iran | 2 | 17 | 10 | 65 | 0.73 | [0.14; 3.71] | 2.5% |
| Alberici, Italy | 1 | 3 | 4 | 17 | 1.62 | [0.11; 22.98] | 1.2% |
| Benelli, Italy | 14 | 60 | 58 | 351 | 1.54 | [0.79; 2.98] | 6.5% |
| Felice, Italy | 7 | 42 | 26 | 91 | 0.50 | [0.20; 1.27] | 5.0% |
| Giacomelli, Italy | 14 | 31 | 34 | 202 | 4.07 | [1.83; 9.03] | 5.7% |
| Giorgi Rossi, Italy | 52 | 368 | 165 | 2285 | 2.11 | [1.51; 2.95] | 8.4% |
| Sardu, Italy | 3 | 21 | 6 | 41 | 0.97 | [0.22; 4.35] | 2.8% |
| Violi, Italy | 4 | 42 | 60 | 277 | 0.38 | [0.13; 1.11] | 4.3% |
| Lee, South Korea | 48 | 941 | 64 | 7325 | 6.10 | [4.17; 8.92] | 8.1% |
| Fernández-Ruiz, Spain | 0 | 3 | 5 | 15 | 0.27 | [0.01; 6.29] | 0.9% |
| Lopez-Otero, Spain | 9 | 134 | 29 | 831 | 1.99 | [0.92; 4.31] | 5.8% |
| Poblador-Plou, Spain | 81 | 356 | 690 | 4056 | 1.44 | [1.11; 1.87] | 8.7% |
| Senkal, Turkey | 5 | 52 | 7 | 104 | 1.47 | [0.44; 4.89] | 3.8% |
| Khera, USA | 345 | 2226 | 785 | 5707 | 1.15 | [1.00; 1.32] | 9.1% |
| **Random effects model** | | **5529** | | **26878** | **1.43** | **[1.05; 1.93]** | **100.0%** |

Heterogeneity: $I^2 = 84\%$, $\tau^2 = 0.2578$, $p < 0.01$

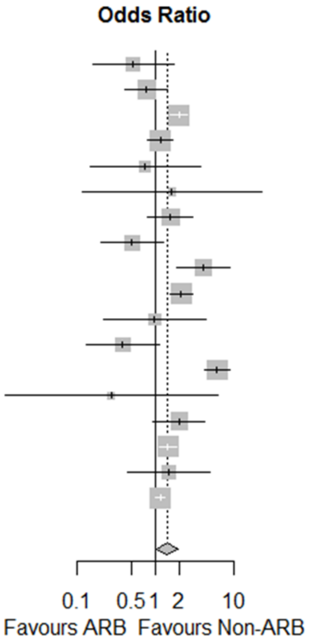


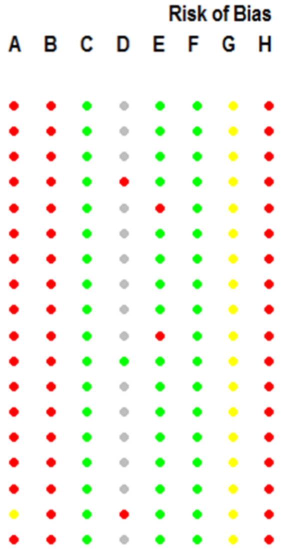


**Figure S44. Forest plots for association between mortality outcomes in COVID-19 patients on an angiotensin receptor blocker (ARB).** Risk of bias legend. A = risk of bias due to confounding, B = risk of bias in selection of participants into the study, C = risk of bias in classification of interventions, D = risk of bias due to deviations from intended interventions, E = risk of bias due to missing data, F = risk of bias in measurement of outcomes, G = risk of bias in selection of the reported result, H = overall risk of bias. Color codes. Colour codes. Red = serious, yellow = moderate, green = low, grey = unclear.

**C. Hypertensive-only cohorts**

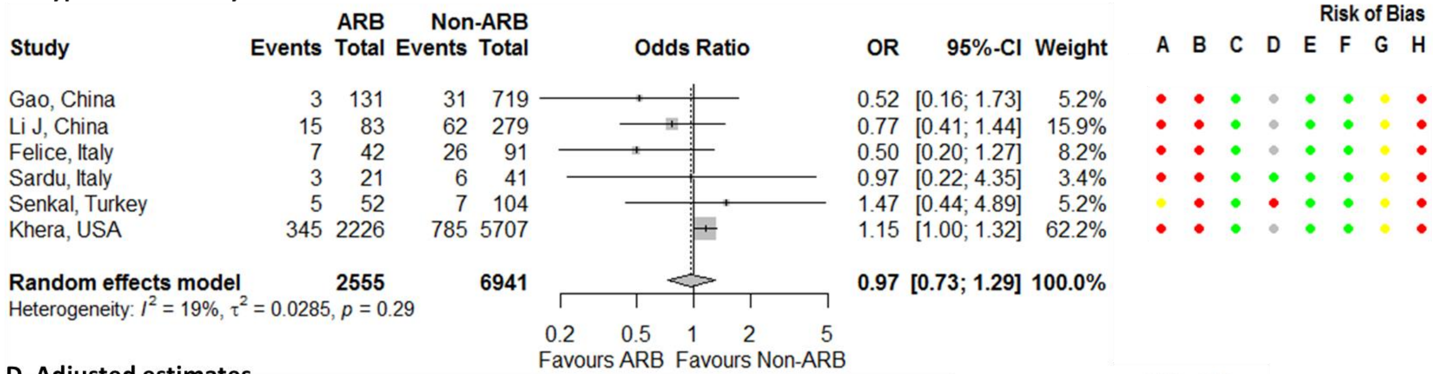


**D. Adjusted estimates**

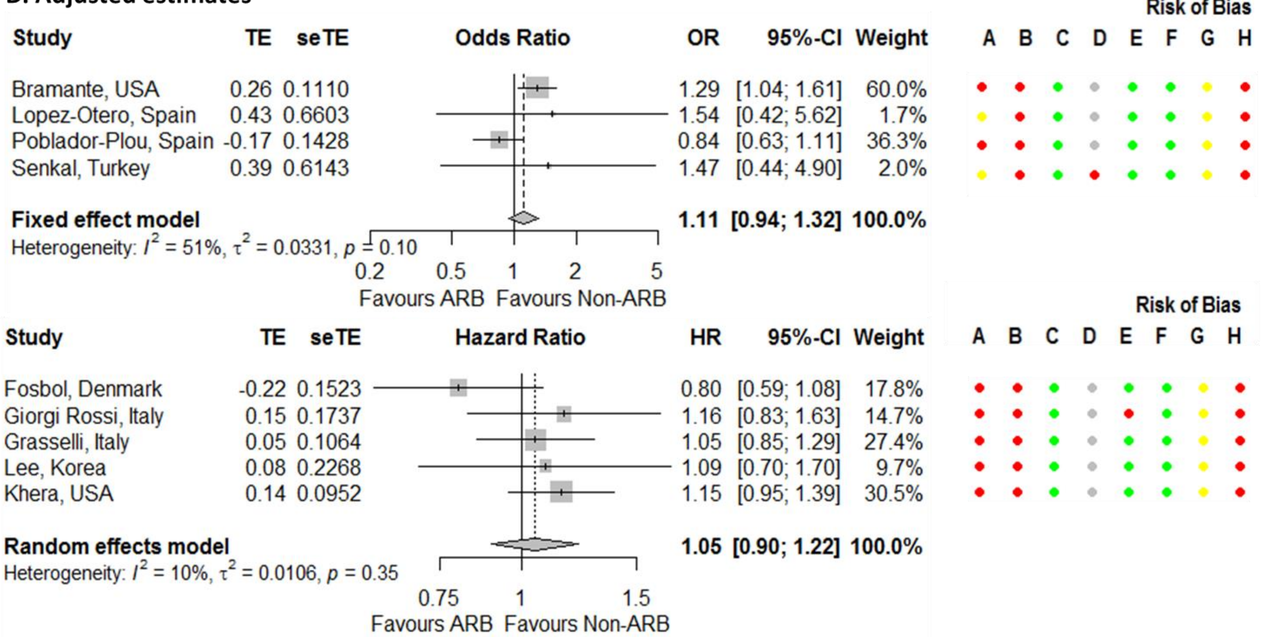


**Figure S44. Continued.**

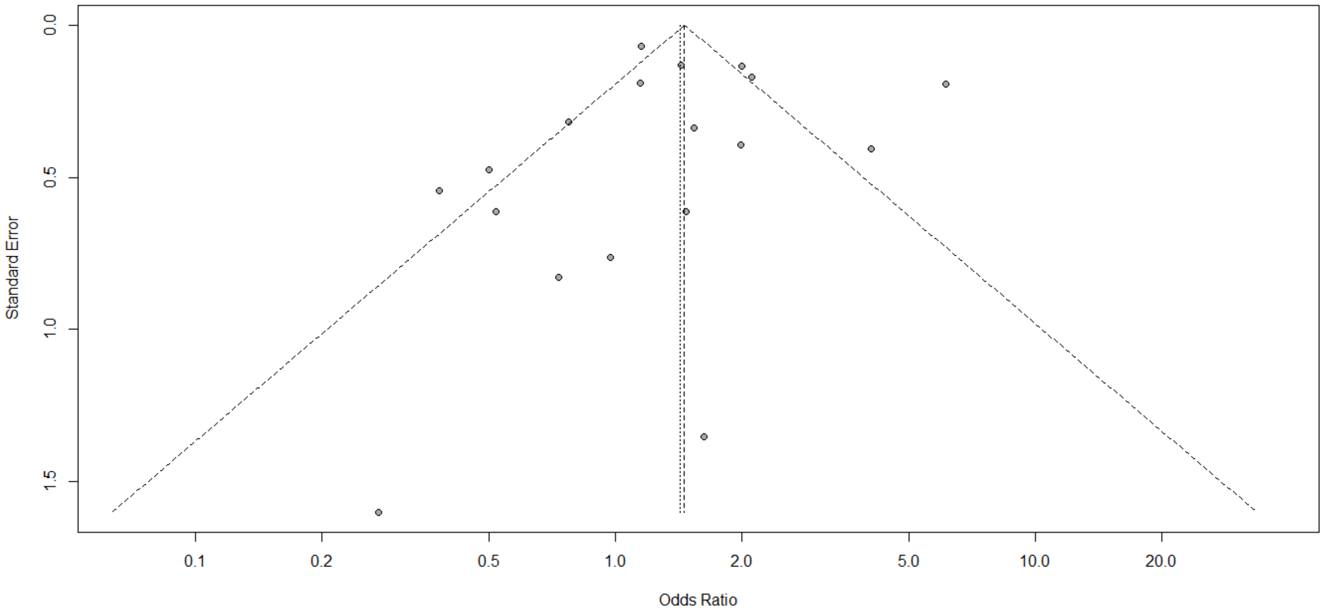


**Figure S45. Funnel plot for association between mortality outcomes in COVID-19 patients on an angiotensin receptor blocker.**

**A. Studies from the same cluster included**

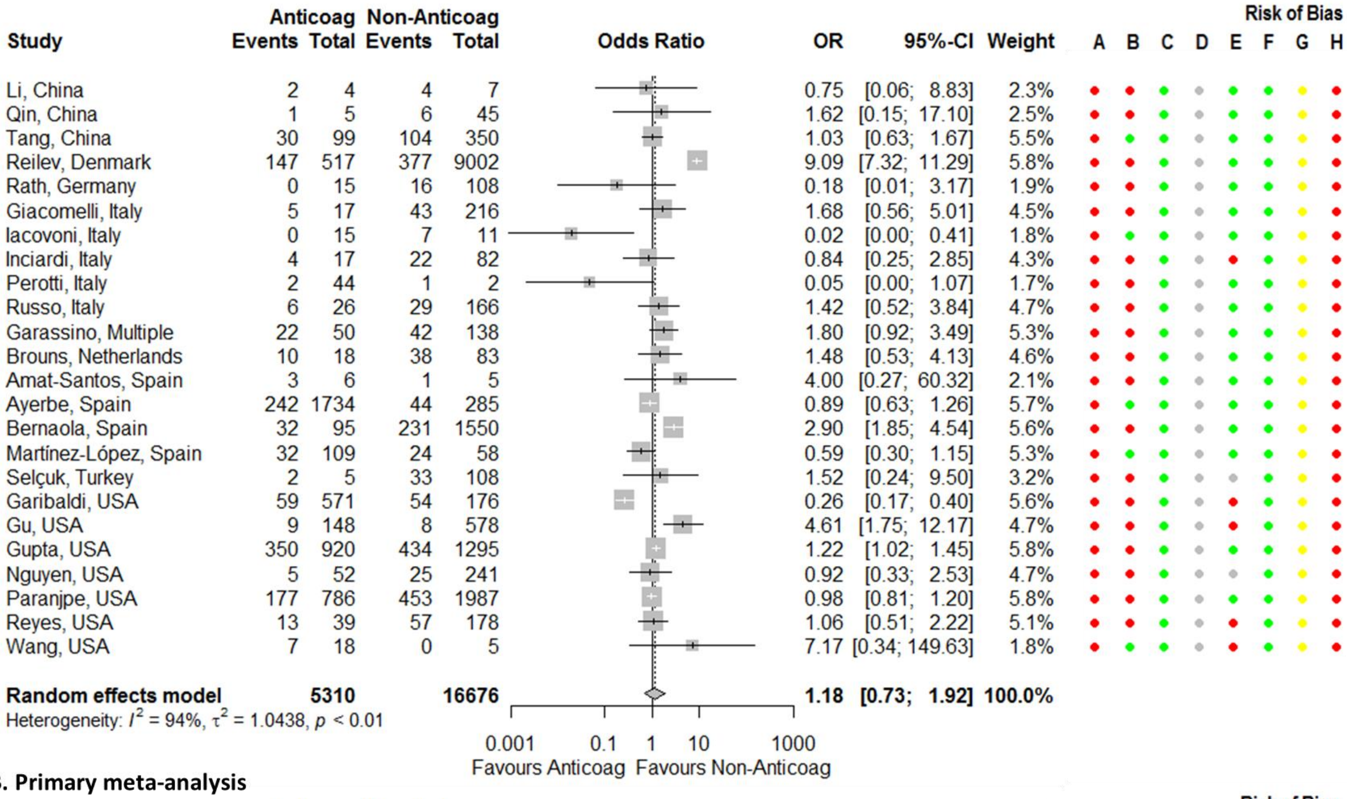

| Study | Anticoag Events | Anticoag Total | Non-Anticoag Events | Non-Anticoag Total | OR | 95%-CI | Weight |
|---|---|---|---|---|---|---|---|
| Li, China | 2 | 4 | 4 | 7 | 0.75 | [0.06; 8.83] | 2.3% |
| Qin, China | 1 | 5 | 6 | 45 | 1.62 | [0.15; 17.10] | 2.5% |
| Tang, China | 30 | 99 | 104 | 350 | 1.03 | [0.63; 1.67] | 5.5% |
| Reilev, Denmark | 147 | 517 | 377 | 9002 | 9.09 | [7.32; 11.29] | 5.8% |
| Rath, Germany | 0 | 15 | 16 | 108 | 0.18 | [0.01; 3.17] | 1.9% |
| Giacomelli, Italy | 5 | 17 | 43 | 216 | 1.68 | [0.56; 5.01] | 4.5% |
| Iacovoni, Italy | 0 | 15 | 7 | 11 | 0.02 | [0.00; 0.41] | 1.8% |
| Inciardi, Italy | 4 | 17 | 22 | 82 | 0.84 | [0.25; 2.85] | 4.3% |
| Perotti, Italy | 2 | 44 | 1 | 2 | 0.05 | [0.00; 1.07] | 1.7% |
| Russo, Italy | 6 | 26 | 29 | 166 | 1.42 | [0.52; 3.84] | 4.7% |
| Garassino, Multiple | 22 | 50 | 42 | 138 | 1.80 | [0.92; 3.49] | 5.3% |
| Brouns, Netherlands | 10 | 18 | 38 | 83 | 1.48 | [0.53; 4.13] | 4.6% |
| Amat-Santos, Spain | 3 | 6 | 1 | 5 | 4.00 | [0.27; 60.32] | 2.1% |
| Ayerbe, Spain | 242 | 1734 | 44 | 285 | 0.89 | [0.63; 1.26] | 5.7% |
| Bernaola, Spain | 32 | 95 | 231 | 1550 | 2.90 | [1.85; 4.54] | 5.6% |
| Martínez-López, Spain | 32 | 109 | 24 | 58 | 0.59 | [0.30; 1.15] | 5.3% |
| Selçuk, Turkey | 2 | 5 | 33 | 108 | 1.52 | [0.24; 9.50] | 3.2% |
| Garibaldi, USA | 59 | 571 | 54 | 176 | 0.26 | [0.17; 0.40] | 5.6% |
| Gu, USA | 9 | 148 | 8 | 578 | 4.61 | [1.75; 12.17] | 4.7% |
| Gupta, USA | 350 | 920 | 434 | 1295 | 1.22 | [1.02; 1.45] | 5.8% |
| Nguyen, USA | 5 | 52 | 25 | 241 | 0.92 | [0.33; 2.53] | 4.7% |
| Paranjpe, USA | 177 | 786 | 453 | 1987 | 0.98 | [0.81; 1.20] | 5.8% |
| Reyes, USA | 13 | 39 | 57 | 178 | 1.06 | [0.51; 2.22] | 5.1% |
| Wang, USA | 7 | 18 | 0 | 5 | 7.17 | [0.34; 149.63] | 1.8% |
| **Random effects model** | | **5310** | | **16676** | **1.18** | **[0.73; 1.92]** | **100.0%** |

Heterogeneity: $I^2 = 94\%$, $\tau^2 = 1.0438$, $p < 0.01$



**B. Primary meta-analysis**

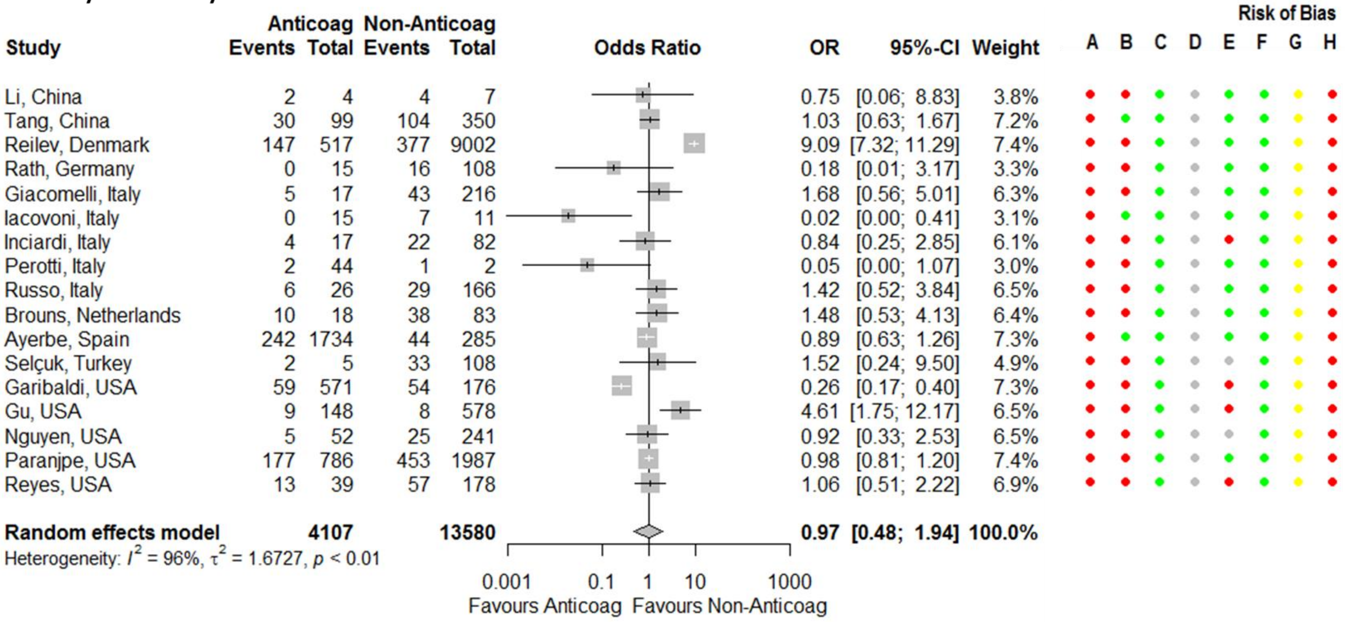

| Study | Anticoag Events | Anticoag Total | Non-Anticoag Events | Non-Anticoag Total | OR | 95%-CI | Weight |
|---|---|---|---|---|---|---|---|
| Li, China | 2 | 4 | 4 | 7 | 0.75 | [0.06; 8.83] | 3.8% |
| Tang, China | 30 | 99 | 104 | 350 | 1.03 | [0.63; 1.67] | 7.2% |
| Reilev, Denmark | 147 | 517 | 377 | 9002 | 9.09 | [7.32; 11.29] | 7.4% |
| Rath, Germany | 0 | 15 | 16 | 108 | 0.18 | [0.01; 3.17] | 3.3% |
| Giacomelli, Italy | 5 | 17 | 43 | 216 | 1.68 | [0.56; 5.01] | 6.3% |
| Iacovoni, Italy | 0 | 15 | 7 | 11 | 0.02 | [0.00; 0.41] | 3.1% |
| Inciardi, Italy | 4 | 17 | 22 | 82 | 0.84 | [0.25; 2.85] | 6.1% |
| Perotti, Italy | 2 | 44 | 1 | 2 | 0.05 | [0.00; 1.07] | 3.0% |
| Russo, Italy | 6 | 26 | 29 | 166 | 1.42 | [0.52; 3.84] | 6.5% |
| Brouns, Netherlands | 10 | 18 | 38 | 83 | 1.48 | [0.53; 4.13] | 6.4% |
| Ayerbe, Spain | 242 | 1734 | 44 | 285 | 0.89 | [0.63; 1.26] | 7.3% |
| Selçuk, Turkey | 2 | 5 | 33 | 108 | 1.52 | [0.24; 9.50] | 4.9% |
| Garibaldi, USA | 59 | 571 | 54 | 176 | 0.26 | [0.17; 0.40] | 7.3% |
| Gu, USA | 9 | 148 | 8 | 578 | 4.61 | [1.75; 12.17] | 6.5% |
| Nguyen, USA | 5 | 52 | 25 | 241 | 0.92 | [0.33; 2.53] | 6.5% |
| Paranjpe, USA | 177 | 786 | 453 | 1987 | 0.98 | [0.81; 1.20] | 7.4% |
| Reyes, USA | 13 | 39 | 57 | 178 | 1.06 | [0.51; 2.22] | 6.9% |
| **Random effects model** | | **4107** | | **13580** | **0.97** | **[0.48; 1.94]** | **100.0%** |

Heterogeneity: $I^2 = 96\%$, $\tau^2 = 1.6727$, $p < 0.01$



**Figure S46. Forest plots for association between mortality outcomes in COVID-19 patients on any form of anticoagulation (prophylactic and/or therapeutic).** Risk of bias legend. A = risk of bias due to confounding, B = risk of bias in selection of participants into the study, C = risk of bias in classification of interventions, D = risk of bias due to deviations from intended interventions, E = risk of bias due to missing data, F = risk of bias in measurement of outcomes, G = risk of bias in selection of the reported result, H = overall risk of bias. Color codes. Colour codes. Red = serious, yellow = moderate, green = low, grey = unclear.

**C. Hospitalized patients only**

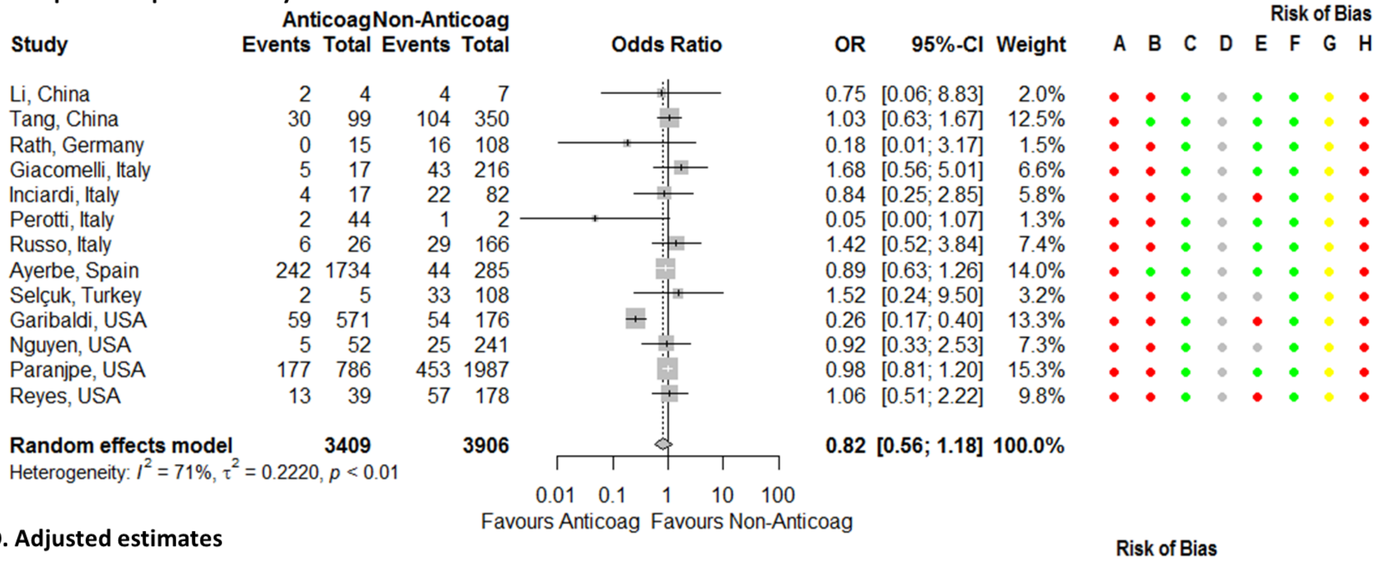


**D. Adjusted estimates**

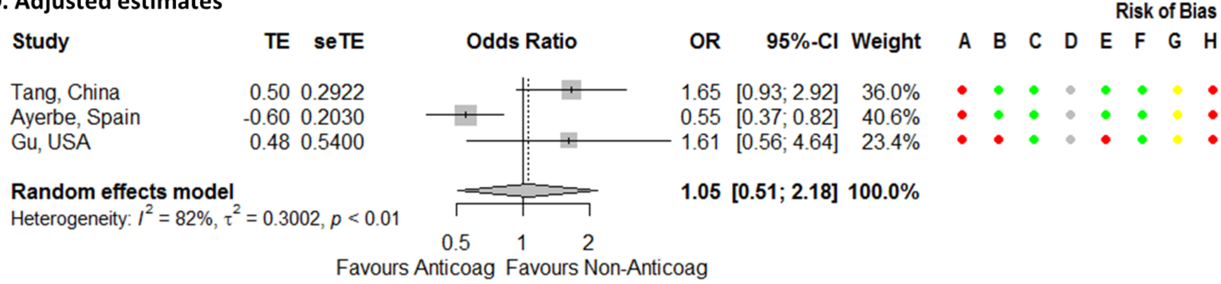


**Figure S46. Continued.**

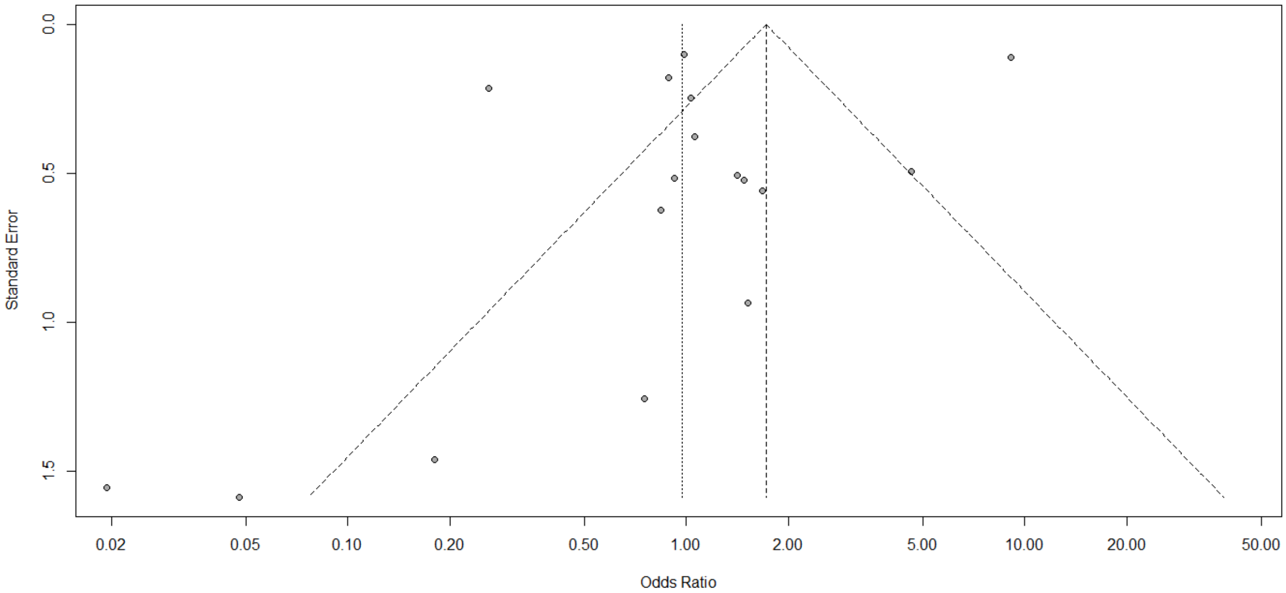


**Figure S47. Funnel plot for association between mortality outcomes in COVID-19 patients on anticoagulants.**

**A. Studies from the same cluster included**

| Study | Antiplatelets Events | Antiplatelets Total | Non-Antiplatelets Events | Non-Antiplatelets Total | OR | 95%-CI | Weight |
|---|---|---|---|---|---|---|---|
| Li, China | 3 | 6 | 3 | 5 | 0.67 | [0.06; 7.35] | 2.9% |
| Qin, China | 1 | 14 | 6 | 36 | 0.38 | [0.04; 3.52] | 3.2% |
| Reilev, Denmark | 180 | 777 | 344 | 8742 | 7.36 | [6.04; 8.98] | 8.9% |
| Rath, Germany | 3 | 27 | 13 | 96 | 0.80 | [0.21; 3.03] | 5.4% |
| Giacomelli, Italy | 13 | 33 | 35 | 200 | 3.06 | [1.39; 6.74] | 7.3% |
| Russo, Italy | 10 | 55 | 25 | 137 | 1.00 | [0.44; 2.24] | 7.2% |
| Viecca, Italy | 1 | 5 | 3 | 5 | 0.17 | [0.01; 2.82] | 2.3% |
| Violi, Italy | 15 | 46 | 49 | 273 | 2.21 | [1.11; 4.41] | 7.6% |
| Ayed, Kuwait | 6 | 7 | 39 | 96 | 8.77 | [1.02; 75.72] | 3.3% |
| Garassino, Multiple | 8 | 37 | 56 | 151 | 0.47 | [0.20; 1.09] | 7.1% |
| Brouns, Netherlands | 14 | 34 | 34 | 67 | 0.68 | [0.30; 1.56] | 7.2% |
| Bernaola, Spain | 48 | 149 | 215 | 1496 | 2.83 | [1.95; 4.11] | 8.6% |
| Selçuk, Turkey | 16 | 35 | 19 | 78 | 2.61 | [1.13; 6.07] | 7.1% |
| Gupta, USA | 234 | 491 | 550 | 1724 | 1.94 | [1.58; 2.38] | 8.9% |
| Nguyen, USA | 10 | 69 | 20 | 224 | 1.73 | [0.77; 3.90] | 7.2% |
| Wang, USA | 8 | 34 | 6 | 24 | 0.92 | [0.27; 3.12] | 5.8% |
| **Random effects model** | | **1819** | | **13354** | **1.59** | **[0.97; 2.61]** | **100.0%** |

Heterogeneity: $I^2 = 90\%$, $\tau^2 = 0.7066$, $p < 0.01$

Odds Ratio axis: 0.01, 0.1, 1, 10, 100 — Favours Antiplatelets / Favours Non-Antiplatelets

Risk of Bias: A B C D E F G H

**B. Primary meta-analysis**

| Study | Antiplatelets Events | Antiplatelets Total | Non-Antiplatelets Events | Non-Antiplatelets Total | OR | 95%-CI | Weight |
|---|---|---|---|---|---|---|---|
| Li, China | 3 | 6 | 3 | 5 | 0.67 | [0.06; 7.35] | 3.5% |
| Qin, China | 1 | 14 | 6 | 36 | 0.38 | [0.04; 3.52] | 3.9% |
| Reilev, Denmark | 180 | 777 | 344 | 8742 | 7.36 | [6.04; 8.98] | 11.3% |
| Rath, Germany | 3 | 27 | 13 | 96 | 0.80 | [0.21; 3.03] | 6.7% |
| Giacomelli, Italy | 13 | 33 | 35 | 200 | 3.06 | [1.39; 6.74] | 9.2% |
| Russo, Italy | 10 | 55 | 25 | 137 | 1.00 | [0.44; 2.24] | 9.1% |
| Viecca, Italy | 1 | 5 | 3 | 5 | 0.17 | [0.01; 2.82] | 2.7% |
| Violi, Italy | 15 | 46 | 49 | 273 | 2.21 | [1.11; 4.41] | 9.6% |
| Ayed, Kuwait | 6 | 7 | 39 | 96 | 8.77 | [1.02; 75.72] | 4.0% |
| Brouns, Netherlands | 14 | 34 | 34 | 67 | 0.68 | [0.30; 1.56] | 9.0% |
| Bernaola, Spain | 48 | 149 | 215 | 1496 | 2.83 | [1.95; 4.11] | 10.9% |
| Selçuk, Turkey | 16 | 35 | 19 | 78 | 2.61 | [1.13; 6.07] | 8.9% |
| Gupta, USA | 234 | 491 | 550 | 1724 | 1.94 | [1.58; 2.38] | 11.3% |
| **Random effects model** | | **1679** | | **12955** | **1.85** | **[1.08; 3.16]** | **100.0%** |

Heterogeneity: $I^2 = 91\%$, $\tau^2 = 0.6488$, $p < 0.01$

Odds Ratio axis: 0.01, 0.1, 1, 10, 100 — Favours Antiplatelets / Favours Non-Antiplatelets

Risk of Bias: A B C D E F G H

**C. Peer-reviewed studies**

| Study | Antiplatelets Events | Antiplatelets Total | Non-Antiplatelets Events | Non-Antiplatelets Total | OR | 95%-CI | Weight |
|---|---|---|---|---|---|---|---|
| Li, China | 3 | 6 | 3 | 5 | 0.67 | [0.06; 7.35] | 2.3% |
| Qin, China | 1 | 14 | 6 | 36 | 0.38 | [0.04; 3.52] | 2.7% |
| Rath, Germany | 3 | 27 | 13 | 96 | 0.80 | [0.21; 3.03] | 6.3% |
| Giacomelli, Italy | 13 | 33 | 35 | 200 | 3.06 | [1.39; 6.74] | 12.5% |
| Russo, Italy | 10 | 55 | 25 | 137 | 1.00 | [0.44; 2.24] | 12.1% |
| Viecca, Italy | 1 | 5 | 3 | 5 | 0.17 | [0.01; 2.82] | 1.7% |
| Violi, Italy | 15 | 46 | 49 | 273 | 2.21 | [1.11; 4.41] | 14.3% |
| Brouns, Netherlands | 14 | 34 | 34 | 67 | 0.68 | [0.30; 1.56] | 11.7% |
| Selçuk, Turkey | 16 | 35 | 19 | 78 | 2.61 | [1.13; 6.07] | 11.6% |
| Gupta, USA | 234 | 491 | 550 | 1724 | 1.94 | [1.58; 2.38] | 24.9% |
| **Random effects model** | | **746** | | **2621** | **1.50** | **[1.02; 2.19]** | **100.0%** |

Heterogeneity: $I^2 = 48\%$, $\tau^2 = 0.1415$, $p = 0.04$

Odds Ratio axis: 0.01, 0.1, 1, 10, 100 — Favours Antiplatelets / Favours Non-Antiplatelets

Risk of Bias: A B C D E F G H

**Figure S48. Forest plots for association between mortality outcomes in COVID-19 patients on an antiplatelet.** Risk of bias legend. A = risk of bias due to confounding, B = risk of bias in selection of participants into the study, C = risk of bias in classification of interventions, D = risk of bias due to deviations from intended interventions, E = risk of bias due to missing data, F = risk of bias in measurement of outcomes, G = risk of bias in selection of the reported result, H = overall risk of bias. Color codes. Colour codes. Red = serious, yellow = moderate, green = low, grey = unclear.

**D. Preprints**

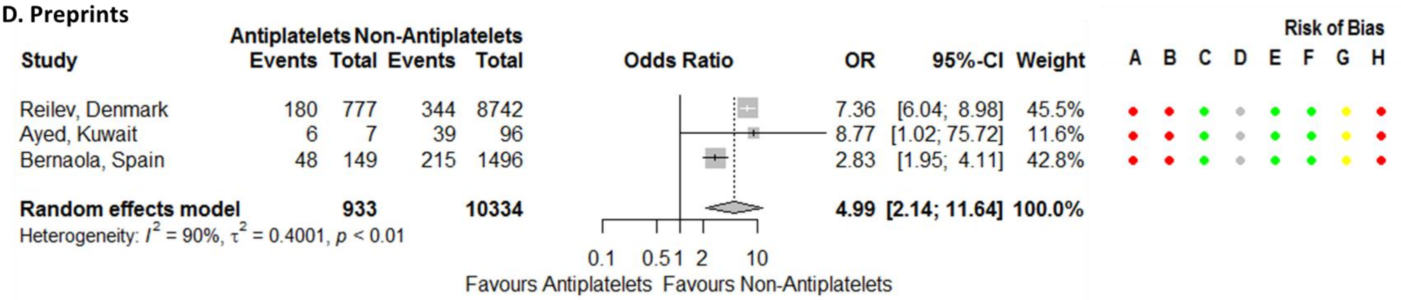

| Study | Antiplatelets Events | Antiplatelets Total | Non-Antiplatelets Events | Non-Antiplatelets Total | OR | 95%-CI | Weight | A | B | C | D | E | F | G | H |
|---|---|---|---|---|---|---|---|---|---|---|---|---|---|---|---|
| Reilev, Denmark | 180 | 777 | 344 | 8742 | 7.36 | [6.04; 8.98] | 45.5% | ● | ● | ● | ● | ● | ● | ● | ● |
| Ayed, Kuwait | 6 | 7 | 39 | 96 | 8.77 | [1.02; 75.72] | 11.6% | ● | ● | ● | ● | ● | ● | ● | ● |
| Bernaola, Spain | 48 | 149 | 215 | 1496 | 2.83 | [1.95; 4.11] | 42.8% | ● | ● | ● | ● | ● | ● | ● | ● |
| **Random effects model** | | **933** | | **10334** | **4.99** | **[2.14; 11.64]** | **100.0%** | | | | | | | | |

Heterogeneity: $I^2 = 90\%$, $\tau^2 = 0.4001$, $p < 0.01$



**Figure S48. Continued.**

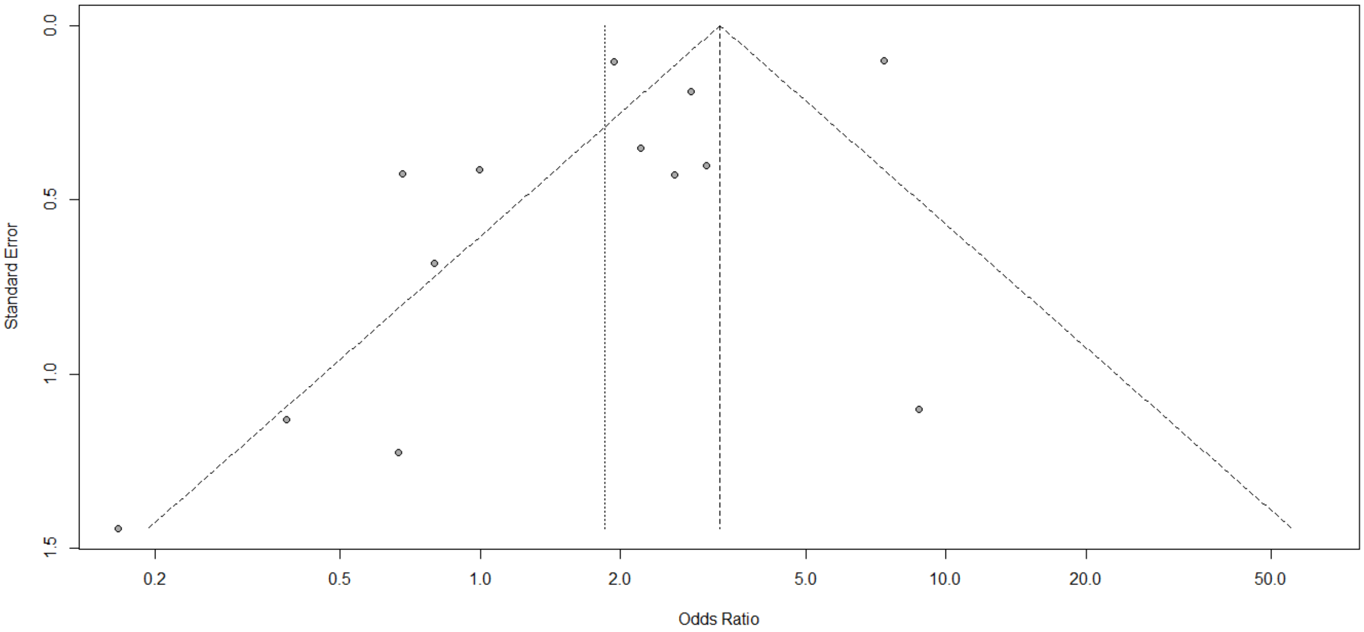


**Figure S49. Funnel plot for association between mortality outcomes in COVID-19 patients on antiplatelets.**

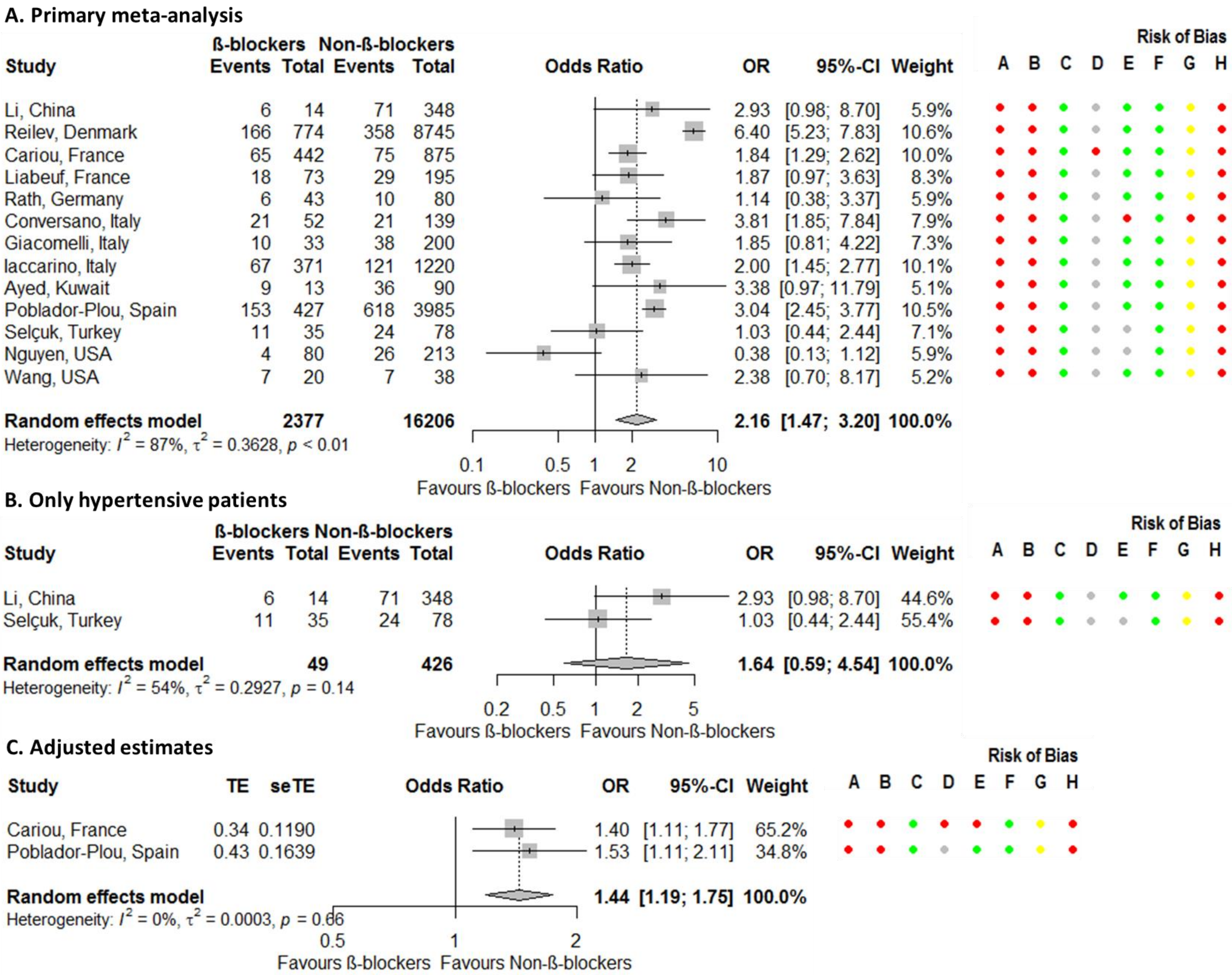


**Figure S50. Forest plots for association between mortality outcomes in COVID-19 patients on a beta-blocker.** Risk of bias legend. A = risk of bias due to confounding, B = risk of bias in selection of participants into the study, C = risk of bias in classification of interventions, D = risk of bias due to deviations from intended interventions, E = risk of bias due to missing data, F = risk of bias in measurement of outcomes, G = risk of bias in selection of the reported result, H = overall risk of bias. Color codes. Colour codes. Red = serious, yellow = moderate, green = low, grey = unclear.

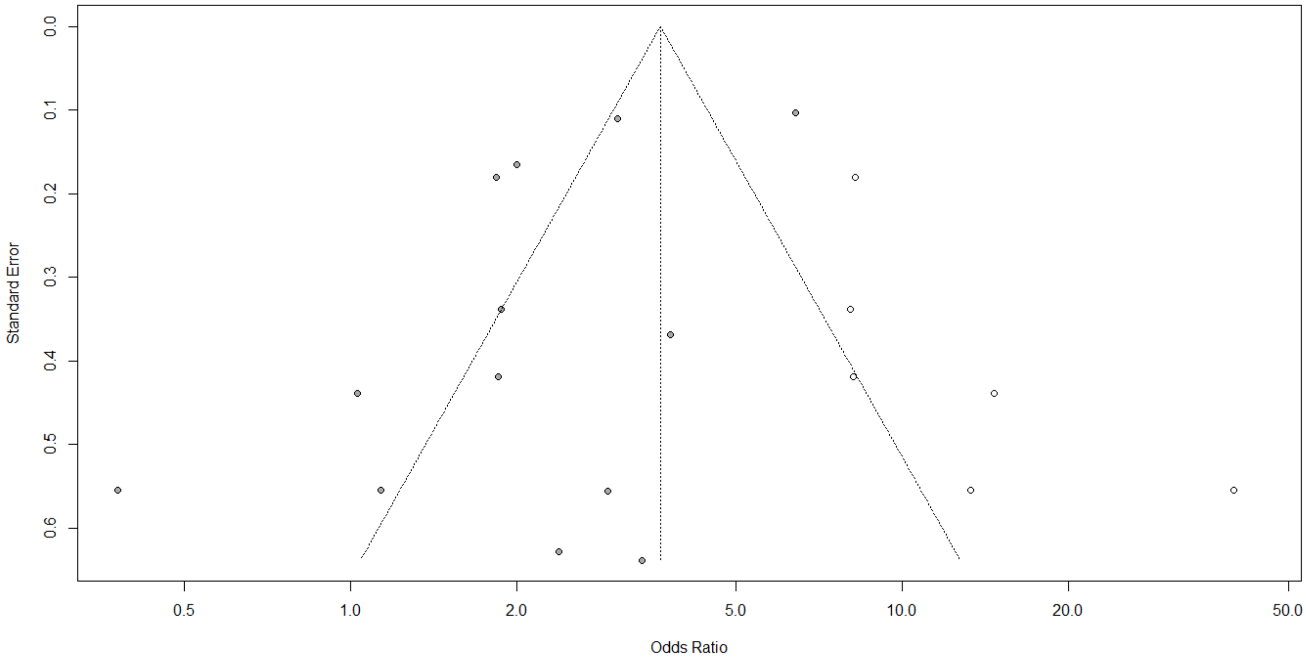


**Figure S51. Funnel plot for association between mortality outcomes in COVID-19 patients on a beta-blocker.** Solid dots represent the 13 studies included in the primary meta-analysis whereas the open dots, the 6 'filled' studies.

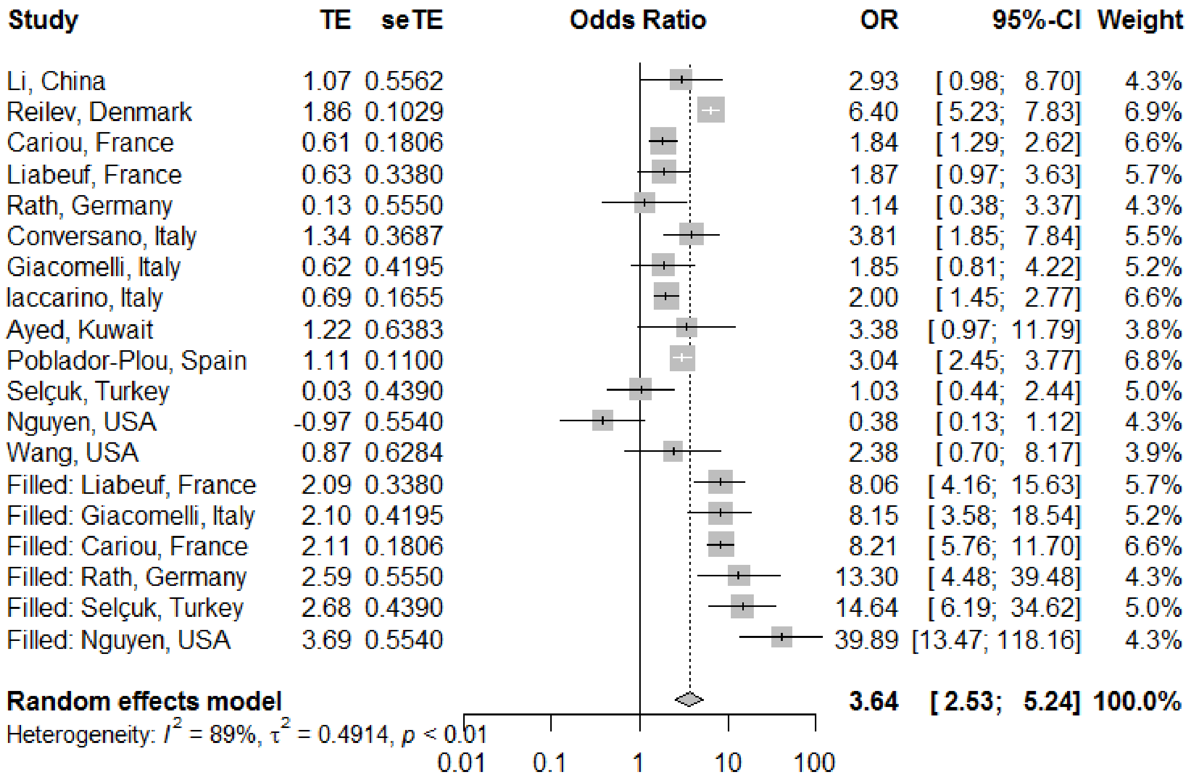


**Figure S52. Forest plot for the association between mortality outcomes in COVID-19 patients on a beta-blocker after adjusting for publication bias.**

**A. Studies from the same cluster included**

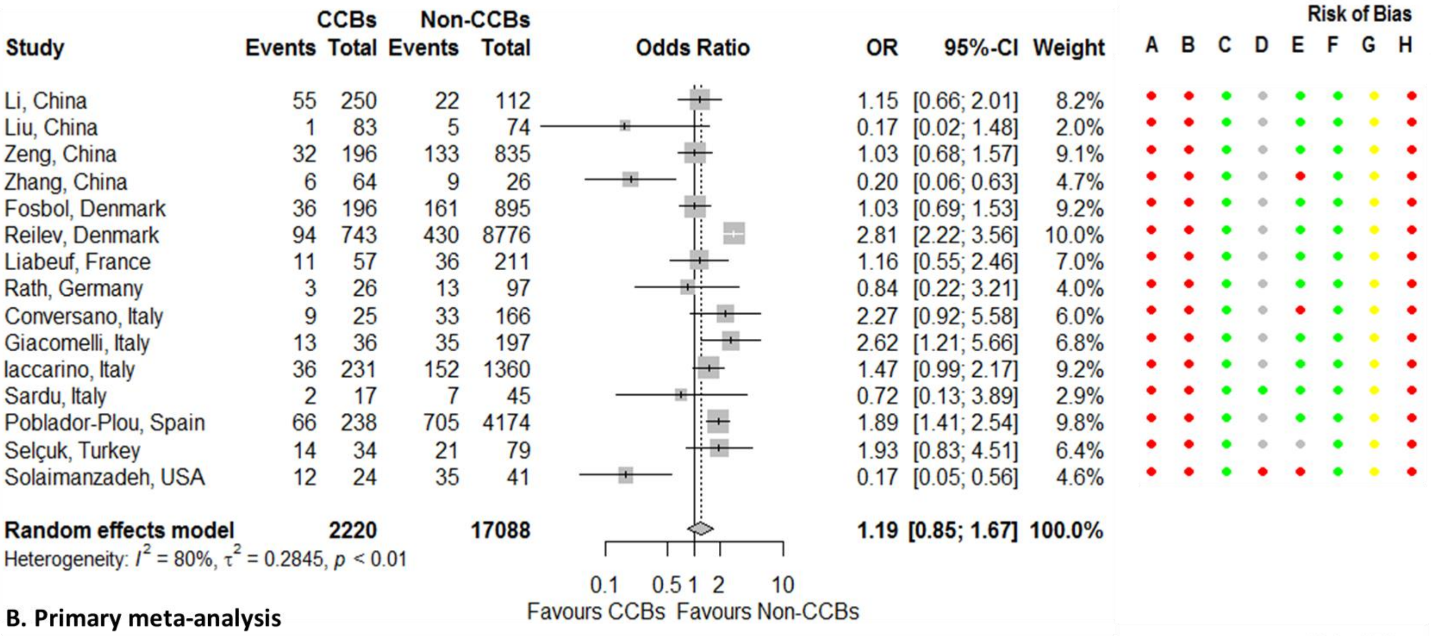


**B. Primary meta-analysis**

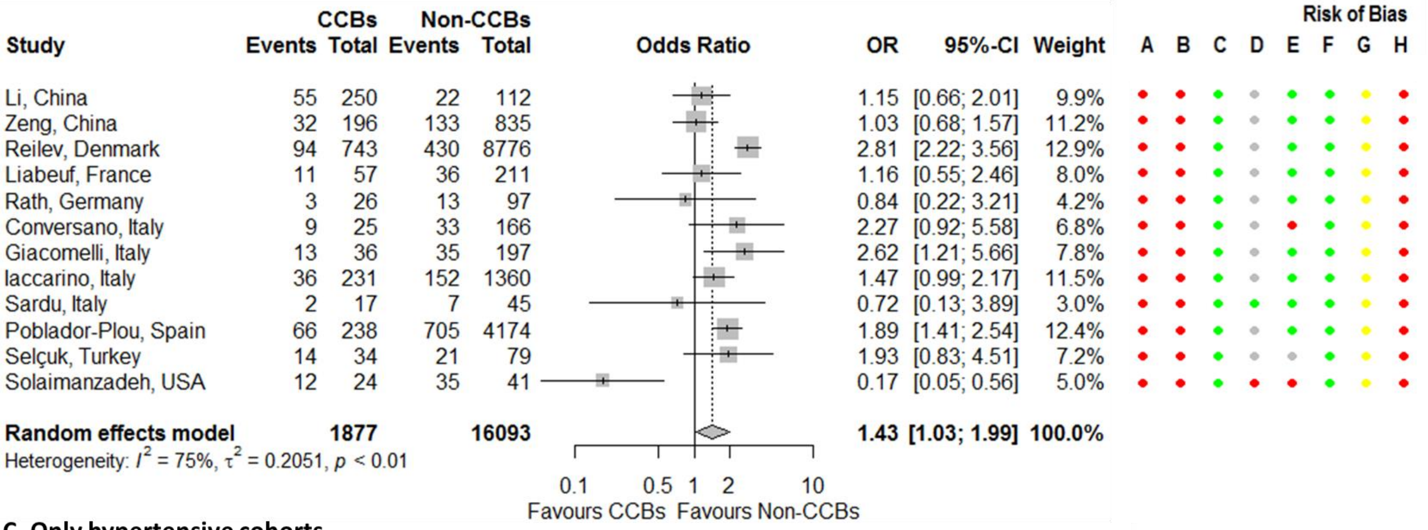


**C. Only hypertensive cohorts**

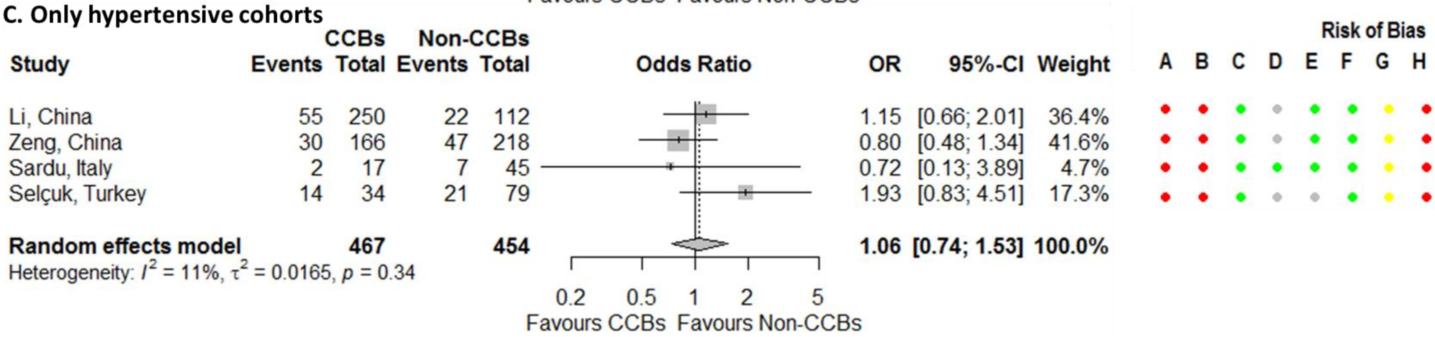


**Figure S53. Forest plots for association between mortality outcomes in COVID-19 patients on a calcium channel blocker.** Risk of bias legend. A = risk of bias due to confounding, B = risk of bias in selection of participants into the study, C = risk of bias in classification of interventions, D = risk of bias due to deviations from intended interventions, E = risk of bias due to missing data, F = risk of bias in measurement of outcomes, G = risk of bias in selection of the reported result, H = overall risk of bias. Color codes. Colour codes. Red = serious, yellow = moderate, green = low, grey = unclear.

**D. Adjusted estimates**

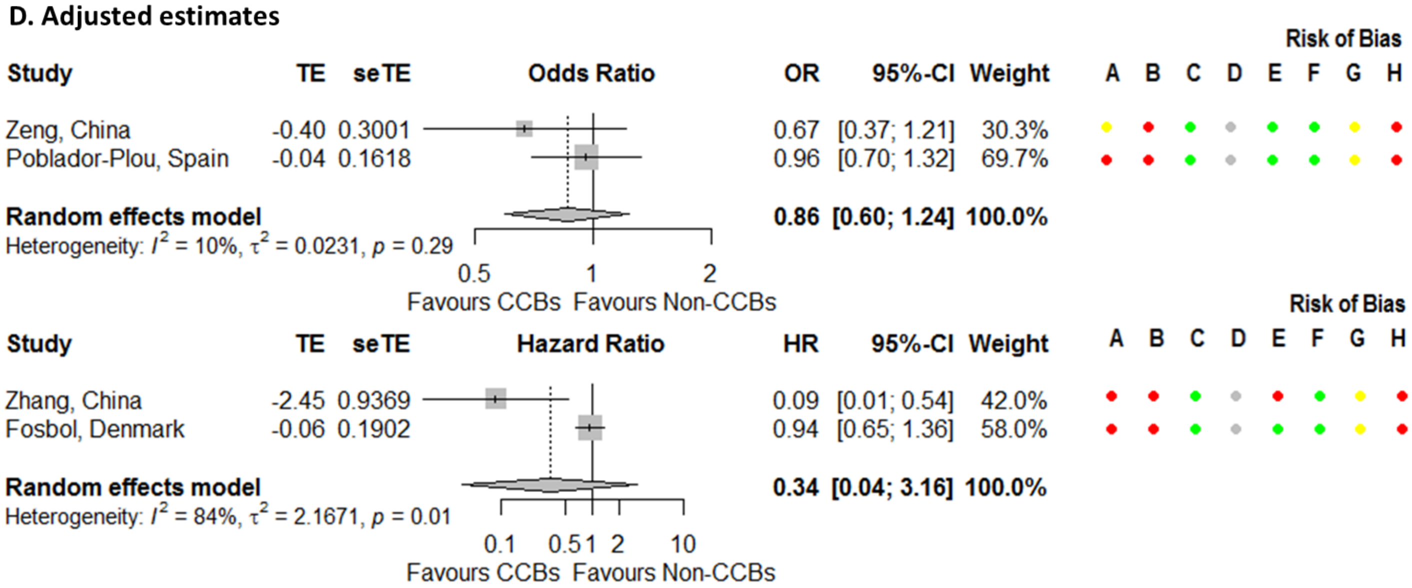


**Figure S53. Continued.**

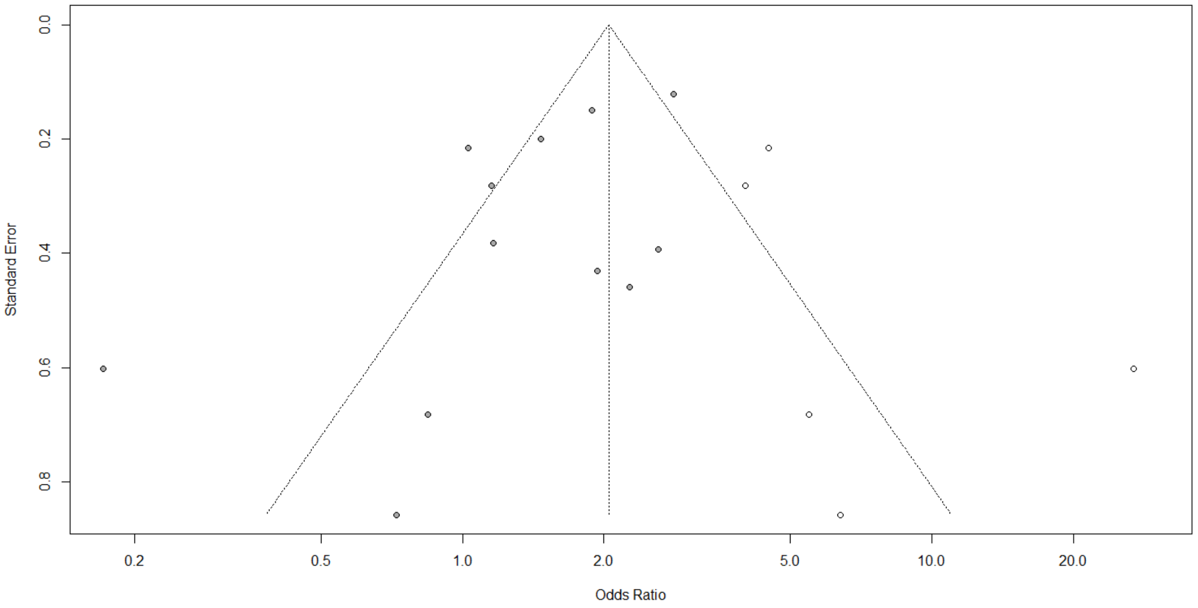


**Figure S54. Funnel plot for association between mortality outcomes in COVID-19 patients on a calcium channel blocker.** Solid dots represent the 12 studies included in the primary meta-analysis whereas the open dots, the 5 'filled' studies.

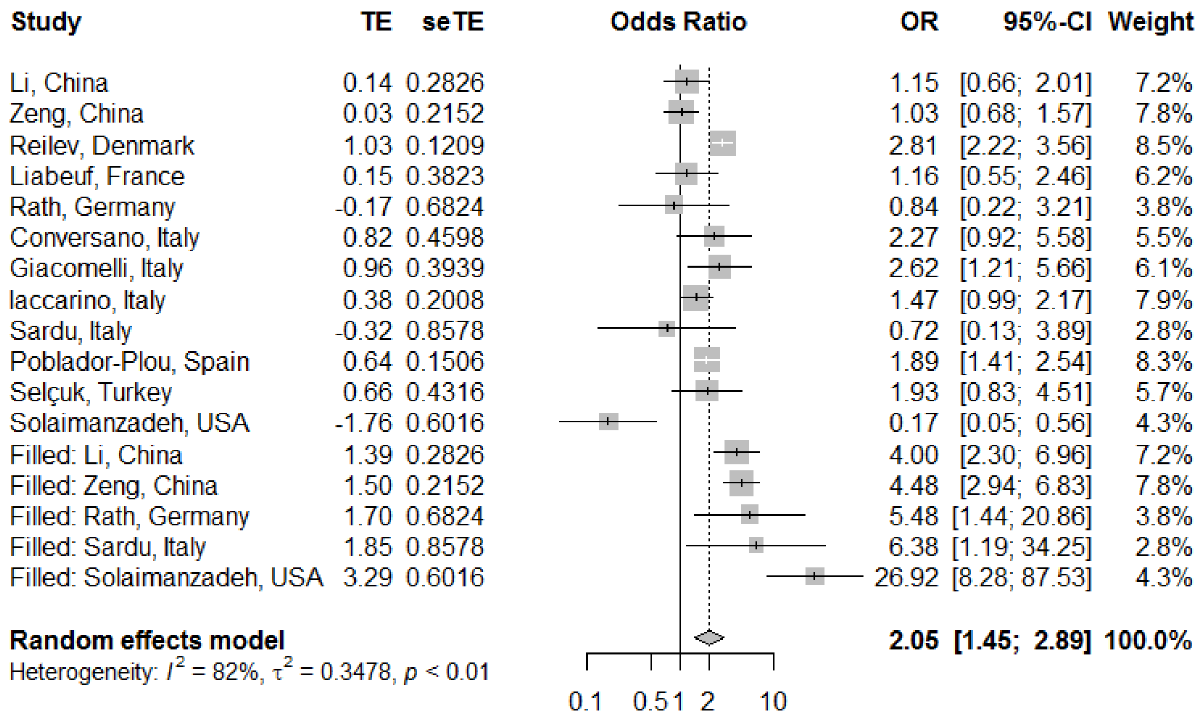


**Figure S55. Forest plot for the association between mortality outcomes in COVID-19 patients on a calcium channel blocker after adjusting for publication bias.**

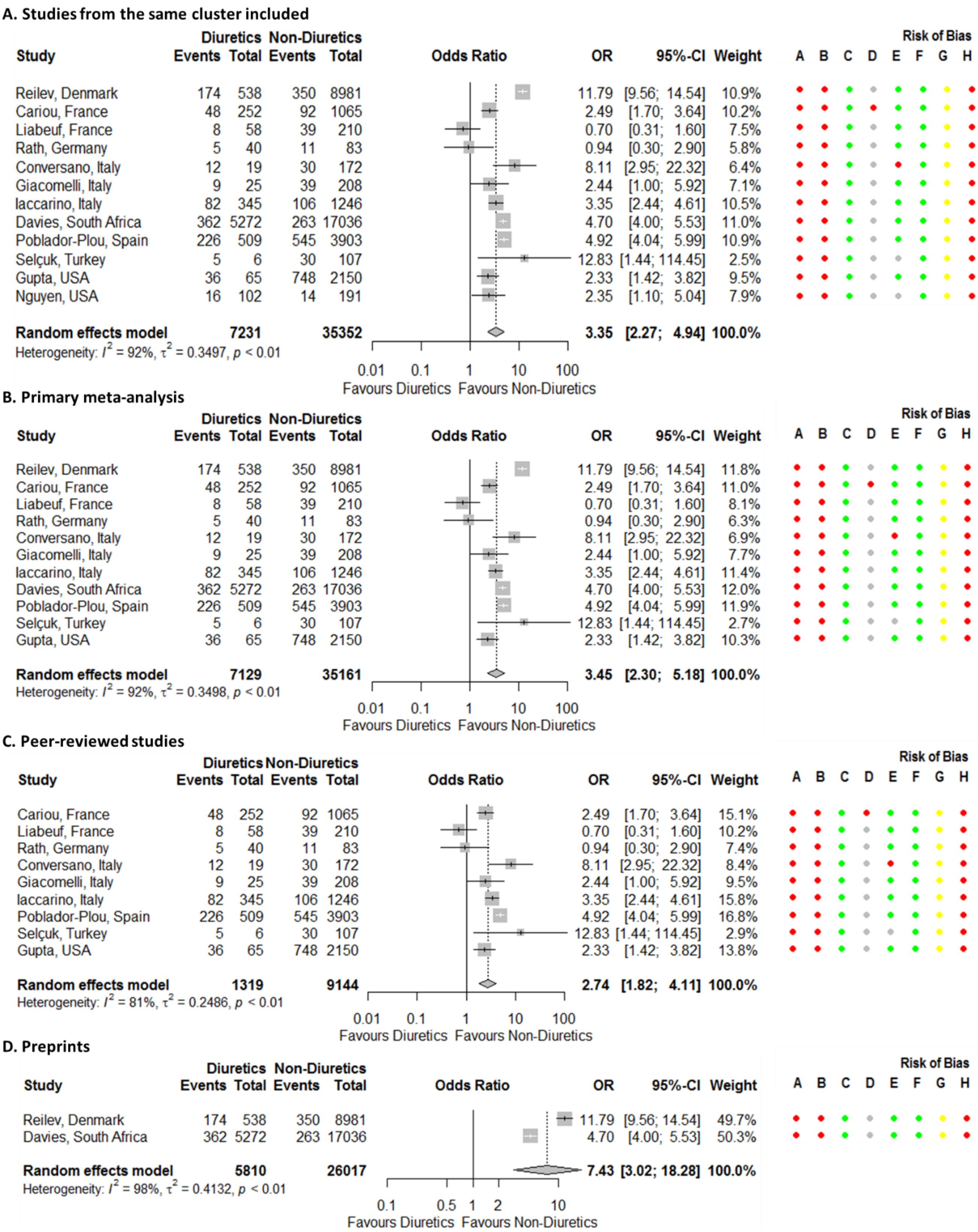


**Figure S56. Forest plots for association between mortality outcomes in COVID-19 patients on a diuretic.** Risk of bias legend. A = risk of bias due to confounding, B = risk of bias in selection of participants into the study, C = risk of bias in classification of interventions, D = risk of bias due to deviations from intended interventions, E = risk of bias due to missing data, F = risk of bias in measurement of outcomes, G = risk of bias in selection of the reported result, H = overall risk of bias. Color codes. Colour codes. Red = serious, yellow = moderate, green = low, grey = unclear.

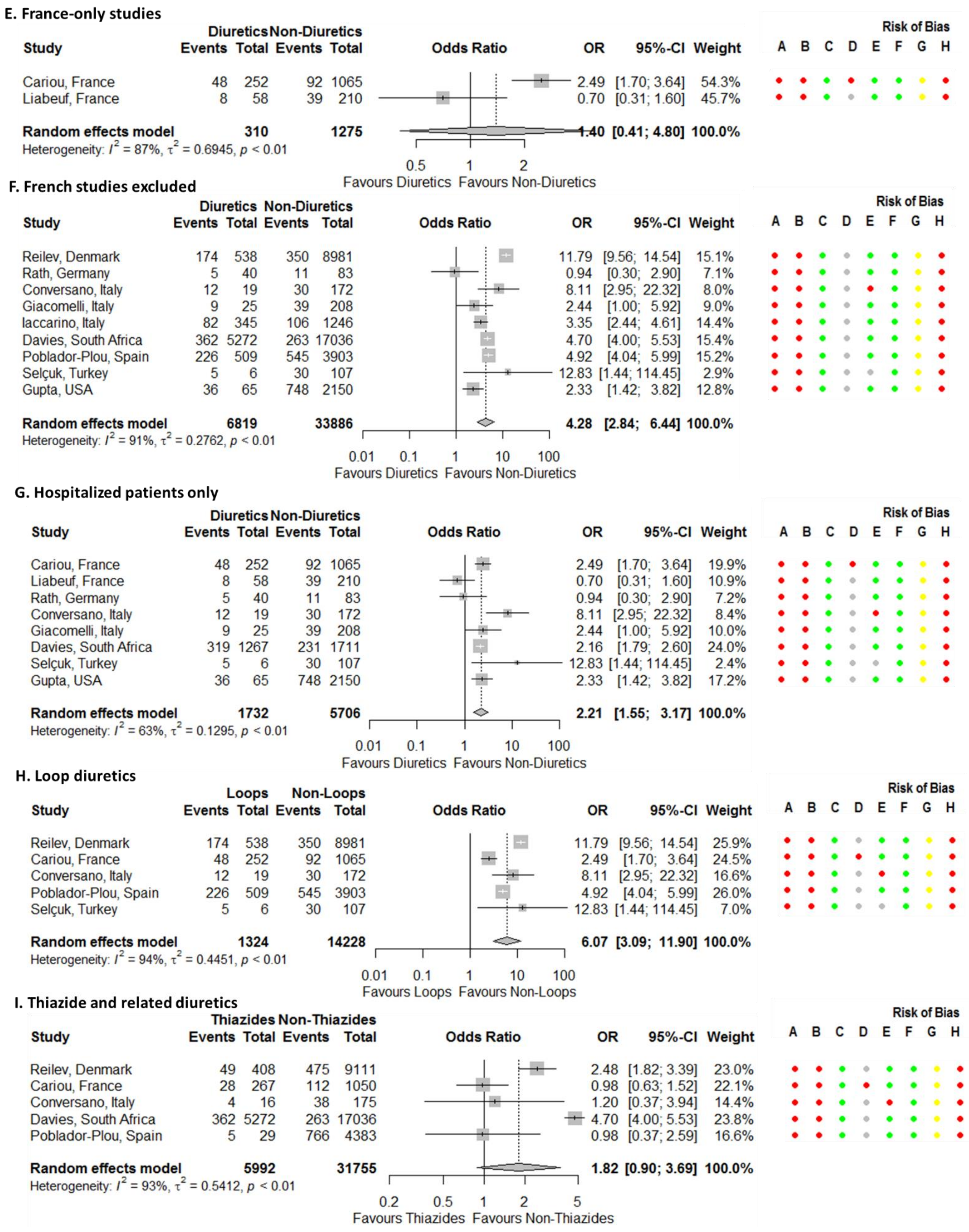


**Figure S56. Continued.**

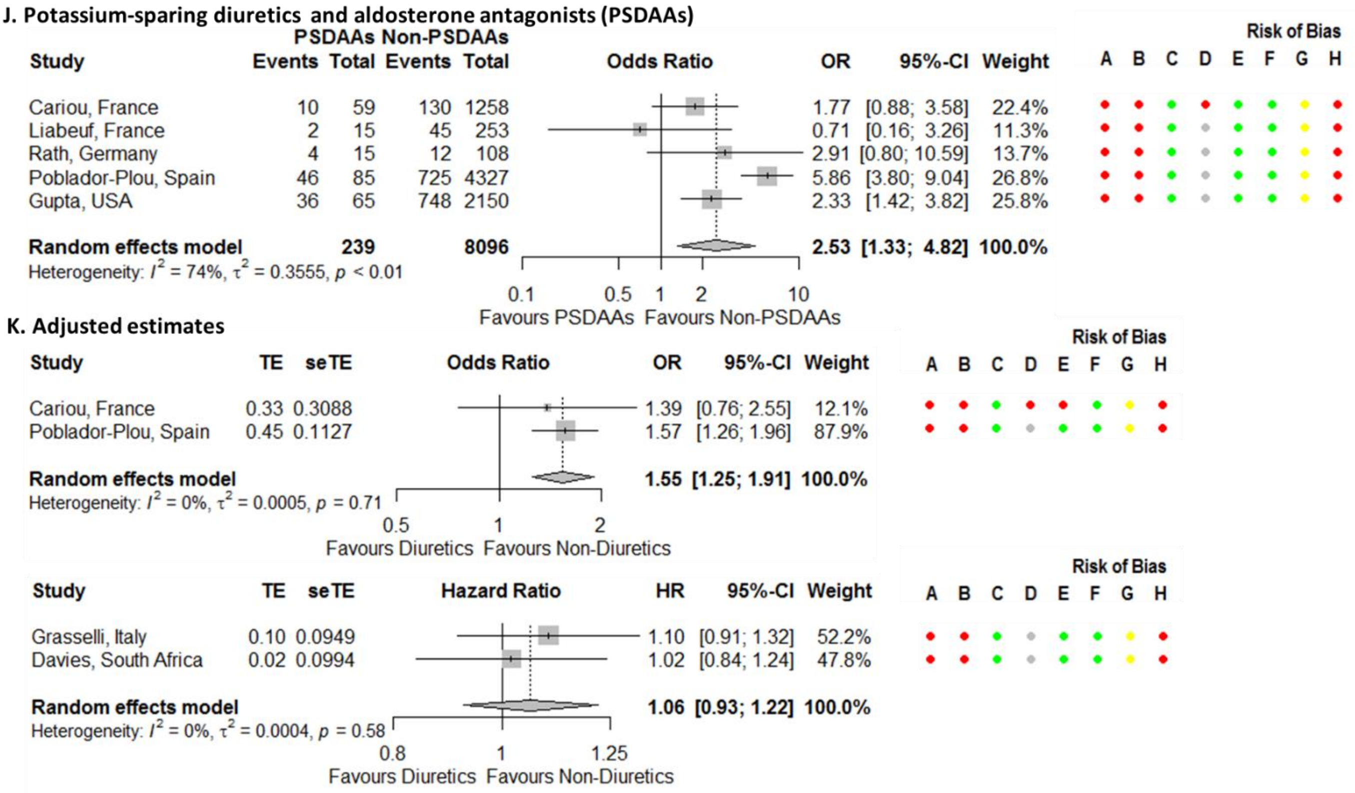


**Figure S56. Continued.**

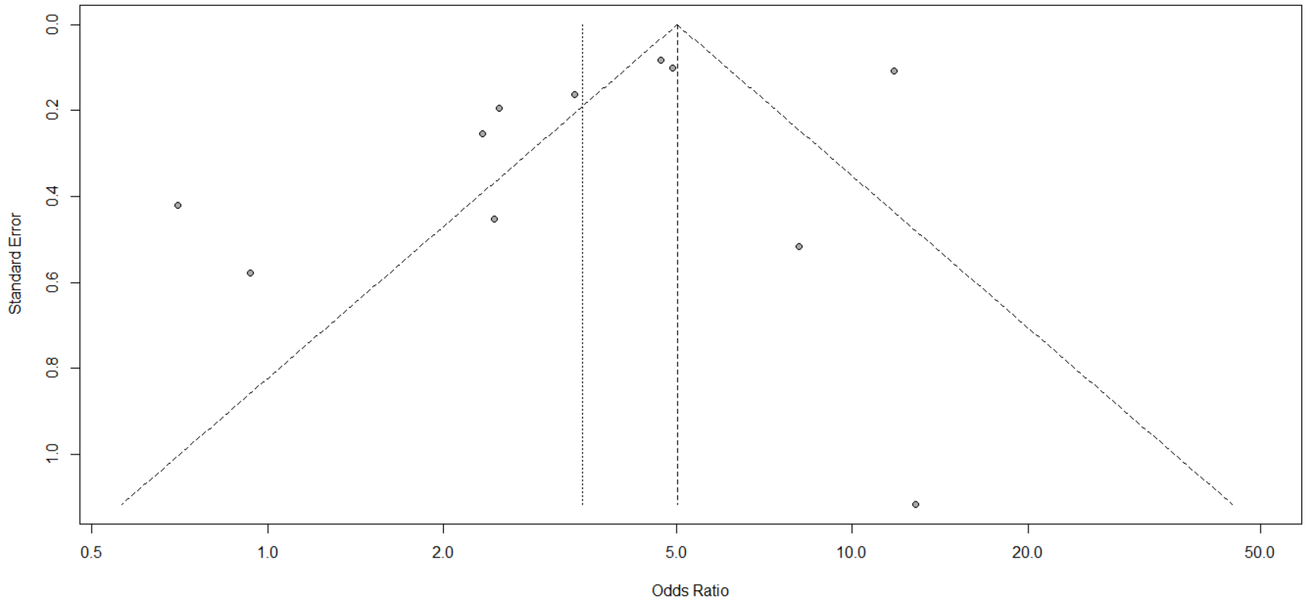


**Figure S57. Funnel plot for association between mortality outcomes in COVID-19 patients on a diuretic.**

**A. Studies from the same cluster included**

| Study | LMDs Events | LMDs Total | Non-LMDs Events | Non-LMDs Total | OR | 95%-CI | Weight |
|---|---|---|---|---|---|---|---|
| Qin, China | 1 | 12 | 6 | 38 | 0.48 | [0.05; 4.49] | 1.5% |
| Zeng, China | 5 | 38 | 160 | 993 | 0.79 | [0.30; 2.05] | 4.5% |
| Zhang, China | 67 | 1219 | 868 | 12762 | 0.80 | [0.62; 1.03] | 7.8% |
| Reilev, Denmark | 174 | 1277 | 350 | 8242 | 3.56 | [2.93; 4.31] | 8.0% |
| Cariou, France | 72 | 627 | 68 | 690 | 1.19 | [0.84; 1.68] | 7.4% |
| Rath, Germany | 3 | 41 | 13 | 82 | 0.42 | [0.11; 1.56] | 3.2% |
| Conversano, Italy | 9 | 21 | 33 | 170 | 3.11 | [1.21; 8.00] | 4.5% |
| Giacomelli, Italy | 11 | 39 | 37 | 194 | 1.67 | [0.76; 3.65] | 5.3% |
| Inciardi, Italy | 9 | 25 | 17 | 74 | 1.89 | [0.71; 5.02] | 4.4% |
| Violi, Italy | 13 | 46 | 51 | 273 | 1.71 | [0.84; 3.49] | 5.6% |
| Ayed, Kuwait | 3 | 10 | 42 | 93 | 0.52 | [0.13; 2.14] | 2.9% |
| Poblador-Plou, Spain | 163 | 723 | 608 | 3689 | 1.47 | [1.21; 1.79] | 8.0% |
| Selçuk, Turkey | 10 | 21 | 25 | 92 | 2.44 | [0.92; 6.44] | 4.4% |
| Garibaldi, USA | 43 | 271 | 70 | 476 | 1.09 | [0.72; 1.65] | 7.1% |
| Gu, USA | 11 | 144 | 6 | 582 | 7.94 | [2.88; 21.85] | 4.3% |
| Gupta, USA | 364 | 840 | 420 | 1375 | 1.74 | [1.45; 2.08] | 8.0% |
| Nguyen, USA | 10 | 90 | 20 | 203 | 1.14 | [0.51; 2.55] | 5.2% |
| Rodriguez-Nava, USA | 23 | 47 | 25 | 40 | 0.57 | [0.24; 1.36] | 4.9% |
| Wang, USA | 11 | 27 | 3 | 31 | 6.42 | [1.56; 26.46] | 2.9% |
| **Random effects model** | | **5518** | | **30099** | **1.50** | **[1.11; 2.03]** | **100.0%** |

Heterogeneity: $I^2 = 86\%$, $\tau^2 = 0.2823$, $p < 0.01$

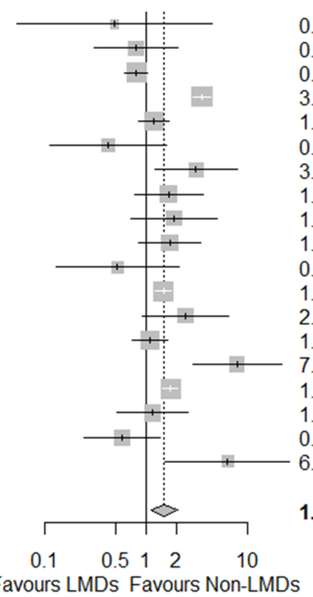


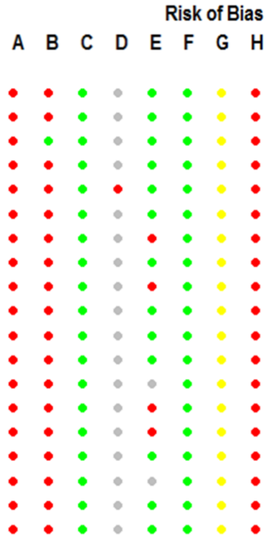


**B. Primary meta-analysis**

| Study | LMDs Events | LMDs Total | Non-LMDs Events | Non-LMDs Total | OR | 95%-CI | Weight |
|---|---|---|---|---|---|---|---|
| Zhang, China | 67 | 1219 | 868 | 12762 | 0.80 | [0.62; 1.03] | 11.3% |
| Reilev, Denmark | 174 | 1277 | 350 | 8242 | 3.56 | [2.93; 4.31] | 11.6% |
| Cariou, France | 72 | 627 | 68 | 690 | 1.19 | [0.84; 1.68] | 10.8% |
| Rath, Germany | 3 | 41 | 13 | 82 | 0.42 | [0.11; 1.56] | 4.5% |
| Conversano, Italy | 9 | 21 | 33 | 170 | 3.11 | [1.21; 8.00] | 6.4% |
| Giacomelli, Italy | 11 | 39 | 37 | 194 | 1.67 | [0.76; 3.65] | 7.5% |
| Inciardi, Italy | 9 | 25 | 17 | 74 | 1.89 | [0.71; 5.02] | 6.2% |
| Violi, Italy | 13 | 46 | 51 | 273 | 1.71 | [0.84; 3.49] | 8.1% |
| Ayed, Kuwait | 3 | 10 | 42 | 93 | 0.52 | [0.13; 2.14] | 4.1% |
| Poblador-Plou, Spain | 163 | 723 | 608 | 3689 | 1.47 | [1.21; 1.79] | 11.6% |
| Selçuk, Turkey | 10 | 21 | 25 | 92 | 2.44 | [0.92; 6.44] | 6.3% |
| Gupta, USA | 364 | 840 | 420 | 1375 | 1.74 | [1.45; 2.08] | 11.7% |
| **Random effects model** | | **4889** | | **27736** | **1.54** | **[1.09; 2.19]** | **100.0%** |

Heterogeneity: $I^2 = 90\%$, $\tau^2 = 0.2653$, $p < 0.01$

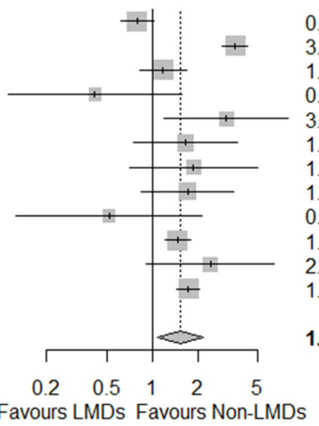


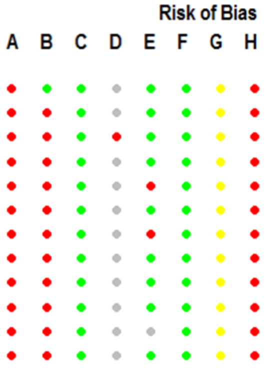


**C. Statins only**

| Study | Statins Events | Statins Total | Non-Statins Events | Non-Statins Total | OR | 95%-CI | Weight |
|---|---|---|---|---|---|---|---|
| Zhang, China | 67 | 1219 | 868 | 12762 | 0.80 | [0.62; 1.03] | 19.0% |
| Cariou, France | 72 | 627 | 68 | 690 | 1.19 | [0.84; 1.68] | 17.6% |
| Rath, Germany | 3 | 41 | 13 | 82 | 0.42 | [0.11; 1.56] | 5.8% |
| Conversano, Italy | 9 | 21 | 33 | 170 | 3.11 | [1.21; 8.00] | 8.9% |
| Inciardi, Italy | 9 | 25 | 17 | 74 | 1.89 | [0.71; 5.02] | 8.5% |
| Violi, Italy | 13 | 46 | 51 | 273 | 1.71 | [0.84; 3.49] | 11.8% |
| Selçuk, Turkey | 10 | 21 | 25 | 92 | 2.44 | [0.92; 6.44] | 8.6% |
| Gupta, USA | 364 | 840 | 420 | 1375 | 1.74 | [1.45; 2.08] | 19.9% |
| **Random effects model** | | **2840** | | **15518** | **1.41** | **[0.97; 2.04]** | **100.0%** |

Heterogeneity: $I^2 = 79\%$, $\tau^2 = 0.1717$, $p < 0.01$

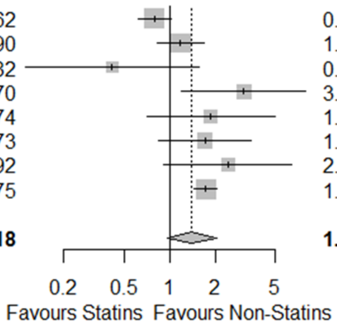


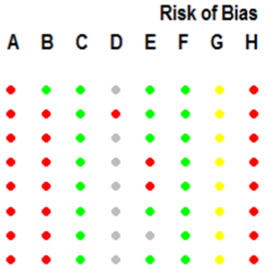


**Figure S58. Forest plots for association between mortality outcomes in COVID-19 patients on a lipid modifying drug.** Risk of bias legend. A = risk of bias due to confounding, B = risk of bias in selection of participants into the study, C = risk of bias in classification of interventions, D = risk of bias due to deviations from intended interventions, E = risk of bias due to missing data, F = risk of bias in measurement of outcomes, G = risk of bias in selection of the reported result, H = overall risk of bias. Color codes. Colour codes. Red = serious, yellow = moderate, green = low, grey = unclear.

**D. Adjusted estimates**

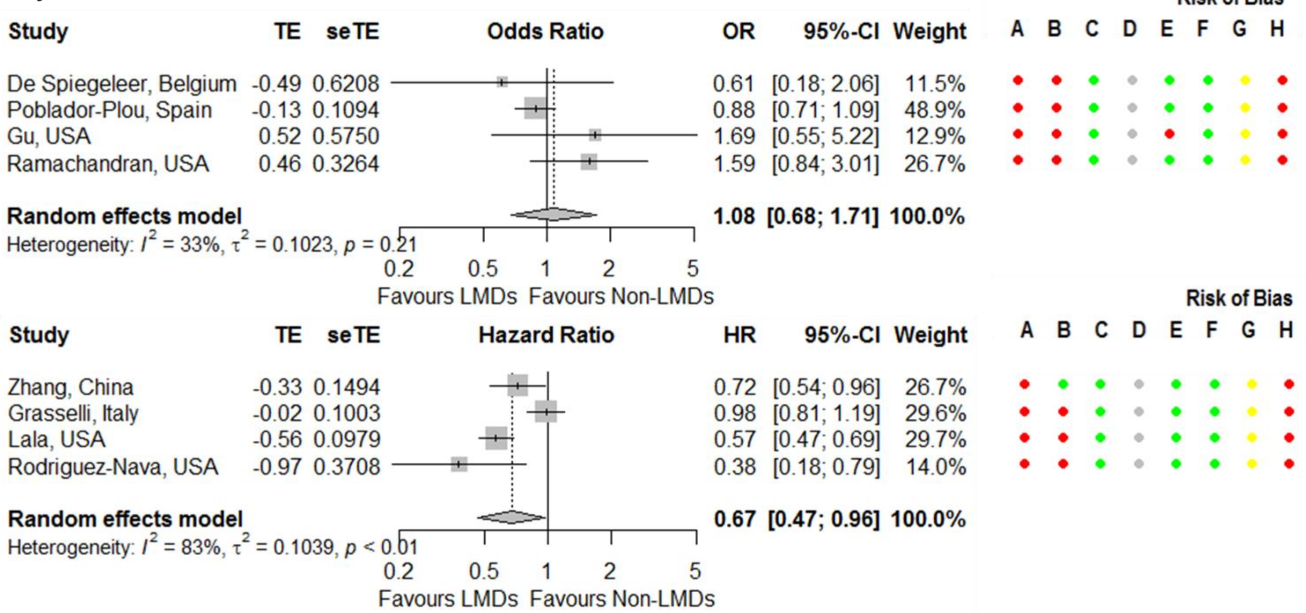


**Figure S58. Continued.**

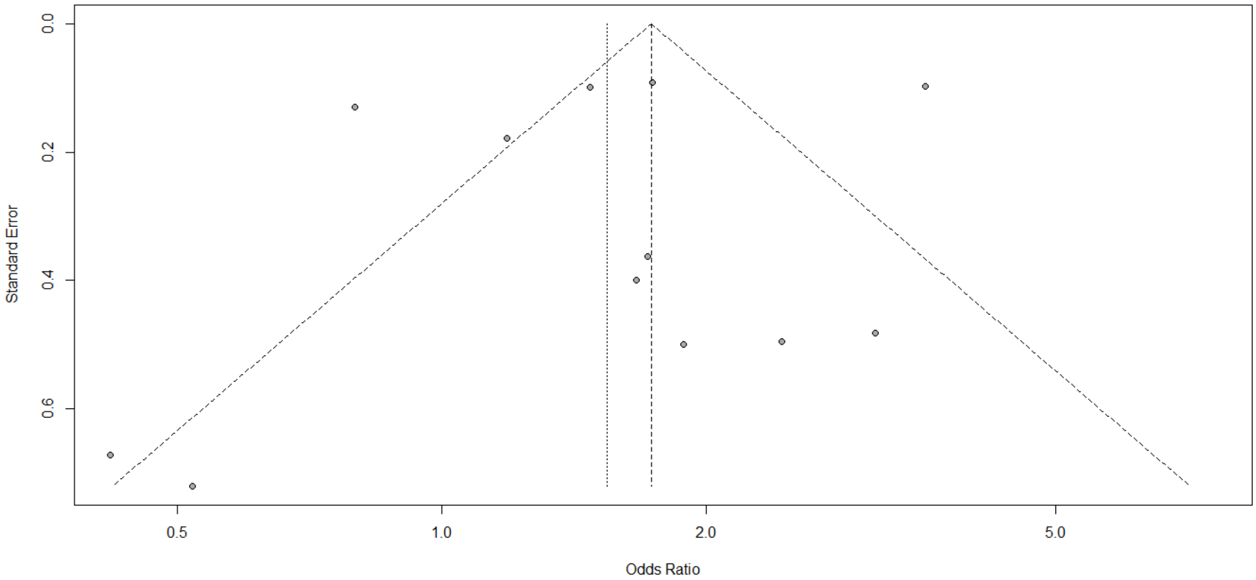


**Figure S59. Funnel plot for association between mortality outcomes in COVID-19 patients on a lipid modifying drug.**

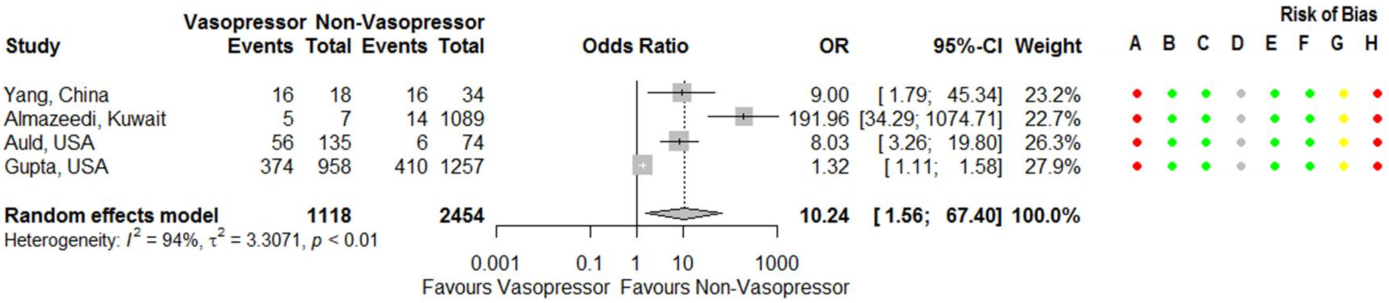


**Figure S60. Forest plots for association between mortality outcomes in COVID-19 patients receiving vasopressor treatment.** Risk of bias legend. A = risk of bias due to confounding, B = risk of bias in selection of participants into the study, C = risk of bias in classification of interventions, D = risk of bias due to deviations from intended interventions, E = risk of bias due to missing data, F = risk of bias in measurement of outcomes, G = risk of bias in selection of the reported result, H = overall risk of bias. Color codes. Colour codes. Red = serious, yellow = moderate, green = low, grey = unclear.

### Supplementary References


1. rvest: Easily Harvest (Scrape) Web Pages. R package version 0.3.5. [program]. online, 2019.
2. R: A language and environment for statistical computing. [program]. Vienna: R Foundation for Statistical Computing, 2019.
3. Xiong F, Tang H, Liu L, et al. Clinical Characteristics of and Medical Interventions for COVID-19 in Hemodialysis Patients in Wuhan, China. *Journal of the American Society of Nephrology : JASN* 2020;31(7):1387-97. doi: 10.1681/ASN.2020030354
4. Ebinger JE, Achamallah N, Ji H, et al. Pre-existing traits associated with Covid-19 illness severity. *PLoS ONE* 2020;15(7):1-16. doi: 10.1371/journal.pone.0236240
5. Dauchet L, Lambert M, Gauthier V, et al. ACE inhibitors, AT1 receptor blockers and COVID-19: clinical epidemiology evidences for a continuation of treatments. The ACER-COVID study. *MedRxiv* 2020 doi: 10.1101/2020.04.28.20078071 [published Online First: 1 May]
6. Joint Formulary Committee. British National Formulary 78 September 2019 – March 2020. 78 ed. London: BMJ Group and Pharmaceutical Press 2019.
7. Fosbøl EL, Butt JH, Østergaard L, et al. Association of Angiotensin-Converting Enzyme Inhibitor or Angiotensin Receptor Blocker Use With COVID-19 Diagnosis and Mortality. *JAMA: Journal of the American Medical Association* 2020;324(2):168-77. doi: 10.1001/jama.2020.11301
8. Grasselli G, Greco M, Zanella A, et al. Risk Factors Associated With Mortality Among Patients With COVID-19 in Intensive Care Units in Lombardy, Italy. *JAMA internal medicine* 2020 doi: 10.1001/jamainternmed.2020.3539
9. Schneeweiss MC, Leonard S, Weckstein A, et al. Renin-Angiotensin-Aldosterone-System inhibitor use in patients with COVID-19 infection and prevention of serious events: a cohort study in commercially insured patients in the US. *medRxiv* 2020 doi: 10.1101/2020.07.22.20159855 [published Online First: July 24]
10. Ashraf MA, Shokouhi N, Shirali E, et al. COVID-19 in Iran, a comprehensive investigation from exposure to treatment outcomes. *MedRxiv* 2020 doi: 10.1101/2020.04.20.20072421 [published Online First: 24 April]
11. Benotmane I, Vargas GG, Wendling M, et al. In-depth virological assessment of kidney transplant recipients with COVID-19. *medRxiv* 2020 doi: 10.1101/2020.06.17.20132076 [published Online First: June 19]
12. Feng Y, Ling Y, Bai T, et al. COVID-19 with Different Severities: A Multicenter Study of Clinical Features. *American journal of respiratory and critical care medicine* 2020;201(11):1380-88. doi: 10.1164/rccm.202002-0445OC
13. Gao C, Cai Y, Zhang K, et al. Association of hypertension and antihypertensive treatment with COVID-19 mortality: a retrospective observational study. *European heart journal* 2020;41(22):2058-66. doi: 10.1093/eurheartj/ehaa433
14. Garibaldi BT, Fiksel J, Muschelli J, et al. Patient trajectories and risk factors for severe outcomes among persons hospitalized for COVID-19 in the Maryland/DC region. *MedRxiv* 2020 doi: 10.1101/2020.05.24.20111864 [published Online First: 26 May]
15. Jung C, Bruno RR, Wernly B, et al. Inhibitors of the Renin-Angiotensin-Aldosterone System and Covid-19 in critically ill elderly patients. *European heart journal Cardiovascular pharmacotherapy* 2020 doi: 10.1093/ehjcvp/pvaa083
16. Oussalah A, Gleye S, Clerc Urmes I, et al. Long-Term ACE Inhibitor/ARB Use Is Associated with Severe Renal Dysfunction and Acute Kidney Injury in Patients with severe COVID-19: Results from a Referral Center Cohort in the North East of France. *Clinical infectious diseases : an official publication of the Infectious Diseases Society of America* 2020 doi: 10.1093/cid/ciaa677
17. Richardson S, Hirsch JS, Narasimhan M, et al. Presenting Characteristics, Comorbidities, and Outcomes Among 5700 Patients Hospitalized With COVID-19 in the New York City Area. *JAMA* 2020 doi: 10.1001/jama.2020.6775 [published Online First: 2020/04/23]

18. Sardu C, Maggi P, Messina V, et al. Could anti-hypertensive drug therapy affect the clinical prognosis of hypertensive patients with COVID-19 infection? Data from centers of southern Italy. *Journal of the American Heart Association* 2020:e016948. doi: 10.1161/JAHA.120.016948
19. Tan N-D, Qiu Y, Xing X-B, et al. Associations between Angiotensin Converting Enzyme Inhibitors and Angiotensin II Receptor Blocker Use, Gastrointestinal Symptoms, and Mortality among Patients with COVID-19. *Gastroenterology* 2020 doi: 10.1053/j.gastro.2020.05.034
20. Bravi F, Flacco ME, Carradori T, et al. Predictors of severe or lethal COVID-19, including Angiotensin Converting Enzyme Inhibitors and Angiotensin II Receptor Blockers, in a sample of infected Italian citizens. *MedRxiv* 2020 doi: 10.1101/2020.05.21.20109082 [published Online First: 23 May]
21. Cariou B, Hadjadj S, Wargny M, et al. Phenotypic characteristics and prognosis of inpatients with COVID-19 and diabetes: the CORONADO study. *Diabetologia* 2020 doi: 10.1007/s00125-020-05180-x
22. Jung S-Y, Choi JC, You S-H, et al. Association of renin-angiotensin-aldosterone system inhibitors with COVID-19-related outcomes in Korea: a nationwide population-based cohort study. *Clinical infectious diseases : an official publication of the Infectious Diseases Society of America* 2020 doi: 10.1093/cid/ciaa624
23. Li J, Wang X, Chen J, et al. Association of Renin-Angiotensin System Inhibitors With Severity or Risk of Death in Patients With Hypertension Hospitalized for Coronavirus Disease 2019 (COVID-19) Infection in Wuhan, China. *JAMA cardiology* 2020 doi: 10.1001/jamacardio.2020.1624
24. López-Otero D, López-Pais J, Cacho-Antonio CE, et al. Impact of angiotensin-converting enzyme inhibitors and angiotensin receptor blockers on COVID-19 in a western population. CARDIOVID registry. *Revista espanola de cardiologia (English ed)* 2020 doi: 10.1016/j.rec.2020.05.018
25. Mehta N, Kalra A, Nowacki AS, et al. Association of Use of Angiotensin-Converting Enzyme Inhibitors and Angiotensin II Receptor Blockers With Testing Positive for Coronavirus Disease 2019 (COVID-19). *JAMA cardiology* 2020 doi: 10.1001/jamacardio.2020.1855
26. Rentsch CT, Kidwai-Khan F, Tate JP, et al. Covid-19 Testing, Hospital Admission, and Intensive Care Among 2,026,227 United States Veterans Aged 54-75 Years. *MedRxiv* 2020 doi: 10.1101/2020.04.09.20059964 [published Online First: 14 April]
27. Reynolds HR, Adhikari S, Pulgarin C, et al. Renin-Angiotensin-Aldosterone System Inhibitors and Risk of Covid-19. *The New England journal of medicine* 2020 doi: 10.1056/NEJMoa2008975
28. Şenkal N, Meral R, Medetalibeyoğlu A, et al. Association between chronic ACE inhibitor exposure and decreased odds of severe disease in patients with COVID-19. *Anatolian journal of cardiology* 2020;24(1):21-29. doi: 10.14744/AnatolJCardiol.2020.57431
29. Xu J, Huang C, Fan G, et al. Use of angiotensin-converting enzyme inhibitors and angiotensin II receptor blockers in context of COVID-19 outbreak: a retrospective analysis. *Frontiers of Medicine* 2020:1. doi: 10.1007/s11684-020-0800-y
30. Thompson SG, Higgins JP. How should meta-regression analyses be undertaken and interpreted? *Stat Med* 2002;21(11):1559-73. doi: 10.1002/sim.1187 [published Online First: 2002/07/12]
31. Schwarzer G. meta: An R package for meta-analysis. *R News* 2007;7(3):40-45.
32. Moher D, Liberati A, Tetzlaff J, et al. Preferred reporting items for systematic reviews and meta-analyses: the PRISMA statement. *PLoS Med* 2009;6(7):e1000097. doi: 10.1371/journal.pmed.1000097 [published Online First: 2009/07/22]
33. Trubiano JA, Vogrin S, Smibert OC, et al. COVID-MATCH65 – A prospectively derived clinical decision rule for severe acute respiratory syndrome coronavirus. *medRxiv* 2020 doi: 10.1101/2020.06.30.20143818 [published Online First: July 2]

34. De Spiegeleer A, Bronselaer A, Teo JT, et al. The Effects of ARBs, ACEis, and Statins on Clinical Outcomes of COVID-19 Infection Among Nursing Home Residents. *Journal of the American Medical Directors Association* 2020;21(7):909-14. doi: 10.1016/j.jamda.2020.06.018
35. Mazzoleni L, Ghafari C, Mestrez F, et al. COVID-19 Outbreak in a Hemodialysis Center: A Retrospective Monocentric Case Series. *Canadian Journal of Kidney Health & Disease* 2020:1.
36. Chen M, Fan Y, Wu X, et al. Clinical Characteristics And Risk Factors For Fatal Outcome in Patients With 2019-Coronavirus Infected Disease (COVID-19) in Wuhan, China (2/27/2020). *Preprints with Lancet (SSRN)* 2020 doi: 10.2139/ssrn.3546069 [published Online First: 3 Mar]
37. Chen Y, Yang D, Cheng B, et al. Clinical Characteristics and Outcomes of Patients With Diabetes and COVID-19 in Association With Glucose-Lowering Medication. *Diabetes care* 2020 doi: 10.2337/dc20-0660
38. Feng Z, Li J, Yao S, et al. The Use of Adjuvant Therapy in Preventing Progression to Severe Pneumonia in Patients with Coronavirus Disease 2019: A Multicenter Data Analysis. *MedRxiv* 2020 doi: 10.1101/2020.04.08.20057539 [published Online First: 10 April]
39. Guo T, Fan Y, Chen M, et al. Cardiovascular Implications of Fatal Outcomes of Patients With Coronavirus Disease 2019 (COVID-19). *JAMA Cardiology* 2020;5(7):811.
40. Hu J, Zhang X, Zhang X, et al. COVID-19 patients with hypertension have more severity condition, and ACEI/ARB treatment have no infulence on the clinical severity and outcome. *The Journal of infection* 2020 doi: 10.1016/j.jinf.2020.05.056
41. Huang Z, Cao J, Yao Y, et al. The effect of RAS blockers on the clinical characteristics of COVID-19 patients with hypertension. *Annals of translational medicine* 2020;8(7):430. doi: 10.21037/atm.2020.03.229
42. Jiang S, Wang R, Li L, et al. Liver Injury in Critically Ill and Non-critically Ill COVID-19 Patients: A Multicenter, Retrospective, Observational Study. *Frontiers in medicine* 2020;7:347. doi: 10.3389/fmed.2020.00347
43. Li T, Lu L, Zhang W, et al. Clinical characteristics of 312 hospitalized older patients with COVID-19 in Wuhan, China. *Archives of gerontology and geriatrics* 2020;91:104185. doi: 10.1016/j.archger.2020.104185
44. Li X, Xu S, Yu M, et al. Risk factors for severity and mortality in adult COVID-19 inpatients in Wuhan. *The Journal of Allergy and Clinical Immunology* 2020;146(1):110-18. doi: 10.1016/j.jaci.2020.04.006
45. Li Y, Li M, Wang M, et al. Acute cerebrovascular disease following COVID-19: a single center, retrospective, observational study. *Stroke and vascular neurology* 2020 doi: 10.1136/svn-2020-000431
46. Liu Y, Huang F, Xu J, et al. Anti-hypertensive Angiotensin II receptor blockers associated to mitigation of disease severity in elderly COVID-19 patients. *MedRxiv* 2020 doi: 10.1101/2020.03.20.20039586 [published Online First: 27 March]
47. Liu X, Zhou H, Zhou Y, et al. Risk factors associated with disease severity and length of hospital stay in COVID-19 patients. *Journal of Infection* 2020;81(1):e95-e97. doi: 10.1016/j.jinf.2020.04.008
48. Liu X, Liu Y, Chen K, et al. Efficacy of ACEIs/ARBs versus CCBs on the progression of COVID-19 patients with hypertension in Wuhan: A hospital-based retrospective cohort study. *Journal of medical virology* 2020 doi: 10.1002/jmv.26315
49. Meng J, Xiao G, Zhang J, et al. Renin-angiotensin system inhibitors improve the clinical outcomes of COVID-19 patients with hypertension. *Emerg Microbes Infect* 2020;9(1):757-60. doi: 10.1080/22221751.2020.1746200 [published Online First: 2020/04/02]
50. Peng YD, Meng K, Guan HQ, et al. Clinical characteristics and outcomes of 112 cardiovascular disease patients infected by 2019-nCoV. *Zhonghua xin xue guan bing za zhi* 2020;48:E004. doi: 10.3760/cma.j.cn112148-20200220-00105
51. Qin C, Zhou L, Hu Z, et al. Clinical Characteristics and Outcomes of COVID-19 Patients With a History of Stroke in Wuhan, China, 2020:2219-23.

52. SHI C, WANG C, WANG H, et al. The potential of low molecular weight heparin to mitigate cytokine storm in severe COVID-19 patients: a retrospective clinical study. *MedRxiv* 2020 doi: 10.1101/2020.03.28.20046144 [published Online First: 1 April]
53. Tang N, Bai H, Chen X, et al. Anticoagulant treatment is associated with decreased mortality in severe coronavirus disease 2019 patients with coagulopathy. *J Thromb Haemost* 2020;18(5):1094-99. doi: 10.1111/jth.14817 [published Online First: 2020/03/29]
54. Xie Y, Chen S, Wang X, et al. Early Diagnosis and Clinical Significance of Acute Cardiac Injury - Under the Iceberg: A Retrospective Cohort Study of 619 Non-critically Ill Hospitalized COVID-19 Pneumonia Patients. *medRxiv* 2020 doi: 10.1101/2020.07.06.20147256 [published Online First: July 7]
55. Xie Y, You Q, Wu C, et al. Impact of Cardiovascular Disease on Clinical Characteristics and Outcomes of Coronavirus Disease 2019 (COVID-19). *Circulation journal : official journal of the Japanese Circulation Society* 2020;84(8):1277-83. doi: 10.1253/circj.CJ-20-0348
56. Yan H, Valdes AM, Vijay A, et al. Role of Drugs Affecting the Renin-Angiotensin-Aldosterone System on Susceptibility and Severity of COVID-19: A Large Case-Control Study from Zheijang Province, China. *MedRxiv* 2020 doi: 10.1101/2020.04.24.20077875 [published Online First: 29 April]
57. Yang G, Tan Z, Peng L, et al. Effects of angiotensin II receptor blockers and ACE (angiotensin-converting enzyme) inhibitors on virus infection, inflammatory status, and clinical outcomes in patients with COVID-19 and hypertension: A single-center retrospective study. *Hypertension* 2020:51-58. doi: 10.1161/HYPERTENSIONAHA.120.15143
58. Yang W, Xiaofan L, Yongsheng L, et al. Clinical Course and Outcomes of 344 Intensive Care Patients with COVID-19. *American Journal of Respiratory & Critical Care Medicine* 2020;201(11):1430.
59. Yang X, Yu Y, Xu J, et al. Clinical course and outcomes of critically ill patients with SARS-CoV-2 pneumonia in Wuhan, China: a single-centered, retrospective, observational study. *Lancet Respiratory Medicine* 2020;8(5):475-81. doi: 10.1016/S2213-2600(20)30079-5 [published Online First: MAY 01]
60. Yao Y, Cao J, Wang Q, et al. D-dimer as a biomarker for disease severity and mortality in COVID-19 patients: a case control study. *Journal of Intensive Care* 2020;8(1) doi: 10.1186/s40560-020-00466-z
61. Ye C, Zhang S, Zhang X, et al. Impact of comorbidities on patients with COVID-19: A large retrospective study in Zhejiang, China. *Journal of medical virology* 2020 doi: 10.1002/jmv.26183
62. Yin R, Yang Z, Wei Y, et al. Clinical characteristics of 106 patients with neurological diseases and co-morbid coronavirus disease 2019: a retrospective study *MedRxiv* 2020 doi: 10.1101/2020.04.29.20085415 [published Online First: 5 May]
63. Zhang X, Yu J, Pan L-Y, et al. ACEI/ARB use and risk of infection or severity or mortality of COVID-19: A systematic review and meta-analysis. *Pharmacological research* 2020;158:104927. doi: 10.1016/j.phrs.2020.104927
64. Zeng H, Zhang T, He X, et al. Impact of Chronic Comorbidities on Progression and Prognosis in Patients with COVID-19: A Retrospective Cohort Study in 1031 Hospitalized Cases in Wuhan, China. *MedRxiv* 2020 doi: 10.1101/2020.06.14.20125997 [published Online First: 16 June]
65. Zeng Z, Sha T, Zhang Y, et al. Hypertension in patients hospitalized with COVID-19 in Wuhan, China: A single-center retrospective observational study. *MedRxiv* 2020 doi: 10.1101/2020.04.06.20054825 [published Online First: 11 April]
66. Zhang L, Sun Y, Zeng H-L, et al. Calcium channel blocker amlodipine besylate is associated with reduced case fatality rate of COVID-19 patients with hypertension. *MedRxiv* 2020 doi: 10.1101/2020.04.08.20047134 [published Online First: 14 April]
67. Zhang P, Zhu L, Cai J, et al. Association of Inpatient Use of Angiotensin-Converting Enzyme Inhibitors and Angiotensin II Receptor Blockers With Mortality Among Patients With

Hypertension Hospitalized With COVID-19. *Circulation Research* 2020(12):1671. doi: 10.1161/CIRCRESAHA.120.317134
68. Zhang X-J, Qin J-J, Cheng X, et al. In-Hospital Use of Statins Is Associated with a Reduced Risk of Mortality among Individuals with COVID-19. *Cell metabolism* 2020 doi: 10.1016/j.cmet.2020.06.015
69. Zhou H, Xiao X, Wang X, et al. Clinical Features of Hemodialysis (HD) patients confirmed with Coronavirus Disease 2019 (COVID-19): a Retrospective Case-Control Study. *medRxiv* 2020 doi: 10.1101/2020.07.06.20147827 [published Online First: July 10]
70. Zhou X, Zhu J, Xu T. Clinical characteristics of coronavirus disease 2019 (COVID-19) patients with hypertension on renin-angiotensin system inhibitors. *Clinical and experimental hypertension (New York, NY : 1993)* 2020:1-5. doi: 10.1080/10641963.2020.1764018
71. Reilev M, Kristensen KB, Pottegaard A, et al. Characteristics and predictors of hospitalization and death in the first 9,519 cases with a positive RT-PCR test for SARS-CoV-2 in Denmark: A nationwide cohort. *medRxiv* 2020 doi: 10.1101/2020.05.24.20111823 [published Online First: 26 May]
72. Feuth T, Saaresranta T, Karlsson A, et al. Is sleep apnoea a risk factor for Covid-19? Findings from a retrospective cohort study. *MedRxiv* 2020 doi: 10.1101/2020.05.14.20098319 [published Online First: 18 May]
73. Allenbach Y, Saadoun D, Maalouf G, et al. Multivariable prediction model of intensive care unit transfer and death: a French prospective cohort study of COVID-19 patients. *MedRxiv* 2020 doi: 10.1101/2020.05.04.20090118 [published Online First: 8 May]
74. Bar S, Lecourtois A, Diouf M, et al. The association of lung ultrasound images with COVID-19 infection in an emergency room cohort. *Anaesthesia* 2020 doi: 10.1111/anae.15175
75. Basse C, Diakite S, Servois V, et al. Characteristics and outcome of SARS-CoV-2 infection in cancer patients. *MedRxiv* 2020 doi: 10.1101/2020.05.14.20101576 [published Online First: 19 May]
76. Khider L, Gendron N, Goudot G, et al. Curative anticoagulation prevents endothelial lesion in COVID-19 patients. *J Thromb Haemost* 2020 doi: 10.1111/jth.14968 [published Online First: 2020/06/20]
77. Kibler M, Carmona A, Marchandot B, et al. Risk and severity of COVID-19 and ABO blood group in transcatheter aortic valve patients. *MedRxiv* 2020 doi: 10.1101/2020.06.13.20130211 [published Online First: 16 June]
78. Liabeuf S, Moragny J, Bennis Y, et al. Association between renin-angiotensin system inhibitors and COVID-19 complications. *European heart journal Cardiovascular pharmacotherapy* 2020 doi: 10.1093/ehjcvp/pvaa062
79. Meszaros M, Meunier L, Morquin D, et al. Abnormal liver tests in patients hospitalized with Coronavirus disease 2019: Should we worry? *Liver international : official journal of the International Association for the Study of the Liver* 2020 doi: 10.1111/liv.14557
80. Rath D, Petersen-Uribe Á, Avdiu A, et al. Impaired cardiac function is associated with mortality in patients with acute COVID-19 infection. *Clinical Research in Cardiology* 2020:1. doi: 10.1007/s00392-020-01683-0
81. Rieder M, Goller I, Jeserich M, et al. Rate of venous thromboembolism in a prospective all-comers cohort with COVID-19, 2020.
82. Sacco V, Rauch B, Gar C, et al. Overweight/obesity as the potentially most important lifestyle factor associated with signs of pneumonia in COVID-19. *medRxiv* 2020 doi: 10.1101/2020.07.23.20161042 [published Online First: July 24]
83. Cheung KS, Hung IFN, Leung WK. Association between angiotensin blockade and COVID-19 severity in Hong Kong. *CMAJ* 2020;192(23):E635. doi: 10.1503/cmaj.75865 [published Online First: 2020/06/24]
84. Zhou J, Tse G, Lee S, et al. Identifying main and interaction effects of risk factors to predict intensive care admission in patients hospitalized with COVID-19: a retrospective cohort

study in Hong Kong. *medRxiv* 2020 doi: 10.1101/2020.06.30.20143651 [published Online First: July 2]
85. Amit M, Sorkin A, Chen J, et al. Clinical Course and Outcomes of Severe Covid-19: A National Scale Study. *Journal of Clinical Medicine* 2020;9(7):2282.
86. Chodick G, Nutman A, Yiekutiel N, et al. Angiotensin-converting enzyme inhibitors and angiotensin-receptor blockers are not associated with increased risk of SARS-CoV-2 infection. *J Travel Med* 2020;27(5) doi: 10.1093/jtm/taaa069 [published Online First: 2020/05/15]
87. Alberici F, Delbarba E, Manenti C, et al. A report from the Brescia Renal COVID Task Force on the clinical characteristics and short-term outcome of hemodialysis patients with SARS-CoV-2 infection. *Kidney International* 2020;98(1):20-26. doi: 10.1016/j.kint.2020.04.030
88. Alberici F, Delbarba E, Manenti C, et al. A single center observational study of the clinical characteristics and short-term outcome of 20 kidney transplant patients admitted for SARS-CoV2 pneumonia. *Kidney International* 2020;97(6):1083-88. doi: 10.1016/j.kint.2020.04.002
89. Benelli G, Buscarini E, Canetta C, et al. SARS-COV-2 comorbidity network and outcome in hospitalized patients in Crema, Italy. *MedRxiv* 2020 doi: 10.1101/2020.04.14.20053090 [published Online First: 30 April]
90. Cannata F, Chiarito M, Reimers B, et al. Continuation versus discontinuation of ACE inhibitors or angiotensin II receptor blockers in COVID-19: effects on blood pressure control and mortality. *European heart journal Cardiovascular pharmacotherapy* 2020 doi: 10.1093/ehjcvp/pvaa056
91. Conversano A, Melillo F, Napolano A, et al. RAAs inhibitors and outcome in patients with SARS-CoV-2 pneumonia. A case series study. *Hypertension (Dallas, Tex : 1979)* 2020 doi: 10.1161/HYPERTENSIONAHA.120.15312
92. Di Micco P, Russo V, Carannante N, et al. Clotting Factors in COVID-19: Epidemiological Association and Prognostic Values in Different Clinical Presentations in an Italian Cohort. *Journal of Clinical Medicine* 2020;9(5):1371.
93. Fasano A, Cereda E, Barichella M, et al. COVID-19 in Parkinson's Disease Patients Living in Lombardy, Italy. *Movement disorders : official journal of the Movement Disorder Society* 2020;35(7):1089-93. doi: 10.1002/mds.28176
94. Felice C, Nardin C, Di Tanna GL, et al. Use of RAAS inhibitors and risk of clinical deterioration in COVID-19: results from an Italian cohort of 133 hypertensives. *American journal of hypertension* 2020 doi: 10.1093/ajh/hpaa096
95. Ferrante G, Fazzari F, Cozzi O, et al. Risk factors for myocardial injury and death in patients with COVID-19: insights from a cohort study with chest computed tomography. *Cardiovascular research* 2020 doi: 10.1093/cvr/cvaa193
96. Giacomelli A, Ridolfo AL, Milazzo L, et al. 30-day mortality in patients hospitalized with COVID-19 during the first wave of the Italian epidemic: A prospective cohort study. *Pharmacol Res* 2020;158:104931. doi: 10.1016/j.phrs.2020.104931 [published Online First: 2020/05/25]
97. Giorgi Rossi P, Marino M, Formisano D, et al. Characteristics and outcomes of a cohort of SARS-CoV-2 patients in the Province of Reggio Emilia, Italy. *MedRxiv* 2020 doi: 10.1101/2020.04.13.20063545 [published Online First: 16 April]
98. Gnavi R, Demaria M, Picariello R, et al. Therapy with agents acting on the renin-angiotensin system and risk of SARS-CoV-2 infection. *Clinical infectious diseases : an official publication of the Infectious Diseases Society of America* 2020 doi: 10.1093/cid/ciaa634
99. Iaccarino G, Grassi G, Borghi C, et al. Age and Multimorbidity Predict Death Among COVID-19 Patients: Results of the SARS-RAS Study of the Italian Society of Hypertension. *Hypertension* 2020;76(2):366-72. doi: 10.1161/HYPERTENSIONAHA.120.15324 [published Online First: 2020/06/23]
100. Iacovoni A, Boffini M, Pidello S, et al. A case series of novel coronavirus infection in heart transplantation from 2 centers in the pandemic area in the North of Italy. *The Journal of*

*heart and lung transplantation : the official publication of the International Society for Heart Transplantation* 2020 doi: 10.1016/j.healun.2020.06.016
101. Inciardi RM, Adamo M, Lupi L, et al. Characteristics and outcomes of patients hospitalized for COVID-19 and cardiac disease in Northern Italy. *European heart journal* 2020;41(19):1821-29. doi: 10.1093/eurheartj/ehaa388
102. Mancia G, Rea F, Ludergnani M, et al. Renin-Angiotensin-Aldosterone System Blockers and the Risk of Covid-19. *The New England journal of medicine* 2020 doi: 10.1056/NEJMoa2006923
103. Oliva A, Siccardi G, Migliarini A, et al. Co-infection of SARS-CoV-2 with Chlamydia or Mycoplasma pneumoniae: a case series and review of the literature. *Infection: A Journal of Infectious Diseases* 2020:1. doi: 10.1007/s15010-020-01483-8
104. Parigi TL, Vespa E, Pugliese N. COVID-19, ACEI/ARBs and gastrointestinal symptoms: the jury is still out on the association. *Gastroenterology* 2020 doi: 10.1053/j.gastro.2020.06.095 [published Online First: 2020/07/20]
105. Perotti C, Baldanti F, Bruno R, et al. Mortality reduction in 46 severe Covid-19 patients treated with hyperimmune plasma. A proof of concept single arm multicenter interventional trial. *MedRxiv* 2020 doi: 10.1101/2020.05.26.20113373 [published Online First: 29 May]
106. Russo V, Di Maio M, Attena E, et al. Clinical impact of pre-admission antithrombotic therapy in hospitalized patients with COVID-19: A multicenter observational study. *Pharmacological research* 2020;159:104965. doi: 10.1016/j.phrs.2020.104965
107. Tedeschi S, Giannella M, Bartoletti M, et al. Clinical Impact of Renin-angiotensin System Inhibitors on In-hospital Mortality of Patients With Hypertension Hospitalized for Coronavirus Disease 2019. *Clin Infect Dis* 2020;71(15):899-901. doi: 10.1093/cid/ciaa492 [published Online First: 2020/04/28]
108. Trecarichi EM, Mazzitelli M, Serapide F, et al. Characteristics, outcome and predictors of in-hospital mortality in an elderly population from a SARS-CoV-2 outbreak in a long-term care facility. *medRxiv* 2020 doi: 10.1101/2020.06.30.20143701 [published Online First: July 2]
109. Viecca M, Radovanovic D, Forleo GB, et al. Enhanced platelet inhibition treatment improves hypoxemia in patients with severe Covid-19 and hypercoagulability. A case control, proof of concept study. *Pharmacological research* 2020;158:104950. doi: 10.1016/j.phrs.2020.104950
110. Violi F, Cangemi R, Romiti GF, et al. Is Albumin Predictor of Mortality in COVID-19? *Antioxidants & redox signaling* 2020 doi: 10.1089/ars.2020.8142
111. Garassino MC, Whisenant JG, Huang L-C, et al. COVID-19 in patients with thoracic malignancies (TERAVOLT): first results of an international, registry-based, cohort study. *The Lancet Oncology* 2020;21(7):914-22. doi: 10.1016/S1470-2045(20)30314-4
112. Higuchi T, Nishida T, Iwahashi H, et al. Early Clinical Factors Predicting the Development of Critical Disease in Japanese Patients with COVID-19: A Single-Center Retrospective, Observational Study. *MedRxiv* 2020 doi: 10.1101/2020.07.29.20159442 [published Online First: July 30]
113. Almazeedi S, Al-Youha S, Jamal MH, et al. Characteristics, risk factors and outcomes among the first consecutive 1096 patients diagnosed with COVID-19 in Kuwait. *EClinicalMedicine* 2020;24 doi: 10.1016/j.eclinm.2020.100448
114. Ayed M, Borahmah AA, Yazdani A, et al. Assessment of clinical characteristics and mortality-associated factors in COVID-19 Critical cases in Kuwait. *medRxiv* 2020 doi: 10.1101/2020.06.17.20134007 [published Online First: June 20]
115. Brouns SH, Brüggemann R, Linkens AEMJH, et al. Mortality and the Use of Antithrombotic Therapies Among Nursing Home Residents with COVID-19. *Journal of the American Geriatrics Society* 2020 doi: 10.1111/jgs.16664
116. Middeldorp S, Coppens M, van Haaps TF, et al. Incidence of venous thromboembolism in hospitalized patients with COVID-19. *Journal of thrombosis and haemostasis : JTH* 2020 doi: 10.1111/jth.14888

117. Davies MA. HIV and risk of COVID-19 death: a population cohort study from the Western Cape Province, South Africa. *medRxiv* 2020 doi: 10.1101/2020.07.02.20145185 [published Online First: July 3]
118. Choi HK, Koo H-J, Seok H, et al. ARB/ACEI use and severe COVID-19: a nationwide case-control study. *MedRxiv* 2020 doi: 10.1101/2020.06.12.20129916 [published Online First: 13 June]
119. Choi MH, Ahn H, Ryu HS, et al. Clinical Characteristics and Disease Progression in Early-Stage COVID-19 Patients in South Korea. *Journal of Clinical Medicine* 2020;9(6):1959. doi: 10.3390/jcm9061959
120. Chung SM, Lee YY, Ha E, et al. The Risk of Diabetes on Clinical Outcomes in Patients with Coronavirus Disease 2019: A Retrospective Cohort Study. *Diabetes Metab J* 2020;44(3):405-13. doi: 10.4093/dmj.2020.0105 [published Online First: 2020/07/01]
121. Huh K, Ji W, Kang M, et al. Association of previous medications with the risk of COVID-19: a nationwide claims-based study from South Korea. *MedRxiv* 2020 doi: 10.1101/2020.05.04.20089904 [published Online First: 8 May]
122. Hwang J-m, Kim J-H, Park J-S, et al. Neurological diseases as mortality predictive factors for patients with COVID-19: a retrospective cohort study. *Neurological Sciences* 2020:1. doi: 10.1007/s10072-020-04541-z
123. Kim J, Kim DW, Kim KI, et al. Compliance of Antihypertensive Medication and Risk of Coronavirus Disease 2019: a Cohort Study Using Big Data from the Korean National Health Insurance Service. *Journal of Korean medical science* 2020;35(25):e232. doi: 10.3346/jkms.2020.35.e232
124. Lee H, Ahn J, Kang C, et al. Association of Angiotensin II Receptor Blockers and Angiotensin-Converting Enzyme Inhibitors on COVID-19-Related Outcome (4/1/2020). . *Preprints with Lancet (SSRN)* 2020 doi: 10.2139/ssrn.3569837 [published Online First: 14 April]
125. Rhee SY, Lee J, Nam H, et al. Effects of a DPP-4 inhibitor and RAS blockade on clinical outcomes of patients with diabetes and COVID-19. *MedRxiv* 2020 doi: 10.1101/2020.05.20.20108555 [published Online First: 23 May]
126. Amat-Santos IJ, Santos-Martinez S, López-Otero D, et al. Ramipril in High Risk Patients with COVID-19. *Journal of the American College of Cardiology* 2020 doi: 10.1016/j.jacc.2020.05.040
127. Ayerbe L, Risco C, Ayis S. The association between treatment with heparin and survival in patients with Covid-19. *J Thromb Thrombolysis* 2020;50(2):298-301. doi: 10.1007/s11239-020-02162-z [published Online First: 2020/06/02]
128. Bernaola N, Mena R, Bernaola A, et al. Observational Study of the Efficiency of Treatments in Patients Hospitalized with Covid-19 in Madrid. *medRxiv* 2020 doi: 10.1101/2020.07.17.20155960 [published Online First: July 21]
129. de Abajo FJ, Rodríguez-Martín S, Lerma V, et al. Use of renin-angiotensin-aldosterone system inhibitors and risk of COVID-19 requiring admission to hospital: a case-population study. *Lancet (London, England)* 2020;395(10238):1705-14. doi: 10.1016/S0140-6736(20)31030-8
130. Fernández-Ruiz M, Andrés A, Loinaz C, et al. COVID-19 in solid organ transplant recipients: A single-center case series from Spain. *American journal of transplantation : official journal of the American Society of Transplantation and the American Society of Transplant Surgeons* 2020 doi: 10.1111/ajt.15929
131. Golpe R, Pérez-de-Llano LA, Dacal D, et al. Risk of severe COVID-19 in hypertensive patients treated with renin-angiotensin-aldosterone system inhibitors. *Medicina clinica* 2020 doi: 10.1016/j.medcli.2020.06.013
132. Jurado A, Martin MC, Abad-Molina C, et al. COVID-19: age, Interleukin-6, C-Reactive Protein and lymphocytes as key clues from a multicentre retrospective study in Spain. *MedRxiv* 2020 doi: 10.1101/2020.05.13.20101345 [published Online First: 16 May]

133. Lorente-Ros A, Monteagudo Ruiz JM, Rincón LM, et al. Myocardial injury determination improves risk stratification and predicts mortality in COVID-19 patients. *Cardiology journal* 2020 doi: 10.5603/CJ.a2020.0089
134. Marcos M, Belhassen-García M, Sánchez-Puente A, et al. Development of a severity of disease score and classification model by machine learning for hospitalized COVID-19 patients. *medRxiv* 2020 doi: 10.1101/2020.07.13.20150177 [published Online First: July 14]
135. Martínez-López j, Mateos M, Encinas C, et al. Multiple Myeloma and SARS-CoV-2 Infection: Clinical Characteristics and Prognostic Factors of Inpatient Mortality. *medRxiv* 2020 doi: 10.1101/2020.06.29.20142455 [published Online First: June 30]
136. Pérez-Sáez MJ, Blasco M, Redondo-Pachón D, et al. Use of tocilizumab in kidney transplant recipients with COVID-19. *American journal of transplantation : official journal of the American Society of Transplantation and the American Society of Transplant Surgeons* 2020 doi: 10.1111/ajt.16192
137. Poblador-Plou B, Carmona-Pírez J, Ioakeim-Skoufa I, et al. Baseline Chronic Comorbidity and Mortality in Laboratory-Confirmed COVID-19 Cases: Results from the PRECOVID Study in Spain. *International journal of environmental research and public health* 2020;17(14) doi: 10.3390/ijerph17145171
138. Regina J, Papadimitriou-Olivgeris M, Burger R, et al. Epidemiology, risk factors and clinical course of SARS-CoV-2 infected patients in a Swiss university hospital: an observational retrospective study. *MedRxiv* 2020 doi: 10.1101/2020.05.11.20097741 [published Online First: 14 May]
139. Pongpirul WA, Wiboonchutikul S, Charoenpong L, et al. Clinical course and potential predicting factors of pneumonia of adult patients with coronavirus disease 2019 (COVID-19): A retrospective observational analysis of 193 confirmed cases in Thailand. *medRxiv* 2020 doi: 10.1101/2020.06.24.20139642 [published Online First: June 26]
140. Selcuk M, Cinar T, Keskin M, et al. Is the use of ACE inb/ARBs associated with higher in-hospital mortality in Covid-19 pneumonia patients?, 2020.
141. Baker KF, Hanrath AT, Schim van der Loeff I, et al. COVID-19 management in a UK NHS Foundation Trust with a High Consequence Infectious Diseases centre: a detailed descriptive analysis. *MedRxiv* 2020 doi: 10.1101/2020.05.14.20100834 [published Online First: 19 May]
142. Bataille V, Visconti A, Rossi N, et al. Diagnostic value of skin manifestation of SARS-CoV-2 infection. *medRxiv* 2020 doi: 10.1101/2020.07.10.20150656 [published Online First: July 11]
143. Bean DM, Kraljevic Z, Searle T, et al. ACE-inhibitors and Angiotensin-2 Receptor Blockers are not associated with severe SARS-COVID19 infection in a multi-site UK acute Hospital Trust. *European journal of heart failure* 2020 doi: 10.1002/ejhf.1924
144. Fletcher RA, Matcham T, Tiburcio M, et al. Risk factors for clinical progression in patients with COVID-19: a retrospective study of electronic health record data in the United Kingdom. *MedRxiv* 2020 doi: 10.1101/2020.05.11.20093096 [published Online First: 15 May]
145. Ho FK, Celis-Morales CA, Gray SR, et al. Modifiable and non-modifiable risk factors for COVID-19: results from UK Biobank. *MedRxiv* 2020 doi: 10.1101/2020.04.28.20083295 [published Online First: 2 May]
146. Khawaja AP, Warwick AN, Hysi PG, et al. Associations with covid-19 hospitalisation amongst 406,793 adults: the UK Biobank prospective cohort study. *MedRxiv* 2020 doi: 10.1101/2020.05.06.20092957 [published Online First: 11 May]
147. Kolin DA, Kulm S, Elemento O. Clinical and Genetic Characteristics of Covid-19 Patients from UK Biobank. *MedRxiv : the preprint server for health sciences* 2020 doi: 10.1101/2020.05.05.20075507
148. Raisi-Estabragh Z, McCracken C, Ardissino M, et al. NON-WHITE ETHNICITY, MALE SEX, AND HIGHER BODY MASS INDEX, BUT NOT MEDICATIONS ACTING ON THE RENIN-ANGIOTENSIN SYSTEM ARE ASSOCIATED WITH CORONAVIRUS DISEASE 2019 (COVID-19) HOSPITALISATION:

REVIEW OF THE FIRST 669 CASES FROM THE UK BIOBANK. *MedRxiv* 2020 doi: 10.1101/2020.05.10.20096925 [published Online First: 15 May]
149. Raisi-Estabragh Z, Celeste M, Maddalena A, et al. Renin-Angiotensin-Aldosterone System Blockers Are Not Associated With Coronavirus Disease 2019 (COVID-19) Hospitalization: Study of 1,439 UK Biobank Cases. *Frontiers in Cardiovascular Medicine* 2020;7 doi: 10.3389/fcvm.2020.00138
150. Russell B, Moss C, Papa S, et al. Factors affecting COVID-19 outcomes in cancer patients - A first report from Guys Cancer Centre in London. *MedRxiv* 2020 doi: 10.1101/2020.05.12.20094219 [published Online First: 19 May]
151. Sivaloganathan H, Ladikou EE, Chevassut T. COVID-19 mortality in patients on anticoagulants and antiplatelet agents. *British journal of haematology* 2020 doi: 10.1111/bjh.16968
152. McKeigue PM, Kennedy S, Weir A, et al. Associations of severe COVID-19 with polypharmacy in the REACT-SCOT case-control study. *medRxiv* 2020 doi: 10.1101/2020.07.23.20160747 [published Online First: July 27]
153. Argenziano MG, Bruce SL, Slater CL, et al. Characterization and clinical course of 1000 patients with coronavirus disease 2019 in New York: retrospective case series. *BMJ: British Medical Journal (Online content)* 2020:1.
154. Auld SC, Caridi-Scheible M, Blum JM, et al. ICU and Ventilator Mortality Among Critically Ill Adults With Coronavirus Disease 2019. *Crit Care Med* 2020;48(9):e799-e804. doi: 10.1097/CCM.0000000000004457 [published Online First: 2020/05/27]
155. Bae DJ, Tehrani DM, Rabadia SV, et al. Angiotensin Converting Enzyme Inhibitor and Angiotensin II Receptor Blocker Use Among Outpatients Diagnosed With COVID-19. *Am J Cardiol* 2020 doi: 10.1016/j.amjcard.2020.07.007 [published Online First: 2020/08/21]
156. Bramante CT, Ingraham NE, Murray TA, et al. Observational Study of Metformin and Risk of Mortality in Persons Hospitalized with Covid-19. *medRxiv* 2020 doi: 10.1101/2020.06.19.20135095 [published Online First: June 20]
157. Castro VM, Ross RA, McBride SM, et al. Identifying common pharmacotherapies associated with reduced COVID-19 morbidity using electronic health records. *MedRxiv* 2020 doi: 10.1101/2020.04.11.20061994 [published Online First: 16 April]
158. Chang TS, Ding Y, Freund MK, et al. Prior diagnoses and medications as risk factors for COVID-19 in a Los Angeles Health System. *medRxiv* 2020 doi: 10.1101/2020.07.03.20145581 [published Online First: July 4]
159. Dublin S, Walker R, Floyd JS, et al. Renin-angiotensin-aldosterone system inhibitors and COVID-19 infection or hospitalization: a cohort study. *medRxiv* 2020 doi: 10.1101/2020.07.06.20120386 [published Online First: July 7]
160. Ferguson J, Rosser JI, Quintero O, et al. Characteristics and Outcomes of Coronavirus Disease Patients under Nonsurge Conditions, Northern California, USA, March-April 2020. *Emerging infectious diseases* 2020;26(8) doi: 10.3201/eid2608.201776
161. Goshua G, Pine AB, Meizlish ML, et al. Endotheliopathy in COVID-19-associated coagulopathy: evidence from a single-centre, cross-sectional study. *The Lancet Haematology* 2020 doi: 10.1016/S2352-3026(20)30216-7
162. Gu T, Mack JA, Salvatore M, et al. COVID-19 outcomes, risk factors and associations by race: a comprehensive analysis using electronic health records data in Michigan Medicine. *MedRxiv* 2020 doi: 10.1101/2020.06.16.20133140 [published Online First: 18 June]
163. Gupta S, Hayek SS, Wang W, et al. Factors Associated With Death in Critically Ill Patients With Coronavirus Disease 2019 in the US. *JAMA internal medicine* 2020 doi: 10.1001/jamainternmed.2020.3596
164. Hippensteel JA, Burnham EL, Jolley SE. Prevalence of venous thromboembolism in critically ill patients with COVID-19. *British journal of haematology* 2020 doi: 10.1111/bjh.16908

165. Imam Z, Odish F, Gill I, et al. Older age and comorbidity are independent mortality predictors in a large cohort of 1305 COVID-19 patients in Michigan, United States. *Journal of internal medicine* 2020 doi: 10.1111/joim.13119
166. Ip A, Parikh K, Parrillo JE, et al. Hypertension and Renin-Angiotensin-Aldosterone System Inhibitors in Patients with Covid-19. *MedRxiv* 2020 doi: 10.1101/2020.04.24.20077388 [published Online First: 29 April]
167. Jillella DV, Janocko NJ, Nahab F, et al. Ischemic Stroke in COVID-19: An Urgent Need for Early Identification and Management. *MedRxiv* 2020 doi: 10.1101/2020.05.25.20111047 [published Online First: 26 May]
168. Khera R, Clark C, Lu Y, et al. Association of Angiotensin-Converting Enzyme Inhibitors and Angiotensin Receptor Blockers with the Risk of Hospitalization and Death in Hypertensive Patients with Coronavirus Disease-19. *MedRxiv : the preprint server for health sciences* 2020 doi: 10.1101/2020.05.17.20104943
169. Kim L, Garg S, O'Halloran A, et al. Risk Factors for Intensive Care Unit Admission and In-hospital Mortality among Hospitalized Adults Identified through the U.S. Coronavirus Disease 2019 (COVID-19)-Associated Hospitalization Surveillance Network (COVID-NET). *Clinical infectious diseases : an official publication of the Infectious Diseases Society of America* 2020 doi: 10.1093/cid/ciaa1012
170. King CS, Sahjwani D, Brown AW, et al. Outcomes of Mechanically Ventilated Patients with COVID-19 Associated Respiratory Failure. *medRxiv* 2020 doi: 10.1101/2020.07.16.20155580 [published Online First: July 18]
171. Lala A, Johnson KW, Januzzi JL, et al. Prevalence and Impact of Myocardial Injury in Patients Hospitalized with COVID-19 Infection. *Journal of the American College of Cardiology* 2020 doi: 10.1016/j.jacc.2020.06.007
172. Lam KW, Chow KW, Vo J, et al. Continued in-hospital ACE inhibitor and ARB Use in hypertensive COVID-19 patients is associated with positive clinical outcomes. *The Journal of infectious diseases* 2020 doi: 10.1093/infdis/jiaa447
173. Lobelo F, Bienvenida A, Leung S, et al. Clinical, Behavioral and Social Factors Associated with Racial Disparities in Hospitalized and Ambulatory COVID-19 Patients from an Integrated Health Care System in Georgia. *medRxiv* 2020 doi: 10.1101/2020.07.08.20148973 [published Online First: July 10]
174. Lubetzky M, Aull M, Craig-Shapiro R, et al. Kidney Allograft Recipients Diagnosed with Coronavirus Disease-2019: A Single Center Report. *MedRxiv* 2020 doi: 10.1101/2020.04.30.20086462 [published Online First: 5 May]
175. Morales DR, Conover MM, You SC, et al. Renin-angiotensin system blockers and susceptibility to COVID-19: a multinational open science cohort study. *MedRxiv* 2020 doi: 10.1101/2020.06.11.20125849 [published Online First: 12 June]
176. Nguyen AB, Upadhyay GA, Chung B, et al. OUTCOMES AND CARDIOVASCULAR COMORBIDITIES IN A PREDOMINANTLY AFRICAN AMERICAN POPULATION WITH COVID-19. *medRxiv* 2020 doi: 10.1101/2020.06.28.20141929 [published Online First: June 29]
177. Palaiodimos L, Kokkinidis DG, Li W, et al. Severe obesity, increasing age and male sex are independently associated with worse in-hospital outcomes, and higher in-hospital mortality, in a cohort of patients with COVID-19 in the Bronx, New York. *Metabolism* 2020;108 doi: 10.1016/j.metabol.2020.154262
178. Paranjpe I, Fuster V, Lala A, et al. Association of Treatment Dose Anticoagulation with In-Hospital Survival Among Hospitalized Patients with COVID-19. *Journal of the American College of Cardiology (JACC)* 2020;75(17):N.PAG-N.PAG. doi: 10.1016/j.jacc.2020.05.001
179. Ramachandran P, Perisetti A, Gajendran M, et al. Prehospitalization Proton Pump Inhibitor (PPI) use and Clinical Outcomes in COVID-19. *medRxiv* 2020 doi: 10.1101/2020.07.12.20151084 [published Online First: July 14]

180. Reyes Gil M, Gonzalez-Lugo JD, Rahman S, et al. Correlation of coagulation parameters with clinical outcomes in Coronavirus-19 affected minorities in United States: Observational cohort. *MedRxiv* 2020 doi: 10.1101/2020.05.01.20087932 [published Online First: 6 May]
181. Rodriguez-Nava G, Trelles-Garcia DP, Yanez-Bello MA, et al. Atorvastatin associated with decreased hazard for death in COVID-19 patients admitted to an ICU: a retrospective cohort study. *Critical Care* 2020;24(1) doi: 10.1186/s13054-020-03154-4
182. Shah SJ, Barish PN, Prasad PA, et al. Clinical features, diagnostics, and outcomes of patients presenting with acute respiratory illness: a comparison of patients with and without COVID-19. 2020 doi: 10.1101/2020.05.02.20082461 [published Online First: 6 May]
183. Solaimanzadeh I. Nifedipine and Amlodipine Are Associated With Improved Mortality and Decreased Risk for Intubation and Mechanical Ventilation in Elderly Patients Hospitalized for COVID-19. *Cureus* 2020;12(5):e8069. doi: 10.7759/cureus.8069
184. Wang B, Van Oekelen O, Mouhieddine TH, et al. A tertiary center experience of multiple myeloma patients with COVID-19: lessons learned and the path forward. *Journal of Hematology & Oncology* 2020;13(1) doi: 10.1186/s13045-020-00934-x
185. Sterne JAC, Savovic J, Page MJ, et al. RoB 2: a revised tool for assessing risk of bias in randomised trials. *BMJ* 2019;366:l4898. doi: 10.1136/bmj.l4898 [published Online First: 2019/08/30]
